# Dried blood spot proteomics as a diagnostic framework for citrin deficiency

**DOI:** 10.64898/2026.05.26.26354012

**Authors:** Eriko Totsune, Daisuke Nakajima, Ryo Konno, Yasuko Mikami-Saito, Natsuko Arai-Ichinoi, Hikaru Nishida, Hiroko Yagi, Takashi Ishige, Hoshiro Suzuki, Matsuyuki Shirota, Jun Takayama, Chika Takano-Asai, Masaru Shimura, Hideo Sasai, Tomoko Lee, Jun Kido, Yoko Nakajima, Hironori Kobayashi, Atsuo Kikuchi, Chikahiko Numakura, Takashi Hamazaki, Kimihiko Oishi, Kimitoshi Nakamura, Yusuke Kawashima, Osamu Ohara, Yoichi Wada

**Affiliations:** Department of Pediatrics, Tohoku University Graduate School of Medicine, Sendai, Japan; Department of Applied Genomics, Kazusa DNA Research Institute, Kisarazu, Japan; Department of Pediatrics, The Jikei University School of Medicine, Tokyo, Japan; Department of Pediatrics, Hirosaki University Graduate School of Medicine, Hirosaki, Japan; Department of Pediatrics, Gunma University Graduate School of Medicine, Maebashi, Japan; Department of Pediatrics, Iwaki City Medical Center, Iwaki, Japan; Department of AI and Innovative Medicine, Tohoku University Graduate School of Medicine, Sendai, Japan; Tohoku Medical Megabank Organization, Tohoku University, Sendai, Japan; Statistical Genetics Team, RIKEN Center for Advanced Intelligence Project, Tokyo, Japan; Advanced Research Center for Innovations in Next-Generation Medicine, Tohoku University, Sendai, Japan; Department of Pediatrics and Child Health, Nihon University School of Medicine, Tokyo, Japan; Department of Metabolism, Chiba Children’s Hospital, Chiba, Japan; Department of Early Diagnosis and Preventive Medicine for Rare Intractable Pediatric Diseases, Graduate School of Medicine, Gifu University, Gifu, Japan; Department of Pediatrics, Hyogo Medical University, Nishinomiya, Japan; Department of Pediatrics, Faculty of Life Sciences, Kumamoto University, Kumamoto, Japan; Department of Pediatrics, Fujita Health University School of Medicine, Toyoake, Japan; Laboratories Division, Shimane University Hospital, Shimane, Japan; Department of Clinical Genomics, Saitama Medical University, Saitama, Japan; Department of Pediatrics, Yamagata University Faculty of Medicine, Yamagata, Japan; Department of Pediatrics, Osaka Metropolitan University Graduate School of Medicine, Osaka, Japan; Department of Medical Science and Innovation, SiRIUS Institute of Medical Research, Tohoku University, Sendai, Japan

**Author notes:** Corresponding authors: Yusuke Kawashima, Department of Applied Genomics, Kazusa DNA Research Institute, Kisarazu, Chiba 292-0818, Japan, Osamu Ohara, Department of Applied Genomics, Kazusa DNA Research Institute, Kisarazu, Chiba 292-0818, Japan, Yoichi Wada, Department of Medical Science and Innovation, SiRIUS Institute of Medical Research, Tohoku University, 1-1 Seiryomachi, Aobaku, Sendai, Miyagi 980-8574, Japan. These authors contributed equally (Eriko Totsune, Daisuke Nakajima).

**Keywords:** Proteomics, Proteome, Dried Blood Spot Testing, Citrin Deficiency, Proteomic Signature, SLC25A13, Diagnosis

## Abstract

**Background:** Citrin deficiency, caused by biallelic pathogenic variants in *SLC25A13*, must be identified early to prevent serious complications such as hyperammonemia and liver failure. However, clinical diagnosis is often delayed due to its nonspecific presentation and limited sensitivity of amino acid–based newborn screening methods. Although genome-based evaluations are being investigated to address these issues, concerns about their cost, turnaround time, variant interpretation ability, and data handling highlight the need for a more practical yet reliable alternative. We investigated the feasibility of applying proteomic approach on dried blood spots (DBS), which are routinely used in newborn screening.

**Methods:** We performed untargeted liquid chromatography-tandem mass spectrometry to analyze the proteome of DBS using a previously developed “non-targeted analysis of non-specifically DBS-absorbed proteins” (NANDA) workflow. SLC25A13 protein abundance was quantified in individuals with biallelic loss-of-function mutations, compound loss-of-function/missense mutations, and heterozygous carriers; this was also evaluated in healthy and diseased controls representing relevant differential diagnoses. To leverage proteomic information, we derived a multivariate proteomic signature using feature selection and evaluated its performance with leave-one-out cross-validation. Biological relevance was assessed by enrichment analysis, and complementary transcriptomics was performed using RNA sequencing.

**Results:** A total of 7,474 proteins, including SLC25A13, were consistently detected in DBS. SLC25A13 was undetectable in individuals with biallelic loss-of-function mutations. However, individuals with compound loss-of-function/missense genotypes showed reduced but measurable SLC25A13 levels, comparable to those observed in heterozygous carriers. In contrast, a compact 15-protein signature accurately identified individuals with compound loss-of-function/missense genotypes (AUC, 0.99; sensitivity, 1.00; specificity, 0.95). The signature was enriched for Ca^2+^-response, and transcriptomics showed downregulation of genes related to multimodal ion channels in affected individuals compared to controls.

**Conclusions:** DBS-based proteomic profiling may assist in the diagnosis of citrin deficiency through SLC25A13-quantification and a biologically plausible multivariate signature. More broadly, this strategy offers a promising new diagnostic layer for protein disorders, providing a proteomic readout in a clinically practical DBS format with potential utility for future diagnostic and screening applications.

## Background

*SLC25A13* (HGNC:10983) encodes citrin, a calcium-binding aspartate–glutamate carrier localized to the inner mitochondrial membrane and is prominently expressed in the liver.^1^ Citrin is an essential component of the malate–aspartate shuttle, where it contributes to the cytosolic–mitochondrial redox homeostasis by supporting NADH/NAD^+^ cycling.^2–4^ Biallelic pathogenic variants of *SLC25A13* cause citrin deficiency, an autosomal recessive inborn error of metabolism with age-dependent clinical manifestations.^1,5^ The phenotypic spectrum typically includes neonatal intrahepatic cholestasis caused by citrin deficiency (NICCD, MIM #605814), an apparent compensation period, and adult-onset type II citrullinemia (CTLN2, MIM #603471).^6,7^ Although NICCD is often transient, some affected infants experience severe or irreversible liver injury, including cirrhosis.^8–10^ CTLN2 is a later-onset phenotype that can present with recurrent hyperammonemia, encephalopathy, or liver failure, causing substantial morbidity and potentially fatal outcomes.^11–13^ Although the NICCD–CTLN2 interval is often clinically asymptomatic, a few develop manifestations such as hypoglycemia or short stature.^14–17^ As appropriate dietary interventions, including medium-chain triglyceride supplementation and/or a high-fat, high-protein and low-carbohydrate diet, can mitigate disease progression, early and accurate diagnosis is critical.^18–21^

Despite this clinical need, citrin deficiency remains difficult to identify in routine practice because its presentation is often non-specific, and the currently used newborn evaluation strategies have limited sensitivity.^9,22,23^ Citrulline, an indirect downstream marker of citrin dysfunction and the most widely used screening analyte for citrin deficiency, may yield false-negative results.^24,25^ Although genetic tests are the diagnostic standard for citrin deficiency, genome-based screening of newborns is being explored; its use as a primary population-level evaluation modality remains limited by cost, turnaround requirements, challenges in large-scale variant interpretation, and operational issues, including secondary findings and genomic data stewardship.^26–28^ Together, these limitations highlight the need for a complementary diagnostic approach combining analytical reliability with practical scalability.

To address the drawbacks of metabolite- and genotype-based strategies, we profiled the proteome of dried blood spots (DBSs) as a practical, complementary approach for diagnosing citrin deficiency. DBSs are minimally invasive, routinely collected in neonatal care, and well-suited for storage, transport, and large-scale implementation. Using a workflow we previously established for the non-targeted analysis of non-specifically DBS-absorbed proteins (NANDA),^29^ we examined whether DBS proteomics can detect SLC25A13 loss and distinguish affected individuals from controls with clinically meaningful diagnostic performance.

## Methods

### Study participants and data collection

For this study, 23 individuals with citrin deficiency and their guardians were recruited at seven institutions. Clinical and laboratory data, including genotypes, comorbidities, and metabolic profiles, were obtained from their family physicians. The diagnostic and carrier statuses were confirmed by analyzing mutations prevalent among the Japanese population or by panel sequencing. Further, six healthy adults were recruited as controls. For differential diagnosis, we included two individuals with Fanconi–Bickel syndrome (MIM #227810), two with ornithine transcarbamylase deficiency (MIM #311250), one with argininosuccinic aciduria (MIM #207900), three with citrullinemia type I (MIM #215700), two with galactosemia type I (MIM #230400), two with galactosemia type II (MIM #230200), three with galactosemia type III (MIM #230350), four with galactosemia type IV (MIM #618881), and two with portosystemic shunts. The diagnosis of portosystemic shunts was based on abdominal ultrasound findings, elevated blood bile acid levels, and the absence of findings suggestive of other conditions.

### RNA sequencing

Peripheral blood mononuclear cells (PBMCs) were isolated from venous blood samples in a heparin sodium tube. Total RNA from PBMCs was extracted using TRIzol™ Reagent (Thermo Fisher Scientific, Carlsbad, CA, USA) according to the manufacturer’s protocol. Poly(A)+ RNA was subsequently isolated from the total RNA using the NEBNext™ Poly(A) mRNA Magnetic Isolation Module (New England Biolabs, Ipswich, MA, USA).

RNA sequencing (RNA-seq) libraries were prepared from the poly(A)+ RNA using the xGen Broad-Range RNA Library Prep Kit (Integrated DNA Technologies, Coralville, IA, USA) and sequenced on the NextSeq 2000 or NovaSeq X Plus platform in 150-bp paired-end mode (Illumina, San Diego, CA, USA). Before sequencing, mRNA transcripts derived from causative genes of rare diseases, including inborn errors of metabolism, by hybridization capture using probes synthesized by TWIST Bioscience (South San Francisco, CA, USA). Gene-level read counts were generated using featureCounts (Subread v.2.1.1) from paired-end RNA-seq data with the GENCODE v.44 annotation (GRCh38).

All downstream analyses were conducted in R (v.4.5.2). Count data were imported into DESeq2 (v.1.48.2). Sample-level metadata were derived from sample names and used to define genotype, clinical status, and pre-specified technical batch. Differential gene expression was analyzed within the DESeq2 negative binomial generalized linear model framework using the design formula ∼ batch + status. Normalization was performed by estimating size factors using the median-of-ratios method. Dispersion estimation and model fitting were performed using the standard DESeq2 workflow. The primary contrast of interest was patients versus controls, with the control group set as the reference. Shrinkage of log2 fold changes was performed using apeglm when available.

For exploratory analysis, expression values were subjected to variance-stabilizing transformation (VST). Principal component analysis (PCA) was performed using both the unmodified VST matrix and a batch-residualized VST matrix. The batch-residualized matrix was obtained by fitting a linear model for each gene with batch as the explanatory variable and extracting the residuals. Residualized PCA was used only for visualization, whereas batch effects in differential expression testing were adjusted by explicitly including batch in the DESeq2 design. Volcano and MA plots were generated from the DESeq2 results for the patient-versus-control comparison. Significant categories were defined based on adjusted *P* values and effect–size thresholds. For gene set enrichment analysis (GSEA), ranked gene lists were generated from DESeq2 Wald statistics after mapping Ensembl identifiers to Entrez Gene IDs with org.Hs.eg.db. For Ensembl IDs mapping to the same Entrez Gene ID, the entry with the largest absolute test statistic was retained. Gene Ontology enrichment analyses for Biological Process, Molecular Function, and Cellular Component were performed using clusterProfiler,^30^ and Reactome pathway enrichment analysis was performed using ReactomePA. Analyses were run with minGSSize = 10 and maxGSSize = 500, and *P* values were adjusted using the Benjamini-Hochberg procedure.

### DBS proteome analysis

DBS samples were obtained from venous blood of the participants. Protein extraction and digestion from DBS was performed according to our previously reported method, the automated preparation of proteins for NANDA.^29^ In brief, 96-well plates compatible with the Maelstrom 9610 instrument (TANBead) was prepared. The first plate was loaded with 500 µL of 1× TBST, 25 µL of freshly prepared iron powder suspension containing 5 mg of iron powder in 50% glycerol (particle size 3–5 µm; Kojundo Chemical Lab. Co., Ltd., Japan), and one 3.2 mm-diameter disk punched from DBS. The second plate was filled with 500 µL of 1× TBST, whereas the third and fourth plates each received 500 µL of 50 mM Tris-HCl (pH 8.0). The fifth plate was filled with 200 µL of 50 mM Tris-HCl (pH 8.0), 10 mM CaCl_2_, and 0.012% lauryl maltose neopentyl glycol (Anatrace Products, Maumee, OH, USA).^31^ Subsequently, the plates were mounted on the Maelstrom 9610, and protein extraction from DBS was initiated. A spin tip was inserted into the first plate, and the DBS was disrupted by agitating in TBST for 30 min at 3,500 rpm with inversion every 10 s, forming a pulp-like DBS–iron powder complex. A magnetic rod was then inserted into the spin tip, and the complex was captured and collected on the tip for 60 s before being transferred to the second plate containing TBST. After removal of the magnetic rod, the complex was washed with agitation for 10 min at 3,500 rpm with inversion every 10 s. The complex was subsequently transferred to the third plate containing 50 mM Tris-HCl (pH 8.0) and washed under the same conditions for 10 min. This washing step was repeated in the fourth plate containing 50 mM Tris-HCl (pH 8.0), with agitation for 2 min. Finally, the complex was transferred to the fifth plate and suspended by agitation for 2 min at 3,500 rpm, with inversion every 10 s.

For protein digestion, 1 µg of Trypsin Platinum (CAT# VA9000; Promega, Madison, WI, USA) was gently mixed with the sample for 14 h at 37 °C. After the filter paper was removed from the digested sample using the Maelstrom 8-Autostage/Maelstrom 9610, the isolated supernatant was acidified with 50 µL of 5% trifluoroacetic acid (TFA). The sample was then desalted using a styrene–divinylbenzene polymer stop-and-go extraction tip (SDB-STAGE tip; GL Sciences, Tokyo, Japan). The tip was washed with 25 µL of 80% acetonitrile (ACN) in 0.1% TFA and equilibrated with 50 µL of 3% ACN in 0.1% TFA. The sample was loaded onto the tip, washed with 80 µL of 3% ACN in 0.1% TFA, and eluted with 50 µL of 36% ACN in 0.1% TFA. The eluate was dried in a centrifugal evaporator (miVac Duo concentrator; Genevac, Ipswich, UK). The dried sample was then redissolved in 0.02% decyl maltose neopentyl glycol (Anatrace Products, LLC, Maumee, OH, USA) and 0.1% TFA. Peptide concentration was measured using a Lunatic instrument (Unchained Labs, Pleasanton, CA, USA) and the sample was transferred to an LC vial (Thermo Fisher Scientific, Waltham, MA, USA).

Redissolved peptides, corresponding to 200 ng, were directly injected into a 75 µm × 30 cm C18 column (1.7 µm particle size, 100 Å; CoAnn Technologies, Richland, WA, USA) maintained at 50 °C, and separated using a 90 min gradient on an UltiMate 3000 RSLCnano system (Thermo Fisher Scientific) at a flow rate of 150 nL/min. Solvent A was 0.1% formic acid in water, and solvent B was 0.1% formic acid in 80% acetonitrile. The gradient was as follows: 6% B at 0 min, 34% B at 76 min, 70% B at 83 min, and 70% B at 90 min, Eluted peptides were analyzed by data-independent acquisition liquid chromatography–tandem mass spectrometry (DIA LC-MS/MS) using an Orbitrap Exploris 480 mass spectrometer (Thermo Fisher Scientific) equipped with an InSpIon system (AMR, Tokyo, Japan).^32^ Full-scan spectra were acquired over an *m/z* range of 495–705 range at a resolution of 60,000, with an automatic gain control target of 300% and a maximum injection time set to “Auto.” Fragment ion spectra were acquired over an *m/z* range of 200–1,800 at a resolution of 60,000, with an automatic gain control target of 3,000%, a maximum injection time set to “Auto,” and stepped normalized collision energies of 22%, 26%, and 30%. The optimized window was set at 4 Th, and the acquisition method was configured using Xcalibur 4.3 (Thermo Fisher Scientific).

The mass spectrometry data were queried against an *in silico* human spectral library using DIA-NN v.2.2.0 (https://github.com/vdemichev/DiaNN).^33^ Initially, the spectral library was generated from the UniProt human protein sequence database (proteome ID UP000005640; 20,644 entries; downloaded on April 4, 2025) using DIA-NN. The parameters for spectral library generation were as follows: digestion enzyme, trypsin; missed cleavages, 1; peptide length range, 7–45; precursor charge range, 2–4; precursor mass range, 495–705; and fragment ion *m/z* range, 200–1,800. Additionally, “FASTA digest for library-free search/library generation,” “deep learning-based spectra, RTs and IM prediction,” and “n-term M excision” were enabled. For the DIA-NN search, the following parameters were applied: MS2 accuracy, 10 ppm; MS1 accuracy, 10 ppm; quantification strategy, QuantUMS high precision; and cross-run normalization, RT-dependent. Precursor- and protein-level false discovery rates were controlled at less than or equal to 1%. Protein quantities were calculated by aggregating the quantities of unique peptides as determined by DIA-NN.

### Proteomics preprocessing

Proteomic intensities were curated using sample metadata, including batch and clinical group labels. Intensities were log2-transformed, and zero values were treated as missing. ACTB (HGNC:132)-based normalization was applied; for each sample, all log2-transformed protein intensities were centered by subtracting the log2-transformed ACTB intensity. Proteins were retained only if supported at least three unique peptides. Missing values were imputed using MinProb imputation (imputeLCMD::impute.MinProb; q = 0.01; seed = 2025), which replaces missing observations with low-intensity values consistent with left-censored proteomics data. Batch correction was then performed using ComBat (sva::ComBat).

For downstream modeling, the corrected protein matrix was transposed into a sample-by-feature format. Features containing non-finite values across samples or showing zero variance were removed. Feature-wise standardization was then applied before model fitting. In cross-validation analyses, means and standard deviations were estimated using the training subset only and then applied to the held-out sample, thereby preventing information leakage. Separately, row-wise z-scores were used only for visualization, such as heatmaps, and were not used for model fitting.

### Model development and feature selection

The primary classification task was to discriminate the compound loss-of-function/missense (LoF/Mis) group from individuals without biallelic LoF/LoF genotypes. The positive class was defined as the LoF/Mis group. Individuals with LoF/LoF genotypes were excluded from model training and evaluation. Accordingly, the classifier was trained to distinguish individuals with LoF/Mis genotypes from those with reference/reference (Ref/Ref) or Ref/LoF genotypes, while individuals with LoF/LoF genotypes were excluded. A penalized logistic regression classifier was trained using elastic net regularization implemented in glmnet, with the mixing parameter fixed at α = 0.5. To account for class imbalance, inverse-prevalence class weighting was applied during model fitting. The regularization strength, λ, was selected by stratified 3-fold cross-validation using the area under the receiver operating characteristic curve (AUC) as the optimization metric. Feature ranking was performed by stability selection. Specifically, model fitting was repeated across B = 30 class-stratified subsamples, each containing 80% of samples from each class. Within each subsample, proteins were ranked according to whether their coefficients were non-zero at the λ value selected by cross-validation. Stability was quantified as the selection frequency, defined as the proportion of subsamples in which a feature received a non-zero coefficient. Candidate panel sizes were pre-specified as m = 5, 10, 15, 20, 30, 40, and 50, and these candidate panels were evaluated by leave-one-out cross-validation (LOOCV). In the final analysis script, the panel size was fixed *a priori* at m = 30 for the final model. Discrimination was summarized by the AUC. The decision threshold, τ, was chosen to maximize sensitivity subject to a specificity of at least 0.90.

### Final model and scoring rule

After the panel size had been prespecified as 30, the highest-ranked proteins from the stability analysis were selected as the panel, and the elastic net model was refit using all samples in the analysis set.

Let x_ij_ denote the preprocessed log-intensity of feature *j* for sample *i*. For each feature *j*, standardization was performed as

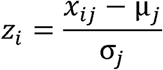

, where μ*_j_*and σ*j* denote the mean and standard deviation, respectively, estimated from the training data. The linear predictor was calculated as

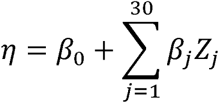

, and converted to a predicted probability using the logistic function:

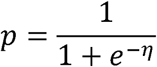

. The operating threshold τ was then applied to classify each sample as positive or negative.

### Multivariate visualization

Sparse partial least squares discriminant analysis (sPLS-DA) was performed as an auxiliary visualization analysis using the top 1,000 proteins ranked by elastic net stability selection, implemented in mixOmics. Models were fit with two components, and keepX, defined as the number of proteins retained per component, was evaluated over 5, 10, 15, 20, 30, 40, and 50. Performance was assessed by LOOCV using the balanced error rate, and selection stability was summarized across LOOCV refits.

### Statistical environment

All statistical analyses and visualizations were performed in R version 4.3.2 (R Foundation for Statistical Computing, Vienna, Austria). Unless otherwise specified, two-sided tests were used, and *P* value < 0.05 was considered statistically significant.

## Results

### Proteomic analysis of DBS enables robust quantification of SLC25A13 but does not fully resolve genotype subgroups

We first investigated whether proteomic analysis of DBS, a convenient and minimally invasive specimen type routinely used in newborn screening, could serve as a diagnostic platform for citrin deficiency (Fig. 1). DBS samples were obtained from five groups: 6 healthy controls, 10 heterozygous carriers, 23 individuals with citrin deficiency (16 with LoF/LoF genotypes and 7 with LoF/Mis genotypes), and 21 disease controls representing relevant differential diagnoses encountered in newborn screening or clinical practice (Tables 1, S1, and S2). DIA LC-MS/MS analysis using the NANDA pipeline detected 7,474 proteins, including SLC25A13, across DBS samples (Table S3).

**Figure 1.**
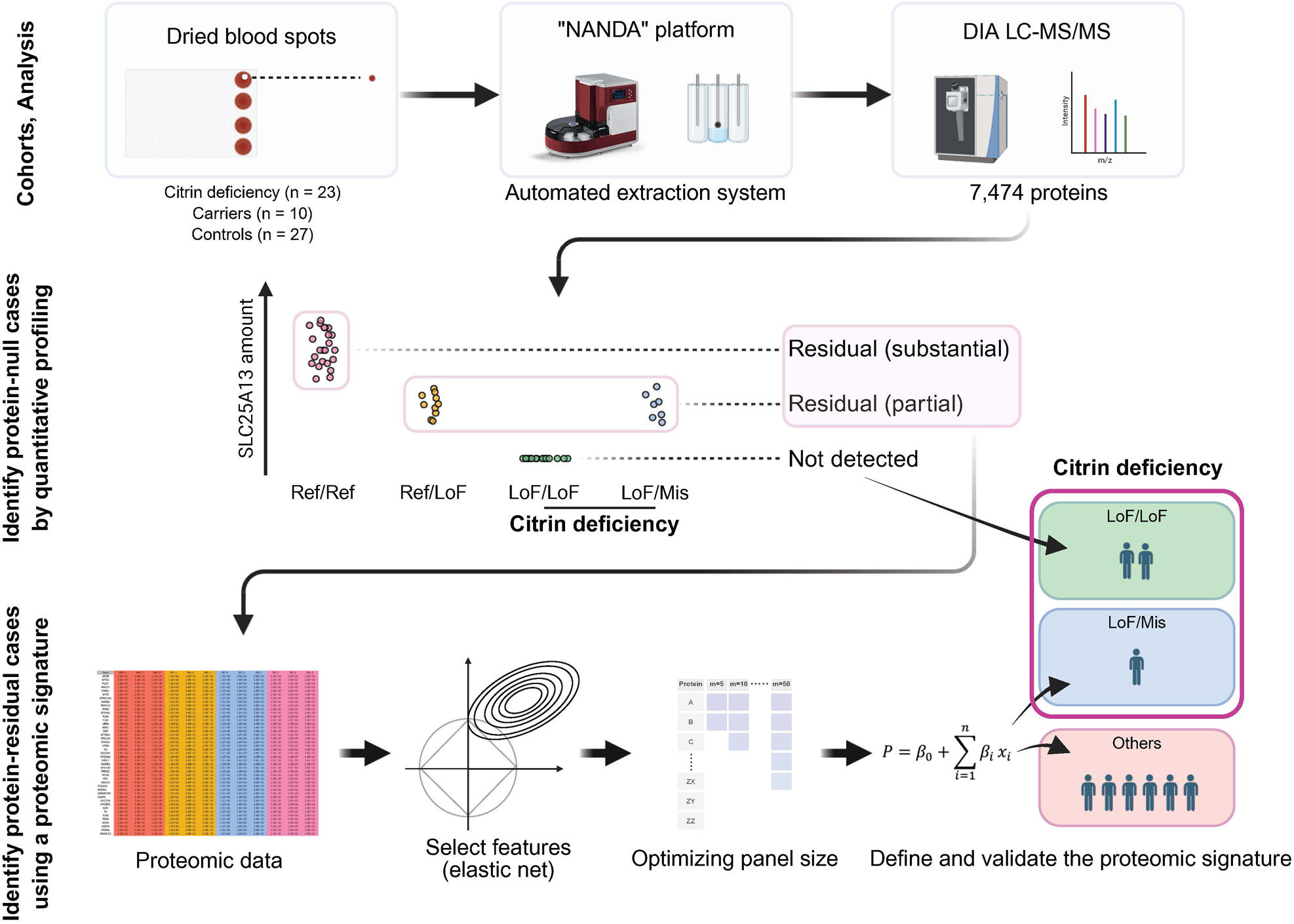
Study design Dried blood spot specimens from the participants were processed using the non-targeted analysis of non-specifically DBS-absorbed proteins (NANDA) platform, an automated extraction system for data-independent acquisition-liquid chromatography-tandem mass spectrometry (DIA LC-MS/MS)-based proteomic analysis. Quantitative proteomic data were used to identify individuals with biallelic loss-of-function (LoF/LoF) genotypes. In the next step, individuals with compound loss-of-function/missense (LoF/Mis) genotypes, in whom SLC25A13 levels decreased but not absent, were identified using a proteomic signature derived by elastic net regression and validated by leave-one-LoF/Mis-out stability analysis. NANDA, non-targeted Analysis of Non-specifically DBS-Absorbed proteins; DIA LC-MS/MS, data-independent acquisition-liquid chromatography-tandem mass spectrometry; LoF, loss-of-function; Mis, missense; Ref, reference. Created with BioRender.com.

SLC25A13 was undetectable in individuals with LoF/LoF genotypes (Fig. 2A). In contrast, SLC25A13 abundance in heterozygous carriers and individuals with LoF/Mis genotypes was reduced to approximately half of that observed in healthy controls and disease controls (Fig. 2A). Accordingly, SLC25A13 abundance alone was insufficient to distinguish individuals with LoF/Mis genotypes from heterozygous carriers (Fig. 2A). Furthermore, neither the pattern of detected SLC25A13 peptides, the broader proteomic profile, nor the available clinical and biochemical findings reliably discriminated between individuals with LoF/LoF genotypes and those with LoF/Mis genotypes (Figs. S1, S2, and S3; Tables S4 and S5).

**Figure 2.**
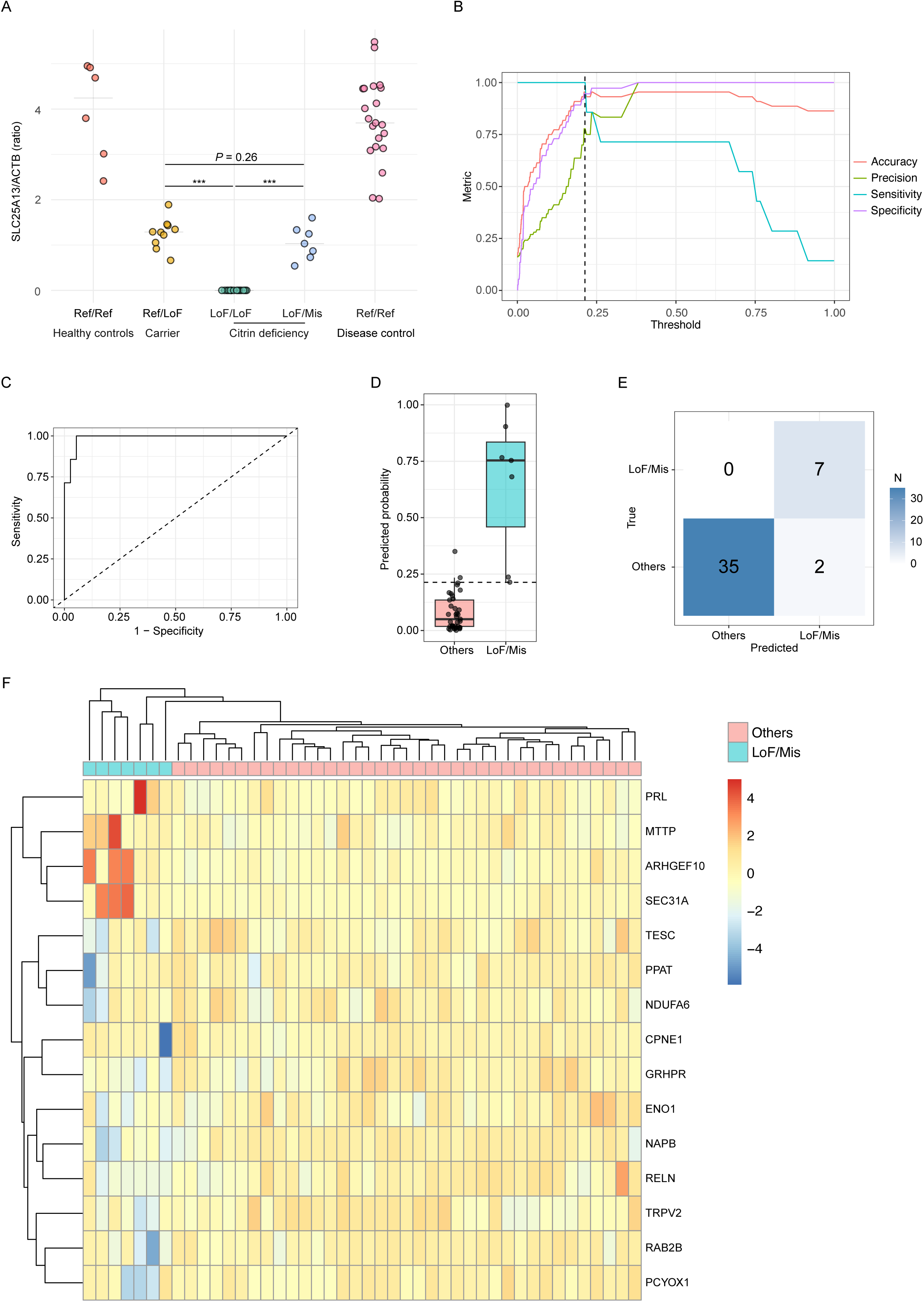
Proteomic analysis for identifying citrin deficiency. **A.** ACTB-normalized SLC25A13 abundance in dried blood spots of healthy controls (n = 6), carriers (n = 10), individuals with LoF/LoF *SLC25A13* variants (n = 16), individuals with LoF/Mis *SLC25A13* variants (n = 7), and disease controls (n = 21). Each dot represents an individual. Group comparisons were evaluated using Welch’s t-test; *** *P* < 0.001. **B.** Threshold-sweep plot evaluating the ability of the panel to identify LoF/Mis genotypes. The x-axis represents the decision threshold applied to predicted probabilities from the elastic net model, and the y-axis shows the corresponding performance metrics: sensitivity, specificity, precision, and accuracy. The dashed vertical line marks the selected threshold, chosen to maximize sensitivity while maintaining specificity ≥ 0.90. **C.** Receiver operating characteristic curve of the 30-protein panel for distinguishing LoF/Mis genotypes from the other groups after excluding LoF/LoF samples. The y-axis represents sensitivity, and the x-axis indicates 1 – specificity. **D.** Elastic net-predicted probabilities of LoF/Mis genotypes based on the 30-protein panel. The Others group denotes healthy controls, carriers, and disease controls after exclusion of LoF/LoF samples. The y-axis represents the predicted probability of the LoF/Mis genotype. Each dot represents an individual sample. Boxplots indicate the interquartile range, with the center line representing the median. The dashed horizontal line corresponds to a predictive probability cut-off of 0.213 used to classify LoF/Mis versus Others. **E.** Confusion matrix of the elastic net classification model at the selected decision threshold. Each cell shows the number of samples in the corresponding true/predicted category. **F.** Heatmap of the 15 proteins with non-zero coefficients in the final elastic net model derived from the 30-protein panel. Expression is shown as standardized log-intensities (z-scores). LoF, loss-of-function; Mis, missense; Ref, reference.

### A multivariate proteomic signature enables accurate identification of individuals with LoF/Mis genotypes

To identify individuals with LoF/Mis genotypes, we next leveraged the multivariate structure of the proteomic data rather than relying solely on SLC25A13 quantification (Fig. 1). Elastic net regularization (α = 0.5) was applied to classify this genotype class against the other groups after excluding samples with LoF/LoF genotypes. The regularization parameter, λ, was selected by stratified 3-fold cross-validation, and proteins were ranked by stability selection across 30 class-stratified subsampling runs, each including 80% of samples from each class. Pre-specified candidate panel sizes (m = 5, 10, 15, 20, 30, 40, and 50) were then evaluated by LOOCV, with the decision threshold chosen to maximize sensitivity subject to a specificity constraint of at least 0.90.

The 30-protein panel showed the best performance, achieving an AUC of 0.99, a sensitivity of 1.00, and a specificity of 0.95 at the selected decision threshold (Fig. 2B, 2C; Table S6). Consistent with these metrics, LOOCV-predicted probabilities were markedly higher in samples with LoF/Mis genotypes than in the other samples (Fig. 2D). The corresponding LOOCV confusion matrix confirmed correct identification of all samples with LoF/Mis genotypes, with only two samples from the other groups misclassified as positive (Fig. 2E). These results indicated that the 30-protein panel provided robust sample-level discrimination of LoF/Mis genotypes.

Among the 30 proteins, 15 retained non-zero coefficients in the final elastic net model, whereas the remaining 15 were shrunk to zero, yielding a compact 15-protein core signature within the selected panels (Fig. 2F; Table S7). Consistent with this result, sPLS-DA using two components reproduced separation of samples with LoF/Mis genotypes from the remaining groups in a low-dimensional feature space. Evaluation across the pre-specified keepX grid showed progressive improvement in classification performance up to keepX = 30, beyond which no further gain was observed, supporting 30 proteins as a parsimonious and appropriate panel size (Table S8). Additionally, leave-out stability analysis restricted to samples with LoF/Mis genotypes demonstrated complete retention of all proteins in the final panel across all omission runs, indicating that the selected proteomic signature was not driven by any single sample with LoF/Mis genotype (Table S9). Together, these results demonstrate that a stable multivariate proteomic signature can accurately distinguish individuals with LoF/Mis genotypes from other clinical and genetic groups.

### Proteomic and transcriptomic analyses converge on biologically coherent alterations related to calcium ion homeostasis

To examine the biological relevance of the proteomic signature, we performed ontology analysis of the 15-protein core signature. This analysis identified significant enrichment of the “response to calcium ion” category (Table S10), a finding of particular interest given the emerging link between SLC25A13/citrin and calcium-related biology.^34^ To obtain orthogonal molecular support for this observation, we then conducted RNA-seq analysis of PBMCs. Although *SLC25A13* transcript levels did not differ between samples with LoF/LoF genotypes (n = 7) and Ref/Ref (n = 6) samples (Fig. 3A, 3B, S4), GSEA revealed biologically coherent alterations in affected individuals, including signatures related not only to mitochondrial dysfunction, consistent with SLC25A13 deficiency, but also to calcium ion binding and calmodulin binding (Table S11). Pathways related to voltage-gated ion channel activity and transmembrane ion transport were also prominently enriched (Fig. 3C, 3D, S5), further supporting disruption of calcium ion homeostasis. Together, these transcriptomic findings provide orthogonal support for the biological validity of the proteomic signature.

**Figure 3.**
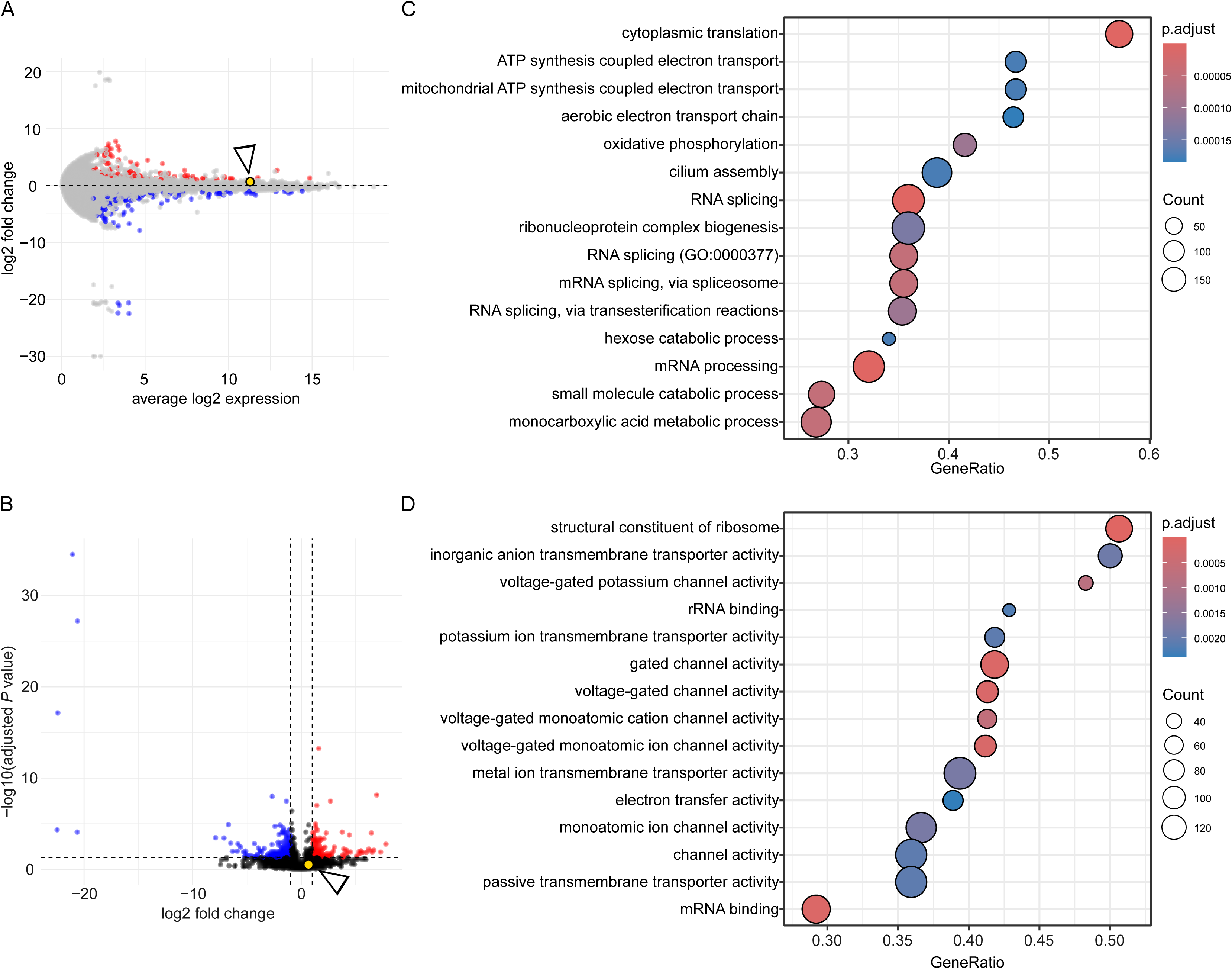
Transcriptomic signatures associated with citrin deficiency. **A.** MA plot of RNA-seq data comparing individuals with LoF/LoF genotypes and Ref/Ref controls. The y-axis denotes log2 fold change, and the x-axis represents the average log2 expression across samples. Each point represents one gene. Genes meeting the significance threshold, defined as adjusted *P* value < 0.05 and |log2 fold change| ≥ 1, are shown in red and blue when upregulated and downregulated, respectively. The disease-associated gene *SLC25A13* is highlighted in yellow with a white triangle. **B.** Volcano plot. The y-axis represents −log10(adjusted *P* value), and the x-axis indicates log2 fold change. Genes meeting the significance threshold are colored as in panel A. *SLC25A13* is highlighted as in panel A. **C, D.** Gene set enrichment analysis of Gene Ontology terms using the ranked RNA-seq differential expression from the LoF/LoF versus Ref/Ref comparison. The x-axis represents the gene ratio, and each point corresponds to an enriched GO term. Point size indicates the number of contributing genes, and point color denotes statistical significance based on adjusted *P* value. Selected biological process terms are shown in panel C, and molecular function terms are shown in panel D. LoF, loss-of-function; Ref, reference.

## Discussion

In this study, we evaluated DBS-based proteomic profiling as a protein-level approach for identifying individuals with citrin deficiency. In our cohort, SLC25A13 was undetectable in individuals with LoF/LoF genotypes, clearly distinguishing them from controls and supporting SLC25A13 abundance as a mechanism-proximal marker. Among individuals carrying missense variants who retained residual SLC25A13 protein, discrimination based on SLC25A13 abundance alone was incomplete; however, a parsimonious 15-protein panel improved separation in our dataset. Collectively, these findings support DBS proteomics as a diagnostic strategy that provides a direct protein-level disease signal complementary to genotype- and metabolite-based approaches.

This proof of concept is clinically important because current diagnosis still depends largely on genetic testing and biochemical surrogates, both of which have practical limitations. Genetic testing may leave uncertainty when variants of uncertain significance are identified, and biochemical markers do not directly measure the defining molecular consequence of disease, namely loss or reduction of citrin protein.^35,36^ By contrast, SLC25A13 abundance represents a mechanism-proximal readout, while the DBS format offers additional practical advantages, including minimally invasive sampling, compatibility with decentralized collection, and potential applicability in neonatal or pediatric settings. With further work to establish age-appropriate reference ranges and evaluate the effects of clinical state and pre-analytical conditions, DBS proteomics may provide a clinically useful adjunct in settings where rapid and biologically direct assessment is desirable.

Proteome-wide profiling added diagnostic information beyond single-analyte quantification. In individuals with residual SLC25A13 protein attributable to missense alleles, SLC25A13 abundance alone provided only partial separation, whereas a compact multivariate protein panel improved discrimination in our dataset. This finding suggests that proteome-level context may help resolve diagnostically challenging cases in which the primary protein is not completely absent.^37,38^ Because this panel was derived and evaluated in a cohort with a limited sample size, external validation in independent samples using pre-specified decision criteria will be essential for clinical implementation.

As supportive biological context, pathway-level enrichment of calcium ion response-related terms in the proteomic signature, together with concordant transcriptomic signals, is consistent with prior literature linking citrin deficiency and mitochondrial metabolism to altered calcium homeostasis.^2,39^ At the same time, these findings should be interpreted with caution; they are associative and do not establish a direct calcium-dependent regulatory mechanism for SLC25A13 transport.^34^ Accordingly, the calcium-related and ion channel-related signals identified here should be regarded as biologically coherent ancillary observations rather than mechanistic conclusions.

Several limitations of this study should be acknowledged. First, because our extraction protocol captures proteins retained on filter paper, their cellular origin of the detected proteins could not be determined. Second, although DBS generally stabilizes blood components, including proteins, pre-analytical variables such as hematocrit, spot volume, storage temperature, humidity, and storage duration should be systematically evaluated in future studies. Third, although consumable costs are modest, untargeted mass spectrometry remains resource-intensive; once a minimal diagnostic protein set has been analytically and clinically validated, targeted proteomic platforms with fixed decision thresholds may facilitate scalable implementation. Fourth, the RNA-seq analysis in this study may have been affected by bias introduced by experimental design bias because the panel was enriched for genes related to inborn errors of metabolism, although substantial transcriptomic breadth was retained, with 4,456 genes detected. Finally, the calcium-related signals identified by enrichment analysis were derived from DBS proteomic data and PBMC transcriptomic data, neither of which directly reflects the liver, the principal tissue in which SLC25A13 functions. These observations therefore should not be overinterpreted as direct evidence of hepatic pathophysiology.

In conclusion, DBS-based proteomic profiling offers a promising complementary strategy for the diagnosis of citrin deficiency by providing a protein-level readout that can stratify patients beyond single-analyte measurement. Recent applications in inborn errors of immunity suggest that DBS proteomics can capture clinically meaningful protein-level disease signatures, supporting its potential extension to other inherited metabolic disorders.^40,41^ With rigorous external validation and further optimization toward targeted workflows, DBS-based proteomics may become a practical adjunct in clinical settings where rapid and biologically informative assessment is needed.

**Table 1.** Clinical information and genetic background of individuals with citrin deficiency in this study.

| Individuals | SS_1 | SS_2 | SS_3 | SS_4 | SS_5 | SS_6 | SS_7 | SS_8 | SS_9 | SS_10 |
| --- | --- | --- | --- | --- | --- | --- | --- | --- | --- | --- |
| Age | 36 y | 21 y | 12 y | 4 y | 22 y | 9 y | 24 y | 4 y | 6 y | 9 y |
| Sex | woman | woman | boy | girl | woman | boy | woman | girl | boy | boy |
| Weight (kg) (SD) | 54.2 | 46.0 | 47.2 (0.33) | 15.6 (0.10) | 48.0 | 26.7 (-0.44) | 60.7 | 17.5 (0.33) | 20.6 (-0.02) | 25.3 (-0.79) |
| Height (cm) (SD) | 152.6 | 152.6 | 165.7 (1.52) | 100.2 (-0.10) | 155.8 | 128.6 (-0.64) | 159.3 | 100.7 (-1.05) | 113.4 (-0.55) | 133.1 (0.15) |
| Body Mass Index (kg/m<sup>2</sup>) | 23.3 | 19.8 | 17.2 | 15.5 | 19.8 | 16.1 | 23.9 | 17.3 | 16.0 | 14.3 |
| Birth week | 38 | 39 | 40 | 41 | 37 | 38 | 40 | 40 | 39 | 39 |
| Birth weight (g) | 2674 | 2560 | 2680 | 3245 | 2460 | 2570 | 2840 | 2732 | 2584 | 2446 |
| Birth height (cm) | NA | 46.7 | 50.0 | 49.0 | 48.0 | 49.5 | 49.0 | 48.0 | 46.5 | 47.0 |
| Age at diagnosis | 13 y | 2 m | 3 m | 55 d | 3 m | 56 d | 1 y 3 m | 57 d | 18 d | 17 d |
| Genotype |  |  |  |  |  |  |  |  |  |  |
| SLC25A13 mutation | c.[1177+1G>A];<br>[1177+1G>A] | c.[1177+1G>A];<br>[1177+1G>A] | c.852_855del(;)<br>1311+1G>A | c.852_855del(;)<br>1177+1G>A | c.[852_855del];<br>[852_855del] | c.[852_855del];<br>[852_855del] | c.852_855del(;)<br>1177+1G>A | c.852_855del(;)<br>1177+1G>A | c.852_855del(;)<br>1799dup | c.1177+1G>A(;)<br>674C>A |
| Clinical information |  |  |  |  |  |  |  |  |  |  |
| NBS-positive | - | + | + | + | + | + | + | + | + | + |
| NICCD | + | + | + | + | + | + | + | + | + | + |
| FTTDCD | - | - | - | - | - | - | - | - | - | - |
| CTLN2 | - | - | - | - | - | - | - | - | - | - |
| Treatment |  |  |  |  |  |  |  |  |  |  |
| Protein-rich, lipid-rich, and low-carbohydrate diet | + | + | + | + | + | + | + | + | + | + |
| MCT supplementation | - | - | + | + | + | + | + | + | + | + |
| Medication for cholestasis | - | - | - | - | - | - | - | - | - | - |

NA, not available; y, years; m, months; d, days; NBS, newborn screening; NICCD, neonatal intrahepatic cholestasis by citrin deficiency; FTTDCD, failure to thrive and dyslipidemia by citrin deficiency; CTLN2, adult-onset type II citrullinemia; MCT, medium-chain triglyceride; \* Before the start of NBS.

| SS_11 | SS_12 | SS_13 | SS_14 | SS_15 | SS_16 | SM_1 | SM_2 | SM_3 | SM_4 | SM_5 | SM_6 | SM_7 |
| --- | --- | --- | --- | --- | --- | --- | --- | --- | --- | --- | --- | --- |
| 4 m | 11 y | 2 y | 1 y 8 m | 23 y | 76 y | 12 y | 10 y | 3 y | 6 y | 8 y | 14 y | 51 y |
| boy | boy | girl | girl | man | man | boy | boy | girl | boy | boy | boy | man |
| 7.09 (0.12) | 27.9 (-1.37) | 11.95 (1.0) | 8.3 (-2.13) | 62.9 | 51.8 | 45.1 (0.0) | 28.2 (-0.83) | 14.1 (0.05) | 20 (-0.16) | 34 (1.48) | 67.6 (1.10) | 55.1 |
| 63.0 (-0.51) | 135.7 (-0.98) | 81.7 (-1.0) | 73.7 (-2.67) | 184.7 | 162.3 | 155.8 (0.28) | 141 (0.46) | 98.3 (0.67) | 117.3 (0.35) | 134.7 (1.58) | 175.3 (1.42) | 162.8 |
| 17.9 | 15.2 | 17.9 | 15.3 | 18.4 | 19.7 | 18.6 | 14.2 | 14.6 | 14.5 | 18.7 | 22.0 | 20.8 |
| 36 | 39 | 40 | 37 | NA | NA | 40 | 38 | 40 | 38 | 41 | 39 | NA |
| 2588 | 2448 | 2615 | 1768 | NA | NA | 2635 | 2438 | 2.664 | 2964 | 3610 | 2580 | NA |
| 48.0 | 47.6 | 47 | 42.2 | NA | NA | 45 | 48 | 50 | 48.5 | 51 | 45 | NA |
| 35 d | 14 d | 3 m | 2 m | 1 m | 62 y | 2 m | 1 y 0 m | 5 m | 1 y 2 m | 3 y | 2 m | 34 y |
| c.1638_1660dup(;) 1177+1G>A | c.852_855del(;) IVS16ins3kb | c.852_855del(;) 1177+1G>A | c.852_855del(;) 1177+1G>A | c.1801G>A(;) 1638_1660dup | c.674C>A(;) 1478A>G | c.852_855del(;) 1178G>T | c.852_855del(;) 701G>C | c.852_855del(;) 701G>C | c.1177+1G>A(;) 1592G>A | c.1177+1G>A(;) 1592G>A | c.1177+1G>A(;) 1801G>T | c.674C>A(;) 1645C>T |
| + | + | - | + | + | * | + | + | - | - | - | - | * |
| + | + | + | - | - | NA | + | + | - | + | - | + | + |
| - | - | - | NA | - | NA | - | - | NA | - | - | - | - |
| - | - | - | NA | - | + | - | - | NA | NA | NA | - | + |
| + | + | - | NA | + | + | + | - | - | + | + | + | - |
| + | + | - | + | - | + | + | + | + | + | + | + | + |
| - | - | - | - | - | - | - | - | - | - | - | - | - |

erides; SS, loss-of-function/loss-of-function; SM, loss-of-function/missense.
CGCCCCCC.

## List of abbreviations

ACN: acetonitrile
AUC: area under the receiver operating characteristic curve
CTLN2: adult-onset type II citrullinemia
DBS: dried blood spots
DIA LC-MS/MS: data-independent acquisition liquid chromatography-tandem mass spectrometry
LoF: loss-of-function
LOOCV: leave-one-out cross-validation
Mis: missense
NANDA: non-targeted analysis of non-specifically DBS-absorbed proteins
NICCD: neonatal intrahepatic cholestasis caused by citrin deficiency
PBMCs: peripheral blood mononuclear cells
Ref: reference
RNA-seq: RNA sequencing
sPLS-DA: sparse partial least squares discriminant analysis
TFA: trifluoroacetic acid
VST: variance-stabilizing transformation

## Declarations

### Ethics approval and consent to participate

This study was approved by the Ethics Committee of the Faculty of Medicine in Tohoku University (Approval number: 2023-1-390, 2025-1-257). Written informed consent was obtained from the participants or their parents. Residual DBS samples were collected in accordance with an opt-out consent procedure.

Y.W. used ChatGPT 5.5 to improve the readability and clarity of the manuscript and to assist in organizing the analytical code for RNA-seq and proteomic analyses. After using this tool, the authors reviewed and edited the content as needed and take full responsibility for the the final content of the manuscript.

### Consent for publication

Not applicable.

### Availability of data and materials

Raw RNA-seq data are not publicly available because of informed consent restrictions and the potential risk of re-identification in this rare disease cohort. Access to de-identified processed data, including processed RNA-seq data, may be considered by the corresponding author upon reasonable request, subject to approval by the relevant ethics committee and completion of an appropriate data use agreement.

### Competing interests

The authors declare no competing interests.

### Funding

This work was supported in part by the institutional research funds of the SiRIUS Institute of Medical Research, Tohoku University, to Y.W., JSPS KAKENHI (Grant number: 24K11013) to D.N., the Kazusa DNA Research Institute Foundation to D.N., R.K, O.O, and Y.K., JST Moonshot R&D Program (Grant number JPMJMS2023) to J.T., and a Grant-in-Aid for Practical Research Project for Rare/Intractable Diseases from the AMED (Grant number: 26ek0109873s1001) to O.O. and T.H.

### Authors’ contributions

E.T., Y.-M.S., N.-A.I., H.N., H.Y., T.I., H.S., and C.N. collected specimens and clinical information. D.N., R.K., and Y.K. performed the proteomic analysis. O.O. conducted RNA-seq. E.T., M.S., J.T., and Y.W. analyzed the data. E.T. and Y.W. drafted the initial manuscript. Y.K., O.O., and Y.W. supervised the study. All authors participated in the study design, reviewed and revised the manuscript, approved the final manuscript, and agreed to be accountable for all aspects of the work.

## Acknowledgments

We thank all patients and their family members who participated in this study. We acknowledge the clinical personnel at each collaborating institution for their dedicated patient care and assistance with sample collection. We are grateful to Tomoko Kasanuki (The Jikei University School of Medicine, Tokyo, Japan) and Kumi Ito and Yoko Chiba (Department of Pediatrics, Tohoku University Graduate School of Medicine, Sendai, Japan) for their excellent technical support. We also acknowledge the Kazusa DNA Research Institute for help with RNA-seq and DBS proteome analyses. We thank the Biomedical Research Core of Tohoku University Graduate School of Medicine and the Biomedical Research Unit of Tohoku University Hospital for technical assistance.

## Notes

### Competing Interest Statement

The authors have declared no competing interest.

### Author Declarations

This study was approved by the Ethics Committee of the Faculty of Medicine in Tohoku University (Approval number: 2023-1-390, 2025-1-257).

