## Supplemental Figure for "Dried blood spot proteomics as a diagnostic framework for citrin deficiency"

### Supplementary Figures

**Figure S1.** Differential proteomic profile across *SLC25A13* genotype groups

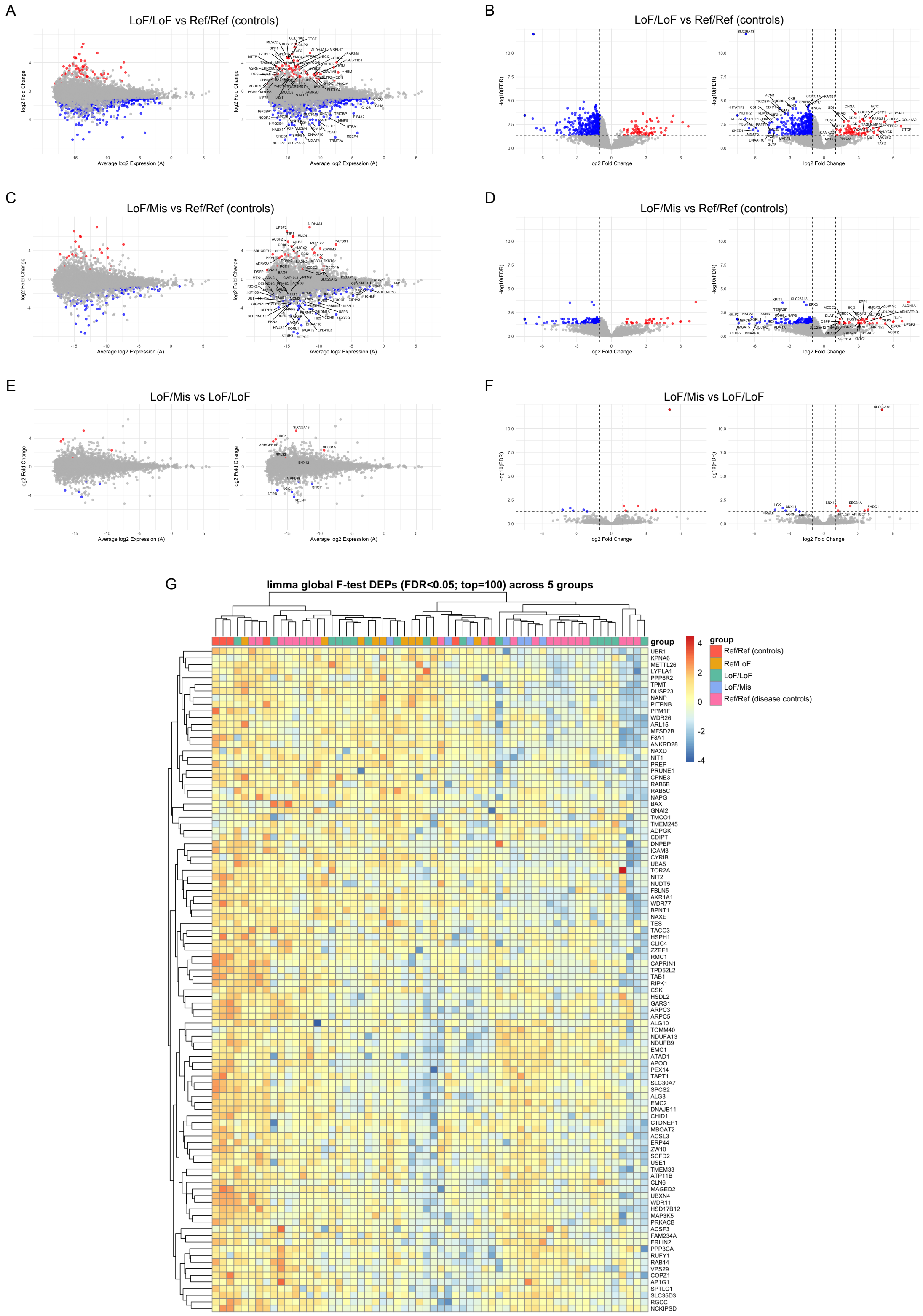

**A, B.** Proteomic differential expression analysis comparing individuals with LoF/LoF genotypes and Ref/Ref controls. **A.** MA plot showing the relationship between average protein abundance and log2 fold change. The y-axis represents log2 fold change, and the x-axis represents average log2 expression across samples. **B.** Volcano plot showing statistical significance versus effect size for the same comparison. The x-axis represents log2 fold change, and the y-axis represents −log10 adjusted *P* value. Each point represents one protein. Proteins meeting the significance threshold, defined as adjusted *P* value < 0.05 and |log2 fold change| ≥ 1, are shown in red for proteins upregulated in individuals with LoF/LoF genotypes and in blue for proteins downregulated in individuals with LoF/LoF genotypes.

**C, D.** Proteomic differential expression analysis comparing individuals with LoF/Mis genotypes and Ref/Ref controls. **C.** MA plot and **D.** volcano plot are presented as described for panels A and B.

**E, F.** Proteomic differential expression analysis comparing individuals with LoF/Mis genotypes and those with LoF/LoF genotypes. **E.** MA plot and **F.** volcano plot are presented as described for panels A and B.

**G.** Heatmap of the top 100 DEPs identified by the limma global F-test (FDR < 0.05) across five groups. Group colors indicate Ref/Ref controls in red, Ref/LoF in orange, LoF/LoF in green, LoF/Mis in blue, and Ref/Ref disease controls in pink. Protein expression levels are shown as standardized log-intensities, expressed as z-scores across samples.

LoF, loss-of-function; Mis, missense; Ref, reference; DEPs, differentially expressed proteins; FDR, false discovery rate.

**Figure S2.** Clinical laboratory parameters in individuals with LoF/LoF and LoF/Mis genotypes

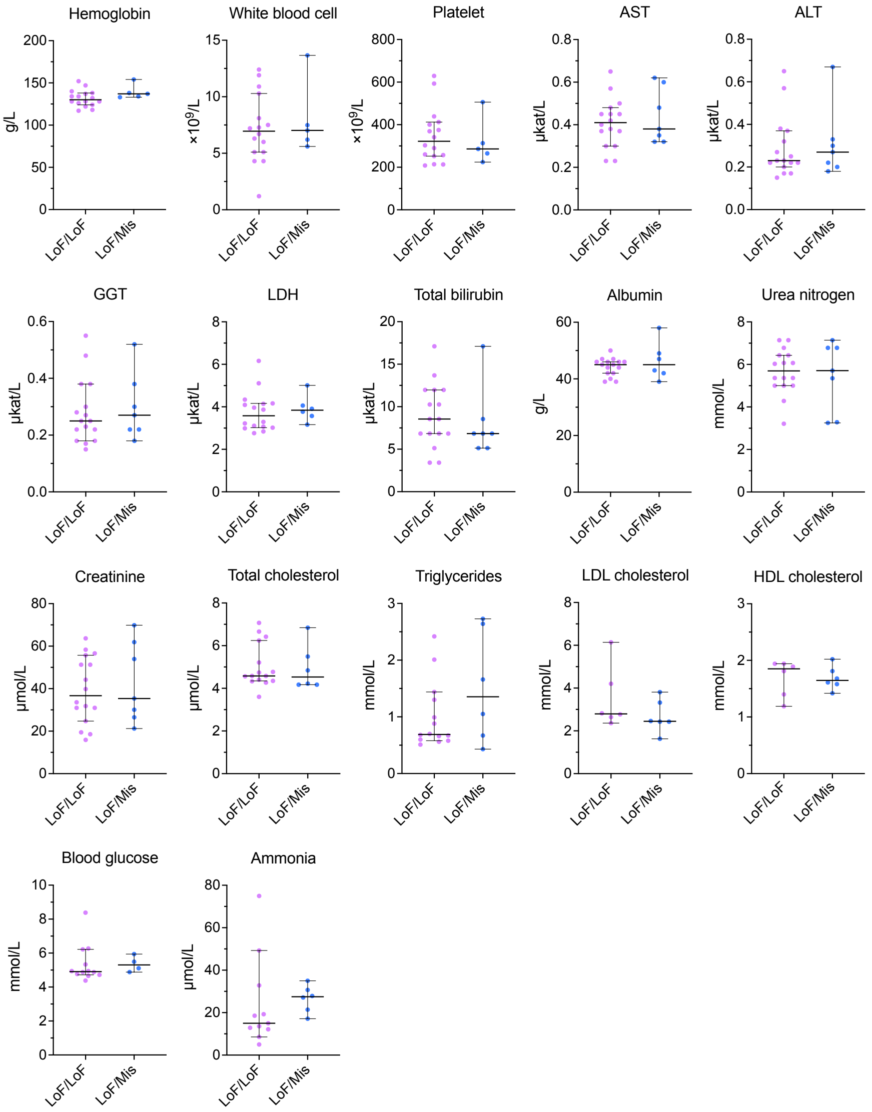


Laboratory parameters are shown as dot plots for individuals with LoF/LoF and LoF/Mis genotypes. Each point represents one individual. Horizontal lines indicate medians with 95% confidence interval. Statistical comparisons between groups were performed using Welch's *t*-test. No statistically significant differences were observed between the two groups. LoF, loss-of-function; Mis, missense; AST, aspartate aminotransferase; ALT, alanine aminotransferase; GGT, γ-glutamyltransferase; LDH, lactate dehydrogenase; LDL, low-density lipoprotein; HDL, high-density lipoprotein.

**Figure S3.** Plasma amino acid levels in individuals with LoF/LoF and LoF/Mis genotypes

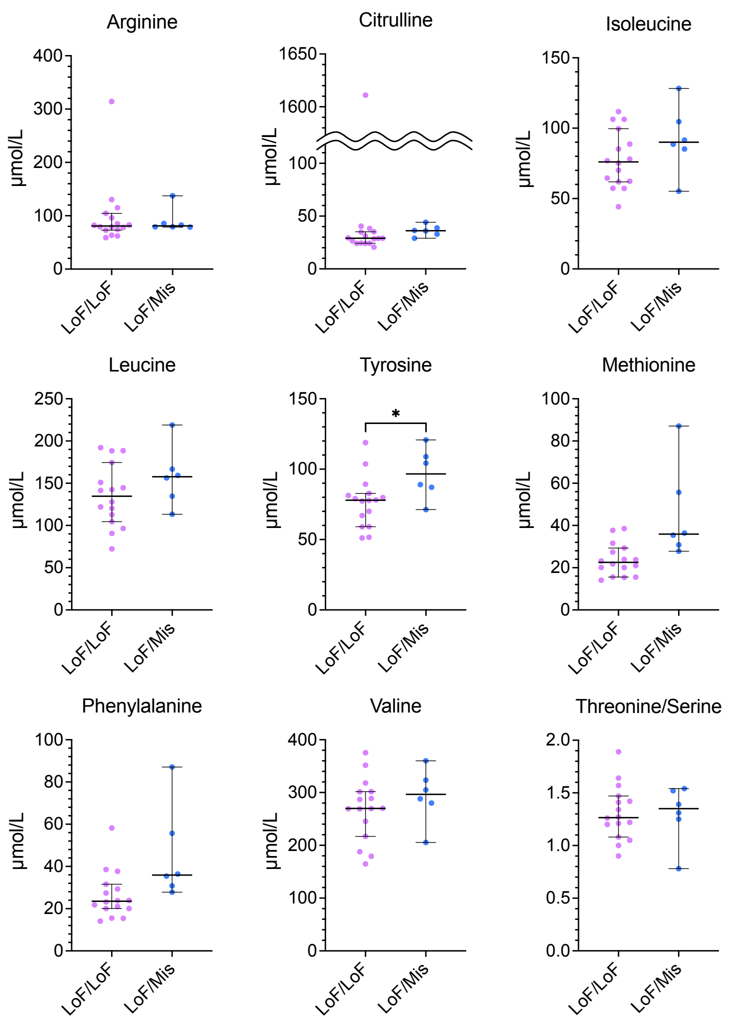


Plasma amino acid levels are shown as dot plots for individuals with LoF/LoF and LoF/Mis genotypes. Each point represents one individual. Horizontal lines indicate medians with 95% confidence intervals. Statistical comparisons between groups were performed using Welch's *t*-test; * *P* value < 0.05.

LoF, loss-of-function; Mis, missense.

**Figure S4.** Gene Ontology enrichment analysis of differentially expressed proteins across *SLC25A13* genotype comparisons

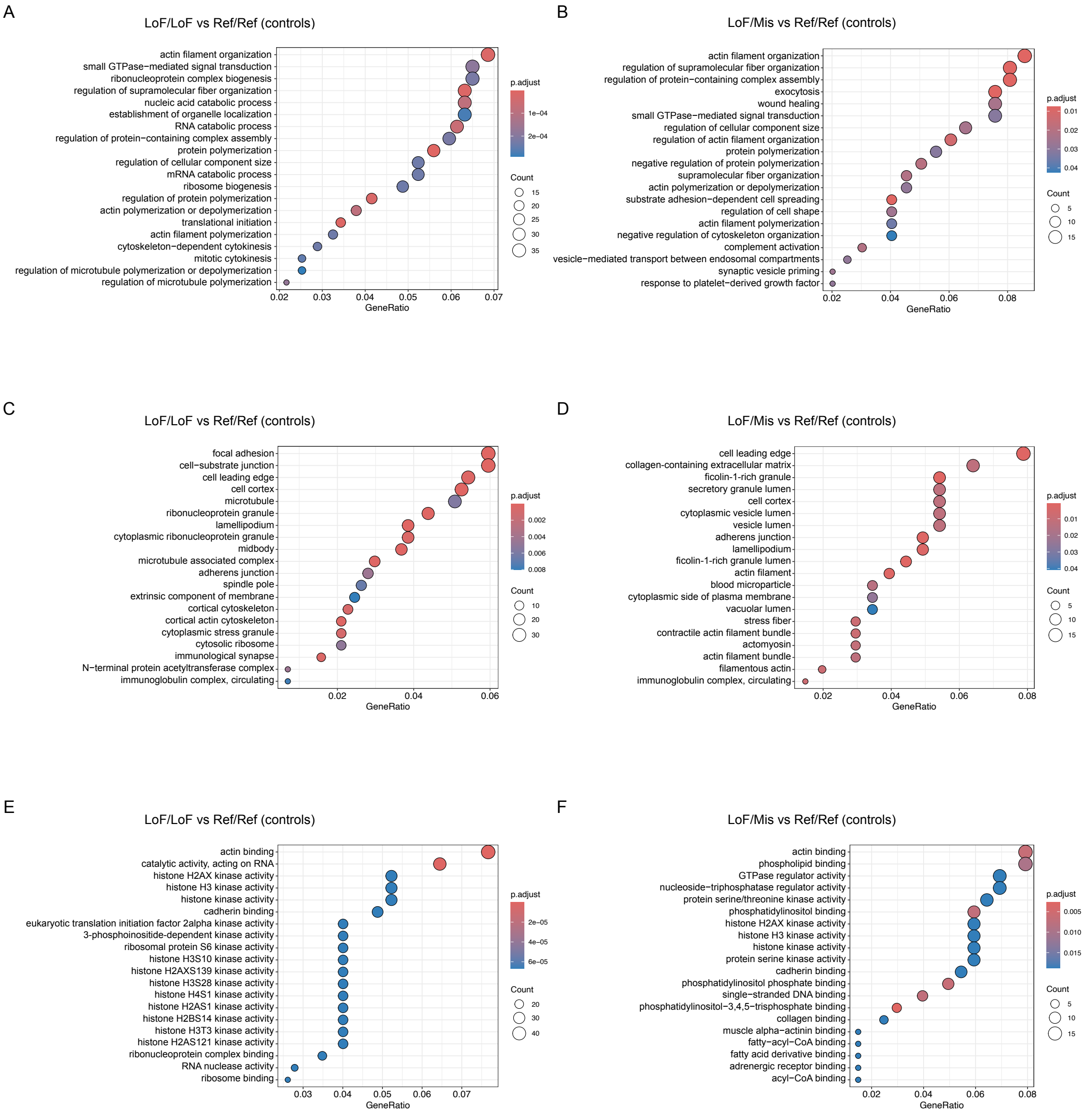


The x-axis represents the GeneRatio, and each point represents one enriched GO term. Point size is proportional to the number of counting genes, and point color indicates statistical significance based on adjusted *P* value. The y-axis lists enriched GO terms. **A, B.** GO biological process enrichment. **C, D.** GO cellular component enrichment. **E, F.** GO molecular function enrichment. **A, C, E.** Comparison of individuals with LoF/LoF genotypes versus Ref/Ref controls. **B, D, F.** Comparison of individuals with LoF/Mis genotypes versus Ref/Ref controls.

LoF, loss-of-function; Mis, missense; Ref, reference; GO, Gene Ontology.

**Figure S5.** Gene Ontology and Reactome enrichment analysis of RNA-seq differential expression signatures

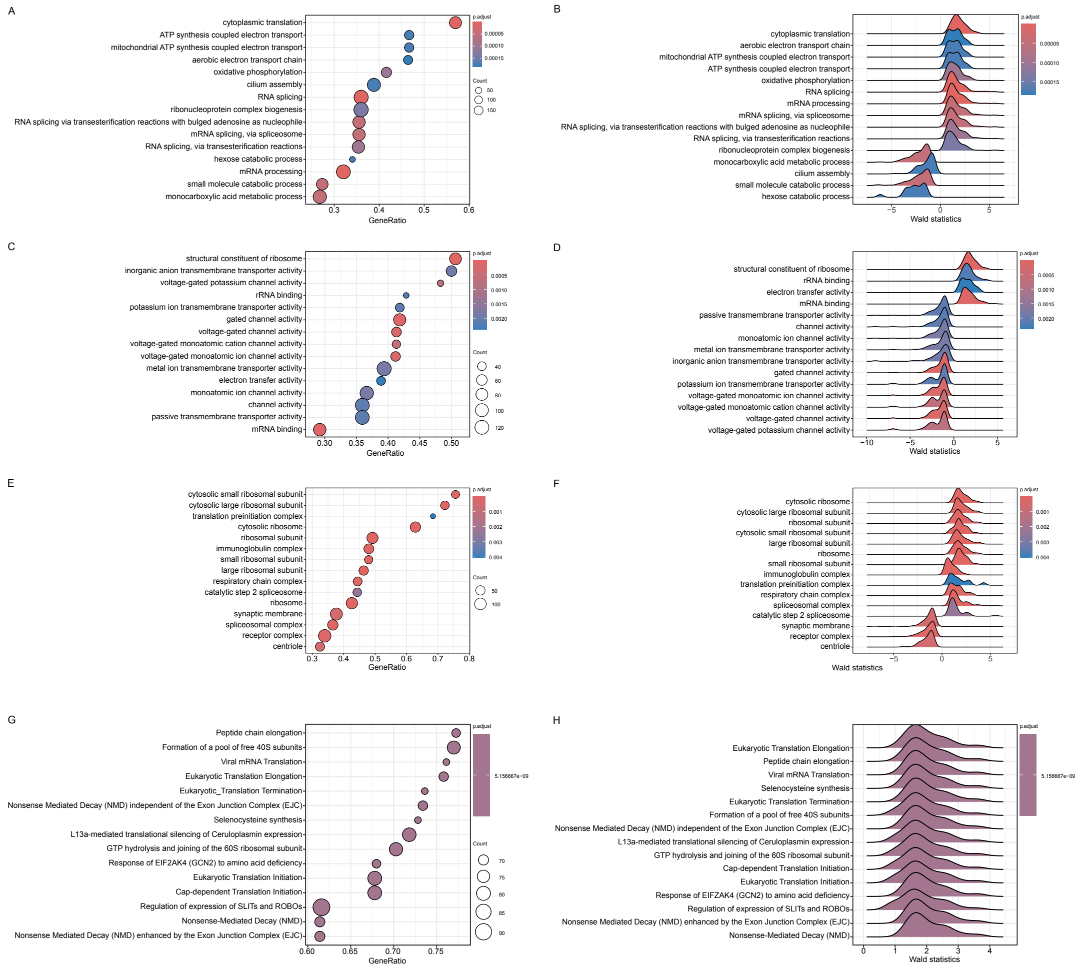


**A, B.** GO biological process enrichment analysis based on ranked gene lists derived from RNA-seq differential expression analysis.

**A.** Dot plot showing enriched GO biological process terms. The x-axis represents the GeneRatio, and each point represents one enriched GO term. Point size is proportional to the number of contributing genes, and point color indicates statistical significance based on the adjusted *P* value. **B.** Ridge plot showing the distribution of gene-level Wald statistics for genes within each enriched GO biological process term. Each curve corresponds to one enriched GO term, and color indicates statistical significance based on the adjusted *P* value.

**C, D.** GO molecular function enrichment analysis based on ranked gene lists derived from RNA-seq analysis. **C.** Dot plot and **D.** ridge plot are presented as described for panels A and B.

**E, F.** GO cellular component enrichment analysis based on ranked gene lists derived from RNA-seq analysis. **E.** Dot plot and **F.** ridge plot are presented as described for panels A and B.

**G, H.** Reactome pathway enrichment analysis based on ranked gene lists derived from RNA-seq differential expression analysis. **G.** Dot plot showing enriched Reactome pathways. **H.** Ridge plot showing the distribution of gene-level Wald statistics for genes within each enriched Reactome pathway. Plot formats are presented as described for panels A and B.

GO, Gene Ontology.
