## Supplemental Table for "Dried blood spot proteomics as a diagnostic framework for citrin deficiency"

Table S1. Laboratory data and clinincal manifestation of individuals with citrin deficiency in this study.

| Individuals | SS_1 | SS_2 | SS_3 | SS_4 | SS_5 | SS_6 | SS_7 | SS_8 | SS_9 | SS_10 | SS_11 | SS_12 | SS_13 | SS_14 | SS_15 | SS_16 | SM_1 | SM_2 | SM_3 | SM_4 | SM_5 | SM_6 | SM_7 |
|---|---|---|---|---|---|---|---|---|---|---|---|---|---|---|---|---|---|---|---|---|---|---|---|
| Plasma amino acids analysis | | | | | | | | | | | | | | | | | | | | | | | |
| Arginine (μmol/L) | 62.1 | 115.1 | 104.6 | 63.5 | 130.4 | 78.9 | 82.2 | 58.9 | 79.4 | 78 | 74.3 | 82.8 | 84.9 | 96.2 | 85.4 | 79.3 | 137.5 | NA | 78.7 | 82.5 | 79.3 | 73.1 | 314.3 |
| Citrulline (μmol/L) | 40.5 | 28.5 | 26.5 | 31.0 | 24.2 | 29.1 | 29.4 | 35.2 | 20.6 | 34.8 | 24.3 | 38.3 | 24.5 | 28.9 | 44.2 | 38.8 | 35.9 | NA | 29 | 36.4 | 32.9 | 24.2 | 1611 |
| Isoleucine (μmol/L) | 88.7 | 99.6 | 106.3 | 62.3 | 78.0 | 44.2 | 70.1 | 75.2 | 106.3 | 111.8 | 61.9 | 64.5 | 85.2 | 57.3 | 104.6 | 88.6 | 128.2 | NA | 55.2 | 91.5 | 85.2 | 76.9 | 57.2 |
| Leucine (μmol/L) | 150.9 | 188.5 | 192.2 | 112.7 | 141.5 | 72.3 | 122.0 | 127.8 | 174.5 | 188.4 | 96.4 | 120.2 | 144.6 | 104.5 | 166.8 | 159.2 | 218.9 | NA | 113.3 | 156.3 | 134.6 | 142.4 | 90.6 |
| Tyrosine (μmol/L) | 51.1 | 82.8 | 89.3 | 77.4 | 79.8 | 78.1 | 59.0 | 59.1 | 103.7 | 79 | 81.3 | 70 | 77.8 | 118.9 | 89 | 104.3 | 120.8 | NA | 71.3 | 108.9 | 87.1 | 51.6 | 67 |
| Methionine (μmol/L) | 20.1 | 37.7 | 29.3 | 14.1 | 27.4 | 15.4 | 21.0 | 15.5 | 38.5 | 23.2 | 23.8 | 20.1 | 31.6 | 21.8 | 27.8 | 35.4 | 55.7 | NA | 30.8 | 36.4 | 29.4 | 23.9 | 15.6 |
| Phenylalanine (μmol/L) | 56 | 82.5 | 81 | 55.9 | 94.3 | 48.7 | 59.1 | 56.5 | 86.1 | 68.1 | 57.9 | 69.9 | 71.3 | 78.2 | 78.6 | 71.9 | 93.6 | NA | 62.3 | 89.6 | 70.2 | 74.6 | 58.2 |
| Valine (μmol/L) | 286.9 | 351.6 | 301.6 | 245.5 | 269.3 | 164.7 | 269.7 | 288.7 | 318.2 | 375.5 | 179.3 | 269 | 301.4 | 217.1 | 304.9 | 288 | 360 | NA | 205.5 | 323.5 | 280 | 270 | 187.9 |
| Threonine/Serine | 1.89 | 1.64 | 1.34 | 1.20 | 1.22 | 1.57 | 1.41 | 1.00 | 1.27 | 0.90 | 1.47 | 1.05 | 1.21 | 1.42 | 1.39 | 1.52 | 1.54 | NA | 0.78 | 1.31 | 1.25 | 1.08 | 1.26 |
| CBC | | | | | | | | | | | | | | | | | | | | | | | |
| Hemoglobin (g/L) | 128.0 | 128.0 | 147.0 | 117.0 | 132.0 | 134.0 | 140.0 | 128.0 | 124.0 | 138.0 | 126.0 | 137.0 | 122.0 | 118.0 | 154.0 | NA | 138.0 | 134.0 | 133.0 | 137.0 | NA | 152.0 | 134.0 |
| White blood cell ($\times 10^9$/L) | 5.10 | 5.10 | 8.10 | 7.20 | 4.30 | 7.50 | 6.00 | 10.90 | 6.30 | 6.70 | 12.40 | 4.30 | 1.20 | 10.29 | 5.60 | NA | 6.20 | 13.66 | 7.02 | 7.49 | NA | 7.28 | 11.9 |
| Platelet ($\times 10^9$/L) | 213 | 341 | 257 | 412 | 369 | 400 | 290 | 439 | 375 | 252 | 629 | 260 | 303 | 593 | 224 | NA | 313 | 286 | 265 | 506 | NA | 214 | 208 |
| Chemistry | | | | | | | | | | | | | | | | | | | | | | | |
| Aspartate aminotransferase (μkat/L) | 0.23 | 0.30 | 0.30 | 0.40 | 0.23 | 0.50 | 0.45 | 0.37 | 0.48 | 0.38 | 0.65 | 0.45 | 0.45 | 0.57 | 0.32 | 0.62 | 0.32 | 0.48 | 0.6 | 0.38 | 0.35 | 0.37 | 0.42 |
| Alanine aminotransferase (μkat/L) | 0.20 | 0.17 | 0.23 | 0.17 | 0.15 | 0.25 | 0.65 | 0.22 | 0.37 | 0.22 | 0.38 | 0.27 | 0.23 | 0.22 | 0.22 | 0.67 | 0.18 | 0.27 | 0.2 | 0.33 | 0.3 | 0.32 | 0.57 |
| γ-Glutamyltransferase (μkat/L) | 0.22 | 0.15 | 0.22 | 0.28 | 0.30 | 0.25 | 0.48 | 0.18 | 0.25 | 0.18 | 0.55 | 0.27 | 0.17 | 0.23 | 0.22 | 0.52 | 0.38 | 0.27 | 0.18 | 0.3 | 0.22 | 0.38 | 0.38 |
| Lactate dehydrogenase (μkat/L) | 2.76 | 2.99 | 3.11 | 3.87 | 3.29 | 6.16 | 3.02 | 4.34 | 4.09 | 3.07 | 5.11 | 3.96 | 3.22 | 4.16 | NA | 3.91 | 3.16 | 4.06 | 5.01 | 3.77 | 3.57 | 2.84 | 4.14 |
| Total bilirubin (μmol/L) | 17.10 | 10.26 | 13.68 | 8.55 | 11.97 | 8.55 | 11.97 | 6.84 | 5.13 | 10.26 | 3.42 | 6.84 | 3.42 | 6.84 | 5.13 | 17.1 | 8.55 | 6.84 | 5.13 | 6.84 | 6.84 | 11.97 | 6.84 |
| Direct bilirubin (μmol/L) | NA | < 1.71 | NA | < 1.71 | 1.71 | < 1.71 | NA | < 1.71 | < 1.71 | < 1.71 | 1.71 | < 1.71 | 1.71 | NA | 1.71 | NA | 1.03 | 1.71 | 1.71 | 1.71 | 1.71 | 1.71 | NA |
| Total bile acids (μmol/L) | NA | NA | NA | NA | NA | NA | NA | NA | NA | NA | NA | NA | NA | NA | 13.5 | 4.6 | 12.3 | 5.2 | 6.1 | 9.4 | 6.4 | 2.3 | NA |
| Albumin (g/L) | 42.0 | 40.0 | 47.0 | 45.0 | 44.0 | 50.0 | 46.0 | 45.0 | 47.0 | 46.0 | 46.0 | 46.0 | 42.0 | 39.0 | NA | 39.0 | 58.0 | 47.0 | 49.0 | 43.0 | 42.0 | 44.0 | 39.0 |
| Urea nitrogen (mmol/L) | 7.14 | 6.07 | 5.36 | 4.28 | 5.00 | 5.36 | 5.00 | 5.36 | 6.43 | 6.43 | 3.21 | 7.14 | 6.03 | 6.78 | 6.78 | 6.78 | 5.71 | 3.25 | 3.28 | 7.14 | 5.35 | 5.00 | 6.07 |
| Creatinine (μmol/L) | 63.65 | 55.69 | 56.58 | 18.56 | 51.27 | 33.59 | 44.20 | 19.45 | 30.94 | 30.94 | 15.91 | 39.78 | 31.82 | 24.75 | 69.84 | 61.88 | 53.92 | 30.06 | 26.52 | 21.22 | 35.36 | 58.34 | 51.27 |
| Total cholesterol (mmol/L) | 7.07 | 5.21 | 4.40 | 4.27 | 4.58 | 4.74 | 6.66 | 6.24 | 4.58 | 6.42 | 4.77 | 4.35 | NA | 3.6 | 6.84 | 5.49 | 4.17 | NA | 4.22 | 4.84 | 4.17 | 4.33 | 4.56 |
| Triglycerides (mmol/L) | 0.68 | 1.44 | 0.99 | 2.01 | 0.88 | 0.67 | 0.70 | 0.66 | 0.60 | 0.58 | 2.42 | 0.51 | NA | NA | 2.64 | 1.05 | 0.67 | NA | 0.43 | 2.73 | 1.66 | 1.30 | 0.56 |
| Low-density cholesterol (mmol/L) | NA | 2.77 | NA | NA | 2.82 | NA | 4.20 | NA | NA | NA | NA | 6.14 | NA | NA | 3.81 | 3.32 | 1.63 | NA | 2.43 | 2.46 | 2.43 | 2.64 | 2.36 |
| High-density cholesterol (mmol/L) | 1.94 | 1.94 | NA | NA | 1.40 | NA | 1.81 | NA | NA | NA | NA | NA | NA | NA | 1.81 | 1.68 | 2.02 | NA | 1.58 | 1.61 | 1.42 | 1.19 | 1.89 |
| Blood glucose (mmol/L) | 4.38 | 4.72 | 4.94 | 4.66 | NA | 4.94 | 4.77 | NA | 6.27 | NA | 6.22 | 5.33 | NA | 4.88 | 4.88 | NA | 5.94 | NA | NA | 5.11 | 5.49 | NA | 8.38 |
| Ammonia (μmol/L) | 5.00 | 12.14 | 18.56 | NA | 14.99 | 8.57 | Low | NA | NA | 12.85 | 74.97 | 13.57 | 32.84 | NA | 30.70 | 17.14 | NA | 21.42 | 27.85 | 27.13 | 34.99 | 19.28 | 49.27 |
| Lactate (mmol/L) | NA | NA | NA | NA | NA | NA | NA | NA | NA | NA | NA | NA | 2.02 | NA | NA | NA | 1.89 | NA | NA | NA | NA | NA | NA |
| Pyruvate (μmol/L) | NA | NA | NA | NA | NA | NA | NA | NA | NA | NA | NA | NA | 136.27 | NA | NA | NA | 102.20 | NA | NA | NA | NA | NA | NA |
| Prothrombin time (%) | NA | 123.2 | NA | NA | NA | NA | NA | NA | NA | NA | NA | NA | NA | NA | 100 < | NA | NA | 98 | NA | NA | NA | NA | NA |
| Activated partial thromboplastin time (sec) | NA | 25.0 | NA | NA | NA | NA | NA | NA | NA | NA | NA | NA | NA | NA | 26.3 | NA | NA | 30 | NA | NA | NA | NA | NA |
| Fibrinogen (g/L) | NA | 3.24 | NA | NA | NA | NA | NA | NA | NA | NA | NA | NA | NA | NA | NA | NA | NA | NA | NA | NA | NA | NA | NA |
| Ultrasound testing | | | | | | | | | | | | | | | | | | | | | | | |
| Fatty liver | - | - | NA | - | - | - | + | - | - | - | NA | - | NA | NA | - | - | - | NA | NA | - | - | - | - |
| Hepatomegaly | - | - | NA | - | - | - | - | - | - | - | NA | - | NA | NA | - | - | - | NA | NA | - | - | - | - |
| Clinical manifestation | | | | | | | | | | | | | | | | | | | | | | | |
| Food preferences | + | + | + | + | + | + | + | + | + | + | NA | + | - | NA | + | + | + | - | - | + | + | + | NA |
| Growth restriction | - | - | - | - | - | - | - | - | - | - | - | - | - | + | - | - | - | - | - | - | - | - | NA |
| Pancreatitis | - | - | - | - | - | - | - | - | - | - | - | - | - | - | - | - | - | - | - | - | - | - | NA |
| Fatigue | - | - | - | - | - | - | - | - | - | - | - | - | - | NA | - | - | - | - | - | + | + | - | NA |
| Anorexia | - | - | - | - | - | - | - | - | - | - | - | - | - | - | - | - | - | - | - | - | - | - | NA |

CBC, complete blood count; NA, not available; SS, loss-of-function/loss-of-function; SM, loss-of-function/missense.

Table S2. Character of disease controls.

| Individuals | DD_1 | DD_2 | DD_3 | DD_4 | DD_5 | DD_6 | DD_7 |
|---|---|---|---|---|---|---|---|
| Age | 8 y | 27 y | 40 y | 36 y | 3 y | 14 y | 3 y |
| Sex | girl | woman | woman | man | boy | boy | girl |
| Diagnosis | Fanconi-Bickel syndrome | Ornithine transcarbamylase deficiency | Ornithine transcarbamylase deficiency | Argininosuccinic aciduria | Galactosemia type III | Galactosemia type II | Galactosemia type II |
| Gene | *SLC2A2* | *OTC* | *OTC* | *ASL* | *GALE* | *GALK1* | *GALK1* |
| Mutations | NM_000340.2: c.[682C>T];[682C>T] | del(X)(p11.4p21.1) | NM_000531.6:c.533C>T | NM_000048.4: c.[347A>G];[637C>T] | NM_001008216.2: c.308A>G(;)788G>A | NM_000154.2: c.401C>A(;)830G>A | NM_000154.2: c.1016A>C(;)(793+1_794-1)_(944+1_945-1)del |

| Individuals | DD_8 | DD_9 | DD_10 | DD_11 | DD_12 | DD_13 | DD_14 |
|---|---|---|---|---|---|---|---|
| Age | 16 y | 13 y | 9 y | 8 y | 8 y | 4 y | 5 y |
| Sex | woman | boy | boy | boy | boy | girl | girl |
| Diagnosis | Galactosemia type III | Galactosemia type III | Galactosemia type IV | Galactosemia type IV | Galactosemia type IV | Galactosemia type I | Galactosemia type I |
| Gene | *GALE* | *GALE* | *GALM* | *GALM* | *GALM* | *GALT* | *GALT* |
| Mutations | NM_001008216.2: c.715C>T(;)796-15T>C | NM_001008216.2: c.715C>T(;)796-15T>C | NM_138801.3: c.244C>T(;)294del | NM_138801.3: c.[424G>A];[424G>A] | NM_138801.3: c.[294del];[294del] | NM_000155.4: c.207C>A(;)328G>A | NM_000155.4: c.207C>A(;)784C>T |

| Individuals | DD_15 | DD_16 | DD_17 | DD_18 | DD_19 | DD_20 | DD_21 |
|---|---|---|---|---|---|---|---|
| Age | 1 y 6 m | 19 y | 16 y | 9 y | 26 d | 7 y | 45 y |
| Sex | girl | woman | woman | girl | woman | boy | woman |
| Diagnosis | Portosystemic shunt | Citrullinemia type I | Citrullinemia type I | Citrullinemia type I | Portosystemic shunt | Galactosemia type IV | Fanconi-Bickel syndrome |
| Gene | | *ASS1* | *ASS1* | *ASS1* | | *GALM* | *SLC2A2* |
| Mutations | | NM_054012.4: c.496-2A>G(;)814C>T | NM_054012.4: c.496-2A>G(;)814C>T | NM_054012.4: c.496-2A>G(;)814C>T | | NM_138801.3: c.294del(;)799C>G | NM_000340.2: c.[1268T>A];[1268T>A] |

Table S3. Proteomic quantification data.

| Number | # of Grouped | Accession n | Master acce | Master prote | Master Gene | Species | MW(kDa) | Peptides | Unique peptic | WW_1 | WW_2 | WW_3 | WW_4 | WW_5 | WW_6 | WS_1 |
|---|---|---|---|---|---|---|---|---|---|---|---|---|---|---|---|---|
| 1 | 1 | P04114 | P04114 | Apolipoprotei | APOB | Homo sapien | 515.553 | 238 | 238 | 1.0E+10 | 5.7E+09 | 8.3E+09 | 1.3E+10 | 6.9E+09 | 1.6E+10 | 6.8E+09 |
| 2 | 1 | P02549 | P02549 | Spectrin alph | SPTA1 | Homo sapien | 280.019 | 154 | 154 | 1.0E+10 | 1.0E+10 | 8.5E+09 | 1.3E+10 | 1.3E+10 | 1.6E+10 | 1.7E+10 |
| 3 | 1 | Q15149 | Q15149 | Plectin | PLEC | Homo sapien | 531.798 | 157 | 152 | 1.3E+08 | 1.8E+08 | 1.3E+08 | 1.0E+08 | 2.9E+08 | 8.2E+07 | 1.1E+08 |
| 4 | 1 | Q9UPN3 | Q9UPN3 | Microtubule-a | MACF1 | Homo sapien | 838.323 | 153 | 151 | 5.3E+07 | 6.0E+07 | 1.2E+07 | 4.8E+07 | 5.8E+07 | 2.4E+07 | 7.9E+06 |
| 5 | 1 | Q8NF91 | Q8NF91 | Nesprin-1 | SYNE1 | Homo sapien | 1011.107 | 129 | 129 | 9.2E+06 | 1.6E+07 | 1.5E+07 | 7.1E+06 | 3.0E+07 | 1.4E+07 | 1.4E+07 |
| 6 | 1 | P11277 | P11277 | Spectrin beta | SPTB | Homo sapien | 246.47 | 128 | 121 | 9.8E+09 | 1.1E+10 | 8.8E+09 | 1.3E+10 | 1.3E+10 | 1.5E+10 | 1.6E+10 |
| 7 | 1 | Q14204 | Q14204 | Cytoplasmic c | DYNC1H1 | Homo sapien | 532.418 | 112 | 112 | 4.3E+07 | 4.2E+07 | 3.2E+07 | 2.1E+07 | 2.8E+07 | 3.2E+07 | 4.2E+07 |
| 8 | 1 | Q09666 | Q09666 | Neuroblast di | AHNAK | Homo sapien | 629.112 | 111 | 111 | 5.1E+07 | 5.0E+07 | 4.2E+07 | 4.0E+07 | 8.9E+07 | 4.4E+07 | 2.7E+07 |
| 9 | 1 | Q63HN8 | Q63HN8 | E3 ubiquitin-p | RNF213 | Homo sapien | 591.42 | 105 | 105 | 9.4E+06 | 5.5E+07 | 7.2E+07 | 3.1E+07 | 3.2E+07 | 1.8E+07 | 1.4E+07 |
| 10 | 1 | P35579 | P35579 | Myosin-9 | MYH9 | Homo sapien | 226.536 | 111 | 95 | 2.2E+09 | 2.7E+09 | 2.9E+09 | 2.3E+09 | 2.1E+09 | 2.2E+09 | 4.8E+09 |
| 11 | 1 | Q13813 | Q13813 | Spectrin alph | SPTAN1 | Homo sapien | 284.544 | 94 | 94 | 5.3E+07 | 8.1E+07 | 7.2E+07 | 6.7E+07 | 5.2E+07 | 5.7E+07 | 4.6E+07 |
| 12 | 1 | P21333 | P21333 | Filamin-A | FLNA | Homo sapien | 280.742 | 96 | 88 | 1.0E+09 | 1.1E+09 | 1.1E+09 | 1.0E+09 | 1.0E+09 | 8.5E+08 | 1.9E+09 |
| 13 | 1 | Q9Y490 | Q9Y490 | Talin-1 | TLN1 | Homo sapien | 269.773 | 99 | 87 | 2.9E+09 | 3.1E+09 | 2.8E+09 | 2.5E+09 | 2.2E+09 | 2.2E+09 | 2.8E+09 |
| 14 | 1 | Q5T4S7 | Q5T4S7 | E3 ubiquitin-p | UBR4 | Homo sapien | 573.853 | 85 | 85 | 4.6E+07 | 5.9E+07 | 5.2E+07 | 7.0E+07 | 5.0E+07 | 5.8E+07 | 1.8E+08 |
| 15 | 1 | P16157 | P16157 | Ankyrin-1 | ANK1 | Homo sapien | 206.27 | 84 | 79 | 6.2E+09 | 7.1E+09 | 6.2E+09 | 9.7E+09 | 8.7E+09 | 1.0E+10 | 9.1E+09 |
| 16 | 1 | P15924 | P15924 | Desmoplakin | DSP | Homo sapien | 331.78 | 78 | 78 | 8.3E+06 | 1.1E+07 | 7.6E+06 | 1.4E+07 | 1.2E+07 | 2.5E+07 | 1.1E+07 |
| 17 | 1 | Q01082 | Q01082 | Spectrin beta | SPTBN1 | Homo sapien | 274.614 | 80 | 78 | 7.4E+07 | 8.1E+07 | 8.1E+07 | 7.0E+07 | 7.5E+07 | 7.6E+07 | 5.9E+07 |
| 18 | 1 | Q709C8 | Q709C8 | Intermembran | VPS13C | Homo sapien | 422.398 | 78 | 78 | 2.4E+07 | 2.8E+07 | 3.2E+07 | 3.3E+07 | 3.1E+07 | 3.1E+07 | 2.7E+07 |
| 19 | 1 | P35580 | P35580 | Myosin-10 | MYH10 | Homo sapien | 229.002 | 79 | 74 | 1.8E+07 | 2.5E+07 | 3.0E+07 | 2.8E+07 | 2.6E+07 | 3.5E+07 | 2.4E+07 |
| 20 | 1 | P46939 | P46939 | Utrophin | UTRN | Homo sapien | 394.474 | 76 | 74 | 9.6E+06 | 1.0E+07 | 1.1E+07 | 8.4E+06 | 9.7E+06 | 1.1E+07 | 1.6E+07 |
| 21 | 1 | P01024 | P01024 | Complement | C3 | Homo sapien | 187.152 | 72 | 72 | 4.0E+09 | 3.5E+09 | 2.9E+09 | 3.4E+09 | 2.9E+09 | 2.9E+09 | 3.3E+09 |
| 22 | 1 | Q14789 | Q14789 | Golgin subfar | GOLGB1 | Homo sapien | 376.027 | 72 | 72 | 4.2E+07 | 3.3E+07 | 3.4E+07 | 3.7E+07 | 5.1E+07 | 2.3E+07 | 3.4E+07 |
| 23 | 1 | Q92614 | Q92614 | Unconvention | MYO18A | Homo sapien | 233.116 | 71 | 71 | 1.7E+08 | 1.5E+08 | 1.2E+08 | 1.6E+08 | 2.6E+08 | 1.9E+08 | 2.3E+08 |
| 24 | 1 | Q8WXH0 | Q8WXH0 | Nesprin-2 | SYNE2 | Homo sapien | 796.458 | 70 | 70 | 3.9E+06 | 8.1E+06 | 7.9E+06 | 4.1E+06 | 1.7E+07 | 2.1E+06 | 4.1E+06 |
| 25 | 1 | Q7Z6Z7 | Q7Z6Z7 | E3 ubiquitin-p | HUWE1 | Homo sapien | 481.899 | 65 | 65 | 8.8E+06 | 8.7E+06 | 9.0E+06 | 1.1E+07 | 9.1E+06 | 9.6E+06 | 2.2E+07 |
| 26 | 1 | Q96RL7 | Q96RL7 | Intermembran | VPS13A | Homo sapien | 360.285 | 63 | 63 | 4.1E+07 | 5.2E+07 | 3.8E+07 | 5.7E+07 | 4.6E+07 | 5.4E+07 | 4.5E+07 |
| 27 | 1 | P78527 | P78527 | DNA-depend | PRKDC | Homo sapien | 469.099 | 62 | 62 | 1.6E+07 | 2.1E+07 | 1.8E+07 | 2.2E+07 | 1.8E+07 | 1.5E+07 | 1.7E+07 |
| 28 | 1 | P42345 | P42345 | Serine/threon | MTOR | Homo sapien | 288.896 | 61 | 61 | 9.1E+06 | 1.0E+07 | 8.4E+06 | 1.7E+07 | 1.1E+07 | 1.0E+07 | 1.7E+07 |
| 29 | 1 | P02751 | P02751 | Fibronectin | FN1 | Homo sapien | 272.323 | 57 | 57 | 5.1E+09 | 5.3E+09 | 4.3E+09 | 6.5E+09 | 5.9E+09 | 6.5E+09 | 6.7E+09 |
| 30 | 1 | Q8N3C0 | Q8N3C0 | Activating sig | ASCC3 | Homo sapien | 251.464 | 57 | 57 | 1.7E+07 | 1.5E+07 | 1.1E+07 | 1.3E+07 | 1.9E+07 | 1.7E+07 | 1.3E+07 |
| 31 | 1 | P12111 | P12111 | Collagen alph | COL6A3 | Homo sapien | 343.671 | 56 | 56 | 1.4E+07 | 2.6E+07 | 1.5E+07 | 2.1E+07 | 2.1E+07 | 1.8E+07 | 3.0E+07 |
| 32 | 1 | Q14980 | Q14980 | Nuclear mitot | NUMA1 | Homo sapien | 238.263 | 56 | 56 | 8.8E+06 | 9.6E+06 | 1.2E+07 | 9.5E+06 | 1.4E+07 | 1.1E+07 | 1.5E+07 |
| 33 | 1 | O75643 | O75643 | U5 small nucl | SNRNP200 | Homo sapien | 244.511 | 55 | 55 | 6.2E+06 | 6.1E+06 | 6.9E+06 | 4.8E+06 | 9.3E+06 | 5.9E+06 | 4.8E+06 |
| 34 | 1 | Q14008 | Q14008 | Cytoskeleton | CKAP5 | Homo sapien | 225.499 | 55 | 55 | 1.0E+08 | 1.2E+08 | 8.9E+07 | 1.0E+08 | 9.9E+07 | 1.0E+08 | 1.1E+08 |
| 35 | 1 | Q9Y4D8 | Q9Y4D8 | Probable E3 u | HECTD4 | Homo sapien | 439.353 | 52 | 52 | 1.0E+07 | 1.4E+07 | 9.8E+06 | 1.6E+07 | 1.4E+07 | 2.3E+07 | 1.2E+07 |
| 36 | 1 | O75592 | O75592 | E3 ubiquitin-p | MYCBP2 | Homo sapien | 513.646 | 51 | 51 | 9.4E+06 | 1.2E+07 | 1.2E+07 | 1.0E+07 | 1.4E+07 | 1.2E+07 | 8.4E+06 |
| 37 | 1 | P01023 | P01023 | Alpha-2-macr | A2M | Homo sapien | 163.292 | 53 | 50 | 1.6E+09 | 1.7E+09 | 1.5E+09 | 1.9E+09 | 1.3E+09 | 1.2E+09 | 2.3E+09 |
| 38 | 1 | P12259 | P12259 | Coagulation f | F5 | Homo sapien | 251.709 | 50 | 50 | 3.5E+08 | 1.9E+08 | 2.3E+08 | 3.3E+08 | 3.7E+08 | 3.4E+08 | 1.9E+08 |
| 39 | 1 | Q9Y4G6 | Q9Y4G6 | Talin-2 | TLN2 | Homo sapien | 271.62 | 50 | 50 | 2.2E+07 | 2.7E+07 | 2.2E+07 | 1.9E+07 | 2.0E+07 | 2.4E+07 | 1.3E+07 |
| 40 | 1 | P22105 | P22105 | Tenascin-X | TNXB | Homo sapien | 458.395 | 53 | 49 | 2.1E+06 | 5.2E+06 | 3.0E+06 | 3.7E+06 | 4.1E+06 | 3.0E+06 | 3.5E+06 |
| 41 | 1 | Q92616 | Q92616 | Stalled riboso | GCN1 | Homo sapien | 292.713 | 49 | 49 | [illegible] | [illegible] | [illegible] | [illegible] | [illegible] | [illegible] | [illegible] |

| | | | | | | | | | | |
|---|---|---|---|---|---|---|---|---|---|---|
| 522 | 1 | Q9BSJ2 | Q9BSJ2 | Gamma-tubu | TUBGCP2 | Homo sapien | 102.536 | 18 | 18 | [illegible] |
| 523 | 1 | Q9C0E2 | Q9C0E2 | Exportin-4 | XPO4 | Homo sapien | 130.139 | 18 | 18 | [illegible] |
| 524 | 1 | Q9C0J8 | Q9C0J8 | pre-mRNA 3' | WDR33 | Homo sapien | 145.891 | 18 | 18 | [illegible] |
| 525 | 1 | Q9NRW7 | Q9NRW7 | Vacuolar pro | VPS45 | Homo sapien | 65.075 | 18 | 18 | [illegible] |
| 526 | 1 | Q9NRY4 | Q9NRY4 | Rho GTPase | ARHGAP35 | Homo sapien | 170.516 | 18 | 18 | [illegible] |
| 527 | 1 | Q9NRY5 | Q9NRY5 | Protein FAM1 | FAM114A2 | Homo sapien | 55.467 | 18 | 18 | [illegible] |
| 528 | 1 | Q9NVE7 | Q9NVE7 | 4'-phosphop | PANK4 | Homo sapien | 85.992 | 18 | 18 | [illegible] |
| 529 | 1 | Q9NVP1 | Q9NVP1 | ATP-depend | DDX18 | Homo sapien | 75.406 | 18 | 18 | [illegible] |
| 530 | 1 | Q9NZB2 | Q9NZB2 | Constitutive | FAM120A | Homo sapien | 121.888 | 19 | 18 | [illegible] |
| 531 | 1 | Q9P260 | Q9P260 | RAB11-bindi | RELCH | Homo sapien | 134.633 | 18 | 18 | [illegible] |
| 532 | 1 | Q9UHD8 | Q9UHD8 | Septin-9 | SEPTIN9 | Homo sapien | 65.402 | 18 | 18 | [illegible] |
| 533 | 1 | Q9UHP3 | Q9UHP3 | Ubiquitin car | USP25 | Homo sapien | 122.218 | 18 | 18 | [illegible] |
| 534 | 1 | Q9UI08 | Q9UI08 | Ena/VASP-lik | EVL | Homo sapien | 44.621 | 18 | 18 | [illegible] |
| 535 | 1 | Q9UKV3 | Q9UKV3 | Apoptotic ch | ACIN1 | Homo sapien | 151.864 | 18 | 18 | [illegible] |
| 536 | 1 | Q9ULH1 | Q9ULH1 | Arf-GAP with | ASAP1 | Homo sapien | 125.499 | 20 | 18 | [illegible] |
| 537 | 1 | Q9Y6D6 | Q9Y6D6 | Brefeldin A-in | ARFGEF1 | Homo sapien | 208.77 | 25 | 18 | [illegible] |
| 538 | 1 | A0FGR8 | A0FGR8 | Extended sy | ESYT2 | Homo sapien | 102.358 | 17 | 17 | [illegible] |
| 539 | 1 | O00231 | O00231 | 26S proteas | PSMD11 | Homo sapien | 47.464 | 17 | 17 | [illegible] |
| 540 | 1 | O00391 | O00391 | Sulfhydryl ox | QSOX1 | Homo sapien | 82.576 | 17 | 17 | [illegible] |
| 541 | 1 | O43143 | O43143 | ATP-depend | DHX15 | Homo sapien | 90.934 | 18 | 17 | [illegible] |
| 542 | 1 | O43290 | O43290 | U4/U6.U5 tri- | SART1 | Homo sapien | 90.255 | 17 | 17 | [illegible] |
| 543 | 1 | O75083 | O75083 | WD repeat-c | WDR1 | Homo sapien | 66.194 | 17 | 17 | [illegible] |
| 544 | 1 | O75691 | O75691 | Small subuni | UTP20 | Homo sapien | 318.389 | 17 | 17 | [illegible] |
| 545 | 1 | O75955 | O75955 | Flotillin-1 | FLOT1 | Homo sapien | 47.356 | 17 | 17 | [illegible] |
| 546 | 1 | P01008 | P01008 | Antithrombin- | SERPINC1 | Homo sapien | 52.604 | 17 | 17 | [illegible] |
| 547 | 1 | P07237 | P07237 | Protein disulf | P4HB | Homo sapien | 57.118 | 17 | 17 | [illegible] |
| 548 | 1 | P09327 | P09327 | Villin-1 | VIL1 | Homo sapien | 92.695 | 18 | 17 | [illegible] |
| 549 | 1 | P11215 | P11215 | Integrin alpha | ITGAM | Homo sapien | 127.182 | 17 | 17 | [illegible] |
| 550 | 1 | P13796 | P13796 | Plastin-2 | LCP1 | Homo sapien | 70.291 | 23 | 17 | [illegible] |
| 551 | 1 | P14868 | P14868 | Aspartate--tR | DARS1 | Homo sapien | 57.136 | 17 | 17 | [illegible] |
| 552 | 1 | P17858 | P17858 | ATP-depend | PFKL | Homo sapien | 85.018 | 18 | 17 | [illegible] |
| 553 | 1 | P23246 | P23246 | Splicing facto | SFPQ | Homo sapien | 76.151 | 17 | 17 | [illegible] |
| 554 | 1 | P28331 | P28331 | NADH-ubiqui | NDUFS1 | Homo sapien | 79.469 | 17 | 17 | [illegible] |
| 555 | 1 | P30041 | P30041 | Peroxiredoxi | PRDX6 | Homo sapien | 25.034 | 17 | 17 | [illegible] |
| 556 | 1 | P30101 | P30101 | Protein disulf | PDIA3 | Homo sapien | 56.783 | 17 | 17 | [illegible] |
| 557 | 1 | P30876 | P30876 | DNA-directe | POLR2B | Homo sapien | 133.897 | 17 | 17 | [illegible] |
| 558 | 1 | P38606 | P38606 | V-type proto | ATP6V1A | Homo sapien | 68.301 | 17 | 17 | [illegible] |
| 559 | 1 | P39880 | P39880 | Homeobox pr | CUX1 | Homo sapien | 164.19 | 18 | 17 | [illegible] |
| 560 | 1 | P43403 | P43403 | Tyrosine-pro | ZAP70 | Homo sapien | 69.875 | 17 | 17 | [illegible] |
| 561 | 1 | P46379 | P46379 | Large proline | BAG6 | Homo sapien | 119.41 | 17 | 17 | [illegible] |
| 562 | 1 | P48444 | P48444 | Coatomer su | ARCN1 | Homo sapien | 57.212 | 17 | 17 | [illegible] |
| 563 | 1 | P49588 | P49588 | Alanine--tRN | AARS1 | Homo sapien | 106.813 | 17 | 17 | [illegible] |
| 564 | 1 | P49748 | P49748 | Very long-ch | ACADVL | Homo sapien | 70.392 | 17 | 17 | [illegible] |
| 565 | 1 | P51608 | P51608 | Methyl-CpG- | MECP2 | Homo sapien | 52.441 | 17 | 17 | [illegible] |
| 566 | 1 | Q01804 | Q01804 | OTU domain- | OTUD4 | Homo sapien | 124.046 | 17 | 17 | [illegible] |
| 567 | 1 | Q02108 | Q02108 | Guanylate cy | GUCY1A1 | Homo sapien | 77.452 | 17 | 17 | [illegible] |
| 568 | 1 | Q05823 | Q05823 | 2-5A-depend | RNASEL | Homo sapien | 83.534 | 17 | 17 | [illegible] |
| 569 | 1 | Q06124 | Q06124 | Tyrosine-pro | PTPN11 | Homo sapien | 68.012 | 17 | 17 | [illegible] |
| 570 | 1 | Q08495 | Q08495 | Dematin | DMTN | Homo sapien | 45.516 | 17 | 17 | [illegible] |
| 571 | 1 | Q0ZGT2 | Q0ZGT2 | Nexilin | NEXN | Homo sapien | 80.661 | 17 | 17 | [illegible] |
| 572 | 1 | Q13435 | Q13435 | Splicing facto | SF3B2 | Homo sapien | 100.23 | 17 | 17 | [illegible] |
| 573 | 1 | Q14554 | Q14554 | Protein disulf | PDIA5 | Homo sapien | 59.595 | 17 | 17 | [illegible] |
| 574 | 1 | Q14667 | Q14667 | Bridge-like lip | BLTP2 | Homo sapien | 253.703 | 17 | 17 | [illegible] |
| 575 | 1 | Q14764 | Q14764 | Major vault p | MVP | Homo sapien | 99.328 | 17 | 17 | [illegible] |
| 576 | 1 | Q14766 | Q14766 | Latent-transf | LTBP1 | Homo sapien | 186.797 | 17 | 17 | [illegible] |
| 577 | 1 | Q14974 | Q14974 | Importin sub | KPNB1 | Homo sapien | 97.172 | 17 | 17 | [illegible] |
| 578 | 1 | Q2M389 | Q2M389 | WASH comp | WASHC4 | Homo sapien | 136.404 | 17 | 17 | [illegible] |
| 579 | 1 | Q2M3D2 | Q2M3D2 | Exocyst com | EXOC3L2 | Homo sapien | 87.482 | 17 | 17 | [illegible] |
| 580 | 1 | Q32MZ4 | Q32MZ4 | Leucine-rich | LRRFIP1 | Homo sapien | 89.253 | 18 | 17 | [illegible] |
| 581 | 1 | Q32P44 | Q32P44 | Echinoderm | EML3 | Homo sapien | 95.2 | 17 | 17 | [illegible] |
| 582 | 1 | Q5M775 | Q5M775 | Cytospin-B | SPECC1 | Homo sapien | 118.587 | 17 | 17 | [illegible] |
| 583 | 1 | Q5SW79 | Q5SW79 | Centrosomal | CEP170 | Homo sapien | 175.295 | 25 | 17 | [illegible] |
| 584 | 1 | Q5T0F9 | Q5T0F9 | Coiled-coil an | CC2D1B | Homo sapien | 94.224 | 17 | 17 | [illegible] |
| 585 | 1 | Q5VZK9 | Q5VZK9 | F-actin-unca | CARMIL1 | Homo sapien | 151.557 | 17 | 17 | [illegible] |
| 586 | 1 | Q6WKZ4 | Q6WKZ4 | Rab11 family | RAB11FIP1 | Homo sapien | 137.168 | 17 | 17 | [illegible] |
| 587 | 1 | Q86VS8 | Q86VS8 | Protein Hook | HOOK3 | Homo sapien | 83.129 | 17 | 17 | [illegible] |
| 588 | 1 | Q8IWJ2 | Q8IWJ2 | GRIP and co | GCC2 | Homo sapien | 195.912 | 18 | 17 | [illegible] |
| 589 | 1 | Q8IY17 | Q8IY17 | Patatin-like p | PNPLA6 | Homo sapien | 150.958 | 18 | 17 | [illegible] |
| 590 | 1 | Q8IY33 | Q8IY33 | MICAL-like p | MICALL2 | Homo sapien | 97.503 | 17 | 17 | [illegible] |
| 591 | 1 | Q8N1F7 | Q8N1F7 | Nuclear pore | NUP93 | Homo sapien | 93.489 | 17 | 17 | [illegible] |
| 592 | 1 | Q8TEX9 | Q8TEX9 | Importin-4 | IPO4 | Homo sapien | 118.717 | 17 | 17 | [illegible] |
| 593 | 1 | Q93100 | Q93100 | Phosphoryla | PHKB | Homo sapien | 124.885 | 17 | 17 | [illegible] |
| 594 | 1 | Q969R2 | Q969R2 | Oxysterol-bi | OSBP2 | Homo sapien | 101.268 | 18 | 17 | [illegible] |
| 595 | 1 | Q96BY7 | Q96BY7 | Autophagy-r | ATG2B | Homo sapien | 232.768 | 17 | 17 | [illegible] |
| 596 | 1 | Q96EK7 | Q96EK7 | Constitutive | FAM120B | Homo sapien | 103.785 | 17 | 17 | [illegible] |
| 597 | 1 | Q96KP1 | Q96KP1 | Exocyst com | EXOC2 | Homo sapien | 104.066 | 17 | 17 | [illegible] |
| 598 | 1 | Q96S38 | Q96S38 | Ribosomal pr | RPS6KC1 | Homo sapien | 118.683 | 17 | 17 | [illegible] |
| 599 | 1 | Q99798 | Q99798 | Aconitate hy | ACO2 | Homo sapien | 85.426 | 17 | 17 | [illegible] |
| 600 | 1 | Q9BPX3 | Q9BPX3 | Condensin c | NCAPG | Homo sapien | 114.337 | 17 | 17 | [illegible] |
| 601 | 1 | Q9BQG0 | Q9BQG0 | Myb-binding | MYBBP1A | Homo sapien | 148.856 | 17 | 17 | [illegible] |
| 602 | 1 | Q9H3U1 | Q9H3U1 | Protein unc-4 | UNC45A | Homo sapien | 103.077 | 17 | 17 | [illegible] |
| 603 | 1 | Q9H9A6 | Q9H9A6 | Leucine-rich | LRRC40 | Homo sapien | 68.25 | 17 | 17 | [illegible] |
| 604 | 1 | Q9HBR0 | Q9HBR0 | Solute carrier | SLC38A10 | Homo sapien | 119.764 | 17 | 17 | [illegible] |
| 605 | 1 | Q9HC35 | Q9HC35 | Echinoderm | EML4 | Homo sapien | 108.916 | 17 | 17 | [illegible] |
| 606 | 1 | Q9NP58 | Q9NP58 | ATP-binding | ABCB6 | Homo sapien | 93.885 | 17 | 17 | [illegible] |
| 607 | 1 | Q9NRL2 | Q9NRL2 | Bromodomai | BAZ1A | Homo sapien | 178.703 | 17 | 17 | [illegible] |
| 608 | 1 | Q9Y3P9 | Q9Y3P9 | Rab GTPase | RABGAP1 | Homo sapien | 121.737 | 18 | 17 | [illegible] |
| 609 | 1 | Q9Y5B9 | Q9Y5B9 | FACT compl | SUPT16H | Homo sapien | 119.916 | 17 | 17 | [illegible] |
| 610 | 1 | A1A4S6 | A1A4S6 | Rho GTPase | ARHGAP10 | Homo sapien | 89.377 | 16 | 16 | [illegible] |
| 611 | 1 | O00267 | O00267 | Transcription | SUPT5H | Homo sapien | 121.002 | 16 | 16 | [illegible] |
| 612 | 1 | O15294 | O15294 | UDP-N-acety | OGT | Homo sapien | 116.925 | 16 | 16 | [illegible] |
| 613 | 1 | O43815 | O43815 | Striatin | STRN | Homo sapien | 86.134 | 16 | 16 | [illegible] |
| 614 | 1 | O60716 | O60716 | Catenin delta | CTNND1 | Homo sapien | 108.171 | 16 | 16 | [illegible] |
| 615 | 1 | O95340 | O95340 | Bifunctional 3 | PAPSS2 | Homo sapien | 69.498 | 16 | 16 | [illegible] |
| 616 | 1 | O95373 | O95373 | Importin-7 | IPO7 | Homo sapien | 119.517 | 16 | 16 | [illegible] |
| 617 | 1 | P00558 | P00558 | Phosphoglyc | PGK1 | Homo sapien | 44.616 | 20 | 16 | [illegible] |
| 618 | 1 | P02649 | P02649 | Apolipoprote | APOE | Homo sapien | 36.153 | 16 | 16 | [illegible] |
| 619 | 1 | P04843 | P04843 | Dolichyl-diph | RPN1 | Homo sapien | 68.57 | 16 | 16 | [illegible] |
| 620 | 1 | P05165 | P05165 | Propionyl-Co | PCCA | Homo sapien | 80.06 | 16 | 16 | [illegible] |
| 621 | 1 | P06681 | P06681 | Complement | C2 | Homo sapien | 83.269 | 16 | 16 | [illegible] |
| 622 | 1 | P08779 | P08779 | Keratin, type | KRT16 | Homo sapien | 51.268 | 26 | 16 | [illegible] |
| 623 | 1 | P10909 | P10909 | Clusterin | CLU | Homo sapien | 52.494 | 16 | 16 | [illegible] |
| 624 | 1 | P12956 | P12956 | X-ray repair c | XRCC6 | Homo sapien | 69.844 | 16 | 16 | [illegible] |
| 625 | 1 | P13010 | P13010 | X-ray repair c | XRCC5 | Homo sapien | 82.705 | 16 | 16 | [illegible] |
| 626 | 1 | P14618 | P14618 | Pyruvate kin | PKM | Homo sapien | 57.938 | 17 | 16 | [illegible] |
| 627 | 1 | P17987 | P17987 | T-complex pr | TCP1 | Homo sapien | 60.343 | 16 | 16 | [illegible] |
| 628 | 1 | P18054 | P18054 | Polyunsatura | ALOX12 | Homo sapien | 75.694 | 16 | 16 | [illegible] |
| 629 | 1 | P19823 | P19823 | Inter-alpha-tr | ITIH2 | Homo sapien | 106.465 | 16 | 16 | [illegible] |
| 630 | 1 | P21399 | P21399 | Cytoplasmic | ACO1 | Homo sapien | 98.403 | 16 | 16 | [illegible] |
| 631 | 1 | P22102 | P22102 | Trifunctional | GART | Homo sapien | 107.766 | 16 | 16 | [illegible] |
| 632 | 1 | P23276 | P23276 | Kell blood gr | KEL | Homo sapien | 82.827 | 16 | 16 | [illegible] |
| 633 | 1 | P23458 | P23458 | Tyrosine-pro | JAK1 | Homo sapien | 133.279 | 16 | 16 | [illegible] |
| 634 | 1 | P23743 | P23743 | Diacylglycer | DGKA | Homo sapien | 82.632 | 16 | 16 | [illegible] |
| 635 | 1 | P26358 | P26358 | DNA (cytosin | DNMT1 | Homo sapien | 183.167 | 16 | 16 | [illegible] |
| 636 | 1 | P26639 | P26639 | Threonine--t | TARS1 | Homo sapien | 83.436 | 18 | 16 | [illegible] |
| 637 | 1 | P27448 | P27448 | MAP/microtu | MARK3 | Homo sapien | 84.429 | 21 | 16 | [illegible] |
| 638 | 1 | P28838 | P28838 | Cytosol amin | LAP3 | Homo sapien | 56.168 | 16 | 16 | [illegible] |
| 639 | 1 | P30740 | P30740 | Leukocyte el | SERPINB1 | Homo sapien | 42.741 | 17 | 16 | [illegible] |
| 640 | 1 | P33992 | P33992 | DNA replicat | MCM5 | Homo sapien | 82.284 | 16 | 16 | [illegible] |
| 641 | 1 | P33993 | P33993 | DNA replicat | MCM7 | Homo sapien | 81.31 | 16 | 16 | [illegible] |
| 642 | 1 | P35241 | P35241 | Radixin | RDX | Homo sapien | 68.564 | 17 | 16 | [illegible] |
| 643 | 1 | P35998 | P35998 | 26S proteas | PSMC2 | Homo sapien | 48.634 | 16 | 16 | [illegible] |
| 644 | 1 | P36915 | P36915 | Guanine nuc | GNL1 | Homo sapien | 68.661 | 16 | 16 | [illegible] |
| 645 | 1 | P42331 | P42331 | Rho GTPase | ARHGAP25 | Homo sapien | 73.435 | 16 | 16 | [illegible] |
| 646 | 1 | P45974 | P45974 | Ubiquitin car | USP5 | Homo sapien | 95.786 | 18 | 16 | [illegible] |
| 647 | 1 | P46100 | P46100 | Transcription | ATRX | Homo sapien | 282.591 | 16 | 16 | [illegible] |
| 648 | 1 | P49641 | P49641 | Alpha-manno | MAN2A2 | Homo sapien | 130.542 | 16 | 16 | [illegible] |
| 649 | 1 | P50416 | P50416 | Carnitine O-p | CPT1A | Homo sapien | 88.367 | 16 | 16 | [illegible] |
| 650 | 1 | P51812 | P51812 | Ribosomal pr | RPS6KA3 | Homo sapien | 83.737 | 19 | 16 | [illegible] |
| 651 | 1 | P55265 | P55265 | Double-stran | ADAR | Homo sapien | 136.067 | 16 | 16 | [illegible] |
| 652 | 1 | P61221 | P61221 | ATP-binding | ABCE1 | Homo sapien | 67.315 | 16 | 16 | [illegible] |
| 653 | 1 | Q01968 | Q01968 | Inositol polyp | OCRL | Homo sapien | 104.204 | 17 | 16 | [illegible] |
| 654 | 1 | Q02153 | Q02153 | Guanylate cy | GUCY1B1 | Homo sapien | 70.514 | 16 | 16 | [illegible] |
| 655 | 1 | Q05086 | Q05086 | Ubiquitin-prot | UBE3A | Homo sapien | 100.691 | 16 | 16 | [illegible] |
| 656 | 1 | Q0VD83 | Q0VD83 | Apolipoprote | APOBR | Homo sapien | 115.634 | 16 | 16 | [illegible] |
| 657 | 1 | Q12906 | Q12906 | Interleukin e | ILF3 | Homo sapien | 95.341 | 19 | 16 | [illegible] |
| 658 | 1 | Q12913 | Q12913 | Receptor-typ | PTPRJ | Homo sapien | 145.944 | 16 | 16 | [illegible] |
| 659 | 1 | Q14247 | Q14247 | Src substrate | CTTN | Homo sapien | 61.585 | 16 | 16 | [illegible] |
| 660 | 1 | Q14677 | Q14677 | Clathrin inter | CLINT1 | Homo sapien | 68.26 | 16 | 16 | [illegible] |
| 661 | 1 | Q16363 | Q16363 | Laminin subu | LAMA4 | Homo sapien | 202.569 | 16 | 16 | [illegible] |
| 662 | 1 | Q16512 | Q16512 | Serine/threo | PKN1 | Homo sapien | 103.933 | 16 | 16 | [illegible] |
| 663 | 1 | Q29RF7 | Q29RF7 | Sister chrom | PDS5A | Homo sapien | 150.829 | 16 | 16 | [illegible] |
| 664 | 1 | Q4KMP7 | Q4KMP7 | TBC1 domai | TBC1D10B | Homo sapien | 87.2 | 16 | 16 | [illegible] |
| 665 | 1 | Q5D862 | Q5D862 | Filaggrin-2 | FLG2 | Homo sapien | 248.077 | 16 | 16 | [illegible] |
| 666 | 1 | Q5T1M5 | Q5T1M5 | FK506-bindi | FKBP15 | Homo sapien | 133.63 | 16 | 16 | [illegible] |
| 667 | 1 | Q5T5C0 | Q5T5C0 | Syntaxin-bin | STXBP5 | Homo sapien | 127.575 | 18 | 16 | [illegible] |
| 668 | 1 | Q5T5Y3 | Q5T5Y3 | Calmodulin-r | CAMSAP1 | Homo sapien | 177.974 | 16 | 16 | [illegible] |
| 669 | 1 | Q66K74 | Q66K74 | Microtubule-a | MAP1S | Homo sapien | 112.213 | 16 | 16 | [illegible] |
| 670 | 1 | Q6GYQ0 | Q6GYQ0 | Ral GTPase- | RALGAPA1 | Homo sapien | 229.837 | 16 | 16 | [illegible] |
| 671 | 1 | Q6P1M0 | Q6P1M0 | Long-chain f | SLC27A4 | Homo sapien | 72.067 | 16 | 16 | [illegible] |
| 672 | 1 | Q6P2E9 | Q6P2E9 | Enhancer of | EDC4 | Homo sapien | 151.664 | 16 | 16 | [illegible] |
| 673 | 1 | Q6PKG0 | Q6PKG0 | La-related pr | LARP1 | Homo sapien | 123.512 | 19 | 16 | [illegible] |
| 674 | 1 | Q6UWP8 | Q6UWP8 | Suprabasin | SBSN | Homo sapien | 60.541 | 16 | 16 | [illegible] |
| 675 | 1 | Q7Z7A1 | Q7Z7A1 | Centriolin | CNTRL | Homo sapien | 268.89 | 16 | 16 | [illegible] |
| 676 | 1 | Q86UE4 | Q86UE4 | Protein LYRI | MTDH | Homo sapien | 63.837 | 16 | 16 | [illegible] |
| 677 | 1 | Q8ND71 | Q8ND71 | GTPase IMA | GIMAP8 | Homo sapien | 74.891 | 16 | 16 | [illegible] |
| 678 | 1 | Q8TAQ2 | Q8TAQ2 | SWI/SNF co | SMARCC2 | Homo sapien | 132.881 | 21 | 16 | [illegible] |
| 679 | 1 | Q8TDB6 | Q8TDB6 | E3 ubiquitin-p | DTX3L | Homo sapien | 83.556 | 16 | 16 | [illegible] |
| 680 | 1 | Q92598 | Q92598 | Heat shock p | HSPH1 | Homo sapien | 96.865 | 16 | 16 | [illegible] |
| 681 | 1 | Q92797 | Q92797 | Symplekin | SYMPK | Homo sapien | 141.151 | 16 | 16 | [illegible] |
| 682 | 1 | Q92954 | Q92954 | Proteoglycan | PRG4 | Homo sapien | 151.064 | 16 | 16 | [illegible] |
| 683 | 1 | Q96CV9 | Q96CV9 | Optineurin | OPTN | Homo sapien | 65.922 | 16 | 16 | [illegible] |
| 684 | 1 | Q96P70 | Q96P70 | Importin-9 | IPO9 | Homo sapien | 115.964 | 16 | 16 | [illegible] |
| 685 | 1 | Q96PK6 | Q96PK6 | RNA-binding | RBM14 | Homo sapien | 69.491 | 16 | 16 | [illegible] |
| 686 | 1 | Q99543 | Q99543 | DnaJ homolo | DNAJC2 | Homo sapien | 71.997 | 16 | 16 | [illegible] |
| 687 | 1 | Q9BTW9 | Q9BTW9 | Tubulin-spec | TBCD | Homo sapien | 132.601 | 16 | 16 | [illegible] |
| 688 | 1 | Q9BZF1 | Q9BZF1 | Oxysterol-bi | OSBPL8 | Homo sapien | 101.197 | 17 | 16 | [illegible] |
| 689 | 1 | Q9BZH6 | Q9BZH6 | WD repeat-c | WDR11 | Homo sapien | 136.687 | 16 | 16 | [illegible] |
| 690 | 1 | Q9BZL4 | Q9BZL4 | Protein phos | PPP1R12C | Homo sapien | 84.883 | 16 | 16 | [illegible] |
| 691 | 1 | Q9C0I1 | Q9C0I1 | Myotubularin- | MTMR12 | Homo sapien | 86.148 | 16 | 16 | [illegible] |
| 692 | 1 | Q9H267 | Q9H267 | Vacuolar pro | VPS33B | Homo sapien | 70.588 | 16 | 16 | [illegible] |
| 693 | 1 | Q9H2P0 | Q9H2P0 | Activity-depe | ADNP | Homo sapien | 123.565 | 16 | 16 | [illegible] |
| 694 | 1 | Q9H307 | Q9H307 | Pinin | PNN | Homo sapien | 81.629 | 16 | 16 | [illegible] |
| 695 | 1 | Q9H8Y5 | Q9H8Y5 | tRNA endon | ANKZF1 | Homo sapien | 80.927 | 16 | 16 | [illegible] |
| 696 | 1 | Q9HBI1 | Q9HBI1 | Beta-parvin | PARVB | Homo sapien | 41.715 | 16 | 16 | [illegible] |
| 697 | 1 | Q9HCM2 | Q9HCM2 | Plexin-A4 | PLXNA4 | Homo sapien | 212.459 | 24 | 16 | [illegible] |
| 698 | 1 | Q9HD20 | Q9HD20 | Endoplasmic | ATP13A1 | Homo sapien | 132.957 | 16 | 16 | [illegible] |
| 699 | 1 | Q9NR30 | Q9NR30 | Nucleolar RN | DDX21 | Homo sapien | 87.344 | 17 | 16 | [illegible] |
| 700 | 1 | Q9NSE4 | Q9NSE4 | Isoleucine--t | IARS2 | Homo sapien | 113.793 | 16 | 16 | [illegible] |
| 701 | 1 | Q9NSK0 | Q9NSK0 | Kinesin light | KLC4 | Homo sapien | 68.641 | 18 | 16 | [illegible] |
| 702 | 1 | Q9NU22 | Q9NU22 | Midasin | MDN1 | Homo sapien | 632.83 | 16 | 16 | [illegible] |
| 703 | 1 | Q9NY33 | Q9NY33 | Dipeptidyl pe | DPP3 | Homo sapien | 82.589 | 16 | 16 | [illegible] |
| 704 | 1 | Q9P2E3 | Q9P2E3 | NFX1-type zi | ZNFX1 | Homo sapien | 220.231 | 16 | 16 | [illegible] |
| 705 | 1 | Q9UH99 | Q9UH99 | SUN domain- | SUN2 | Homo sapien | 80.314 | 16 | 16 | [illegible] |
| 706 | 1 | Q9UI26 | Q9UI26 | Importin-11 | IPO11 | Homo sapien | 112.535 | 16 | 16 | [illegible] |
| 707 | 1 | Q9UKW4 | Q9UKW4 | Guanine nuc | VAV3 | Homo sapien | 97.777 | 16 | 16 | [illegible] |
| 708 | 1 | Q9ULH0 | Q9ULH0 | Kinase D-inte | KIDINS220 | Homo sapien | 196.545 | 16 | 16 | [illegible] |
| 709 | 1 | Q9UNM6 | Q9UNM6 | 26S proteas | PSMD13 | Homo sapien | 42.944 | 16 | 16 | [illegible] |
| 710 | 1 | Q9Y265 | Q9Y265 | RuvB-like 1 | RUVBL1 | Homo sapien | 50.226 | 16 | 16 | [illegible] |
| 711 | 1 | Q9Y2X3 | Q9Y2X3 | Nucleolar pro | NOP58 | Homo sapien | 59.581 | 16 | 16 | [illegible] |
| 712 | 1 | Q9Y6D5 | Q9Y6D5 | Brefeldin A-in | ARFGEF2 | Homo sapien | 202.043 | 16 | 16 | [illegible] |
| 713 | 1 | A1IGU5 | A1IGU5 | Rho guanine | ARHGEF37 | Homo sapien | 76.279 | 15 | 15 | [illegible] |
| 714 | 1 | A4D1P6 | A4D1P6 | WD repeat-c | WDR91 | Homo sapien | 83.344 | 15 | 15 | [illegible] |
| 715 | 1 | O00178 | O00178 | GTP-binding | GTPBP1 | Homo sapien | 72.457 | 15 | 15 | [illegible] |
| 716 | 1 | O15020 | O15020 | Spectrin beta | SPTBN2 | Homo sapien | 271.33 | 16 | 15 | [illegible] |
| 717 | 1 | O15050 | O15050 | TPR and ank | TRANK1 | Homo sapien | 336.227 | 15 | 15 | [illegible] |
| 718 | 1 | O15067 | O15067 | Phosphoribo | PFAS | Homo sapien | 144.738 | 15 | 15 | [illegible] |
| 719 | 1 | O15327 | O15327 | Inositol polyp | INPP4B | Homo sapien | 104.738 | 15 | 15 | [illegible] |
| 720 | 1 | O43166 | O43166 | Signal-induce | SIPA1L1 | Homo sapien | 200.03 | 17 | 15 | [illegible] |
| 721 | 1 | O43491 | O43491 | Band 4.1-like | EPB41L2 | Homo sapien | 112.588 | 16 | 15 | [illegible] |
| 722 | 1 | O60264 | O60264 | SWI/SNF-rel | SMARCA5 | Homo sapien | 121.907 | 24 | 15 | [illegible] |
| 723 | 1 | O60674 | O60674 | Tyrosine-pro | JAK2 | Homo sapien | 130.674 | 16 | 15 | [illegible] |
| 724 | 1 | O75150 | O75150 | E3 ubiquitin-p | RNF40 | Homo sapien | 113.682 | 15 | 15 | [illegible] |
| 725 | 1 | O75746 | O75746 | Electrogenic | SLC25A12 | Homo sapien | 74.762 | 18 | 15 | [illegible] |
| 726 | 1 | P00338 | P00338 | L-lactate deh | LDHA | Homo sapien | 36.688 | 16 | 15 | [illegible] |
| 727 | 1 | P00915 | P00915 | Carbonic anh | CA1 | Homo sapien | 28.87 | 15 | 15 | [illegible] |
| 728 | 1 | P02786 | P02786 | Transferrin r | TFRC | Homo sapien | 84.872 | 15 | 15 | [illegible] |
| 729 | 1 | P02790 | P02790 | Hemopexin | HPX | Homo sapien | 51.676 | 15 | 15 | [illegible] |
| 730 | 1 | P05154 | P05154 | Plasma serin | SERPINA5 | Homo sapien | 45.673 | 15 | 15 | [illegible] |
| 731 | 1 | P05164 | P05164 | Myeloperoxid | MPO | Homo sapien | 83.872 | 15 | 15 | [illegible] |
| 732 | 1 | P06396 | P06396 | Gelsolin | GSN | Homo sapien | 85.699 | 15 | 15 | [illegible] |
| 733 | 1 | P06737 | P06737 | Glycogen ph | PYGL | Homo sapien | 97.151 | 19 | 15 | [illegible] |
| 734 | 1 | P11216 | P11216 | Glycogen ph | PYGB | Homo sapien | 96.699 | 16 | 15 | [illegible] |
| 735 | 1 | P13671 | P13671 | Complement | C6 | Homo sapien | 104.789 | 15 | 15 | [illegible] |
| 736 | 1 | P16050 | P16050 | Polyunsatura | ALOX15 | Homo sapien | 74.805 | 15 | 15 | [illegible] |
| 737 | 1 | P17980 | P17980 | 26S proteas | PSMC3 | Homo sapien | 49.204 | 15 | 15 | [illegible] |
| 738 | 1 | P19320 | P19320 | Vascular cell | VCAM1 | Homo sapien | 81.276 | 15 | 15 | [illegible] |
| 739 | 1 | P23526 | P23526 | Adenosylhon | AHCY | Homo sapien | 47.715 | 15 | 15 | [illegible] |
| 740 | 1 | P25685 | P25685 | DnaJ homolo | DNAJB1 | Homo sapien | 38.043 | 16 | 15 | [illegible] |
| 741 | 1 | P35658 | P35658 | Nuclear pore | NUP214 | Homo sapien | 213.623 | 15 | 15 | [illegible] |
| 742 | 1 | P35749 | P35749 | Myosin-11 | MYH11 | Homo sapien | 227.342 | 15 | 15 | [illegible] |
| 743 | 1 | P38646 | P38646 | Stress-70 pr | HSPA9 | Homo sapien | 73.682 | 15 | 15 | [illegible] |
| 744 | 1 | P49815 | P49815 | Tuberin | TSC2 | Homo sapien | 200.613 | 15 | 15 | [illegible] |
| 745 | 1 | P49959 | P49959 | Double-stran | MRE11 | Homo sapien | 80.594 | 15 | 15 | [illegible] |
| 746 | 1 | P50395 | P50395 | Rab GDP dis | GDI2 | Homo sapien | 50.666 | 21 | 15 | [illegible] |
| 747 | 1 | P53814 | P53814 | Smoothelin | SMTN | Homo sapien | 99.059 | 15 | 15 | [illegible] |
| 748 | 1 | P55010 | P55010 | Eukaryotic tr | EIF5 | Homo sapien | 49.224 | 15 | 15 | [illegible] |
| 749 | 1 | P55786 | P55786 | Puromycin-s | NPEPPS | Homo sapien | 103.277 | 23 | 15 | [illegible] |
| 750 | 1 | Q01780 | Q01780 | Exosome co | EXOSC10 | Homo sapien | 100.833 | 15 | 15 | [illegible] |
| 751 | 1 | Q07866 | Q07866 | Kinesin light | KLC1 | Homo sapien | 65.31 | 19 | 15 | [illegible] |
| 752 | 1 | Q09161 | Q09161 | Nuclear cap- | NCBP1 | Homo sapien | 91.842 | 15 | 15 | [illegible] |
| 753 | 1 | Q12769 | Q12769 | Nuclear pore | NUP160 | Homo sapien | 162.123 | 15 | 15 | [illegible] |
| 754 | 1 | Q13496 | Q13496 | Myotubularin | MTM1 | Homo sapien | 69.931 | 15 | 15 | [illegible] |
| 755 | 1 | Q14258 | Q14258 | E3 ubiquitin/I | TRIM25 | Homo sapien | 70.975 | 15 | 15 | [illegible] |
| 756 | 1 | Q14344 | Q14344 | Guanine nuc | GNA13 | Homo sapien | 44.05 | 16 | 15 | [illegible] |
| 757 | 1 | Q15008 | Q15008 | 26S proteas | PSMD6 | Homo sapien | 45.533 | 15 | 15 | [illegible] |
| 758 | 1 | Q15582 | Q15582 | Transforming | TGFBI | Homo sapien | 74.68 | 15 | 15 | [illegible] |
| 759 | 1 | Q15643 | Q15643 | Thyroid rece | TRIP11 | Homo sapien | 227.591 | 15 | 15 | [illegible] |
| 760 | 1 | Q15813 | Q15813 | Tubulin-spec | TBCE | Homo sapien | 59.348 | 15 | 15 | [illegible] |
| 761 | 1 | Q16204 | Q16204 | Coiled-coil d | CCDC6 | Homo sapien | 53.291 | 15 | 15 | [illegible] |
| 762 | 1 | Q16658 | Q16658 | Fascin | FSCN1 | Homo sapien | 54.531 | 15 | 15 | [illegible] |
| 763 | 1 | Q16799 | Q16799 | Reticulon-1 | RTN1 | Homo sapien | 83.616 | 15 | 15 | [illegible] |
| 764 | 1 | Q16851 | Q16851 | UTP--glucos | UGP2 | Homo sapien | 56.942 | 15 | 15 | [illegible] |
| 765 | 1 | Q5H9R7 | Q5H9R7 | Serine/threo | PPP6R3 | Homo sapien | 97.671 | 15 | 15 | [illegible] |
| 766 | 1 | Q5SSJ5 | Q5SSJ5 | Heterochrom | HP1BP3 | Homo sapien | 61.209 | 15 | 15 | [illegible] |
| 767 | 1 | Q5VTR2 | Q5VTR2 | E3 ubiquitin-p | RNF20 | Homo sapien | 113.663 | 17 | 15 | [illegible] |
| 768 | 1 | Q66GS9 | Q66GS9 | Centrosomal | CEP135 | Homo sapien | 133.491 | 15 | 15 | [illegible] |
| 769 | 1 | Q6BDS2 | Q6BDS2 | Bridge-like lip | BLTP3A | Homo sapien | 159.487 | 15 | 15 | [illegible] |
| 770 | 1 | Q6DN90 | Q6DN90 | IQ motif and | IQSEC1 | Homo sapien | 108.317 | 17 | 15 | [illegible] |
| 771 | 1 | Q6XZF7 | Q6XZF7 | Dynamin-bin | DNMBP | Homo sapien | 177.349 | 15 | 15 | [illegible] |
| 772 | 1 | Q6ZS81 | Q6ZS81 | WD repeat- a | WDFY4 | Homo sapien | 353.616 | 15 | 15 | [illegible] |
| 773 | 1 | Q86UK7 | Q86UK7 | E3 ubiquitin-p | ZNF598 | Homo sapien | 98.638 | 15 | 15 | [illegible] |
| 774 | 1 | Q86V48 | Q86V48 | Leucine zipp | LUZP1 | Homo sapien | 120.276 | 15 | 15 | [illegible] |
| 775 | 1 | Q86X10 | Q86X10 | Ral GTPase- | RALGAPB | Homo sapien | 166.804 | 15 | 15 | [illegible] |
| 776 | 1 | Q8IUR7 | Q8IUR7 | Armadillo rep | ARMC8 | Homo sapien | 75.511 | 15 | 15 | [illegible] |
| 777 | 1 | Q8N122 | Q8N122 | Regulatory-a | RPTOR | Homo sapien | 149.038 | 15 | 15 | [illegible] |
| 778 | 1 | Q8NB90 | Q8NB90 | ATPase fami | AFG2A | Homo sapien | 97.907 | 16 | 15 | [illegible] |
| 779 | 1 | Q8NDX1 | Q8NDX1 | PH and SEC | PSD4 | Homo sapien | 116.25 | 15 | 15 | [illegible] |
| 780 | 1 | Q8TEH3 | Q8TEH3 | DENN domai | DENND1A | Homo sapien | 110.58 | 15 | 15 | [illegible] |
| 781 | 1 | Q8TEU7 | Q8TEU7 | Rap guanine | RAPGEF6 | Homo sapien | 179.426 | 15 | 15 | [illegible] |
| 782 | 1 | Q92541 | Q92541 | RNA polymer | RTF1 | Homo sapien | 80.315 | 15 | 15 | [illegible] |
| 783 | 1 | Q92609 | Q92609 | TBC1 domai | TBC1D5 | Homo sapien | 89.004 | 15 | 15 | [illegible] |
| 784 | 1 | Q92673 | Q92673 | Sortilin-relate | SORL1 | Homo sapien | 248.432 | 15 | 15 | [illegible] |
| 785 | 1 | Q92945 | Q92945 | Far upstream | KHSRP | Homo sapien | 73.117 | 16 | 15 | [illegible] |
| 786 | 1 | Q92974 | Q92974 | Rho guanine | ARHGEF2 | Homo sapien | 111.544 | 15 | 15 | [illegible] |
| 787 | 1 | Q96CN9 | Q96CN9 | GRIP and co | GCC1 | Homo sapien | 87.812 | 15 | 15 | [illegible] |
| 788 | 1 | Q96Q05 | Q96Q05 | Trafficking pr | TRAPPC9 | Homo sapien | 128.532 | 15 | 15 | [illegible] |
| 789 | 1 | Q99832 | Q99832 | T-complex pr | CCT7 | Homo sapien | 59.365 | 15 | 15 | [illegible] |
| 790 | 1 | Q9BZI7 | Q9BZI7 | Regulator of | UPF3B | Homo sapien | 57.76 | 15 | 15 | [illegible] |
| 791 | 1 | Q9C0C2 | Q9C0C2 | 182 kDa tank | TNKS1BP1 | Homo sapien | 181.799 | 15 | 15 | [illegible] |
| 792 | 1 | Q9C0C7 | Q9C0C7 | Activating mo | AMBRA1 | Homo sapien | 142.507 | 15 | 15 | [illegible] |
| 793 | 1 | Q9GZR7 | Q9GZR7 | ATP-depend | DDX24 | Homo sapien | 96.333 | 15 | 15 | [illegible] |
| 794 | 1 | Q9NTK5 | Q9NTK5 | Obg-like ATP | OLA1 | Homo sapien | 44.741 | 15 | 15 | [illegible] |
| 795 | 1 | Q9NXC5 | Q9NXC5 | GATOR2 co | MIOS | Homo sapien | 98.584 | 15 | 15 | [illegible] |
| 796 | 1 | Q9NZW5 | Q9NZW5 | Protein PALS | PALS2 | Homo sapien | 61.118 | 16 | 15 | [illegible] |
| 797 | 1 | Q9UBF2 | Q9UBF2 | Coatomer su | COPG2 | Homo sapien | 97.623 | 15 | 15 | [illegible] |
| 798 | 1 | Q9UBP0 | Q9UBP0 | Spastin | SPAST | Homo sapien | 67.199 | 15 | 15 | [illegible] |
| 799 | 1 | Q9UH65 | Q9UH65 | Switch-asso | SWAP70 | Homo sapien | 68.998 | 15 | 15 | [illegible] |
| 800 | 1 | Q9UHD2 | Q9UHD2 | Serine/threo | TBK1 | Homo sapien | 83.644 | 15 | 15 | [illegible] |
| 801 | 1 | Q9UIG0 | Q9UIG0 | Tyrosine-pro | BAZ1B | Homo sapien | 170.906 | 15 | 15 | [illegible] |
| 802 | 1 | Q9UKU0 | Q9UKU0 | Long-chain-f | ACSL6 | Homo sapien | 77.753 | 17 | 15 | [illegible] |
| 803 | 1 | Q9ULV4 | Q9ULV4 | Coronin-1C | CORO1C | Homo sapien | 53.248 | 15 | 15 | [illegible] |
| 804 | 1 | Q9UPU7 | Q9UPU7 | TBC1 domai | TBC1D2B | Homo sapien | 109.883 | 15 | 15 | [illegible] |
| 805 | 1 | Q9Y2G3 | Q9Y2G3 | Phospholipid- | ATP11B | Homo sapien | 134.189 | 16 | 15 | [illegible] |
| 806 | 1 | Q9Y315 | Q9Y315 | Deoxyribose- | DERA | Homo sapien | 35.231 | 15 | 15 | [illegible] |
| 807 | 1 | Q9Y4K4 | Q9Y4K4 | Mitogen-activ | MAP4K5 | Homo sapien | 95.056 | 15 | 15 | [illegible] |
| 808 | 1 | Q9Y6R4 | Q9Y6R4 | Mitogen-activ | MAP3K4 | Homo sapien | 181.687 | 15 | 15 | [illegible] |
| 809 | 1 | Q9Y6W3 | Q9Y6W3 | Calpain-7 | CAPN7 | Homo sapien | 92.654 | 15 | 15 | [illegible] |
| 810 | 1 | O00471 | O00471 | Exocyst com | EXOC5 | Homo sapien | 81.85 | 14 | 14 | [illegible] |
| 811 | 1 | O00567 | O00567 | Nucleolar pro | NOP56 | Homo sapien | 66.051 | 14 | 14 | [illegible] |
| 812 | 1 | O15117 | O15117 | FYN-binding | FYB1 | Homo sapien | 85.388 | 14 | 14 | [illegible] |
| 813 | 1 | O15372 | O15372 | Eukaryotic tr | EIF3H | Homo sapien | 39.931 | 14 | 14 | [illegible] |
| 814 | 1 | O43847 | O43847 | Nardilysin | NRDC | Homo sapien | 131.705 | 14 | 14 | [illegible] |
| 815 | 1 | O60229 | O60229 | Kalirin | KALRN | Homo sapien | 340.265 | 17 | 14 | [illegible] |
| 816 | 1 | O60568 | O60568 | Multifunction | PLOD3 | Homo sapien | 84.787 | 14 | 14 | [illegible] |
| 817 | 1 | O60885 | O60885 | Bromodomai | BRD4 | Homo sapien | 152.22 | 18 | 14 | [illegible] |
| 818 | 1 | O94973 | O94973 | AP-2 comple | AP2A2 | Homo sapien | 103.961 | 14 | 14 | [illegible] |
| 819 | 1 | O94979 | O94979 | Protein trans | SEC31A | Homo sapien | 133.018 | 14 | 14 | [illegible] |
| 820 | 1 | O95140 | O95140 | Mitofusin-2 | MFN2 | Homo sapien | 86.403 | 14 | 14 | [illegible] |
| 821 | 1 | O95163 | O95163 | Elongator co | ELP1 | Homo sapien | 150.254 | 14 | 14 | [illegible] |
| 822 | 1 | O95352 | O95352 | Ubiquitin-like | ATG7 | Homo sapien | 77.96 | 14 | 14 | [illegible] |
| 823 | 1 | P01871 | P01871 | Immunoglobu | IGHM | Homo sapien | 51.924 | 14 | 14 | [illegible] |
| 824 | 1 | P06576 | P06576 | ATP synthas | ATP5F1B | Homo sapien | 56.559 | 14 | 14 | [illegible] |
| 825 | 1 | P07195 | P07195 | L-lactate deh | LDHB | Homo sapien | 36.639 | 14 | 14 | [illegible] |
| 826 | 1 | P07225 | P07225 | Vitamin K-de | PROS1 | Homo sapien | 75.123 | 14 | 14 | [illegible] |
| 827 | 1 | P08519 | P08519 | Apolipoprote | LPA | Homo sapien | 226.55 | 14 | 14 | [illegible] |
| 828 | 1 | P09960 | P09960 | Leukotriene | LTA4H | Homo sapien | 69.285 | 14 | 14 | [illegible] |
| 829 | 1 | P10586 | P10586 | Receptor-typ | PTPRF | Homo sapien | 212.882 | 16 | 14 | [illegible] |
| 830 | 1 | P13647 | P13647 | Keratin, type | KRT5 | Homo sapien | 62.378 | 23 | 14 | [illegible] |
| 831 | 1 | P16284 | P16284 | Platelet endo | PECAM1 | Homo sapien | 82.524 | 14 | 14 | [illegible] |
| 832 | 1 | P20936 | P20936 | Ras GTPase | RASA1 | Homo sapien | 116.405 | 14 | 14 | [illegible] |
| 833 | 1 | P22059 | P22059 | Oxysterol-bi | OSBP | Homo sapien | 89.422 | 14 | 14 | [illegible] |
| 834 | 1 | P24557 | P24557 | Thromboxan | TBXAS1 | Homo sapien | 60.519 | 14 | 14 | [illegible] |
| 835 | 1 | P25705 | P25705 | ATP synthas | ATP5F1A | Homo sapien | 59.75 | 14 | 14 | [illegible] |
| 836 | 1 | P27105 | P27105 | Stomatin | STOM | Homo sapien | 31.73 | 14 | 14 | [illegible] |
| 837 | 1 | P27824 | P27824 | Calnexin | CANX | Homo sapien | 67.57 | 14 | 14 | [illegible] |
| 838 | 1 | P29622 | P29622 | Kallistatin | SERPINA4 | Homo sapien | 48.543 | 14 | 14 | [illegible] |
| 839 | 1 | P30613 | P30613 | Pyruvate kin | PKLR | Homo sapien | 61.83 | 14 | 14 | [illegible] |
| 840 | 1 | P33897 | P33897 | ATP-binding | ABCD1 | Homo sapien | 82.935 | 14 | 14 | [illegible] |
| 841 | 1 | P36776 | P36776 | Lon protease | LONP1 | Homo sapien | 106.492 | 14 | 14 | [illegible] |
| 842 | 1 | P42338 | P42338 | Phosphatidyl | PIK3CB | Homo sapien | 122.764 | 14 | 14 | [illegible] |
| 843 | 1 | P43686 | P43686 | 26S proteas | PSMC4 | Homo sapien | 47.367 | 14 | 14 | [illegible] |
| 844 | 1 | P46821 | P46821 | Microtubule-a | MAP1B | Homo sapien | 270.639 | 14 | 14 | [illegible] |
| 845 | 1 | P46934 | P46934 | E3 ubiquitin-p | NEDD4 | Homo sapien | 149.118 | 14 | 14 | [illegible] |
| 846 | 1 | P48553 | P48553 | Trafficking pr | TRAPPC10 | Homo sapien | 142.191 | 14 | 14 | [illegible] |
| 847 | 1 | P48735 | P48735 | Isocitrate de | IDH2 | Homo sapien | 50.911 | 14 | 14 | [illegible] |
| 848 | 1 | P49454 | P49454 | Centromere p | CENPF | Homo sapien | 357.532 | 14 | 14 | [illegible] |
| 849 | 1 | P49754 | P49754 | Vacuolar pro | VPS41 | Homo sapien | 98.567 | 14 | 14 | [illegible] |
| 850 | 1 | P49916 | P49916 | DNA ligase 3 | LIG3 | Homo sapien | 112.907 | 14 | 14 | [illegible] |
| 851 | 1 | P51610 | P51610 | Host cell fac | HCFC1 | Homo sapien | 208.734 | 14 | 14 | [illegible] |
| 852 | 1 | P51659 | P51659 | Peroxisomal | HSD17B4 | Homo sapien | 79.686 | 14 | 14 | [illegible] |
| 853 | 1 | P55157 | P55157 | Microsomal t | MTTP | Homo sapien | 99.353 | 14 | 14 | [illegible] |
| 854 | 1 | P55884 | P55884 | Eukaryotic tr | EIF3B | Homo sapien | 92.482 | 14 | 14 | [illegible] |
| 855 | 1 | P57737 | P57737 | Coronin-7 | CORO7 | Homo sapien | 100.607 | 14 | 14 | [illegible] |
| 856 | 1 | P61764 | P61764 | Syntaxin-bin | STXBP1 | Homo sapien | 67.569 | 14 | 14 | [illegible] |
| 857 | 1 | P61978 | P61978 | Heterogeneo | HNRNPK | Homo sapien | 50.979 | 14 | 14 | [illegible] |
| 858 | 1 | P62191 | P62191 | 26S proteas | PSMC1 | Homo sapien | 49.185 | 14 | 14 | [illegible] |
| 859 | 1 | P62826 | P62826 | GTP-binding | RAN | Homo sapien | 24.421 | 14 | 14 | [illegible] |
| 860 | 1 | P78344 | P78344 | Eukaryotic tr | EIF4G2 | Homo sapien | 102.363 | 14 | 14 | [illegible] |
| 861 | 1 | Q01432 | Q01432 | AMP deamin | AMPD3 | Homo sapien | 88.81 | 16 | 14 | [illegible] |
| 862 | 1 | Q02818 | Q02818 | Nucleobindin- | NUCB1 | Homo sapien | 53.879 | 14 | 14 | [illegible] |
| 863 | 1 | Q05193 | Q05193 | Dynamin-1 | DNM1 | Homo sapien | 97.409 | 18 | 14 | [illegible] |
| 864 | 1 | Q06033 | Q06033 | Inter-alpha-tr | ITIH3 | Homo sapien | 99.849 | 14 | 14 | [illegible] |
| 865 | 1 | Q10570 | Q10570 | Cleavage an | CPSF1 | Homo sapien | 160.886 | 14 | 14 | [illegible] |
| 866 | 1 | Q12965 | Q12965 | Unconvention | MYO1E | Homo sapien | 127.063 | 14 | 14 | [illegible] |
| 867 | 1 | Q12986 | Q12986 | Transcription | NFX1 | Homo sapien | 124.396 | 14 | 14 | [illegible] |
| 868 | 1 | Q13283 | Q13283 | Ras GTPase | G3BP1 | Homo sapien | 52.164 | 15 | 14 | [illegible] |
| 869 | 1 | Q13451 | Q13451 | Peptidyl-prol | FKBP5 | Homo sapien | 51.214 | 14 | 14 | [illegible] |
| 870 | 1 | Q13620 | Q13620 | Cullin-4B | CUL4B | Homo sapien | 103.981 | 14 | 14 | [illegible] |
| 871 | 1 | Q14112 | Q14112 | Nidogen-2 | NID2 | Homo sapien | 151.256 | 14 | 14 | [illegible] |
| 872 | 1 | Q14139 | Q14139 | Ubiquitin con | UBE4A | Homo sapien | 122.563 | 14 | 14 | [illegible] |
| 873 | 1 | Q14166 | Q14166 | Tubulin--tyro | TTLL12 | Homo sapien | 74.404 | 14 | 14 | [illegible] |
| 874 | 1 | Q14692 | Q14692 | Ribosome bi | BMS1 | Homo sapien | 145.811 | 14 | 14 | [illegible] |
| 875 | 1 | Q15147 | Q15147 | 1-phosphatid | PLCB4 | Homo sapien | 134.467 | 14 | 14 | [illegible] |
| 876 | 1 | Q15283 | Q15283 | Ras GTPase | RASA2 | Homo sapien | 96.618 | 14 | 14 | [illegible] |
| 877 | 1 | Q4KMQ2 | Q4KMQ2 | Anoctamin-6 | ANO6 | Homo sapien | 106.167 | 14 | 14 | [illegible] |
| 878 | 1 | Q53GS9 | Q53GS9 | Ubiquitin car | USP39 | Homo sapien | 65.379 | 14 | 14 | [illegible] |
| 879 | 1 | Q567U6 | Q567U6 | Coiled-coil d | CCDC93 | Homo sapien | 73.196 | 14 | 14 | [illegible] |
| 880 | 1 | Q5VZE5 | Q5VZE5 | N-alpha-ace | NAA35 | Homo sapien | 83.64 | 14 | 14 | [illegible] |
| 881 | 1 | Q66K14 | Q66K14 | TBC1 domai | TBC1D9B | Homo sapien | 140.526 | 15 | 14 | [illegible] |
| 882 | 1 | Q6N069 | Q6N069 | N-alpha-ace | NAA16 | Homo sapien | 101.465 | 14 | 14 | [illegible] |
| 883 | 1 | Q70CQ4 | Q70CQ4 | Ubiquitin car | USP31 | Homo sapien | 146.654 | 14 | 14 | [illegible] |
| 884 | 1 | Q7LDG7 | Q7LDG7 | RAS guanyl-r | RASGRP2 | Homo sapien | 69.249 | 14 | 14 | [illegible] |
| 885 | 1 | Q86YT6 | Q86YT6 | E3 ubiquitin-p | MIB1 | Homo sapien | 110.137 | 14 | 14 | [illegible] |
| 886 | 1 | Q8IVT5 | Q8IVT5 | Kinase suppr | KSR1 | Homo sapien | 102.161 | 15 | 14 | [illegible] |
| 887 | 1 | Q8IYI6 | Q8IYI6 | Exocyst com | EXOC8 | Homo sapien | 81.799 | 14 | 14 | [illegible] |
| 888 | 1 | Q8IZ83 | Q8IZ83 | Aldehyde de | ALDH16A1 | Homo sapien | 85.127 | 14 | 14 | [illegible] |
| 889 | 1 | Q8NB16 | Q8NB16 | Mixed linea | MLKL | Homo sapien | 54.48 | 14 | 14 | [illegible] |
| 890 | 1 | Q8NDB2 | Q8NDB2 | B-cell scaffo | BANK1 | Homo sapien | 89.282 | 14 | 14 | [illegible] |
| 891 | 1 | Q8NI36 | Q8NI36 | WD repeat-c | WDR36 | Homo sapien | 105.324 | 14 | 14 | [illegible] |
| 892 | 1 | Q8TAG9 | Q8TAG9 | Exocyst com | EXOC6 | Homo sapien | 93.723 | 15 | 14 | [illegible] |
| 893 | 1 | Q8TDY2 | Q8TDY2 | RB1-inducibl | RB1CC1 | Homo sapien | 183.094 | 14 | 14 | [illegible] |
| 894 | 1 | Q8WUA2 | Q8WUA2 | Peptidyl-prol | PPIL4 | Homo sapien | 57.225 | 14 | 14 | [illegible] |
| 895 | 1 | Q8WUM0 | Q8WUM0 | Nuclear pore | NUP133 | Homo sapien | 128.981 | 14 | 14 | [illegible] |
| 896 | 1 | Q8WUX9 | Q8WUX9 | Charged mul | CHMP7 | Homo sapien | 50.912 | 14 | 14 | [illegible] |
| 897 | 1 | Q8WVV4 | Q8WVV4 | Protein POF | POF1B | Homo sapien | 68.064 | 14 | 14 | [illegible] |
| 898 | 1 | Q8WWM7 | Q8WWM7 | Ataxin-2-like | ATXN2L | Homo sapien | 113.376 | 14 | 14 | [illegible] |
| 899 | 1 | Q92499 | Q92499 | ATP-depend | DDX1 | Homo sapien | 82.434 | 14 | 14 | [illegible] |
| 900 | 1 | Q92556 | Q92556 | Engulfment a | ELMO1 | Homo sapien | 83.832 | 15 | 14 | [illegible] |
| 901 | 1 | Q93050 | Q93050 | V-type proto | ATP6V0A1 | Homo sapien | 96.415 | 15 | 14 | [illegible] |
| 902 | 1 | Q96CW1 | Q96CW1 | AP-2 comple | AP2M1 | Homo sapien | 49.657 | 14 | 14 | [illegible] |
| 903 | 1 | Q96G03 | Q96G03 | Phosphopent | PGM2 | Homo sapien | 68.283 | 14 | 14 | [illegible] |
| 904 | 1 | Q96JB2 | Q96JB2 | Conserved o | COG3 | Homo sapien | 94.098 | 14 | 14 | [illegible] |
| 905 | 1 | Q96JM3 | Q96JM3 | Chromosom | CHAMP1 | Homo sapien | 89.102 | 14 | 14 | [illegible] |
| 906 | 1 | Q96RT1 | Q96RT1 | Erbin | ERBIN | Homo sapien | 158.299 | 14 | 14 | [illegible] |
| 907 | 1 | Q96RW7 | Q96RW7 | Hemicentin-1 | HMCN1 | Homo sapien | 613.4 | 14 | 14 | [illegible] |
| 908 | 1 | Q99501 | Q99501 | GAS2-like pr | GAS2L1 | Homo sapien | 72.718 | 14 | 14 | [illegible] |
| 909 | 1 | Q99683 | Q99683 | Mitogen-activ | MAP3K5 | Homo sapien | 154.537 | 17 | 14 | [illegible] |
| 910 | 1 | Q99959 | Q99959 | Plakophilin-2 | PKP2 | Homo sapien | 97.419 | 14 | 14 | [illegible] |
| 911 | 1 | Q9BTC0 | Q9BTC0 | Death-induce | DIDO1 | Homo sapien | 243.876 | 14 | 14 | [illegible] |
| 912 | 1 | Q9BUQ8 | Q9BUQ8 | Probable AT | DDX23 | Homo sapien | 95.582 | 14 | 14 | [illegible] |
| 913 | 1 | Q9H270 | Q9H270 | Vacuolar pro | VPS11 | Homo sapien | 107.839 | 14 | 14 | [illegible] |
| 914 | 1 | Q9H2D6 | Q9H2D6 | TRIO and F-a | TRIOBP | Homo sapien | 261.381 | 14 | 14 | [illegible] |
| 915 | 1 | Q9H2U1 | Q9H2U1 | ATP-depend | DHX36 | Homo sapien | 114.762 | 14 | 14 | [illegible] |
| 916 | 1 | Q9HCE0 | Q9HCE0 | Ectopic P gra | EPG5 | Homo sapien | 292.486 | 14 | 14 | [illegible] |
| 917 | 1 | Q9NPQ8 | Q9NPQ8 | Chaperone | RIC8A | Homo sapien | 59.706 | 15 | 14 | [illegible] |
| 918 | 1 | Q9NR50 | Q9NR50 | Translation in | EIF2B3 | Homo sapien | 50.241 | 14 | 14 | [illegible] |
| 919 | 1 | Q9NR99 | Q9NR99 | Matrix-remod | MXRA5 | Homo sapien | 312.157 | 14 | 14 | [illegible] |
| 920 | 1 | Q9P107 | Q9P107 | GEM-interac | GMIP | Homo sapien | 106.683 | 14 | 14 | [illegible] |
| 921 | 1 | Q9UEY8 | Q9UEY8 | Gamma-add | ADD3 | Homo sapien | 79.157 | 14 | 14 | [illegible] |
| 922 | 1 | Q9UG63 | Q9UG63 | ATP-binding | ABCF2 | Homo sapien | 71.292 | 14 | 14 | [illegible] |
| 923 | 1 | Q9UJU6 | Q9UJU6 | Drebrin-like p | DBNL | Homo sapien | 48.207 | 14 | 14 | [illegible] |
| 924 | 1 | Q9UQ80 | Q9UQ80 | Proliferation- | PA2G4 | Homo sapien | 43.785 | 14 | 14 | [illegible] |
| 925 | 1 | Q9Y2D4 | Q9Y2D4 | Exocyst com | EXOC6B | Homo sapien | 94.201 | 14 | 14 | [illegible] |
| 926 | 1 | Q9Y3I0 | Q9Y3I0 | RNA-splicing | RTCB | Homo sapien | 55.209 | 14 | 14 | [illegible] |
| 927 | 1 | Q9Y3X0 | Q9Y3X0 | Coiled-coil d | CCDC9 | Homo sapien | 59.703 | 14 | 14 | [illegible] |
| 928 | 1 | Q9Y5L0 | Q9Y5L0 | Transportin-3 | TNPO3 | Homo sapien | 104.205 | 14 | 14 | [illegible] |
| 929 | 1 | Q9Y606 | Q9Y606 | Pseudouridyl | PUS1 | Homo sapien | 47.471 | 14 | 14 | [illegible] |
| 930 | 1 | Q9Y6C2 | Q9Y6C2 | EMILIN-1 | EMILIN1 | Homo sapien | 106.696 | 14 | 14 | [illegible] |
| 931 | 1 | A7E2V4 | A7E2V4 | Zinc finger S | ZSWIM8 | Homo sapien | 197.301 | 13 | 13 | [illegible] |
| 932 | 1 | O00139 | O00139 | Kinesin-like p | KIF2A | Homo sapien | 79.955 | 14 | 13 | [illegible] |
| 933 | 1 | O00186 | O00186 | Syntaxin-bin | STXBP3 | Homo sapien | 67.766 | 13 | 13 | [illegible] |
| 934 | 1 | O14727 | O14727 | Apoptotic pro | APAF1 | Homo sapien | 141.843 | 13 | 13 | [illegible] |
| 935 | 1 | O14744 | O14744 | Protein argin | PRMT5 | Homo sapien | 72.684 | 13 | 13 | [illegible] |
| 936 | 1 | O15484 | O15484 | Calpain-5 | CAPN5 | Homo sapien | 73.171 | 13 | 13 | [illegible] |
| 937 | 1 | O15553 | O15553 | Pyrin | MEFV | Homo sapien | 86.443 | 13 | 13 | [illegible] |
| 938 | 1 | O43150 | O43150 | Arf-GAP with | ASAP2 | Homo sapien | 111.65 | 13 | 13 | [illegible] |
| 939 | 1 | O43390 | O43390 | Heterogeneo | HNRNPR | Homo sapien | 70.943 | 17 | 13 | [illegible] |
| 940 | 1 | O43592 | O43592 | Exportin-T | XPOT | Homo sapien | 109.967 | 13 | 13 | [illegible] |
| 941 | 1 | O60506 | O60506 | Heterogeneo | SYNCRIP | Homo sapien | 69.605 | 13 | 13 | [illegible] |
| 942 | 1 | O60890 | O60890 | Oligophrenin- | OPHN1 | Homo sapien | 91.643 | 13 | 13 | [illegible] |
| 943 | 1 | O75069 | O75069 | Transmembr | TMCC2 | Homo sapien | 77.448 | 16 | 13 | [illegible] |
| 944 | 1 | O75694 | O75694 | Nuclear pore | NUP155 | Homo sapien | 155.203 | 13 | 13 | [illegible] |
| 945 | 1 | O75962 | O75962 | Triple functio | TRIO | Homo sapien | 346.905 | 13 | 13 | [illegible] |
| 946 | 1 | O76094 | O76094 | Signal recog | SRP72 | Homo sapien | 74.609 | 13 | 13 | [illegible] |
| 947 | 1 | O94822 | O94822 | E3 ubiquitin-p | LTN1 | Homo sapien | 200.553 | 13 | 13 | [illegible] |
| 948 | 1 | O95155 | O95155 | Ubiquitin con | UBE4B | Homo sapien | 146.184 | 13 | 13 | [illegible] |
| 949 | 1 | O95202 | O95202 | Mitochondria | LETM1 | Homo sapien | 83.357 | 13 | 13 | [illegible] |
| 950 | 1 | P01133 | P01133 | Pro-epiderm | EGF | Homo sapien | 133.996 | 13 | 13 | [illegible] |
| 951 | 1 | P03951 | P03951 | Coagulation f | F11 | Homo sapien | 70.109 | 13 | 13 | [illegible] |
| 952 | 1 | P04406 | P04406 | Glyceraldehy | GAPDH | Homo sapien | 36.054 | 13 | 13 | [illegible] |
| 953 | 1 | P05546 | P05546 | Heparin cofa | SERPIND1 | Homo sapien | 57.071 | 13 | 13 | [illegible] |
| 954 | 1 | P10643 | P10643 | Complement | C7 | Homo sapien | 93.519 | 13 | 13 | [illegible] |
| 955 | 1 | P14317 | P14317 | Hematopoiet | HCLS1 | Homo sapien | 54.014 | 13 | 13 | [illegible] |
| 956 | 1 | P14780 | P14780 | Matrix metall | MMP9 | Homo sapien | 78.458 | 13 | 13 | [illegible] |
| 957 | 1 | P15311 | P15311 | Ezrin | EZR | Homo sapien | 69.414 | 13 | 13 | [illegible] |
| 958 | 1 | P19013 | P19013 | Keratin, type | KRT4 | Homo sapien | 56.146 | 20 | 13 | [illegible] |
| 959 | 1 | P19021 | P19021 | Peptidyl-glyc | PAM | Homo sapien | 108.333 | 13 | 13 | [illegible] |
| 960 | 1 | P19367 | P19367 | Hexokinase- | HK1 | Homo sapien | 102.489 | 16 | 13 | [illegible] |
| 961 | 1 | P22307 | P22307 | Sterol carrier | SCP2 | Homo sapien | 58.995 | 13 | 13 | [illegible] |
| 962 | 1 | P23229 | P23229 | Integrin alpha | ITGA6 | Homo sapien | 126.604 | 13 | 13 | [illegible] |
| 963 | 1 | P23368 | P23368 | NAD-depend | ME2 | Homo sapien | 65.442 | 13 | 13 | [illegible] |
| 964 | 1 | P23786 | P23786 | Carnitine O-p | CPT2 | Homo sapien | 73.779 | 13 | 13 | [illegible] |
| 965 | 1 | P25205 | P25205 | DNA replicat | MCM3 | Homo sapien | 90.981 | 13 | 13 | [illegible] |
| 966 | 1 | P25311 | P25311 | Zinc-alpha-2- | AZGP1 | Homo sapien | 34.26 | 13 | 13 | [illegible] |
| 967 | 1 | P32019 | P32019 | Type II inosit | INPP5B | Homo sapien | 112.853 | 13 | 13 | [illegible] |
| 968 | 1 | P36871 | P36871 | Phosphogluc | PGM1 | Homo sapien | 61.45 | 13 | 13 | [illegible] |
| 969 | 1 | P41218 | P41218 | Myeloid cell n | MNDA | Homo sapien | 45.837 | 13 | 13 | [illegible] |
| 970 | 1 | P42285 | P42285 | Exosome RN | MTREX | Homo sapien | 117.807 | 13 | 13 | [illegible] |
| 971 | 1 | P43034 | P43034 | Platelet-activ | PAFAH1B1 | Homo sapien | 46.639 | 13 | 13 | [illegible] |
| 972 | 1 | P43378 | P43378 | Tyrosine-pro | PTPN9 | Homo sapien | 68.021 | 13 | 13 | [illegible] |
| 973 | 1 | P48643 | P48643 | T-complex pr | CCT5 | Homo sapien | 59.672 | 13 | 13 | [illegible] |
| 974 | 1 | P49411 | P49411 | Elongation fa | TUFM | Homo sapien | 49.876 | 13 | 13 | [illegible] |
| 975 | 1 | P50552 | P50552 | Vasodilator-s | VASP | Homo sapien | 39.83 | 13 | 13 | [illegible] |
| 976 | 1 | P51784 | P51784 | Ubiquitin car | USP11 | Homo sapien | 109.817 | 14 | 13 | [illegible] |
| 977 | 1 | P54619 | P54619 | 5'-AMP-activ | PRKAG1 | Homo sapien | 37.579 | 14 | 13 | [illegible] |
| 978 | 1 | P60228 | P60228 | Eukaryotic tr | EIF3E | Homo sapien | 52.223 | 13 | 13 | [illegible] |
| 979 | 1 | P61011 | P61011 | Signal recog | SRP54 | Homo sapien | 55.703 | 13 | 13 | [illegible] |
| 980 | 1 | P61201 | P61201 | COP9 signal | COPS2 | Homo sapien | 51.597 | 13 | 13 | [illegible] |
| 981 | 1 | P62195 | P62195 | 26S proteas | PSMC5 | Homo sapien | 45.624 | 13 | 13 | [illegible] |
| 982 | 1 | P78509 | P78509 | Reelin | RELN | Homo sapien | 388.395 | 13 | 13 | [illegible] |
| 983 | 1 | Q00839 | Q00839 | Heterogeneo | HNRNPU | Homo sapien | 90.588 | 13 | 13 | [illegible] |
| 984 | 1 | Q01433 | Q01433 | AMP deamin | AMPD2 | Homo sapien | 94.893 | 13 | 13 | [illegible] |
| 985 | 1 | Q01546 | Q01546 | Keratin, type | KRT76 | Homo sapien | 65.844 | 15 | 13 | [illegible] |
| 986 | 1 | Q01813 | Q01813 | ATP-depend | PFKP | Homo sapien | 85.596 | 13 | 13 | [illegible] |
| 987 | 1 | Q02413 | Q02413 | Desmoglein-1 | DSG1 | Homo sapien | 113.749 | 13 | 13 | [illegible] |
| 988 | 1 | Q02952 | Q02952 | A-kinase anc | AKAP12 | Homo sapien | 191.487 | 13 | 13 | [illegible] |
| 989 | 1 | Q04759 | Q04759 | Protein kinas | PRKCQ | Homo sapien | 81.865 | 13 | 13 | [illegible] |
| 990 | 1 | Q12851 | Q12851 | Mitogen-activ | MAP4K2 | Homo sapien | 91.559 | 13 | 13 | [illegible] |
| 991 | 1 | Q12959 | Q12959 | Disks large h | DLG1 | Homo sapien | 100.456 | 14 | 13 | [illegible] |
| 992 | 1 | Q13136 | Q13136 | Liprin-alpha-1 | PPFIA1 | Homo sapien | 135.779 | 17 | 13 | [illegible] |
| 993 | 1 | Q13418 | Q13418 | Scaffold prot | ILK | Homo sapien | 51.419 | 13 | 13 | [illegible] |
| 994 | 1 | Q13596 | Q13596 | Sorting nexin | SNX1 | Homo sapien | 59.069 | 13 | 13 | [illegible] |
| 995 | 1 | Q14149 | Q14149 | MORC family | MORC3 | Homo sapien | 107.115 | 13 | 13 | [illegible] |
| 996 | 1 | Q14156 | Q14156 | Protein EFR3 | EFR3A | Homo sapien | 92.925 | 14 | 13 | [illegible] |
| 997 | 1 | Q14160 | Q14160 | Protein scrib | SCRIB | Homo sapien | 177.727 | 13 | 13 | [illegible] |
| 998 | 1 | Q14694 | Q14694 | Ubiquitin car | USP10 | Homo sapien | 87.137 | 13 | 13 | [illegible] |
| 999 | 1 | Q15027 | Q15027 | Arf-GAP with | ACAP1 | Homo sapien | 81.537 | 13 | 13 | [illegible] |
| 1000 | 1 | Q15154 | Q15154 | Pericentriolar | PCM1 | Homo sapien | 228.562 | 13 | 13 | [illegible] |
| 1001 | 1 | Q15334 | Q15334 | Lethal(2) gia | LLGL1 | Homo sapien | 115.419 | 13 | 13 | [illegible] |
| 1002 | 1 | Q15642 | Q15642 | Cdc42-intera | TRIP10 | Homo sapien | 68.351 | 13 | 13 | [illegible] |
| 1003 | 1 | Q16881 | Q16881 | Thioredoxin r | TXNRD1 | Homo sapien | 70.876 | 13 | 13 | [illegible] |
| 1004 | 1 | Q1KMD3 | Q1KMD3 | Heterogeneo | HNRNPUL2 | Homo sapien | 85.105 | 13 | 13 | [illegible] |
| 1005 | 1 | Q4G0J3 | Q4G0J3 | La-related pr | LARP7 | Homo sapien | 66.898 | 13 | 13 | [illegible] |
| 1006 | 1 | Q53GL7 | Q53GL7 | Protein mono | PARP10 | Homo sapien | 110 | 13 | 13 | [illegible] |
| 1007 | 1 | Q5JRX3 | Q5JRX3 | Presequence | PITRM1 | Homo sapien | 117.415 | 13 | 13 | [illegible] |
| 1008 | 1 | Q5TC84 | Q5TC84 | Opioid growt | OGFRL1 | Homo sapien | 51.251 | 13 | 13 | [illegible] |
| 1009 | 1 | Q5TCZ1 | Q5TCZ1 | SH3 and PX | SH3PXD2A | Homo sapien | 125.292 | 14 | 13 | [illegible] |
| 1010 | 1 | Q6UWE0 | Q6UWE0 | E3 ubiquitin-p | LRSAM1 | Homo sapien | 83.593 | 13 | 13 | [illegible] |
| 1011 | 1 | Q7L2E3 | Q7L2E3 | ATP-depend | DHX30 | Homo sapien | 133.939 | 13 | 13 | [illegible] |
| 1012 | 1 | Q7LBC6 | Q7LBC6 | Lysine-specif | KDM3B | Homo sapien | 191.584 | 14 | 13 | [illegible] |
| 1013 | 1 | Q7Z591 | Q7Z591 | Microtubule o | AKNA | Homo sapien | 155.141 | 13 | 13 | [illegible] |
| 1014 | 1 | Q86TI2 | Q86TI2 | Dipeptidyl pe | DPP9 | Homo sapien | 98.264 | 13 | 13 | [illegible] |
| 1015 | 1 | Q86XA9 | Q86XA9 | HEAT repeat | HEATR5A | Homo sapien | 222.006 | 13 | 13 | [illegible] |
| 1016 | 1 | Q86YS7 | Q86YS7 | C2 domain-c | C2CD5 | Homo sapien | 110.448 | 13 | 13 | [illegible] |
| 1017 | 1 | Q8IWF6 | Q8IWF6 | Protein DEN | DENND6A | Homo sapien | 69.575 | 13 | 13 | [illegible] |
| 1018 | 1 | Q8IXQ6 | Q8IXQ6 | Protein mono | PARP9 | Homo sapien | 96.345 | 13 | 13 | [illegible] |
| 1019 | 1 | Q8IXT5 | Q8IXT5 | RNA-binding | RBM12B | Homo sapien | 118.103 | 13 | 13 | [illegible] |
| 1020 | 1 | Q8N1G4 | Q8N1G4 | Leucine-rich | LRRC47 | Homo sapien | 63.473 | 13 | 13 | [illegible] |
| 1021 | 1 | Q8NEN9 | Q8NEN9 | PDZ domain- | PDZD8 | Homo sapien | 128.563 | 13 | 13 | [illegible] |
| 1022 | 1 | Q8TAT6 | Q8TAT6 | Nuclear prot | NPLOC4 | Homo sapien | 68.121 | 13 | 13 | [illegible] |
| 1023 | 1 | Q8TBA6 | Q8TBA6 | Golgin subfa | GOLGA5 | Homo sapien | 83.025 | 13 | 13 | [illegible] |
| 1024 | 1 | Q8TC07 | Q8TC07 | TBC1 domai | TBC1D15 | Homo sapien | 79.49 | 13 | 13 | [illegible] |
| 1025 | 1 | Q8TDF6 | Q8TDF6 | RAS guanyl-r | RASGRP4 | Homo sapien | 74.885 | 13 | 13 | [illegible] |
| 1026 | 1 | Q8TDZ2 | Q8TDZ2 | [F-actin]-mo | MICAL1 | Homo sapien | 117.875 | 13 | 13 | [illegible] |
| 1027 | 1 | Q8WUJ3 | Q8WUJ3 | Cell migratio | CEMIP | Homo sapien | 152.999 | 13 | 13 | [illegible] |
| 1028 | 1 | Q8WVM8 | Q8WVM8 | Sec1 family | SCFD1 | Homo sapien | 72.381 | 13 | 13 | [illegible] |
| 1029 | 1 | Q92539 | Q92539 | Phosphatida | LPIN2 | Homo sapien | 99.4 | 14 | 13 | [illegible] |
| 1030 | 1 | Q92696 | Q92696 | Geranylgera | RABGGTA | Homo sapien | 65.075 | 13 | 13 | [illegible] |
| 1031 | 1 | Q96AE4 | Q96AE4 | Far upstream | FUBP1 | Homo sapien | 67.561 | 13 | 13 | [illegible] |
| 1032 | 1 | Q96CW5 | Q96CW5 | Gamma-tubu | TUBGCP3 | Homo sapien | 103.571 | 13 | 13 | [illegible] |
| 1033 | 1 | Q96FS4 | Q96FS4 | Signal-induce | SIPA1 | Homo sapien | 112.149 | 13 | 13 | [illegible] |
| 1034 | 1 | Q96GQ7 | Q96GQ7 | Probable AT | DDX27 | Homo sapien | 89.834 | 13 | 13 | [illegible] |
| 1035 | 1 | Q96L91 | Q96L91 | E1A-binding | EP400 | Homo sapien | 343.493 | 13 | 13 | [illegible] |
| 1036 | 1 | Q96NW4 | Q96NW4 | Ankyrin repe | ANKRD27 | Homo sapien | 116.988 | 13 | 13 | [illegible] |
| 1037 | 1 | Q96PE3 | Q96PE3 | Inositol polyp | INPP4A | Homo sapien | 109.956 | 13 | 13 | [illegible] |
| 1038 | 1 | Q96PU5 | Q96PU5 | E3 ubiquitin-p | NEDD4L | Homo sapien | 111.933 | 17 | 13 | [illegible] |
| 1039 | 1 | Q96SB3 | Q96SB3 | Neurabin-2 | PPP1R9B | Homo sapien | 89.336 | 14 | 13 | [illegible] |
| 1040 | 1 | Q96T37 | Q96T37 | RNA-binding | RBM15 | Homo sapien | 107.191 | 13 | 13 | [illegible] |
| 1041 | 1 | Q96T76 | Q96T76 | MMS19 nucl | MMS19 | Homo sapien | 113.292 | 13 | 13 | [illegible] |
| 1042 | 1 | Q99590 | Q99590 | Protein SCAF | SCAF11 | Homo sapien | 164.654 | 13 | 13 | [illegible] |
| 1043 | 1 | Q9BVA0 | Q9BVA0 | Katanin p80 | KATNB1 | Homo sapien | 72.335 | 13 | 13 | [illegible] |
| 1044 | 1 | Q9BYC5 | Q9BYC5 | Alpha-(1,6)-f | FUT8 | Homo sapien | 66.515 | 13 | 13 | [illegible] |
| 1045 | 1 | Q9H0D6 | Q9H0D6 | 5'-3' exoribon | XRN2 | Homo sapien | 108.582 | 13 | 13 | [illegible] |

| | | | | | | | | | | | | | | | | | | | | |
|---|---|---|---|---|---|---|---|---|---|---|---|---|---|---|---|---|---|---|---|---|
| 1046 | 1 Q9H269 | Q9H269 | Vacuolar pro VPS16 | Homo sapien | 94.691 | 13 | 13 | 1.0E+06 | 7.8E+05 | 1.3E+06 | 1.4E+06 | 1.3E+06 | 1.2E+06 | 2.1E+06 | 1.5E+06 | 1.9E+06 | 1.8E+06 | 2.3E+06 | 1.7E+06 | [illegible] |
| 1047 | 1 Q9H4A4 | Q9H4A4 | Aminopeptida RNPEP | Homo sapien | 72.598 | 13 | 13 | 1.8E+06 | 1.8E+06 | 1.6E+06 | 2.0E+06 | 2.2E+06 | 1.5E+06 | 4.5E+06 | 4.0E+06 | 2.9E+06 | 5.9E+06 | 4.3E+06 | 6.2E+06 | [illegible] |
| 1048 | 1 Q9H4M9 | Q9H4M9 | EH domain-c EHD1 | Homo sapien | 60.627 | 25 | 13 | 3.8E+07 | 3.1E+07 | 2.1E+07 | 3.1E+07 | 3.9E+07 | 4.3E+07 | 3.5E+07 | 3.7E+07 | 3.1E+07 | 3.8E+07 | 4.0E+07 | 4.7E+07 | [illegible] |
| 1049 | 1 Q9NPB8 | Q9NPB8 | Glycerophos GPCPD1 | Homo sapien | 76.035 | 13 | 13 | 1.2E+06 | 1.5E+06 | 1.2E+06 | 9.0E+05 | 2.7E+06 | 7.4E+05 | 6.3E+06 | 1.2E+06 | 2.1E+06 | 2.7E+06 | 2.3E+06 | 6.5E+06 | [illegible] |
| 1050 | 1 Q9NRK6 | Q9NRK6 | ATP-binding ABCB10 | Homo sapien | 79.149 | 13 | 13 | 3.1E+05 | 2.2E+05 | 4.3E+05 | 1.7E+05 | 7.0E+05 | 2.0E+05 | 1.7E+05 | 2.0E+05 | 1.6E+05 | 2.6E+05 | 1.9E+05 | 2.1E+05 | [illegible] |
| 1051 | 1 Q9NUQ8 | Q9NUQ8 | ATP-binding ABCF3 | Homo sapien | 79.746 | 13 | 13 | 1.1E+06 | 1.1E+06 | 8.2E+05 | 8.3E+05 | 9.3E+05 | 6.9E+05 | 4.6E+05 | 5.7E+05 | 6.0E+05 | 7.1E+05 | 5.9E+05 | 5.5E+05 | [illegible] |
| 1052 | 1 Q9NXF1 | Q9NXF1 | Testis-expre TEX10 | Homo sapien | 105.674 | 13 | 13 | 2.1E+06 | 2.1E+06 | 2.6E+06 | 2.3E+06 | 3.3E+06 | 9.6E+03 | 0.0E+00 | 5.8E+05 | 2.8E+04 | 4.8E+05 | 1.0E+04 | 2.0E+04 | [illegible] |
| 1053 | 1 Q9UBB9 | Q9UBB9 | Tuftelin-inter TFIP11 | Homo sapien | 96.821 | 13 | 13 | 1.4E+05 | 2.1E+06 | 1.7E+06 | 5.4E+04 | 2.4E+06 | 1.5E+06 | 1.6E+06 | 7.1E+04 | 5.3E+04 | 6.5E+04 | 1.7E+04 | 2.5E+04 | [illegible] |
| 1054 | 1 Q9UBW5 | Q9UBW5 | Bridging integ BIN2 | Homo sapien | 61.877 | 14 | 13 | 5.6E+07 | 6.3E+07 | 6.8E+07 | 5.0E+07 | 4.9E+07 | 5.0E+07 | 6.1E+07 | 6.2E+07 | 5.1E+07 | 6.6E+07 | 5.6E+07 | 6.8E+07 | [illegible] |
| 1055 | 1 Q9UDY6 | Q9UDY6 | Tripartite mot TRIM10 | Homo sapien | 55.036 | 13 | 13 | 7.4E+05 | 1.2E+06 | 7.9E+05 | 1.2E+06 | 1.1E+06 | 1.2E+06 | 1.5E+06 | 9.5E+05 | 1.2E+06 | 1.6E+06 | 1.7E+06 | 1.8E+06 | [illegible] |
| 1056 | 1 Q9UHI6 | Q9UHI6 | Probable ATF DDX20 | Homo sapien | 92.239 | 13 | 13 | 0.0E+00 | 2.1E+04 | 3.2E+04 | 5.3E+03 | 3.8E+04 | 3.6E+03 | 0.0E+00 | 4.7E+03 | 2.5E+04 | 5.8E+04 | 3.6E+04 | 3.0E+04 | [illegible] |
| 1057 | 1 Q9UJX4 | Q9UJX4 | Anaphase-pro ANAPC5 | Homo sapien | 85.079 | 13 | 13 | 9.4E+05 | 1.1E+06 | 8.6E+05 | 1.2E+06 | 9.4E+05 | 1.1E+06 | 1.0E+05 | 5.1E+05 | 8.9E+03 | 2.4E+04 | 7.1E+05 | 3.2E+04 | [illegible] |
| 1058 | 1 Q9UPQ9 | Q9UPQ9 | Trinucleotide TNRC6B | Homo sapien | 194.004 | 13 | 13 | 4.0E+05 | 6.3E+05 | 1.8E+05 | 2.9E+04 | 1.6E+06 | 1.1E+06 | 9.5E+05 | 1.2E+05 | 0.0E+00 | 0.0E+00 | 0.0E+00 | 7.1E+05 | [illegible] |
| 1059 | 1 Q9Y263 | Q9Y263 | Phospholipas PLAA | Homo sapien | 87.158 | 13 | 13 | 1.1E+07 | 4.4E+05 | 1.1E+07 | 2.0E+07 | 6.5E+05 | 5.3E+05 | 1.2E+06 | 1.1E+06 | 1.5E+07 | 8.6E+05 | 1.4E+06 | 1.1E+06 | [illegible] |
| 1060 | 1 Q9Y2L6 | Q9Y2L6 | FERM domai FRMD4B | Homo sapien | 118.049 | 13 | 13 | 6.2E+06 | 6.4E+06 | 5.1E+06 | 6.1E+06 | 3.9E+06 | 5.6E+06 | 3.3E+06 | 4.0E+06 | 4.0E+06 | 3.4E+06 | 2.0E+06 | 2.4E+06 | [illegible] |
| 1061 | 1 Q9Y4F3 | Q9Y4F3 | Meiosis regul MARF1 | Homo sapien | 192.863 | 13 | 13 | 1.5E+05 | 8.5E+05 | 6.8E+06 | 9.1E+06 | 1.2E+07 | 1.1E+07 | 8.6E+05 | 8.8E+05 | 4.3E+04 | 1.7E+07 | 9.8E+05 | 7.6E+05 | [illegible] |
| 1062 | 1 Q9Y4H2 | Q9Y4H2 | Insulin recep IRS2 | Homo sapien | 137.337 | 13 | 13 | 9.6E+06 | 9.2E+06 | 6.7E+06 | 7.3E+06 | 9.3E+06 | 8.2E+06 | 8.3E+06 | 8.2E+06 | 9.2E+06 | 8.0E+06 | 8.0E+06 | 8.2E+06 | [illegible] |
| 1063 | 1 Q9Y5X1 | Q9Y5X1 | Sorting nexin SNX9 | Homo sapien | 66.593 | 13 | 13 | 3.3E+07 | 3.6E+07 | 2.2E+07 | 3.0E+07 | 2.9E+07 | 3.1E+07 | 1.6E+07 | 1.7E+07 | 1.8E+07 | 1.3E+07 | 1.5E+07 | 1.5E+07 | [illegible] |
| 1064 | 1 Q9Y5X2 | Q9Y5X2 | Sorting nexin SNX8 | Homo sapien | 52.571 | 13 | 13 | 3.0E+06 | 2.5E+06 | 3.5E+06 | 2.7E+06 | 1.9E+06 | 3.0E+06 | 2.3E+06 | 8.8E+05 | 1.2E+06 | 1.3E+06 | 3.9E+06 | 1.1E+06 | [illegible] |
| 1065 | 1 O00116 | O00116 | Alkyldihydrox AGPS | Homo sapien | 72.909 | 12 | 12 | 6.4E+06 | 6.3E+06 | 6.3E+06 | 5.3E+06 | 4.3E+06 | 4.4E+06 | 5.7E+06 | 4.8E+06 | 4.4E+06 | 6.0E+06 | 7.8E+06 | 4.7E+06 | [illegible] |
| 1066 | 1 O00329 | O00329 | Phosphatidyl PIK3CD | Homo sapien | 119.479 | 12 | 12 | 1.1E+07 | 1.1E+07 | 2.4E+07 | 6.7E+04 | 3.5E+06 | 2.0E+06 | 9.5E+06 | 5.2E+04 | 1.5E+07 | 2.5E+06 | 1.2E+07 | 2.6E+06 | [illegible] |
| 1067 | 1 O00461 | O00461 | Golgi integral GOLIM4 | Homo sapien | 81.881 | 12 | 12 | 6.7E+04 | 9.2E+03 | 3.3E+04 | 1.9E+04 | 8.5E+03 | 1.7E+04 | 6.3E+05 | 7.1E+04 | 7.7E+04 | 1.0E+06 | 2.4E+04 | 8.0E+04 | [illegible] |
| 1068 | 1 O14523 | O14523 | Phospholipid C2CD2L | Homo sapien | 76.182 | 12 | 12 | 1.5E+07 | 2.2E+07 | 1.5E+07 | 2.3E+07 | 2.2E+07 | 2.3E+07 | 1.6E+07 | 1.9E+07 | 1.7E+07 | 1.7E+07 | 1.5E+07 | 1.6E+07 | [illegible] |
| 1069 | 1 O14920 | O14920 | Inhibitor of nu IKBKB | Homo sapien | 86.565 | 12 | 12 | 4.5E+05 | 6.9E+05 | 1.3E+06 | 1.3E+06 | 1.2E+06 | 9.4E+05 | 1.8E+06 | 2.0E+06 | 1.6E+06 | 2.2E+06 | 2.4E+06 | 1.6E+06 | [illegible] |
| 1070 | 1 O15031 | O15031 | Plexin-B2 PLXNB2 | Homo sapien | 205.13 | 12 | 12 | 3.6E+04 | 4.8E+05 | 7.4E+04 | 7.1E+04 | 6.7E+04 | 5.0E+05 | 5.7E+04 | 3.6E+04 | 1.7E+05 | 5.4E+04 | 1.5E+05 | 2.6E+04 | [illegible] |
| 1071 | 1 O15084 | O15084 | Serine/threon ANKRD28 | Homo sapien | 112.967 | 13 | 12 | 1.0E+06 | 8.0E+05 | 7.0E+05 | 1.1E+06 | 8.6E+05 | 8.3E+05 | 1.8E+06 | 1.3E+06 | 1.1E+06 | 1.8E+06 | 1.8E+06 | 1.7E+06 | [illegible] |
| 1072 | 1 O43395 | O43395 | U4/U6 small r PRPF3 | Homo sapien | 77.528 | 12 | 12 | 2.4E+06 | 1.5E+06 | 3.0E+06 | 2.2E+06 | 3.1E+06 | 1.9E+06 | 5.6E+05 | 1.0E+06 | 9.0E+05 | 1.8E+06 | 1.1E+06 | 7.0E+05 | [illegible] |
| 1073 | 1 O43615 | O43615 | Mitochondrial TIMM44 | Homo sapien | 51.356 | 12 | 12 | 1.2E+06 | 1.4E+06 | 1.2E+06 | 1.3E+06 | 1.3E+06 | 8.3E+05 | 1.5E+06 | 1.1E+06 | 1.6E+06 | 1.2E+06 | 1.2E+06 | 1.4E+06 | [illegible] |
| 1074 | 1 O60231 | O60231 | Pre-mRNA-s DHX16 | Homo sapien | 119.264 | 12 | 12 | 6.7E+06 | 6.3E+06 | 7.0E+06 | 6.1E+06 | 8.7E+06 | 7.5E+06 | 7.2E+04 | 5.8E+06 | 1.1E+05 | 1.8E+05 | 7.7E+04 | 1.8E+05 | [illegible] |
| 1075 | 1 O60307 | O60307 | Microtubule-a MAST3 | Homo sapien | 143.138 | 14 | 12 | 6.6E+04 | 3.9E+06 | 2.1E+06 | 1.6E+05 | 1.5E+05 | 4.6E+06 | 3.2E+06 | 2.5E+06 | 2.5E+05 | 1.3E+05 | 5.5E+06 | 1.1E+05 | [illegible] |
| 1076 | 1 O60331 | O60331 | Phosphatidyl PIP5K1C | Homo sapien | 73.26 | 13 | 12 | 1.2E+07 | 1.8E+07 | 1.5E+07 | 1.6E+07 | 1.3E+07 | 1.5E+07 | 1.2E+07 | 1.1E+07 | 1.4E+07 | 1.0E+07 | 1.1E+07 | 1.5E+07 | [illegible] |
| 1077 | 1 O60333 | O60333 | Kinesin-like p KIF1B | Homo sapien | 204.481 | 20 | 12 | 8.9E+06 | 1.8E+05 | 2.5E+05 | 1.8E+05 | 1.9E+05 | 1.4E+05 | 1.2E+05 | 2.1E+05 | 1.4E+05 | 1.4E+05 | 1.6E+05 | 1.7E+05 | [illegible] |
| 1078 | 1 O60749 | O60749 | Sorting nexin SNX2 | Homo sapien | 58.471 | 14 | 12 | 1.6E+07 | 1.4E+07 | 1.2E+07 | 1.8E+07 | 1.3E+07 | 1.1E+07 | 1.2E+07 | 5.7E+06 | 7.7E+06 | 1.1E+07 | 1.3E+07 | 1.0E+07 | [illegible] |
| 1079 | 1 O60832 | O60832 | H/ACA ribonu DKC1 | Homo sapien | 57.673 | 12 | 12 | 1.3E+06 | 7.4E+05 | 1.2E+06 | 1.0E+06 | 2.0E+06 | 1.0E+06 | 2.7E+05 | 6.7E+05 | 5.5E+05 | 6.6E+05 | 8.2E+05 | 6.3E+05 | [illegible] |
| 1080 | 1 O60879 | O60879 | Protein diaph DIAPH2 | Homo sapien | 125.571 | 12 | 12 | 3.4E+05 | 1.4E+06 | 4.4E+05 | 2.3E+05 | 2.4E+05 | 4.4E+05 | 1.2E+06 | 5.1E+05 | 1.0E+06 | 3.8E+05 | 5.7E+05 | 3.9E+05 | [illegible] |
| 1081 | 1 O75064 | O75064 | DENN domai DENND4B | Homo sapien | 163.846 | 12 | 12 | 1.2E+06 | 1.0E+06 | 9.9E+05 | 1.4E+06 | 1.3E+06 | 1.2E+06 | 5.4E+05 | 3.0E+05 | 4.0E+05 | 7.0E+05 | 7.3E+05 | 6.8E+05 | [illegible] |
| 1082 | 1 O75146 | O75146 | Huntingtin-int HIP1R | Homo sapien | 119.39 | 13 | 12 | 0.0E+00 | 0.0E+00 | 4.4E+04 | 0.0E+00 | 4.0E+04 | 3.3E+04 | 0.0E+00 | 8.1E+04 | 0.0E+00 | 0.0E+00 | 8.9E+03 | 0.0E+00 | [illegible] |
| 1083 | 1 O75400 | O75400 | Pre-mRNA-p PRPF40A | Homo sapien | 108.806 | 12 | 12 | 7.4E+06 | 3.4E+06 | 8.7E+06 | 1.4E+07 | 5.6E+06 | 1.7E+06 | 6.1E+06 | 7.3E+06 | 7.0E+06 | 8.3E+06 | 5.4E+06 | 6.0E+06 | [illegible] |
| 1084 | 1 O75563 | O75563 | Src kinase-as SKAP2 | Homo sapien | 41.218 | 12 | 12 | 6.2E+07 | 6.0E+07 | 4.9E+07 | 4.4E+07 | 4.1E+07 | 3.8E+07 | 3.8E+07 | 3.7E+07 | 4.3E+07 | 4.0E+07 | 3.8E+07 | 3.4E+07 | [illegible] |
| 1085 | 1 O75822 | O75822 | Eukaryotic tra EIF3J | Homo sapien | 29.063 | 12 | 12 | 2.7E+08 | 3.5E+08 | 3.1E+08 | 3.5E+08 | 2.3E+08 | 2.6E+08 | 2.8E+08 | 1.3E+08 | 1.5E+08 | 2.8E+08 | 2.7E+08 | 1.9E+08 | [illegible] |
| 1086 | 1 O95714 | O95714 | E3 ubiquitin-p HERC2 | Homo sapien | 527.236 | 13 | 12 | 2.1E+04 | 5.6E+05 | 1.3E+04 | 1.6E+04 | 4.4E+05 | 2.2E+04 | 2.0E+04 | 0.0E+00 | 3.4E+04 | 6.1E+05 | 0.0E+00 | 4.5E+05 | [illegible] |
| 1087 | 1 P00491 | P00491 | Purine nucleo PNP | Homo sapien | 32.118 | 12 | 12 | 1.4E+08 | 1.5E+08 | 1.2E+08 | 1.6E+08 | 1.4E+08 | 1.1E+08 | 2.1E+08 | 1.9E+08 | 1.3E+08 | 2.1E+08 | 2.1E+08 | 2.1E+08 | [illegible] |
| 1088 | 1 P01011 | P01011 | Alpha-1-antic SERPINA3 | Homo sapien | 47.65 | 12 | 12 | 4.9E+07 | 9.7E+07 | 4.1E+07 | 3.8E+07 | 5.5E+07 | 3.5E+07 | 7.2E+07 | 4.5E+07 | 5.6E+07 | 5.9E+07 | 1.2E+08 | 3.1E+07 | [illegible] |
| 1089 | 1 P02748 | P02748 | Complement C9 | Homo sapien | 63.175 | 12 | 12 | 3.3E+08 | 2.6E+08 | 2.3E+08 | 2.1E+08 | 1.5E+08 | 1.5E+08 | 2.5E+08 | 1.5E+08 | 2.2E+08 | 3.2E+08 | 4.0E+08 | 1.9E+08 | [illegible] |
| 1090 | 1 P04075 | P04075 | Fructose-bis ALDOA | Homo sapien | 39.418 | 13 | 12 | 1.3E+08 | 1.5E+08 | 1.0E+08 | 1.7E+08 | 1.2E+08 | 9.4E+07 | 2.6E+08 | 1.7E+08 | 2.3E+08 | 2.4E+08 | 3.6E+08 | 3.2E+08 | [illegible] |
| 1091 | 1 P04424 | P04424 | Argininosucci ASL | Homo sapien | 51.66 | 12 | 12 | 8.1E+05 | 8.9E+05 | 8.3E+05 | 1.0E+06 | 7.2E+05 | 9.2E+05 | 1.2E+06 | 1.5E+06 | 1.0E+06 | 1.6E+06 | 1.8E+06 | 1.7E+06 | [illegible] |
| 1092 | 1 P04844 | P04844 | Dolichyl-diph RPN2 | Homo sapien | 69.284 | 12 | 12 | 1.5E+07 | 1.2E+07 | 1.8E+07 | 1.6E+07 | 1.8E+07 | 1.7E+07 | 3.0E+07 | 2.6E+07 | 1.8E+07 | 3.0E+07 | 2.1E+07 | 3.8E+07 | [illegible] |
| 1093 | 1 P07093 | P07093 | Glia-derived r SERPINE2 | Homo sapien | 44.005 | 12 | 12 | 3.8E+06 | 4.4E+06 | 2.0E+06 | 1.8E+06 | 3.0E+06 | 1.1E+06 | 2.5E+06 | 2.2E+06 | 2.0E+06 | 2.1E+06 | 1.9E+06 | 2.4E+06 | [illegible] |
| 1094 | 1 P09972 | P09972 | Fructose-bis ALDOC | Homo sapien | 39.458 | 12 | 12 | 4.0E+05 | 6.1E+04 | 5.1E+05 | 1.3E+05 | 1.6E+05 | 5.5E+05 | 9.2E+05 | 2.1E+05 | 1.6E+05 | 1.6E+06 | 3.7E+05 | 2.8E+05 | [illegible] |
| 1095 | 1 P11142 | P11142 | Heat shock c HSPA8 | Homo sapien | 70.899 | 17 | 12 | 1.4E+08 | 1.1E+08 | 8.3E+07 | 1.3E+08 | 1.3E+08 | 1.2E+08 | 1.1E+08 | 1.3E+08 | 8.8E+07 | 1.3E+08 | 1.4E+08 | 1.6E+08 | [illegible] |
| 1096 | 1 P11597 | P11597 | Cholesteryl e CETP | Homo sapien | 54.757 | 12 | 12 | 8.1E+06 | 7.6E+06 | 8.6E+06 | 8.1E+06 | 4.2E+06 | 5.8E+06 | 9.5E+06 | 3.7E+06 | 4.2E+06 | 9.2E+06 | 6.6E+06 | 3.4E+06 | [illegible] |
| 1097 | 1 P13798 | P13798 | Acylamino-ac APEH | Homo sapien | 81.229 | 12 | 12 | 6.3E+07 | 7.6E+07 | 5.2E+07 | 7.3E+07 | 5.8E+07 | 5.9E+07 | 1.5E+08 | 1.5E+08 | 1.3E+08 | 1.6E+08 | 1.8E+08 | 1.8E+08 | [illegible] |
| 1098 | 1 P13807 | P13807 | Glycogen [st GYS1 | Homo sapien | 83.788 | 13 | 12 | 2.1E+06 | 2.4E+06 | 1.6E+06 | 1.7E+06 | 2.4E+06 | 1.4E+06 | 3.2E+06 | 3.5E+06 | 2.3E+06 | 3.6E+06 | 4.0E+06 | 4.7E+06 | [illegible] |
| 1099 | 1 P14923 | P14923 | Junction plak JUP | Homo sapien | 81.745 | 12 | 12 | 3.0E+05 | 3.4E+05 | 7.6E+04 | 6.2E+05 | 6.1E+05 | 2.1E+06 | 0.0E+00 | 0.0E+00 | 0.0E+00 | 2.7E+04 | 5.6E+05 | 8.7E+05 | [illegible] |
| 1100 | 1 P16333 | P16333 | SH2/SH3 ada NCK1 | Homo sapien | 42.865 | 12 | 12 | 4.3E+06 | 4.1E+06 | 4.0E+06 | 3.6E+06 | 3.8E+06 | 4.0E+06 | 2.4E+06 | 3.2E+06 | 2.8E+06 | 2.8E+06 | 3.4E+06 | 2.8E+06 | [illegible] |
| 1101 | 1 P16383 | P16383 | Intron Large r GCFC2 | Homo sapien | 89.387 | 12 | 12 | 8.3E+05 | 2.2E+06 | 4.5E+05 | 4.8E+05 | 6.5E+05 | 2.9E+05 | 1.6E+05 | 3.9E+05 | 6.5E+05 | 3.6E+05 | 4.6E+05 | 1.9E+05 | [illegible] |
| 1102 | 1 P17301 | P17301 | Integrin alpha ITGA2 | Homo sapien | 129.295 | 12 | 12 | 1.1E+06 | 7.1E+05 | 1.0E+06 | 9.1E+05 | 6.3E+05 | 3.6E+05 | 6.7E+05 | 8.2E+05 | 8.3E+05 | 1.7E+06 | 8.5E+05 | 7.5E+05 | [illegible] |
| 1103 | 1 P22234 | P22234 | Bifunctional p PAICS | Homo sapien | 47.077 | 12 | 12 | 4.2E+07 | 4.2E+07 | 3.1E+07 | 4.3E+07 | 3.6E+07 | 3.0E+07 | 7.3E+07 | 8.4E+07 | 6.0E+07 | 8.2E+07 | 6.2E+07 | 9.0E+07 | [illegible] |
| 1104 | 1 P22626 | P22626 | Heterogeneo HNRNPA2B1 | Homo sapien | 37.428 | 13 | 12 | 1.4E+08 | 1.4E+08 | 1.4E+08 | 1.2E+08 | 1.6E+08 | 1.4E+08 | 1.0E+08 | 1.1E+08 | 1.2E+08 | 1.2E+08 | 1.1E+08 | 1.1E+08 | [illegible] |
| [illegible] | | | | | | | | | | | | | | | | | | | | |

| | | | | | | | | | | | | | | | | | | | | | |
|---|---|---|---|---|---|---|---|---|---|---|---|---|---|---|---|---|---|---|---|---|---|
| 1570 | 1 | Q8N0W3 | Q8N0W3 | L-fucose kina | FCSK | Homo sapien | 117.622 | 10 | 10 | 4.2E+05 | 4.0E+05 | 4.1E+05 | 3.8E+05 | 3.5E+05 | 3.6E+05 | 1.0E+06 | 8.2E+05 | 8.0E+05 | 1.0E+06 | 1.4E+06 | 1.2E+06 |
| 1571 | 1 | Q8N684 | Q8N684 | Cleavage an | CPSF7 | Homo sapien | 52.049 | 10 | 10 | 1.4E+06 | 1.3E+06 | 1.4E+06 | 9.8E+05 | 1.4E+06 | 1.2E+06 | 1.1E+06 | 1.2E+06 | 1.1E+06 | 1.4E+06 | 1.1E+06 | 1.0E+06 |
| 1572 | 1 | Q8ND04 | Q8ND04 | Nonsense-m | SMG8 | Homo sapien | 109.684 | 10 | 10 | 5.0E+06 | 7.7E+06 | 4.3E+06 | 1.5E+06 | 8.8E+06 | 5.6E+06 | 4.0E+06 | 9.2E+05 | 1.4E+06 | 1.3E+06 | 1.4E+06 | 3.0E+06 |
| 1573 | 1 | Q8NE86 | Q8NE86 | Calcium unip | MCU | Homo sapien | 39.867 | 10 | 10 | 4.0E+06 | 4.8E+06 | 5.3E+06 | 4.1E+06 | 4.5E+06 | 3.7E+06 | 6.1E+06 | 7.3E+06 | 7.9E+06 | 7.0E+06 | 7.6E+06 | 6.4E+06 |
| 1574 | 1 | Q8NFD5 | Q8NFD5 | AT-rich inter | ARID1B | Homo sapien | 243.945 | 10 | 10 | 1.6E+05 | 1.7E+05 | 2.9E+05 | 1.7E+05 | 2.2E+05 | 7.5E+04 | 3.0E+05 | 5.7E+05 | 5.9E+05 | 7.3E+05 | 4.8E+05 | 4.0E+05 |
| 1575 | 1 | Q8NFQ8 | Q8NFQ8 | Torsin-1A-int | TOR1AIP2 | Homo sapien | 51.264 | 10 | 10 | 8.2E+04 | 1.1E+05 | 1.9E+05 | 1.5E+05 | 9.6E+04 | 1.4E+05 | 2.6E+05 | 3.6E+05 | 4.9E+05 | 3.1E+05 | 2.5E+05 | 2.1E+05 |
| 1576 | 1 | Q8NHP6 | Q8NHP6 | Motile sperm | MOSPD2 | Homo sapien | 59.746 | 10 | 10 | 2.5E+05 | 2.3E+05 | 3.2E+05 | 3.8E+05 | 3.2E+05 | 2.5E+05 | 4.5E+05 | 4.0E+05 | 3.6E+05 | 5.1E+05 | 4.8E+05 | 3.1E+05 |
| 1577 | 1 | Q8NI99 | Q8NI99 | Angiopoietin- | ANGPTL6 | Homo sapien | 51.694 | 10 | 10 | 3.3E+06 | 4.4E+06 | 2.2E+06 | 5.3E+06 | 3.7E+06 | 2.6E+06 | 5.4E+06 | 3.5E+06 | 4.4E+06 | 5.7E+06 | 4.3E+06 | 5.4E+06 |
| 1578 | 1 | Q8TBX8 | Q8TBX8 | Phosphatidyl | PIP4K2C | Homo sapien | 47.3 | 10 | 10 | 6.3E+05 | 6.9E+05 | 7.6E+05 | 8.9E+05 | 6.9E+05 | 7.3E+05 | 6.8E+05 | 3.0E+05 | 2.5E+05 | 8.3E+05 | 4.5E+05 | 4.6E+05 |
| 1579 | 1 | Q8TC05 | Q8TC05 | Nuclear prote | MDM1 | Homo sapien | 80.735 | 10 | 10 | 5.5E+06 | 1.1E+07 | 3.0E+06 | 8.4E+06 | 4.0E+06 | 2.7E+06 | 1.0E+07 | 9.9E+06 | 1.2E+07 | 1.7E+07 | 1.0E+07 | 1.7E+07 |
| 1580 | 1 | Q8TCS8 | Q8TCS8 | Polyribonucle | PNPT1 | Homo sapien | 85.956 | 10 | 10 | 2.1E+04 | 3.0E+04 | 4.5E+04 | 8.9E+04 | 6.9E+03 | 3.1E+04 | 1.6E+05 | 3.8E+04 | 7.9E+04 | 5.4E+04 | 1.8E+05 | 1.3E+05 |
| 1581 | 1 | Q8TF42 | Q8TF42 | Ubiquitin-ass | UBASH3B | Homo sapien | 72.698 | 10 | 10 | 3.2E+06 | 3.2E+06 | 4.9E+06 | 3.7E+06 | 4.3E+06 | 4.3E+06 | 4.5E+06 | 5.1E+06 | 3.5E+06 | 3.4E+06 | 4.2E+06 | 5.8E+06 |
| 1582 | 1 | Q8WUF8 | Q8WUF8 | Cotranscripti | ARB2A | Homo sapien | 47.973 | 10 | 10 | 9.2E+05 | 7.8E+05 | 1.1E+06 | 1.2E+06 | 1.1E+06 | 9.8E+05 | 4.8E+05 | 7.0E+05 | 5.9E+05 | 5.9E+05 | 7.5E+05 | 4.8E+05 |
| 1583 | 1 | Q8WUH2 | Q8WUH2 | Transforming | TGFBRAP1 | Homo sapien | 97.158 | 10 | 10 | 4.0E+04 | 5.8E+04 | 5.5E+04 | 6.7E+05 | 5.1E+04 | 5.1E+04 | 6.1E+04 | 4.3E+04 | 5.1E+04 | 9.2E+04 | 4.6E+04 | 3.8E+04 |
| 1584 | 1 | Q8WXE9 | Q8WXE9 | Stonin-2 | STON2 | Homo sapien | 101.166 | 10 | 10 | 1.7E+06 | 1.6E+06 | 1.4E+06 | 1.4E+06 | 1.1E+06 | 2.0E+06 | 1.4E+06 | 9.7E+05 | 1.6E+06 | 1.1E+06 | 1.3E+06 | 1.3E+06 |
| 1585 | 1 | Q92599 | Q92599 | Septin-8 | SEPTIN8 | Homo sapien | 55.756 | 10 | 10 | 6.8E+06 | 7.7E+06 | 5.0E+06 | 5.1E+06 | 5.8E+06 | 6.4E+06 | 9.6E+06 | 3.2E+06 | 6.4E+06 | 6.5E+06 | 5.2E+06 | 7.2E+06 |
| 1586 | 1 | Q92626 | Q92626 | Peroxidasin h | PXDN | Homo sapien | 165.278 | 10 | 10 | 4.6E+04 | 6.2E+04 | 6.5E+04 | 5.3E+04 | 8.6E+04 | 8.0E+04 | 5.4E+04 | 0.0E+00 | 7.0E+04 | 7.2E+04 | 1.2E+04 | 0.0E+00 |
| 1587 | 1 | Q92738 | Q92738 | USP6 N-term | USP6NL | Homo sapien | 94.104 | 10 | 10 | 5.1E+06 | 5.6E+06 | 2.8E+06 | 3.8E+06 | 4.0E+06 | 3.3E+06 | 4.0E+06 | 1.5E+06 | 3.0E+06 | 2.3E+06 | 2.1E+06 | 4.0E+06 |
| 1588 | 1 | Q92805 | Q92805 | Golgin subfar | GOLGA1 | Homo sapien | 88.185 | 10 | 10 | 5.0E+04 | 2.7E+04 | 1.5E+06 | 2.0E+04 | 3.3E+04 | 0.0E+00 | 5.5E+04 | 7.5E+03 | 3.2E+04 | 8.6E+04 | 1.0E+05 | 1.0E+05 |
| 1589 | 1 | Q92973 | Q92973 | Transportin-1 | TNPO1 | Homo sapien | 102.357 | 13 | 10 | 6.1E+06 | 6.2E+06 | 5.5E+06 | 6.6E+06 | 6.0E+06 | 6.9E+06 | 1.5E+07 | 1.2E+07 | 9.3E+06 | 1.8E+07 | 1.3E+07 | 1.3E+07 |
| 1590 | 1 | Q92990 | Q92990 | Glomulin | GLMN | Homo sapien | 68.208 | 10 | 10 | 2.8E+05 | 4.6E+05 | 4.1E+05 | 4.8E+05 | 3.1E+05 | 3.3E+05 | 1.4E+06 | 1.1E+06 | 1.1E+06 | 1.8E+06 | 1.1E+06 | 1.0E+06 |
| 1591 | 1 | Q96AQ6 | Q96AQ6 | Pre-B-cell leu | PBXIP1 | Homo sapien | 80.642 | 10 | 10 | 6.8E+05 | 7.9E+05 | 9.9E+05 | 7.6E+05 | 1.1E+06 | 6.8E+05 | 2.2E+05 | 9.3E+05 | 6.6E+05 | 3.0E+05 | 8.0E+05 | 6.2E+05 |
| 1592 | 1 | Q96BJ3 | Q96BJ3 | Axin interacti | AIDA | Homo sapien | 35.021 | 10 | 10 | 4.1E+07 | 5.3E+07 | 4.1E+07 | 6.3E+07 | 4.4E+07 | 6.0E+07 | 5.4E+07 | 5.3E+07 | 6.0E+07 | 3.7E+07 | 5.3E+07 | 4.4E+07 |
| 1593 | 1 | Q96I24 | Q96I24 | Far upstream | FUBP3 | Homo sapien | 61.639 | 10 | 10 | 6.8E+06 | 5.3E+06 | 6.9E+06 | 6.9E+06 | 6.1E+06 | 6.0E+06 | 5.0E+05 | 9.3E+06 | 7.0E+06 | 8.0E+06 | 9.1E+06 | 5.4E+05 |
| 1594 | 1 | Q96I99 | Q96I99 | Succinate--C | SUCLG2 | Homo sapien | 46.51 | 10 | 10 | 8.4E+05 | 3.1E+05 | 1.4E+06 | 2.9E+05 | 4.1E+05 | 3.3E+05 | 1.4E+06 | 1.2E+06 | 1.6E+06 | 2.9E+06 | 4.9E+05 | 2.3E+05 |
| 1595 | 1 | Q96KN2 | Q96KN2 | Beta-Ala-His | CNDP1 | Homo sapien | 56.691 | 10 | 10 | 3.5E+06 | 3.8E+06 | 4.7E+06 | 3.3E+06 | 3.0E+06 | 5.4E+06 | 6.2E+05 | 1.1E+06 | 6.4E+05 | 1.3E+06 | 1.2E+06 | 9.2E+05 |
| 1596 | 1 | Q96PL5 | Q96PL5 | Erythroid me | ERMAP | Homo sapien | 52.604 | 10 | 10 | 1.4E+07 | 1.0E+07 | 1.1E+07 | 1.3E+07 | 1.7E+07 | 1.9E+07 | 2.2E+07 | 1.9E+07 | 3.0E+07 | 3.8E+07 | 2.6E+07 | 2.7E+07 |
| 1597 | 1 | Q96RS6 | Q96RS6 | NudC domain | NUDCD1 | Homo sapien | 66.755 | 10 | 10 | 5.7E+05 | 5.7E+05 | 4.6E+05 | 5.9E+05 | 5.2E+05 | 6.3E+05 | 1.2E+06 | 1.5E+06 | 3.2E+05 | 5.0E+05 | 2.0E+06 | 6.8E+05 |
| 1598 | 1 | Q96SB4 | Q96SB4 | SRSF protein | SRPK1 | Homo sapien | 74.327 | 10 | 10 | 1.2E+06 | 8.7E+05 | 6.7E+05 | 9.2E+05 | 8.8E+05 | 1.0E+06 | 5.1E+05 | 3.7E+05 | 7.7E+05 | 9.0E+05 | 4.8E+05 | 5.9E+05 |
| 1599 | 1 | Q99447 | Q99447 | Ethanolamine | PCYT2 | Homo sapien | 43.833 | 10 | 10 | 4.8E+04 | 2.6E+05 | 3.4E+05 | 2.9E+05 | 2.9E+05 | 2.6E+05 | 1.0E+06 | 1.1E+06 | 9.2E+05 | 9.6E+05 | 1.2E+06 | 1.8E+06 |
| 1600 | 1 | Q99567 | Q99567 | Nuclear pore | NUP88 | Homo sapien | 83.54 | 10 | 10 | 2.1E+06 | 0.0E+00 | 2.1E+06 | 1.6E+06 | 2.5E+06 | 1.6E+06 | 5.6E+05 | 8.3E+05 | 5.2E+05 | 1.3E+06 | 9.7E+05 | 8.6E+05 |
| 1601 | 1 | Q99575 | Q99575 | Ribonuclease | POP1 | Homo sapien | 114.711 | 10 | 10 | 1.9E+04 | 1.5E+04 | 2.8E+04 | 3.2E+04 | 6.6E+04 | 1.7E+04 | 8.5E+03 | 8.1E+03 | 0.0E+00 | 0.0E+00 | 0.0E+00 | 7.8E+03 |
| 1602 | 1 | Q99598 | Q99598 | Translin-ass | TSNAX | Homo sapien | 33.112 | 10 | 10 | 2.1E+07 | 2.1E+07 | 2.0E+07 | 3.1E+07 | 2.4E+07 | 2.4E+07 | 3.4E+07 | 2.6E+07 | 2.1E+07 | 2.0E+07 | 2.9E+07 | 3.3E+07 |
| 1603 | 1 | Q99816 | Q99816 | Tumor susce | TSG101 | Homo sapien | 43.944 | 10 | 10 | 1.6E+07 | 1.4E+07 | 1.3E+07 | 1.5E+07 | 1.0E+07 | 7.0E+06 | 6.5E+06 | 7.4E+06 | 6.0E+06 | 1.1E+07 | 1.8E+07 | 8.2E+06 |
| 1604 | 1 | Q9BRX2 | Q9BRX2 | Protein pelot | PELO | Homo sapien | 43.358 | 10 | 10 | 2.3E+08 | 1.8E+08 | 1.6E+08 | 1.3E+08 | 1.9E+08 | 2.0E+08 | 3.4E+08 | 1.3E+08 | 1.6E+08 | 1.4E+08 | 1.3E+08 | 3.4E+08 |
| 1605 | 1 | Q9BSL1 | Q9BSL1 | Ubiquitin-ass | UBAC1 | Homo sapien | 45.338 | 10 | 10 | 1.1E+07 | 2.0E+07 | 1.4E+07 | 1.6E+07 | 1.1E+07 | 1.4E+07 | 3.1E+07 | 2.7E+07 | 2.4E+07 | 3.1E+07 | 2.8E+07 | 2.9E+07 |
| 1606 | 1 | Q9BSQ5 | Q9BSQ5 | Cerebral cav | CCM2 | Homo sapien | 48.836 | 10 | 10 | 1.3E+06 | 1.1E+06 | 1.0E+06 | 1.3E+06 | 1.2E+06 | 1.1E+06 | 6.8E+05 | 4.2E+05 | 5.6E+05 | 6.0E+05 | 6.4E+05 | 8.0E+05 |
| 1607 | 1 | Q9BW27 | Q9BW27 | Nuclear pore | NUP85 | Homo sapien | 75.019 | 10 | 10 | 8.6E+05 | 8.9E+05 | 3.4E+05 | 2.9E+05 | 9.8E+05 | 2.9E+05 | 3.1E+05 | 2.7E+05 | 1.7E+05 | 2.9E+05 | 2.0E+05 | 2.6E+05 |
| 1608 | 1 | Q9BWU0 | Q9BWU0 | Kanadaptin | SLC4A1AP | Homo sapien | 82.892 | 10 | 10 | 1.8E+05 | 2.5E+05 | 2.7E+05 | 2.4E+05 | 1.5E+07 | 2.6E+05 | 1.1E+05 | 1.0E+05 | 1.7E+05 | 1.9E+05 | 1.1E+05 | 1.5E+05 |
| 1609 | 1 | Q9BYM8 | Q9BYM8 | RanBP-type | RBCK1 | Homo sapien | 57.573 | 10 | 10 | 1.2E+06 | 1.2E+06 | 1.9E+06 | 1.0E+06 | 1.0E+06 | 1.1E+06 | 1.5E+06 | 1.4E+06 | 8.9E+06 | 1.9E+06 | 1.2E+07 | 1.7E+06 |
| 1610 | 1 | Q9BZ95 | Q9BZ95 | Histone-lysin | NSD3 | Homo sapien | 161.616 | 10 | 10 | 7.8E+03 | 1.0E+05 | 5.4E+03 | 3.4E+05 | 1.2E+05 | 2.9E+05 | 5.4E+04 | 5.6E+04 | 4.3E+04 | 0.0E+00 | 0.0E+00 | 0.0E+00 |
| 1611 | 1 | Q9BZV1 | Q9BZV1 | UBX domain- | UBXN6 | Homo sapien | 49.756 | 10 | 10 | 2.5E+07 | 1.9E+07 | 2.0E+07 | 2.5E+07 | 2.4E+07 | 2.2E+07 | 1.9E+07 | 1.4E+07 | 1.8E+07 | 1.6E+07 | 1.7E+07 | 1.9E+07 |
| 1612 | 1 | Q9H078 | Q9H078 | Mitochondria | CLPB | Homo sapien | 78.728 | 10 | 10 | 1.2E+07 | 2.0E+06 | 2.4E+07 | 1.7E+06 | 1.5E+07 | 1.5E+07 | 9.7E+06 | 1.2E+07 | 1.2E+07 | 1.3E+07 | 1.7E+07 | 9.0E+06 |
| 1613 | 1 | Q9H1H9 | Q9H1H9 | Kinesin-like p | KIF13A | Homo sapien | 202.313 | 10 | 10 | 3.3E+05 | 6.9E+04 | 5.2E+04 | 3.5E+04 | 4.9E+04 | 6.2E+04 | 1.5E+05 | 7.9E+04 | 5.8E+04 | 5.9E+04 | 2.1E+05 | 1.1E+05 |
| 1614 | 1 | Q9H2U2 | Q9H2U2 | Inorganic pyr | PPA2 | Homo sapien | 37.92 | 10 | 10 | 9.7E+05 | 3.3E+07 | 8.9E+05 | 4.0E+07 | 9.6E+05 | 1.8E+06 | 1.5E+06 | 2.3E+06 | 2.5E+06 | 1.5E+06 | 4.2E+06 | 2.3E+06 |
| 1615 | 1 | Q9H425 | Q9H425 | Uncharacteri | C1orf198 | Homo sapien | 36.346 | 10 | 10 | 2.2E+06 | 2.7E+06 | 2.2E+06 | 2.3E+06 | 2.2E+06 | 2.4E+06 | 2.7E+06 | 2.9E+06 | 3.1E+06 | 2.8E+06 | 2.6E+06 | 2.9E+06 |
| 1616 | 1 | Q9H4B7 | Q9H4B7 | Tubulin beta- | TUBB1 | Homo sapien | 50.327 | 14 | 10 | 5.2E+07 | 6.5E+07 | 5.6E+07 | 5.4E+07 | 3.2E+07 | 4.3E+07 | 7.8E+07 | 9.3E+07 | 4.4E+07 | 1.1E+08 | 1.0E+08 | 7.4E+07 |
| 1617 | 1 | Q9H6A0 | Q9H6A0 | DENN domai | DENND2D | Homo sapien | 53.672 | 11 | 10 | 3.8E+05 | 8.5E+04 | 2.7E+05 | 2.4E+05 | 3.1E+05 | 1.9E+05 | 7.9E+04 | 1.2E+05 | 5.8E+04 | 2.3E+05 | 1.2E+05 | 1.2E+05 |
| 1618 | 1 | Q9H6Z4 | Q9H6Z4 | Ran-binding | RANBP3 | Homo sapien | 60.21 | 10 | 10 | 1.6E+06 | 1.0E+06 | 1.6E+06 | 1.5E+06 | 1.8E+06 | 1.8E+06 | 2.1E+06 | 1.8E+06 | 1.4E+06 | 1.9E+06 | 1.4E+06 | 1.8E+06 |
| 1619 | 1 | Q9H814 | Q9H814 | Phosphorylat | PHAX | Homo sapien | 44.4 | 10 | 10 | 9.2E+06 | 9.2E+06 | 7.6E+06 | 9.4E+06 | 7.4E+06 | 6.9E+06 | 8.9E+06 | 1.0E+07 | 6.7E+06 | 1.0E+07 | 1.0E+07 | 1.0E+07 |
| 1620 | 1 | Q9HB71 | Q9HB71 | Calcyclin-bin | CACYBP | Homo sapien | 26.21 | 10 | 10 | 1.9E+07 | 1.6E+07 | 1.7E+07 | 1.5E+07 | 1.7E+07 | 1.2E+07 | 1.4E+07 | 1.2E+07 | 1.2E+07 | 1.3E+07 | 1.5E+07 | 1.3E+07 |
| 1621 | 1 | Q9HCC0 | Q9HCC0 | Methylcroton | MCCC2 | Homo sapien | 61.332 | 10 | 10 | 7.0E+04 | 3.4E+04 | 7.2E+04 | 6.2E+04 | 5.9E+04 | 1.1E+04 | 2.8E+05 | 1.3E+05 | 3.6E+05 | 2.5E+05 | 2.1E+05 | 4.1E+05 |
| 1622 | 1 | Q9HCD5 | Q9HCD5 | Nuclear rece | NCOA5 | Homo sapien | 65.537 | 10 | 10 | 1.0E+06 | 1.1E+06 | 1.2E+06 | 5.2E+05 | 1.1E+06 | 7.7E+05 | 8.7E+05 | 8.3E+05 | 6.8E+05 | 1.0E+06 | 6.0E+05 | 5.5E+05 |
| 1623 | 1 | Q9NR46 | Q9NR46 | Endophilin-B | SH3GLB2 | Homo sapien | 43.977 | 10 | 10 | 1.3E+07 | 7.4E+06 | 9.2E+06 | 1.5E+07 | 1.7E+07 | 1.3E+07 | 2.4E+07 | 2.1E+07 | 1.9E+07 | 2.1E+07 | 2.9E+07 | 2.6E+07 |
| 1624 | 1 | Q9NRA8 | Q9NRA8 | Eukaryotic tr | EIF4ENIF1 | Homo sapien | 108.203 | 10 | 10 | 1.5E+06 | 1.9E+06 | 1.5E+06 | 8.8E+05 | 1.6E+06 | 1.0E+06 | 1.2E+05 | 1.3E+05 | 1.6E+05 | 8.7E+04 | 1.8E+05 | 5.2E+04 |
| 1625 | 1 | Q9NRH2 | Q9NRH2 | SNF-related | SNRK | Homo sapien | 84.276 | 10 | 10 | 2.8E+05 | 1.2E+05 | 1.9E+05 | 2.4E+05 | 1.7E+06 | 8.2E+05 | 5.7E+04 | 1.9E+05 | 2.0E+05 | 1.5E+05 | 1.3E+05 | 2.5E+05 |
| 1626 | 1 | Q9NSD9 | Q9NSD9 | Phenylalanine | FARSB | Homo sapien | 66.117 | 10 | 10 | 8.7E+06 | 9.1E+06 | 6.4E+06 | 7.8E+06 | 9.4E+06 | 1.1E+07 | 8.1E+06 | 5.3E+06 | 5.0E+06 | 1.0E+07 | 1.1E+07 | 1.0E+07 |
| 1627 | 1 | Q9NVG8 | Q9NVG8 | TBC1 domain | TBC1D13 | Homo sapien | 46.555 | 10 | 10 | 3.9E+06 | 3.8E+05 | 4.0E+05 | 7.0E+05 | 3.1E+06 | 3.0E+06 | 8.7E+06 | 1.0E+07 | 6.7E+06 | 1.2E+07 | 9.7E+06 | 6.7E+06 |
| 1628 | 1 | Q9NVM6 | Q9NVM6 | DnaJ homolo | DNAJC17 | Homo sapien | 34.686 | 10 | 10 | 6.4E+06 | 5.1E+06 | 6.7E+06 | 7.4E+06 | 6.5E+06 | 6.7E+06 | 5.7E+06 | 4.5E+06 | 2.4E+06 | 3.2E+06 | 2.9E+06 | 2.7E+06 |
| 1629 | 1 | Q9NW13 | Q9NW13 | RNA-binding | RBM28 | Homo sapien | 85.738 | 10 | 10 | 1.0E+06 | 7.1E+03 | 8.7E+05 | 7.9E+05 | 1.0E+06 | 3.0E+04 | 0.0E+00 | 2.7E+05 | 0.0E+00 | 2.6E+05 | 2.8E+05 | 0.0E+00 |
| 1630 | 1 | Q9NXR1 | Q9NXR1 | Nuclear distri | NDE1 | Homo sapien | 37.72 | 13 | 10 | 3.6E+06 | 4.5E+06 | 4.0E+06 | 4.1E+06 | 4.7E+06 | 4.6E+06 | 4.0E+06 | 3.2E+06 | 2.3E+06 | 5.2E+06 | 2.5E+06 | 3.0E+06 |
| 1631 | 1 | Q9UGP8 | Q9UGP8 | Translocatio | SEC63 | Homo sapien | 87.998 | 10 | 10 | 3.9E+06 | 4.2E+06 | 3.0E+06 | 2.7E+06 | 4.4E+06 | 3.1E+06 | 4.4E+06 | 4.9E+06 | 4.9E+06 | 4.5E+06 | 6.2E+06 | 4.2E+06 |
| 1632 | 1 | Q9UHD1 | Q9UHD1 | Cysteine and | CHORDC1 | Homo sapien | 37.489 | 10 | 10 | 3.1E+07 | 3.5E+07 | 2.7E+07 | 2.9E+07 | 3.3E+07 | 4.1E+07 | 4.8E+07 | 5.5E+07 | 3.3E+07 | 3.6E+07 | 4.0E+07 | 4.0E+07 |
| 1633 | 1 | Q9UID3 | Q9UID3 | Vacuolar pro | VPS51 | Homo sapien | 86.042 | 10 | 10 | 1.3E+05 | 7.3E+04 | 1.4E+05 | 1.3E+05 | 1.0E+05 | 1.6E+04 | 1.4E+05 | 1.3E+05 | 1.2E+05 | 5.2E+04 | 1.8E+05 | 0.0E+00 |
| 1634 | 1 | Q9UIQ6 | Q9UIQ6 | Leucyl-cystin | LNPEP | Homo sapien | 117.35 | 10 | 10 | 6.3E+05 | 6.2E+04 | 5.8E+05 | 5.2E+05 | 8.8E+05 | 7.3E+05 | 1.7E+05 | 1.8E+05 | 3.2E+05 | 7.9E+05 | 8.5E+05 | 1.0E+06 |
| 1635 | 1 | Q9UIW2 | Q9UIW2 | Plexin-A1 | PLXNA1 | Homo sapien | 211.069 | 12 | 10 | 1.6E+04 | 3.6E+04 | 2.7E+04 | 1.9E+05 | 4.6E+04 | 4.8E+04 | 0.0E+00 | 0.0E+00 | 0.0E+00 | 1.0E+05 | 6.1E+04 | 0.0E+00 |
| 1636 | 1 | Q9UJ70 | Q9UJ70 | N-acetyl-D-g | NAGK | Homo sapien | 37.374 | 10 | 10 | 2.6E+06 | 2.8E+06 | 7.1E+06 | 2.7E+06 | 6.9E+06 | 7.1E+06 | 1.0E+07 | 1.1E+07 | 1.2E+07 | 1.4E+07 | 1.3E+07 | 1.0E+07 |
| 1637 | 1 | Q9UK55 | Q9UK55 | Protein Z-de | SERPINA10 | Homo sapien | 50.707 | 10 | 10 | 1.1E+07 | 1.1E+07 | 8.6E+06 | 9.0E+06 | 1.0E+07 | 1.0E+07 | 6.5E+06 | 6.1E+06 | 7.0E+06 | 8.1E+06 | 7.4E+06 | 6.7E+06 |
| 1638 | 1 | Q9UK61 | Q9UK61 | Protein TASC | TASOR | Homo sapien | 189.034 | 10 | 10 | 1.4E+05 | 3.3E+05 | 1.7E+05 | 2.2E+05 | 2.9E+05 | 2.3E+05 | 0.0E+00 | 8.8E+03 | 0.0E+00 | 1.0E+04 | 0.0E+00 | 0.0E+00 |
| 1639 | 1 | Q9UKG1 | Q9UKG1 | DCC-interact | APPL1 | Homo sapien | 79.664 | 10 | 10 | 7.3E+05 | 6.3E+05 | 7.9E+05 | 6.1E+05 | 7.6E+05 | 7.2E+05 | 1.1E+06 | 9.2E+05 | 1.0E+06 | 1.2E+06 | 1.2E+06 | 1.1E+06 |
| 1640 | 1 | Q9UKK3 | Q9UKK3 | Protein mono | PARP4 | Homo sapien | 192.596 | 10 | 10 | 2.2E+06 | 3.6E+06 | 2.4E+06 | 1.9E+06 | 1.7E+06 | 1.5E+06 | 2.0E+05 | 1.1E+05 | 2.5E+06 | 4.1E+06 | 4.9E+06 | 8.6E+03 |
| 1641 | 1 | Q9UL46 | Q9UL46 | Proteasome | PSME2 | Homo sapien | 27.399 | 10 | 10 | 2.1E+07 | 2.5E+07 | 1.7E+07 | 2.3E+07 | 2.9E+07 | 2.5E+07 | 2.2E+07 | 3.3E+07 | 2.7E+07 | 1.8E+07 | 3.0E+07 | 2.9E+07 |
| 1642 | 1 | Q9UM54 | Q9UM54 | Unconvention | MYO6 | Homo sapien | 149.694 | 10 | 10 | 1.6E+05 | 0.0E+00 | 1.1E+06 | 1.0E+06 | 1.4E+06 | 1.2E+06 | 1.3E+05 | 7.5E+04 | 1.1E+05 | 5.8E+04 | 1.0E+05 | 3.7E+03 |
| 1643 | 1 | Q9UNF0 | Q9UNF0 | Protein kinas | PACSIN2 | Homo sapien | 55.736 | 10 | 10 | 2.6E+07 | 3.1E+07 | 2.8E+07 | 2.8E+07 | 3.0E+07 | 2.6E+07 | 2.8E+07 | 1.8E+07 | 2.5E+07 | 2.3E+07 | 2.8E+07 | 2.4E+07 |
| 1644 | 1 | Q9UNY4 | Q9UNY4 | Transcription | TTF2 | Homo sapien | 129.59 | 10 | 10 | 1.7E+03 | 1.6E+04 | 1.2E+04 | 3.2E+04 | 3.9E+03 | 3.7E+04 | 9.5E+03 | 9.3E+03 | 1.4E+04 | 8.6E+03 | 0.0E+00 | 7.0E+03 |
| 1645 | 1 | Q9UPW5 | Q9UPW5 | Cytosolic car | AGTPBP1 | Homo sapien | 138.45 | 10 | 10 | 1.1E+05 | 9.2E+04 | 3.4E+05 | 1.4E+05 | 1.3E+05 | 7.7E+04 | 0.0E+00 | 4.4E+04 | 6.8E+04 | 3.2E+04 | 2.2E+04 | 0.0E+00 |
| 1646 | 1 | Q9UQ16 | Q9UQ16 | Dynamin-3 | DNM3 | Homo sapien | 97.746 | 10 | 10 | 6.3E+05 | 9.5E+05 | 3.5E+05 | 6.5E+05 | 3.9E+05 | 7.9E+05 | 3.1E+05 | 7.1E+05 | 4.8E+05 | 3.8E+05 | 6.7E+05 | 4.5E+05 |
| 1647 | 1 | Q9Y217 | Q9Y217 | Phosphatidyl | MTMR6 | Homo sapien | 71.968 | 10 | 10 | 5.9E+04 | 7.8E+03 | 6.9E+04 | 2.3E+05 | 1.0E+05 | 3.0E+04 | 4.1E+04 | 0.0E+00 | 7.7E+03 | 0.0E+00 | 7.9E+03 | 0.0E+00 |
| 1648 | 1 | Q9Y285 | Q9Y285 | Phenylalanine | FARSA | Homo sapien | 57.561 | 10 | 10 | 5.5E+06 | 4.8E+06 | 3.6E+06 | 4.0E+06 | 5.5E+06 | 6.5E+06 | 4.6E+06 | 4.3E+06 | 5.4E+06 | 6.8E+06 | 7.7E+06 | 6.1E+06 |
| 1649 | 1 | Q9Y2G2 | Q9Y2G2 | Caspase rec | CARD8 | Homo sapien | 60.652 | 10 | 10 | 2.4E+05 | 1.1E+05 | 7.6E+03 | 1.1E+04 | 3.7E+05 | 1.9E+05 | 7.0E+05 | 1.2E+05 | 1.6E+05 | 1.0E+05 | 5.0E+05 | 3.8E+05 |
| 1650 | 1 | Q9Y2G8 | Q9Y2G8 | DnaJ homolo | DNAJC16 | Homo sapien | 90.592 | 10 | 10 | 4.0E+05 | 3.0E+06 | 2.2E+05 | 5.0E+05 | 2.2E+05 | 1.4E+06 | 2.2E+05 | 2.4E+05 | 2.2E+05 | 1.6E+05 | 1.5E+05 | 2.3E+05 |
| 1651 | 1 | Q9Y2I8 | Q9Y2I8 | WD repeat-c | WDR37 | Homo sapien | 54.665 | 10 | 10 | 1.6E+07 | 1.7E+07 | 1.5E+07 | 6.1E+06 | 1.6E+07 | 1.6E+07 | 6.3E+06 | 1.6E+07 | 6.2E+06 | 6.2E+06 | 7.0E+06 | 6.4E+06 |
| 1652 | 1 | Q9Y305 | Q9Y305 | Acyl-coenzy | ACOT9 | Homo sapien | 49.902 | 10 | 10 | 8.1E+06 | 6.8E+06 | 6.5E+06 | 6.5E+06 | 6.8E+06 | 4.4E+06 | 1.8E+07 | 2.0E+07 | 8.9E+06 | 6.9E+06 | 8.7E+06 | 6.2E+06 |
| 1653 | 1 | Q9Y487 | Q9Y487 | V-type proto | ATP6V0A2 | Homo sapien | 98.085 | 10 | 10 | 2.7E+05 | 2.0E+05 | 2.0E+05 | 1.9E+05 | 3.4E+05 | 2.5E+05 | 3.8E+05 | 3.6E+05 | 1.4E+05 | 3.4E+05 | 2.5E+05 | 3.9E+05 |
| 1654 | 1 | Q9Y4P3 | Q9Y4P3 | Transducin b | TBL2 | Homo sapien | 49.8 | 10 | 10 | 1.0E+06 | 1.0E+06 | 7.8E+05 | 6.0E+05 | 7.9E+05 | 8.9E+05 | 1.2E+06 | 1.0E+06 | 8.5E+04 | 1.4E+06 | 1.0E+06 | 1.0E+05 |
| 1655 | 1 | Q9Y5X3 | Q9Y5X3 | Sorting nexin | SNX5 | Homo sapien | 46.817 | 11 | 10 | 2.0E+07 | 2.1E+07 | 1.6E+07 | 2.1E+07 | 1.7E+07 | 1.3E+07 | 1.8E+07 | 1.2E+07 | 1.5E+07 | 2.1E+07 | 2.4E+07 | 2.0E+07 |
| 1656 | 1 | Q9Y617 | Q9Y617 | Phosphoseri | PSAT1 | Homo sapien | 40.423 | 10 | 10 | 2.5E+05 | 2.5E+07 | 3.0E+07 | 2.7E+07 | 2.3E+07 | 4.0E+05 | 2.8E+04 | 1.0E+04 | 5.6E+04 | 3.3E+04 | 4.8E+04 | 1.9E+04 |
| 1657 | 1 | Q9Y6M1 | Q9Y6M1 | Insulin-like g | IGF2BP2 | Homo sapien | 66.124 | 10 | 10 | 2.6E+06 | 1.8E+06 | 1.8E+06 | 1.9E+06 | 2.4E+06 | 2.1E+06 | 1.9E+06 | 1.9E+06 | 1.9E+06 | 2.4E+06 | 2.1E+06 | 2.2E+06 |
| 1658 | 1 | Q9Y6U3 | Q9Y6U3 | Scinderin | SCIN | Homo sapien | 80.49 | 10 | 10 | 0.0E+00 | 0.0E+00 | 1.6E+06 | 0.0E+00 | 0.0E+00 | 0.0E+00 | 0.0E+00 | 0.0E+00 | 0.0E+00 | 0.0E+00 | 0.0E+00 | 0.0E+00 |
| 1659 | 1 | A1L390 | A1L390 | Pleckstrin ho | PLEKHG3 | Homo sapien | 134.415 | 9 | 9 | 6.3E+05 | 4.9E+04 | 4.2E+04 | 5.1E+04 | 5.7E+04 | 6.9E+04 | 2.8E+04 | 1.8E+05 | 3.9E+04 | 1.2E+04 | 2.1E+04 | 1.4E+05 |
| 1660 | 1 | A1X283 | A1X283 | SH3 and PX | SH3PXD2B | Homo sapien | 101.58 | 9 | 9 | 0.0E+00 | 0.0E+00 | 1.6E+06 | 5.0E+05 | 9.3E+05 | 8.8E+05 | 1.7E+06 | 0.0E+00 | 7.1E+05 | 5.0E+05 | 1.2E+06 | 0.0E+00 |
| 1661 | 1 | A4D126 | A4D126 | D-ribitol-5-ph | CRPPA | Homo sapien | 49.874 | 9 | 9 | 3.3E+05 | 4.6E+06 | 5.0E+06 | 4.6E+06 | 4.1E+05 | 3.2E+04 | 1.1E+07 | 8.9E+06 | 7.7E+06 | 1.1E+07 | 9.5E+05 | 8.2E+06 |
| 1662 | 1 | O00154 | O00154 | Cytosolic acy | ACOT7 | Homo sapien | 41.792 | 9 | 9 | 4.4E+06 | 6.9E+06 | 4.4E+06 | 4.7E+06 | 7.1E+06 | 5.2E+06 | 1.3E+07 | 8.8E+06 | 4.1E+06 | 9.7E+06 | 1.0E+07 | 1.3E+07 |
| 1663 | 1 | O00189 | O00189 | AP-4 comple | AP4M1 | Homo sapien | 49.978 | 9 | 9 | 4.1E+06 | 7.8E+05 | 4.4E+06 | 4.8E+06 | 4.5E+06 | 4.0E+06 | 5.1E+06 | 4.2E+06 | 3.6E+06 | 4.8E+06 | 6.6E+06 | 6.0E+06 |
| 1664 | 1 | O00194 | O00194 | Ras-related p | RAB27B | Homo sapien | 24.608 | 10 | 9 | 7.8E+06 | 9.4E+06 | 7.9E+06 | 5.8E+06 | 7.7E+06 | 7.2E+06 | 1.0E+07 | 8.0E+06 | 9.0E+06 | 1.3E+07 | 1.2E+07 | 9.9E+06 |
| 1665 | 1 | O00214 | O00214 | Galectin-8 | LGALS8 | Homo sapien | 35.804 | 9 | 9 | 5.8E+05 | 6.6E+05 | 3.8E+05 | 5.2E+05 | 4.1E+05 | 9.0E+05 | 4.1E+05 | 3.2E+05 | 4.2E+05 | 3.9E+05 | 5.5E+05 | 3.1E+05 |
| 1666 | 1 | O00562 | O00562 | Membrane-a | PITPNM1 | Homo sapien | 134.848 | 9 | 9 | 9.5E+04 | 1.1E+05 | 1.8E+05 | 8.9E+04 | 4.0E+04 | 2.3E+05 | 1.8E+05 | 1.1E+05 | 1.5E+05 | 8.8E+04 | 9.0E+04 | 1.2E+05 |
| 1667 | 1 | O14745 | O14745 | Na(+)/H(+) ex | NHERF1 | Homo sapien | 38.867 | 9 | 9 | 9.5E+06 | 7.7E+06 | 8.5E+06 | 8.2E+06 | 7.2E+06 | 8.7E+06 | 6.0E+06 | 6.1E+06 | 5.0E+06 | 5.5E+06 | 6.3E+06 | 6.0E+06 |
| 1668 | 1 | O14791 | O14791 | Apolipoprote | APOL1 | Homo sapien | 43.974 | 9 | 9 | 8.8E+07 | 4.5E+07 | 6.6E+07 | 8.2E+07 | 5.4E+07 | 1.2E+08 | 6.7E+07 | 8.0E+07 | 6.1E+07 | 7.0E+07 | 5.3E+07 | 9.2E+07 |
| 1669 | 1 | O15037 | O15037 | Protein KHN | KHNYN | Homo sapien | 74.533 | 9 | 9 | 1.2E+06 | 7.0E+05 | 9.4E+05 | 9.1E+05 | 6.9E+05 | 7.9E+05 | 8.9E+05 | 6.6E+05 | 8.5E+05 | 9.8E+05 | 1.0E+06 | 1.1E+06 |
| 1670 | 1 | O15068 | O15068 | Guanine nucl | MCF2L | Homo sapien | 128.111 | 9 | 9 | 4.5E+04 | 0.0E+00 | 1.1E+04 | 2.6E+04 | 9.1E+03 | 1.3E+04 | 1.3E+04 | 0.0E+00 | 0.0E+00 | 0.0E+00 | 0.0E+00 | 1.0E+04 |
| 1671 | 1 | O15111 | O15111 | Inhibitor of n | CHUK | Homo sapien | 84.641 | 9 | 9 | 2.5E+05 | 3.4E+05 | 5.8E+05 | 4.4E+05 | 5.8E+05 | 6.4E+05 | 5.7E+05 | 2.3E+05 | 2.9E+05 | 7.4E+05 | 2.1E+05 | 4.0E+05 |
| 1672 | 1 | O43172 | O43172 | U4/U6 small | PRPF4 | Homo sapien | 58.45 | 9 | 9 | 5.5E+05 | 1.8E+05 | 5.2E+05 | 1.2E+05 | 1.9E+06 | 4.5E+05 | 1.4E+06 | 1.5E+06 | 1.1E+06 | 1.5E+06 | 1.4E+06 | 8.5E+05 |
| 1673 | 1 | O43306 | O43306 | Adenylate cy | ADCY6 | Homo sapien | 130.619 | 10 | 9 | 8.8E+05 | 9.1E+05 | 8.0E+05 | 8.4E+05 | 1.0E+06 | 8.9E+05 | 6.0E+05 | 7.2E+05 | 6.2E+05 | 1.1E+06 | 1.0E+06 | 7.1E+05 |
| 1674 | 1 | O43399 | O43399 | Tumor protei | TPD52L2 | Homo sapien | 22.237 | 9 | 9 | 1.1E+06 | 1.1E+06 | 1.1E+06 | 1.1E+06 | 1.2E+06 | 1.1E+06 | 1.7E+06 | 1.4E+06 | 1.1E+06 | 1.3E+06 | 1.9E+06 | 1.6E+06 |
| 1675 | 1 | O43516 | O43516 | WAS/WASL-i | WIPF1 | Homo sapien | 51.258 | 9 | 9 | 1.3E+07 | 1.8E+07 | 1.4E+07 | 1.3E+07 | 1.2E+07 | 1.5E+07 | 1.3E+07 | 1.4E+07 | 1.4E+07 | 1.5E+07 | 1.8E+07 | 1.4E+07 |
| 1676 | 1 | O43639 | O43639 | Cytoplasmic | NCK2 | Homo sapien | 42.916 | 9 | 9 | 9.0E+06 | 9.2E+06 | 9.4E+06 | 8.6E+06 | 6.4E+06 | 7.9E+06 | 5.6E+06 | 6.8E+06 | 6.8E+06 | 5.4E+06 | 5.7E+06 | 5.6E+06 |
| 1677 | 1 | O43719 | O43719 | 17S U2 SnR | HTATSF1 | Homo sapien | 85.854 | 9 | 9 | 3.6E+06 | 2.9E+06 | 4.3E+06 | 3.7E+06 | 4.0E+06 | 5.1E+06 | 3.1E+06 | 3.4E+06 | 2.7E+06 | 2.6E+06 | 3.0E+06 | 2.8E+06 |
| 1678 | 1 | O60256 | O60256 | Phosphoribos | PRPSAP2 | Homo sapien | 40.924 | 9 | 9 | 1.5E+07 | 1.7E+07 | 1.3E+07 | 1.6E+07 | 1.9E+07 | 1.5E+07 | 2.1E+07 | 2.4E+07 | 2.3E+07 | 1.6E+07 | 3.4E+07 | 3.0E+07 |
| 1679 | 1 | O60551 | O60551 | Glycylpeptide | NMT2 | Homo sapien | 56.981 | 9 | 9 | 3.5E+05 | 2.5E+05 | 3.0E+05 | 6.2E+04 | 6.9E+05 | 2.8E+04 | 6.9E+03 | 0.0E+00 | 1.8E+05 | 3.3E+04 | 0.0E+00 | 3.2E+04 |
| 1680 | 1 | O60825 | O60825 | 6-phosphofru | PFKFB2 | Homo sapien | 58.477 | 12 | 9 | 1.7E+06 | 4.3E+05 | 4.2E+05 | 3.4E+05 | 1.6E+06 | 1.9E+06 | 7.4E+05 | 5.8E+05 | 1.7E+06 | 5.0E+05 | 5.5E+05 | 6.3E+05 |
| 1681 | 1 | O60884 | O60884 | DnaJ homolo | DNAJA2 | Homo sapien | 45.746 | 9 | 9 | 2.5E+07 | 2.2E+07 | 2.4E+07 | 3.4E+07 | 2.7E+07 | 2.4E+07 | 2.0E+07 | 1.6E+07 | 1.5E+07 | 1.9E+07 | 2.2E+07 | 2.1E+07 |
| 1682 | 1 | O75027 | O75027 | Iron-sulfur cl | ABCB7 | Homo sapien | 82.644 | 9 | 9 | 2.4E+06 | 2.3E+06 | 2.1E+06 | 2.2E+06 | 2.2E+06 | 1.9E+06 | 3.4E+06 | 3.8E+06 | 3.8E+06 | 3.5E+06 | 2.9E+06 | 3.1E+06 |
| 1683 | 1 | O75044 | O75044 | SLIT-ROBO R | SRGAP2 | Homo sapien | 120.871 | 19 | 9 | 1.4E+06 | 2.1E+06 | 1.8E+06 | 1.8E+06 | 1.1E+06 | 1.3E+06 | 1.3E+06 | 8.4E+05 | 8.8E+05 | 1.2E+06 | 6.8E+05 | 9.0E+05 |
| 1684 | 1 | O75175 | O75175 | CCR4-NOT t | CNOT3 | Homo sapien | 81.874 | 9 | 9 | 1.1E+06 | 8.7E+05 | 9.8E+05 | 9.8E+05 | 1.4E+06 | 1.2E+06 | 1.2E+06 | 6.1E+05 | 6.1E+05 | 1.1E+06 | 1.7E+06 | 1.3E+06 |
| 1685 | 1 | O75339 | O75339 | Cartilage inte | CILP | Homo sapien | 132.566 | 9 | 9 | 3.4E+05 | 1.5E+05 | 3.3E+05 | 1.7E+05 | 5.6E+05 | 4.5E+05 | 2.9E+05 | 2.4E+05 | 1.9E+05 | 7.0E+04 | 1.7E+05 | 4.7E+05 |
| 1686 | 1 | O75475 | O75475 | PC4 and SFR | PSIP1 | Homo sapien | 60.104 | 9 | 9 | 5.0E+06 | 4.1E+06 | 4.4E+06 | 5.0E+06 | 5.9E+06 | 4.2E+06 | 5.8E+06 | 4.4E+06 | 4.3E+06 | 7.7E+06 | 5.3E+06 | 3.6E+06 |

| | | | | | | | | | | | | | | | | | | | | |
|---|---|---|---|---|---|---|---|---|---|---|---|---|---|---|---|---|---|---|---|---|
| 2094 | 1 | Q5VVQ6 | Q5VVQ6 | Ubiquitin thioYOD1 | Homo sapien | 38.321 | 8 | 8 | 2.1E+07 | 3.1E+07 | 3.0E+07 | 4.3E+07 | 2.9E+07 | 2.8E+07 | 5.7E+07 | 7.7E+07 | 5.1E+07 | 8.1E+07 | 6.6E+07 | 8.0E+07 |
| 2095 | 1 | Q676U5 | Q676U5 | Autophagy-reATG16L1 | Homo sapien | 68.265 | 8 | 8 | 3.7E+05 | 2.4E+05 | 3.4E+05 | 4.6E+05 | 2.4E+05 | 1.0E+05 | 4.1E+05 | 2.9E+05 | 3.2E+05 | 3.5E+05 | 5.0E+05 | 2.3E+05 |
| 2096 | 1 | Q6BD51 | Q6BD51 | DENN domaiDENND2C | Homo sapien | 106.867 | 8 | 8 | 1.2E+06 | 8.3E+05 | 6.9E+05 | 7.0E+05 | 7.1E+05 | 9.3E+05 | 6.7E+05 | 3.4E+05 | 3.7E+05 | 4.8E+05 | 2.2E+05 | 3.7E+05 |
| 2097 | 1 | Q6IBS0 | Q6IBS0 | Twinfilin-2 TWF2 | Homo sapien | 39.548 | 9 | 8 | 9.7E+06 | 9.2E+06 | 9.7E+06 | 8.6E+06 | 8.5E+06 | 7.0E+06 | 7.6E+06 | 8.6E+06 | 7.2E+06 | 8.4E+06 | 1.0E+07 | 1.0E+07 |
| 2098 | 1 | Q6NUQ1 | Q6NUQ1 | RAD50-interaRINT1 | Homo sapien | 90.636 | 8 | 8 | 2.3E+05 | 2.7E+05 | 1.8E+05 | 3.3E+05 | 2.2E+05 | 2.0E+05 | 2.7E+05 | 3.1E+05 | 4.0E+05 | 4.0E+05 | 3.0E+05 | 2.6E+05 |
| 2099 | 1 | Q6P158 | Q6P158 | Putative ATPDHX57 | Homo sapien | 155.606 | 8 | 8 | 1.1E+04 | 2.9E+03 | 5.2E+04 | 4.8E+03 | 6.9E+04 | 6.9E+04 | 0.0E+00 | 0.0E+00 | 0.0E+00 | 0.0E+00 | 0.0E+00 | 0.0E+00 |
| 2100 | 1 | Q6P1M3 | Q6P1M3 | LLGL scribblLLGL2 | Homo sapien | 113.449 | 8 | 8 | 6.7E+04 | 2.6E+04 | 3.5E+04 | 3.6E+04 | 7.1E+04 | 1.1E+04 | 4.8E+03 | 2.4E+04 | 2.4E+04 | 1.7E+04 | 9.4E+03 | 0.0E+00 |
| 2101 | 1 | Q6P1N9 | Q6P1N9 | DeoxyribonuTATDN1 | Homo sapien | 33.6 | 8 | 8 | 1.1E+07 | 1.1E+07 | 1.2E+07 | 1.2E+07 | 1.1E+07 | 7.8E+06 | 2.0E+07 | 1.1E+07 | 7.8E+06 | 1.1E+07 | 2.2E+07 | 1.9E+07 |
| 2102 | 1 | Q6UB35 | Q6UB35 | MonofunctionMTHFD1L | Homo sapien | 105.788 | 8 | 8 | 5.4E+04 | 6.7E+04 | 4.5E+04 | 4.6E+04 | 0.0E+00 | 4.2E+04 | 0.0E+00 | 0.0E+00 | 3.6E+04 | 5.8E+04 | 0.0E+00 | 3.9E+04 |
| 2103 | 1 | Q6ZS17 | Q6ZS17 | Rho family-intRIPOR1 | Homo sapien | 132.309 | 8 | 8 | 5.1E+05 | 7.2E+04 | 2.8E+05 | 2.6E+05 | 2.8E+05 | 8.0E+04 | 3.6E+05 | 4.2E+05 | 1.4E+05 | 4.8E+05 | 1.6E+05 | 3.5E+05 |
| 2104 | 1 | Q7L591 | Q7L591 | Docking protDOK3 | Homo sapien | 53.29 | 8 | 8 | 8.2E+06 | 9.9E+06 | 9.0E+06 | 9.0E+06 | 8.2E+06 | 8.1E+06 | 6.4E+06 | 6.5E+06 | 7.1E+06 | 8.2E+06 | 7.4E+06 | 7.7E+06 |
| 2105 | 1 | Q7L9B9 | Q7L9B9 | EndonucleaseEEPD1 | Homo sapien | 62.4 | 8 | 8 | 6.4E+04 | 8.4E+07 | 9.0E+04 | 6.5E+05 | 3.9E+04 | 8.8E+05 | 3.1E+07 | 4.1E+07 | 3.4E+07 | 1.9E+05 | 1.2E+05 | 3.4E+07 |
| 2106 | 1 | Q7Z3V4 | Q7Z3V4 | Ubiquitin-protUBE3B | Homo sapien | 123.1 | 8 | 8 | 3.0E+05 | 3.4E+05 | 1.8E+05 | 2.3E+05 | 2.6E+05 | 1.6E+05 | 2.1E+05 | 4.6E+04 | 1.5E+05 | 1.7E+05 | 6.7E+04 | 1.9E+05 |
| 2107 | 1 | Q7Z6B0 | Q7Z6B0 | Coiled-coil doCCDC91 | Homo sapien | 49.972 | 8 | 8 | 4.5E+04 | 3.3E+04 | 2.9E+04 | 0.0E+00 | 5.0E+04 | 4.2E+04 | 8.3E+04 | 0.0E+00 | 5.7E+06 | 8.9E+04 | 7.9E+06 | 1.1E+05 |
| 2108 | 1 | Q7Z6I6 | Q7Z6I6 | Rho GTPaseARHGAP30 | Homo sapien | 118.585 | 9 | 8 | 4.3E+06 | 3.9E+06 | 5.5E+06 | 4.6E+06 | 4.4E+06 | 4.7E+06 | 2.6E+06 | 3.3E+06 | 3.5E+06 | 3.1E+06 | 3.0E+06 | 2.6E+06 |
| 2109 | 1 | Q86SR1 | Q86SR1 | Polypeptide NGALNT10 | Homo sapien | 68.991 | 9 | 8 | 2.4E+05 | 4.6E+05 | 1.6E+05 | 8.9E+04 | 1.2E+05 | 3.7E+04 | 2.0E+05 | 1.7E+05 | 2.7E+05 | 1.9E+05 | 1.5E+05 | 9.3E+04 |
| 2110 | 1 | Q86TV6 | Q86TV6 | TetratricopepTTC7B | Homo sapien | 94.182 | 8 | 8 | 1.3E+05 | 1.1E+05 | 1.3E+05 | 2.2E+05 | 2.3E+05 | 1.8E+05 | 2.6E+05 | 1.9E+05 | 2.0E+05 | 2.5E+05 | 2.4E+05 | 1.8E+05 |
| 2111 | 1 | Q8IUI8 | Q8IUI8 | Cytokine recCRLF3 | Homo sapien | 49.769 | 8 | 8 | 6.6E+06 | 7.5E+06 | 5.5E+06 | 8.1E+06 | 5.4E+06 | 5.3E+06 | 2.5E+07 | 1.7E+07 | 1.3E+07 | 2.5E+07 | 1.9E+07 | 2.3E+07 |
| 2112 | 1 | Q8IWZ8 | Q8IWZ8 | SURP and G-SUGP1 | Homo sapien | 72.472 | 8 | 8 | 1.4E+05 | 4.3E+05 | 4.6E+05 | 1.1E+05 | 5.6E+05 | 1.7E+05 | 2.9E+05 | 2.4E+05 | 1.7E+05 | 3.3E+05 | 2.4E+05 | 3.1E+04 |
| 2113 | 1 | Q8IZL8 | Q8IZL8 | Proline-, glutaPELP1 | Homo sapien | 119.704 | 8 | 8 | 3.1E+05 | 2.8E+04 | 4.1E+05 | 2.5E+04 | 5.1E+05 | 2.2E+05 | 4.1E+04 | 3.1E+04 | 1.8E+04 | 2.0E+04 | 1.6E+04 | 1.6E+04 |
| 2114 | 1 | Q8N0X7 | Q8N0X7 | Spartin SPART | Homo sapien | 72.836 | 8 | 8 | 1.2E+06 | 1.7E+06 | 1.4E+06 | 3.0E+06 | 1.8E+06 | 1.4E+06 | 1.2E+06 | 1.1E+06 | 1.1E+06 | 1.4E+06 | 1.1E+06 | 1.1E+06 |
| 2115 | 1 | Q8N884 | Q8N884 | Cyclic GMP-ACGAS | Homo sapien | 58.815 | 8 | 8 | 4.4E+06 | 0.0E+00 | 2.9E+05 | 7.3E+03 | 0.0E+00 | 2.7E+06 | 4.1E+05 | 3.2E+05 | 0.0E+00 | 5.3E+05 | 5.2E+06 | 4.1E+05 |
| 2116 | 1 | Q8NBF6 | Q8NBF6 | Late secretorAVL9 | Homo sapien | 71.946 | 8 | 8 | 1.0E+05 | 8.6E+05 | 9.7E+04 | 1.4E+06 | 2.8E+04 | 1.3E+06 | 2.2E+06 | 1.2E+06 | 0.0E+00 | 0.0E+00 | 1.8E+04 | 9.2E+04 |
| 2117 | 1 | Q8NBJ5 | Q8NBJ5 | Procollagen gCOLGALT1 | Homo sapien | 71.637 | 8 | 8 | 1.4E+05 | 1.5E+05 | 1.0E+05 | 2.6E+05 | 3.2E+05 | 2.8E+05 | 1.2E+05 | 2.6E+05 | 1.7E+05 | 5.2E+05 | 2.2E+05 | 1.7E+05 |
| 2118 | 1 | Q8NDA8 | Q8NDA8 | Maestro heatMROH1 | Homo sapien | 181.252 | 8 | 8 | 4.0E+04 | 1.0E+05 | 7.5E+04 | 3.9E+04 | 6.7E+04 | 8.0E+05 | 1.5E+06 | 6.0E+04 | 1.9E+06 | 2.0E+05 | 2.0E+06 | 1.3E+05 |
| 2119 | 1 | Q8NHQ9 | Q8NHQ9 | ATP-dependeDDX55 | Homo sapien | 68.548 | 8 | 8 | 4.1E+05 | 4.3E+05 | 4.8E+05 | 4.0E+05 | 5.3E+05 | 2.8E+05 | 1.2E+05 | 1.3E+05 | 4.8E+04 | 3.7E+05 | 6.5E+04 | 2.8E+05 |
| 2120 | 1 | Q8TBB5 | Q8TBB5 | Kelch domainKLHDC4 | Homo sapien | 57.893 | 8 | 8 | 3.4E+06 | 6.1E+06 | 7.3E+06 | 6.4E+06 | 9.8E+06 | 6.8E+06 | 3.8E+04 | 5.1E+06 | 0.0E+00 | 1.3E+07 | 3.7E+04 | 7.0E+06 |
| 2121 | 1 | Q8TBZ3 | Q8TBZ3 | WD repeat-coWDR20 | Homo sapien | 62.894 | 8 | 8 | 4.7E+06 | 6.4E+06 | 4.4E+06 | 8.0E+06 | 7.0E+06 | 7.3E+06 | 7.2E+06 | 5.2E+06 | 5.7E+06 | 5.0E+06 | 3.9E+06 | 8.4E+06 |
| 2122 | 1 | Q8TF65 | Q8TF65 | PDZ domain-GIPC2 | Homo sapien | 34.355 | 8 | 8 | 3.2E+06 | 2.6E+05 | 2.1E+05 | 1.8E+06 | 1.1E+06 | 4.8E+05 | 1.1E+06 | 2.1E+06 | 2.4E+04 | 6.3E+05 | 3.0E+04 | 1.4E+06 |
| 2123 | 1 | Q8TF74 | Q8TF74 | WAS/WASL-irWIPF2 | Homo sapien | 46.288 | 8 | 8 | 1.4E+05 | 1.7E+05 | 2.3E+05 | 3.6E+04 | 0.0E+00 | 5.2E+04 | 0.0E+00 | 0.0E+00 | 4.9E+04 | 0.0E+00 | 1.5E+04 | 9.7E+03 |
| 2124 | 1 | Q8WWP7 | Q8WWP7 | GTPase IMAGIMAP1 | Homo sapien | 34.369 | 8 | 8 | 7.6E+05 | 2.3E+06 | 2.6E+06 | 1.5E+06 | 4.2E+06 | 1.6E+06 | 1.3E+06 | 3.2E+06 | 1.6E+06 | 4.8E+06 | 2.3E+06 | 4.0E+06 |
| 2125 | 1 | Q8WWY3 | Q8WWY3 | U4/U6 small rPRPF31 | Homo sapien | 55.456 | 8 | 8 | 2.0E+06 | 3.2E+05 | 1.7E+06 | 1.4E+06 | 1.7E+06 | 1.5E+06 | 2.4E+05 | 2.5E+05 | 3.3E+05 | 6.6E+05 | 7.4E+05 | 1.9E+05 |
| 2126 | 1 | Q8WX93 | Q8WX93 | Palladin PALLD | Homo sapien | 150.587 | 8 | 8 | 6.6E+03 | 2.1E+04 | 1.1E+04 | 1.9E+06 | 9.3E+03 | 8.8E+03 | 0.0E+00 | 0.0E+00 | 5.3E+03 | 7.8E+03 | 0.0E+00 | 2.2E+04 |
| 2127 | 1 | Q8WXF7 | Q8WXF7 | Atlastin-1 ATL1 | Homo sapien | 63.546 | 9 | 8 | 1.3E+06 | 1.0E+06 | 1.1E+06 | 9.0E+05 | 9.8E+05 | 8.7E+05 | 1.6E+06 | 1.2E+06 | 1.5E+06 | 1.5E+06 | 1.1E+06 | 1.3E+06 |
| 2128 | 1 | Q8WXX5 | Q8WXX5 | DnaJ homoloDNAJC9 | Homo sapien | 29.907 | 8 | 8 | 7.4E+07 | 6.0E+07 | 7.2E+07 | 8.2E+07 | 7.6E+07 | 8.0E+07 | 7.2E+07 | 5.4E+07 | 6.3E+07 | 5.3E+07 | 5.3E+07 | 7.0E+07 |
| 2129 | 1 | Q8WYJ6 | Q8WYJ6 | Septin-1 SEPTIN1 | Homo sapien | 42.384 | 10 | 8 | 8.4E+05 | 6.0E+05 | 1.6E+06 | 8.7E+05 | 1.6E+06 | 8.7E+05 | 3.0E+05 | 7.4E+05 | 3.4E+05 | 4.1E+05 | 7.7E+05 | 3.0E+05 |
| 2130 | 1 | Q8WZ64 | Q8WZ64 | Arf-GAP withARAP2 | Homo sapien | 193.456 | 8 | 8 | 9.2E+03 | 9.3E+03 | 0.0E+00 | 0.0E+00 | 7.7E+03 | 0.0E+00 | 0.0E+00 | 0.0E+00 | 0.0E+00 | 0.0E+00 | 0.0E+00 | 0.0E+00 |
| 2131 | 1 | Q92551 | Q92551 | Inositol hexaIP6K1 | Homo sapien | 50.236 | 8 | 8 | 7.2E+05 | 4.6E+05 | 4.9E+05 | 7.8E+05 | 6.8E+05 | 7.6E+05 | 3.3E+05 | 4.4E+05 | 2.6E+05 | 6.5E+05 | 4.6E+05 | 5.0E+05 |
| 2132 | 1 | Q969G3 | Q969G3 | SWI/SNF-relaSMARCE1 | Homo sapien | 46.65 | 8 | 8 | 8.1E+05 | 6.2E+05 | 1.1E+06 | 4.7E+05 | 9.3E+05 | 5.5E+05 | 6.4E+05 | 9.1E+05 | 5.0E+05 | 7.6E+05 | 6.3E+05 | 4.8E+05 |
| 2133 | 1 | Q969V3 | Q969V3 | BOS complexNCLN | Homo sapien | 62.974 | 8 | 8 | 4.6E+05 | 7.7E+05 | 6.6E+05 | 6.8E+05 | 6.6E+05 | 5.6E+05 | 3.0E+05 | 5.4E+05 | 5.4E+05 | 7.0E+05 | 9.8E+05 | 2.9E+05 |
| 2134 | 1 | Q969V6 | Q969V6 | Myocardin-reMRTFA | Homo sapien | 98.918 | 8 | 8 | 4.6E+05 | 1.9E+05 | 1.9E+05 | 3.4E+05 | 2.4E+05 | 2.3E+05 | 2.7E+05 | 4.3E+05 | 2.4E+05 | 2.3E+05 | 2.2E+05 | 3.3E+05 |
| 2135 | 1 | Q969X5 | Q969X5 | Endoplasmic ERGIC1 | Homo sapien | 32.59 | 8 | 8 | 5.1E+06 | 7.1E+06 | 7.0E+06 | 6.3E+06 | 8.1E+06 | 8.8E+06 | 9.4E+06 | 9.9E+06 | 6.9E+06 | 1.1E+07 | 1.2E+07 | 8.4E+06 |
| 2136 | 1 | Q96A33 | Q96A33 | PAT complexCCDC47 | Homo sapien | 55.874 | 8 | 8 | 5.7E+05 | 6.2E+05 | 7.3E+05 | 8.3E+05 | 1.4E+06 | 8.2E+05 | 9.6E+05 | 1.1E+06 | 8.1E+05 | 1.4E+06 | 8.6E+05 | 1.1E+06 |
| 2137 | 1 | Q96GG9 | Q96GG9 | DCN1-like prDCUN1D1 | Homo sapien | 30.122 | 9 | 8 | 1.6E+06 | 1.7E+06 | 2.2E+06 | 2.2E+06 | 1.7E+06 | 1.7E+06 | 2.6E+06 | 3.7E+06 | 2.7E+06 | 3.6E+06 | 3.6E+06 | 4.0E+06 |

| | | | | | | | | | | | | | | | | | | | | | |
|---|---|---|---|---|---|---|---|---|---|---|---|---|---|---|---|---|---|---|---|---|---|
| 2618 | 1 | Q9UIM3 | Q9UIM3 | FK506-bindin | FKBPL | Homo sapien | 38.174 | 7 | 7 | 2.7E+06 | 6.9E+04 | 1.5E+06 | 1.5E+06 | 1.7E+06 | 1.7E+06 | 2.2E+06 | 2.3E+06 | 1.8E+06 | 2.0E+06 | 1.5E+06 | 2.7E+06 |
| 2619 | 1 | Q9UKI8 | Q9UKI8 | Serine/threon | TLK1 | Homo sapien | 86.698 | 10 | 7 | 5.4E+04 | 2.0E+04 | 3.6E+04 | 3.1E+04 | 3.5E+04 | 2.0E+04 | 7.4E+03 | 3.2E+04 | 3.8E+04 | 2.9E+04 | 3.4E+04 | 1.3E+04 |
| 2620 | 1 | Q9UKX7 | Q9UKX7 | Nuclear pore | NUP50 | Homo sapien | 50.145 | 7 | 7 | 3.1E+05 | 3.3E+05 | 7.8E+05 | 7.1E+05 | 5.4E+05 | 3.4E+05 | 3.5E+05 | 5.2E+05 | 4.3E+05 | 1.1E+06 | 7.5E+05 | 7.4E+05 |
| 2621 | 1 | Q9UL03 | Q9UL03 | Integrator co | INTS6 | Homo sapien | 100.392 | 8 | 7 | 1.4E+05 | 3.3E+05 | 1.8E+05 | 0.0E+00 | 6.3E+05 | 0.0E+00 | 8.7E+03 | 1.6E+05 | 6.8E+03 | 1.1E+05 | 0.0E+00 | 0.0E+00 |
| 2622 | 1 | Q9ULC5 | Q9ULC5 | Long-chain-fa | ACSL5 | Homo sapien | 75.991 | 7 | 7 | 5.0E+06 | 4.7E+06 | 5.9E+06 | 5.2E+06 | 7.7E+06 | 6.3E+06 | 6.9E+06 | 6.8E+06 | 6.4E+06 | 7.8E+06 | 5.4E+06 | 6.6E+06 |
| 2623 | 1 | Q9ULL8 | Q9ULL8 | Protein Shroo | SHROOM4 | Homo sapien | 164.861 | 8 | 7 | 2.8E+04 | 5.6E+04 | 7.0E+04 | 3.2E+04 | 6.2E+04 | 2.2E+04 | 3.0E+04 | 0.0E+00 | 3.3E+04 | 1.1E+06 | 1.3E+04 | 0.0E+00 |
| 2624 | 1 | Q9ULP9 | Q9ULP9 | TBC1 domain | TBC1D24 | Homo sapien | 62.922 | 7 | 7 | 7.0E+06 | 6.0E+06 | 4.3E+06 | 7.7E+06 | 6.8E+06 | 7.5E+06 | 1.1E+07 | 1.2E+07 | 9.8E+06 | 1.2E+07 | 9.6E+06 | 1.1E+07 |
| 2625 | 1 | Q9ULQ0 | Q9ULQ0 | Striatin-intera | STRIP2 | Homo sapien | 95.36 | 7 | 7 | 3.2E+05 | 3.1E+05 | 3.4E+05 | 3.7E+05 | 3.3E+05 | 3.4E+05 | 9.2E+05 | 6.2E+05 | 8.5E+05 | 1.1E+06 | 1.0E+06 | 9.8E+05 |
| 2626 | 1 | Q9ULU4 | Q9ULU4 | MYND-type z | ZMYND8 | Homo sapien | 131.696 | 7 | 7 | 7.2E+04 | 1.6E+05 | 1.0E+05 | 1.3E+05 | 3.4E+05 | 1.0E+05 | 1.3E+04 | 4.1E+04 | 1.1E+05 | 4.5E+04 | 2.0E+04 | 3.4E+04 |
| 2627 | 1 | Q9UM11 | Q9UM11 | Fizzy-related | FZR1 | Homo sapien | 55.178 | 7 | 7 | 5.3E+04 | 0.0E+00 | 0.0E+00 | 0.0E+00 | 2.2E+04 | 1.7E+04 | 0.0E+00 | 0.0E+00 | 0.0E+00 | 0.0E+00 | 0.0E+00 | 0.0E+00 |
| 2628 | 1 | Q9UNH7 | Q9UNH7 | Sorting nexin | SNX6 | Homo sapien | 46.647 | 8 | 7 | 5.4E+06 | 6.4E+06 | 6.1E+06 | 6.6E+06 | 4.7E+06 | 4.3E+06 | 6.5E+06 | 3.9E+06 | 4.3E+06 | 6.1E+06 | 7.6E+06 | 5.7E+06 |
| 2629 | 1 | Q9UNW1 | Q9UNW1 | Multiple inosit | MINPP1 | Homo sapien | 55.051 | 7 | 7 | 2.7E+07 | 5.2E+07 | 4.0E+07 | 6.2E+07 | 7.2E+07 | 5.6E+07 | 2.8E+07 | 2.9E+06 | 1.3E+07 | 1.8E+07 | 2.6E+06 | 2.5E+06 |
| 2630 | 1 | Q9UPP1 | Q9UPP1 | Histone lysin | PHF8 | Homo sapien | 117.886 | 8 | 7 | 1.0E+06 | 7.6E+05 | 6.6E+05 | 0.0E+00 | 1.2E+06 | 8.2E+05 | 0.0E+00 | 0.0E+00 | 7.7E+03 | 0.0E+00 | 0.0E+00 | 0.0E+00 |
| 2631 | 1 | Q9UQL6 | Q9UQL6 | Histone deac | HDAC5 | Homo sapien | 121.978 | 7 | 7 | 1.4E+05 | 7.3E+04 | 1.1E+06 | 1.5E+05 | 9.6E+05 | 1.8E+05 | 2.0E+06 | 7.8E+04 | 5.2E+05 | 0.0E+00 | 4.5E+05 | 4.8E+05 |
| 2632 | 1 | Q9Y224 | Q9Y224 | RNA transcri | RTRAF | Homo sapien | 28.069 | 7 | 7 | 5.8E+06 | 7.9E+06 | 6.6E+06 | 7.6E+06 | 7.2E+06 | 5.4E+06 | 7.9E+05 | 7.2E+06 | 3.9E+06 | 1.0E+07 | 8.9E+06 | 8.2E+05 |
| 2633 | 1 | Q9Y274 | Q9Y274 | Type 2 lactos | ST3GAL6 | Homo sapien | 38.214 | 7 | 7 | 1.4E+06 | 1.8E+06 | 9.9E+05 | 1.4E+06 | 7.9E+05 | 4.2E+05 | 7.5E+05 | 3.9E+05 | 8.3E+05 | 9.9E+05 | 8.7E+05 | 1.3E+06 |
| 2634 | 1 | Q9Y2J2 | Q9Y2J2 | Band 4.1-like | EPB41L3 | Homo sapien | 120.678 | 7 | 7 | 3.1E+06 | 1.5E+06 | 3.2E+06 | 1.5E+05 | 1.6E+06 | 1.4E+06 | 2.2E+05 | 1.7E+05 | 2.1E+05 | 2.2E+05 | 2.6E+05 | 2.2E+04 |
| 2635 | 1 | Q9Y2J8 | Q9Y2J8 | Protein-argini | PADI2 | Homo sapien | 75.563 | 7 | 7 | 3.9E+04 | 1.8E+05 | 1.6E+06 | 9.9E+04 | 7.2E+05 | 1.0E+06 | 2.0E+06 | 1.4E+06 | 1.4E+06 | 1.3E+06 | 1.7E+06 | 1.6E+06 |
| 2636 | 1 | Q9Y2R2 | Q9Y2R2 | Tyrosine-prot | PTPN22 | Homo sapien | 91.705 | 7 | 7 | 4.8E+04 | 3.6E+04 | 4.3E+04 | 0.0E+00 | 1.9E+04 | 4.4E+04 | 0.0E+00 | 0.0E+00 | 0.0E+00 | 0.0E+00 | 0.0E+00 | 0.0E+00 |
| 2637 | 1 | Q9Y2X7 | Q9Y2X7 | ARF GTPase | GIT1 | Homo sapien | 84.339 | 7 | 7 | 2.4E+06 | 2.6E+06 | 2.1E+06 | 2.5E+06 | 2.8E+06 | 2.4E+06 | 1.9E+06 | 1.7E+06 | 2.6E+06 | 1.4E+06 | 2.1E+06 | 1.6E+06 |
| 2638 | 1 | Q9Y3A5 | Q9Y3A5 | Ribosome ma | SBDS | Homo sapien | 28.764 | 7 | 7 | 1.8E+07 | 2.3E+07 | 2.6E+07 | 2.6E+07 | 1.6E+07 | 1.9E+07 | 1.4E+07 | 8.8E+06 | 1.1E+07 | 1.5E+07 | 1.6E+07 | 2.9E+07 |
| 2639 | 1 | Q9Y3D9 | Q9Y3D9 | Small ribosom | MRPS23 | Homo sapien | 21.769 | 7 | 7 | 1.1E+06 | 9.9E+05 | 1.6E+06 | 1.4E+06 | 2.1E+06 | 1.3E+06 | 9.5E+05 | 1.3E+06 | 4.7E+05 | 7.0E+05 | 1.0E+06 | 3.7E+05 |
| 2640 | 1 | Q9Y484 | Q9Y484 | WD repeat do | WDR45 | Homo sapien | 39.869 | 7 | 7 | 6.6E+06 | 5.4E+06 | 3.8E+06 | 6.1E+06 | 5.7E+06 | 5.7E+06 | 2.4E+06 | 2.0E+06 | 4.4E+06 | 2.1E+06 | 2.6E+06 | 2.3E+06 |
| 2641 | 1 | Q9Y4E5 | Q9Y4E5 | E3 SUMO-pr | ZNF451 | Homo sapien | 121.488 | 7 | 7 | 2.6E+04 | 1.3E+05 | 8.2E+04 | 1.7E+05 | 1.1E+05 | 4.2E+04 | 4.7E+04 | 2.2E+05 | 1.4E+05 | 0.0E+00 | 8.9E+04 | 1.1E+05 |
| 2642 | 1 | Q9Y4K3 | Q9Y4K3 | TNF receptor | TRAF6 | Homo sapien | 59.572 | 7 | 7 | 4.1E+05 | 7.6E+04 | 2.9E+05 | 4.0E+05 | 5.9E+05 | 2.7E+05 | 6.6E+05 | 3.4E+05 | 3.0E+05 | 6.0E+05 | 5.8E+05 | 6.8E+05 |
| 2643 | 1 | Q9Y512 | Q9Y512 | Sorting and a | SAMM50 | Homo sapien | 51.978 | 7 | 7 | 2.0E+06 | 2.5E+06 | 2.3E+06 | 2.2E+06 | 2.1E+06 | 2.4E+06 | 2.8E+06 | 2.7E+06 | 2.4E+06 | 1.5E+06 | 2.3E+06 | 2.7E+06 |
| 2644 | 1 | Q9Y608 | Q9Y608 | Leucine-rich r | LRRFIP2 | Homo sapien | 82.173 | 7 | 7 | 1.7E+07 | 2.0E+07 | 2.2E+07 | 2.2E+07 | 1.6E+07 | 2.4E+07 | 3.2E+07 | 2.9E+07 | 3.0E+07 | 3.7E+07 | 2.9E+07 | 3.3E+07 |
| 2645 | 1 | Q9Y6P5 | Q9Y6P5 | Sestrin-1 | SESN1 | Homo sapien | 56.557 | 8 | 7 | 9.3E+03 | 0.0E+00 | 6.2E+05 | 8.2E+04 | 1.1E+04 | 0.0E+00 | 1.0E+06 | 1.2E+06 | 9.3E+04 | 1.1E+06 | 1.2E+06 | 9.4E+05 |
| 2646 | 1 | A0MZ66 | A0MZ66 | Shootin-1 | SHTN1 | Homo sapien | 71.642 | 6 | 6 | 3.6E+04 | 0.0E+00 | 3.6E+04 | 0.0E+00 | 5.4E+04 | 3.9E+04 | 4.9E+04 | 0.0E+00 | 0.0E+00 | 5.2E+04 | 2.5E+05 | 0.0E+00 |
| 2647 | 1 | B1AK53 | B1AK53 | Espin | ESPN | Homo sapien | 91.732 | 6 | 6 | 6.8E+05 | 5.4E+05 | 2.2E+05 | 4.8E+05 | 1.1E+06 | 6.5E+05 | 1.0E+06 | 8.4E+05 | 6.2E+05 | 5.2E+05 | 6.0E+05 | 5.3E+05 |
| 2648 | 1 | O00161 | O00161 | Synaptosom | SNAP23 | Homo sapien | 23.354 | 6 | 6 | 2.6E+06 | 2.8E+06 | 2.3E+06 | 3.1E+06 | 3.2E+06 | 2.7E+06 | 5.3E+06 | 4.4E+06 | 5.6E+06 | 6.1E+06 | 5.0E+06 | 5.5E+06 |
| 2649 | 1 | O00291 | O00291 | Huntingtin-int | HIP1 | Homo sapien | 116.223 | 6 | 6 | 4.5E+06 | 1.0E+06 | 8.2E+05 | 4.6E+05 | 5.8E+06 | 7.4E+05 | 6.2E+05 | 8.3E+05 | 5.9E+05 | 8.5E+05 | 9.3E+05 | 8.7E+05 |
| 2650 | 1 | O00299 | O00299 | Chloride intra | CLIC1 | Homo sapien | 26.922 | 6 | 6 | 4.1E+07 | 2.8E+07 | 3.2E+07 | 3.1E+07 | 4.1E+07 | 2.8E+07 | 7.4E+07 | 6.9E+07 | 6.8E+07 | 9.0E+07 | 6.9E+07 | 7.2E+07 |
| 2651 | 1 | O00330 | O00330 | Pyruvate deh | PDHX | Homo sapien | 54.124 | 6 | 6 | 7.0E+05 | 0.0E+00 | 8.4E+05 | 0.0E+00 | 0.0E+00 | 0.0E+00 | 0.0E+00 | 5.9E+04 | 7.3E+04 | 7.4E+04 | 0.0E+00 | 4.4E+04 |
| 2652 | 1 | O00442 | O00442 | RNA 3'-termi | RTCA | Homo sapien | 39.335 | 6 | 6 | 2.5E+06 | 2.8E+06 | 3.1E+06 | 6.3E+06 | 6.0E+06 | 2.4E+06 | 9.0E+06 | 7.8E+06 | 4.4E+06 | 3.2E+06 | 6.7E+06 | 7.6E+06 |
| 2653 | 1 | O00443 | O00443 | Phosphatidyl | PIK3C2A | Homo sapien | 190.684 | 6 | 6 | 1.7E+04 | 1.7E+04 | 1.2E+04 | 4.2E+04 | 1.2E+04 | 7.4E+03 | 0.0E+00 | 8.1E+03 | 0.0E+00 | 0.0E+00 | 1.1E+04 | 0.0E+00 |
| 2654 | 1 | O00487 | O00487 | 26S proteaso | PSMD14 | Homo sapien | 34.577 | 6 | 6 | 1.9E+07 | 3.0E+07 | 2.5E+07 | 4.3E+07 | 2.6E+07 | 2.7E+07 | 4.7E+07 | 3.5E+07 | 2.5E+07 | 3.4E+07 | 4.1E+07 | 4.7E+07 |
| 2655 | 1 | O00499 | O00499 | Myc box-dep | BIN1 | Homo sapien | 64.701 | 6 | 6 | 0.0E+00 | 2.0E+04 | 0.0E+00 | 2.0E+05 | 0.0E+00 | 1.0E+05 | 1.2E+05 | 0.0E+00 | 0.0E+00 | 0.0E+00 | 1.1E+05 | 2.6E+04 |
| 2656 | 1 | O14562 | O14562 | Ubiquitin dom | UBFD1 | Homo sapien | 33.382 | 6 | 6 | 1.3E+07 | 1.2E+07 | 1.3E+07 | 1.5E+07 | 1.3E+07 | 1.4E+07 | 1.0E+07 | 7.4E+06 | 7.3E+06 | 8.4E+06 | 1.3E+07 | 1.0E+07 |
| 2657 | 1 | O14733 | O14733 | Dual specifici | MAP2K7 | Homo sapien | 47.484 | 6 | 6 | 3.2E+05 | 2.6E+05 | 1.9E+05 | 2.2E+05 | 2.2E+05 | 3.0E+05 | 4.6E+05 | 2.2E+05 | 1.9E+05 | 1.9E+05 | 2.0E+05 | 1.9E+05 |
| 2658 | 1 | O15211 | O15211 | Ral guanine n | RGL2 | Homo sapien | 83.549 | 6 | 6 | 8.7E+05 | 3.0E+04 | 2.3E+04 | 5.5E+04 | 3.5E+04 | 2.6E+04 | 3.3E+04 | 6.7E+04 | 3.2E+04 | 4.5E+04 | 9.0E+04 | 6.6E+04 |
| 2659 | 1 | O15247 | O15247 | Chloride intra | CLIC2 | Homo sapien | 28.355 | 6 | 6 | 3.6E+06 | 2.6E+06 | 2.9E+06 | 4.6E+06 | 2.0E+06 | 2.5E+06 | 4.2E+06 | 2.8E+06 | 4.4E+06 | 5.8E+06 | 5.3E+06 | 4.0E+06 |
| 2660 | 1 | O15270 | O15270 | Serine palmit | SPTLC2 | Homo sapien | 62.926 | 6 | 6 | 5.2E+05 | 2.4E+05 | 1.9E+05 | 4.0E+05 | 6.9E+05 | 6.7E+04 | 2.6E+05 | 5.5E+05 | 3.2E+05 | 5.6E+05 | 4.2E+05 | 3.6E+05 |
| 2661 | 1 | O15498 | O15498 | Synaptobrevi | YKT6 | Homo sapien | 22.417 | 6 | 6 | 7.9E+05 | 8.7E+06 | 7.8E+06 | 4.3E+06 | 4.1E+06 | 9.4E+06 | 1.0E+07 | 1.1E+07 | 6.4E+06 | 1.1E+07 | 1.4E+07 | 1.4E+07 |
| 2662 | 1 | O43396 | O43396 | Thioredoxin-l | TXNL1 | Homo sapien | 32.251 | 6 | 6 | 8.2E+06 | 9.9E+06 | 9.6E+06 | 1.0E+07 | 9.0E+06 | 8.2E+06 | 1.5E+07 | 1.7E+07 | 1.2E+07 | 1.5E+07 | 1.7E+07 | 2.1E+07 |
| 2663 | 1 | O43405 | O43405 | Cochlin | COCH | Homo sapien | 59.485 | 6 | 6 | 1.1E+05 | 7.6E+04 | 1.3E+05 | 0.0E+00 | 1.6E+05 | 3.6E+04 | 0.0E+00 | 0.0E+00 | 0.0E+00 | 2.1E+05 | 0.0E+00 | 0.0E+00 |
| 2664 | 1 | O43493 | O43493 | Trans-Golgi n | TGOLN2 | Homo sapien | 45.88 | 6 | 6 | 1.1E+05 | 6.7E+04 | 1.2E+05 | 8.2E+04 | 1.1E+05 | 1.0E+05 | 1.3E+05 | 8.7E+04 | 1.2E+05 | 2.7E+05 | 5.6E+04 | 1.9E+05 |
| 2665 | 1 | O43681 | O43681 | ATPase GET | GET3 | Homo sapien | 38.792 | 6 | 6 | 6.2E+06 | 4.9E+06 | 5.1E+06 | 5.9E+06 | 4.8E+06 | 5.3E+06 | 7.0E+06 | 7.8E+06 | 6.1E+06 | 8.2E+06 | 9.2E+06 | 8.9E+06 |
| 2666 | 1 | O43809 | O43809 | Cleavage and | NUDT21 | Homo sapien | 26.228 | 6 | 6 | 7.7E+05 | 7.2E+05 | 6.2E+05 | 4.2E+05 | 4.9E+05 | 5.5E+05 | 4.6E+05 | 5.0E+05 | 4.0E+05 | 5.4E+05 | 4.4E+05 | 3.3E+05 |
| 2667 | 1 | O43818 | O43818 | U3 small nucl | RRP9 | Homo sapien | 51.841 | 6 | 6 | 3.7E+05 | 5.2E+05 | 7.2E+05 | 5.9E+05 | 7.5E+05 | 4.4E+05 | 3.2E+05 | 4.9E+05 | 3.4E+05 | 4.2E+05 | 2.7E+05 | 2.1E+04 |
| 2668 | 1 | O43826 | O43826 | Glucose-6-ph | SLC37A4 | Homo sapien | 46.36 | 6 | 6 | 7.6E+05 | 9.3E+05 | 7.4E+05 | 8.1E+05 | 1.0E+06 | 7.9E+05 | 8.9E+05 | 1.6E+06 | 1.3E+06 | 1.6E+06 | 8.3E+05 | 1.2E+06 |
| 2669 | 1 | O43861 | O43861 | Probable pho | ATP9B | Homo sapien | 129.306 | 8 | 6 | 2.7E+05 | 2.8E+05 | 2.2E+05 | 2.0E+05 | 2.5E+05 | 1.2E+05 | 1.3E+05 | 1.5E+05 | 1.7E+05 | 1.8E+05 | 1.8E+05 | 1.4E+05 |
| 2670 | 1 | O43933 | O43933 | Peroxisomal | PEX1 | Homo sapien | 142.87 | 6 | 6 | 0.0E+00 | 6.4E+03 | 5.4E+03 | 0.0E+00 | 1.6E+04 | 0.0E+00 | 3.4E+04 | 0.0E+00 | 0.0E+00 | 9.9E+03 | 1.4E+04 | 8.1E+03 |
| 2671 | 1 | O75113 | O75113 | NEDD4-bindi | N4BP1 | Homo sapien | 100.38 | 6 | 6 | 4.2E+04 | 1.8E+04 | 9.3E+03 | 0.0E+00 | 2.1E+05 | 2.7E+05 | 4.6E+04 | 2.7E+04 | 3.7E+04 | 3.3E+05 | 3.5E+05 | 3.3E+04 |
| 2672 | 1 | O75167 | O75167 | Phosphatase | PHACTR2 | Homo sapien | 69.7 | 6 | 6 | 3.9E+05 | 1.8E+05 | 3.7E+05 | 4.3E+05 | 1.8E+05 | 1.9E+05 | 1.5E+05 | 1.2E+05 | 1.3E+05 | 1.3E+05 | 8.3E+04 | 8.5E+04 |
| 2673 | 1 | O75326 | O75326 | Semaphorin-7 | SEMA7A | Homo sapien | 74.824 | 6 | 6 | 4.4E+06 | 6.6E+06 | 1.2E+06 | 2.4E+06 | 2.2E+06 | 8.2E+06 | 1.0E+07 | 1.2E+07 | 6.0E+06 | 9.9E+06 | 6.5E+06 | 1.2E+07 |
| 2674 | 1 | O75368 | O75368 | Adapter SH3 | SH3BGRL | Homo sapien | 12.772 | 6 | 6 | 2.2E+06 | 1.8E+06 | 2.6E+06 | 2.4E+06 | 2.6E+06 | 2.2E+06 | 1.9E+06 | 2.4E+06 | 3.2E+06 | 3.8E+06 | 2.2E+06 | 1.7E+06 |
| 2675 | 1 | O75449 | O75449 | Katanin p60 | KATNA1 | Homo sapien | 55.967 | 6 | 6 | 9.0E+05 | 8.0E+05 | 8.1E+05 | 1.4E+06 | 8.4E+05 | 7.8E+05 | 7.4E+05 | 4.0E+05 | 5.2E+05 | 9.0E+05 | 5.7E+05 | 6.7E+05 |
| 2676 | 1 | O75477 | O75477 | Erlin-1 | ERLIN1 | Homo sapien | 39.172 | 8 | 6 | 7.7E+05 | 7.8E+05 | 7.8E+05 | 2.9E+06 | 9.3E+05 | 7.3E+05 | 1.0E+06 | 5.6E+06 | 1.0E+06 | 1.1E+06 | 9.7E+05 | 1.4E+06 |

[illegible]

| | | | | | | | | | | | | | | | | | |
|---|---|---|---|---|---|---|---|---|---|---|---|---|---|---|---|---|---|
| 3142 | 1 | O15240 | O15240 | Neurosecret | VGF | Homo sapien | 67.26 | 5 | 5 | 0.0E+00 | 0.0E+00 | 0.0E+00 | 0.0E+00 | 0.0E+00 | 0.0E+00 | [illegible] | [illegible] |
| 3143 | 1 | O15519 | O15519 | CASP8 and F | CFLAR | Homo sapien | 55.345 | 5 | 5 | 1.8E+04 | 0.0E+00 | 1.8E+04 | 1.8E+04 | 0.0E+00 | 0.0E+00 | [illegible] | [illegible] |
| 3144 | 1 | O15523 | O15523 | ATP-depend | DDX3Y | Homo sapien | 73.154 | 5 | 5 | 0.0E+00 | 0.0E+00 | 0.0E+00 | 0.0E+00 | 2.7E+06 | 2.8E+06 | [illegible] | [illegible] |
| 3145 | 1 | O43148 | O43148 | mRNA cap g | RNMT | Homo sapien | 54.844 | 5 | 5 | 7.4E+04 | 1.6E+05 | 1.7E+05 | 2.7E+05 | 1.5E+05 | 8.2E+04 | [illegible] | [illegible] |
| 3146 | 1 | O43157 | O43157 | Plexin-B1 | PLXNB1 | Homo sapien | 232.302 | 5 | 5 | 0.0E+00 | 0.0E+00 | 1.3E+06 | 1.7E+06 | 0.0E+00 | 1.6E+06 | [illegible] | [illegible] |
| 3147 | 1 | O43189 | O43189 | PHD finger pr | PHF1 | Homo sapien | 62.106 | 5 | 5 | 2.6E+04 | 2.5E+04 | 4.9E+04 | 3.7E+04 | 3.1E+04 | 1.8E+04 | [illegible] | [illegible] |
| 3148 | 1 | O43237 | O43237 | Cytoplasmic | DYNC1LI2 | Homo sapien | 54.101 | 5 | 5 | 5.5E+05 | 3.2E+04 | 2.8E+05 | 4.9E+05 | 3.0E+05 | 2.5E+05 | [illegible] | [illegible] |
| 3149 | 1 | O43294 | O43294 | Transforming | TGFB1I1 | Homo sapien | 49.814 | 5 | 5 | 2.6E+06 | 2.7E+06 | 2.0E+06 | 2.8E+06 | 2.3E+06 | 2.4E+06 | [illegible] | [illegible] |
| 3150 | 1 | O43324 | O43324 | Eukaryotic tr | EEF1E1 | Homo sapien | 19.807 | 5 | 5 | 1.7E+06 | 1.6E+06 | 1.7E+06 | 1.8E+06 | 2.9E+06 | 3.5E+06 | [illegible] | [illegible] |
| 3151 | 1 | O43929 | O43929 | Origin recogr | ORC4 | Homo sapien | 50.38 | 5 | 5 | 7.0E+04 | 4.1E+04 | 6.7E+03 | 5.2E+04 | 5.5E+04 | 2.7E+04 | [illegible] | [illegible] |
| 3152 | 1 | O60318 | O60318 | Germinal-cer | MCM3AP | Homo sapien | 218.405 | 5 | 5 | 3.7E+04 | 0.0E+00 | 4.5E+04 | 2.4E+04 | 1.9E+04 | 0.0E+00 | [illegible] | [illegible] |
| 3153 | 1 | O60486 | O60486 | Plexin-C1 | PLXNC1 | Homo sapien | 175.744 | 5 | 5 | 2.7E+05 | 0.0E+00 | 2.7E+05 | 0.0E+00 | 6.4E+05 | 3.2E+05 | [illegible] | [illegible] |
| 3154 | 1 | O60733 | O60733 | 85/88 kDa ca | PLA2G6 | Homo sapien | 89.906 | 5 | 5 | 1.7E+05 | 7.3E+04 | 1.2E+05 | 8.1E+04 | 2.9E+05 | 3.6E+05 | [illegible] | [illegible] |
| 3155 | 1 | O60861 | O60861 | Growth arres | GAS7 | Homo sapien | 54.373 | 5 | 5 | 1.3E+06 | 3.3E+06 | 9.3E+05 | 9.7E+05 | 8.5E+05 | 1.2E+06 | [illegible] | [illegible] |
| 3156 | 1 | O60870 | O60870 | DNA/RNA-bir | KIN | Homo sapien | 45.372 | 5 | 5 | 1.0E+05 | 2.3E+05 | 1.8E+05 | 1.9E+05 | 2.5E+05 | 1.1E+05 | [illegible] | [illegible] |
| 3157 | 1 | O75063 | O75063 | Glycosamino | FAM20B | Homo sapien | 46.435 | 5 | 5 | 4.2E+04 | 4.1E+04 | 4.0E+04 | 4.8E+04 | 4.1E+04 | 4.7E+04 | [illegible] | [illegible] |
| 3158 | 1 | O75298 | O75298 | Reticulon-2 | RTN2 | Homo sapien | 59.265 | 5 | 5 | 1.4E+06 | 1.6E+06 | 7.2E+05 | 9.5E+05 | 6.9E+05 | 6.2E+05 | [illegible] | [illegible] |
| 3159 | 1 | O75448 | O75448 | Mediator of R | MED24 | Homo sapien | 110.306 | 5 | 5 | 9.2E+05 | 0.0E+00 | 5.6E+05 | 8.9E+05 | 8.7E+05 | 1.4E+04 | [illegible] | [illegible] |
| 3160 | 1 | O75554 | O75554 | WW domain-b | WBP4 | Homo sapien | 42.506 | 5 | 5 | 1.4E+06 | 1.4E+06 | 1.3E+06 | 1.6E+06 | 1.4E+06 | 2.0E+06 | [illegible] | [illegible] |
| 3161 | 1 | O75688 | O75688 | Protein phos | PPM1B | Homo sapien | 52.643 | 5 | 5 | 1.6E+05 | 1.5E+05 | 1.8E+05 | 2.1E+05 | 2.8E+05 | 2.4E+05 | [illegible] | [illegible] |
| 3162 | 1 | O75689 | O75689 | Arf-GAP with | ADAP1 | Homo sapien | 43.395 | 6 | 5 | 3.5E+06 | 3.4E+06 | 3.6E+06 | 8.2E+04 | 3.1E+06 | 3.6E+06 | [illegible] | [illegible] |
| 3163 | 1 | O75695 | O75695 | Protein XRP2 | RP2 | Homo sapien | 39.641 | 5 | 5 | 4.2E+06 | 3.8E+06 | 2.7E+06 | 3.6E+06 | 3.0E+06 | 2.6E+06 | [illegible] | [illegible] |
| 3164 | 1 | O75718 | O75718 | Cartilage-ass | CRTAP | Homo sapien | 46.562 | 5 | 5 | 8.9E+03 | 0.0E+00 | 1.1E+04 | 3.3E+04 | 2.3E+05 | 4.1E+04 | [illegible] | [illegible] |
| 3165 | 1 | O75787 | O75787 | Renin recept | ATP6AP2 | Homo sapien | 39.008 | 5 | 5 | 2.1E+05 | 2.5E+05 | 2.2E+05 | 3.0E+05 | 1.8E+05 | 2.4E+05 | [illegible] | [illegible] |
| 3166 | 1 | O75909 | O75909 | Cyclin-K | CCNK | Homo sapien | 64.242 | 5 | 5 | 2.2E+05 | 2.0E+05 | 2.9E+05 | 6.8E+04 | 2.3E+05 | 6.0E+04 | [illegible] | [illegible] |
| 3167 | 1 | O75995 | O75995 | SAM and SH | SASH3 | Homo sapien | 41.594 | 5 | 5 | 3.9E+06 | 4.4E+06 | 3.8E+06 | 3.5E+06 | 4.7E+06 | 3.7E+06 | [illegible] | [illegible] |
| 3168 | 1 | O94769 | O94769 | Extracellular | ECM2 | Homo sapien | 79.79 | 5 | 5 | 2.1E+05 | 2.2E+05 | 1.5E+05 | 2.3E+05 | 3.9E+05 | 2.5E+05 | [illegible] | [illegible] |
| 3169 | 1 | O94880 | O94880 | PHD finger pr | PHF14 | Homo sapien | 106.987 | 5 | 5 | 7.6E+03 | 1.1E+04 | 6.8E+05 | 2.4E+04 | 7.5E+05 | 5.6E+05 | [illegible] | [illegible] |
| 3170 | 1 | O94889 | O94889 | Kelch-like pr | KLHL18 | Homo sapien | 63.639 | 5 | 5 | 1.3E+05 | 3.2E+05 | 1.0E+05 | 1.5E+05 | 1.3E+05 | 1.2E+05 | [illegible] | [illegible] |
| 3171 | 1 | O94916 | O94916 | Nuclear facto | NFAT5 | Homo sapien | 165.765 | 5 | 5 | 1.4E+06 | 9.8E+05 | 1.5E+06 | 1.6E+06 | 1.8E+06 | 1.6E+05 | [illegible] | [illegible] |
| 3172 | 1 | O95182 | O95182 | NADH dehyd | NDUFA7 | Homo sapien | 12.551 | 5 | 5 | 1.1E+05 | 1.1E+05 | 3.1E+05 | 1.6E+05 | 1.2E+05 | 1.4E+05 | [illegible] | [illegible] |
| 3173 | 1 | O95208 | O95208 | Epsin-2 | EPN2 | Homo sapien | 68.48 | 5 | 5 | 4.6E+06 | 9.8E+05 | 8.3E+05 | 8.0E+05 | 9.0E+05 | 1.6E+06 | [illegible] | [illegible] |
| 3174 | 1 | O95400 | O95400 | CD2 antigen | CD2BP2 | Homo sapien | 37.647 | 5 | 5 | 3.1E+06 | 3.0E+06 | 2.4E+06 | 8.4E+04 | 2.6E+06 | 3.4E+06 | [illegible] | [illegible] |
| 3175 | 1 | O95630 | O95630 | STAM-bindin | STAMBP | Homo sapien | 48.077 | 5 | 5 | 5.2E+06 | 3.4E+06 | 4.5E+06 | 4.2E+06 | 5.3E+06 | 6.3E+06 | [illegible] | [illegible] |
| 3176 | 1 | O95822 | O95822 | Malonyl-CoA | MLYCD | Homo sapien | 55.004 | 5 | 5 | 0.0E+00 | 0.0E+00 | 6.5E+03 | 0.0E+00 | 0.0E+00 | 0.0E+00 | [illegible] | [illegible] |
| 3177 | 1 | O95825 | O95825 | Quinone oxid | CRYZL1 | Homo sapien | 38.697 | 5 | 5 | 1.6E+06 | 1.2E+06 | 1.4E+06 | 1.5E+06 | 1.0E+06 | 1.4E+06 | [illegible] | [illegible] |
| 3178 | 1 | O96008 | O96008 | Mitochondrial | TOMM40 | Homo sapien | 37.893 | 5 | 5 | 4.8E+05 | 6.1E+05 | 8.6E+05 | 6.9E+05 | 8.9E+05 | 5.3E+05 | [illegible] | [illegible] |
| 3179 | 1 | O96019 | O96019 | Actin-like pro | ACTL6A | Homo sapien | 47.46 | 8 | 5 | 4.7E+05 | 6.0E+05 | 6.6E+05 | 5.5E+05 | 6.7E+05 | 3.6E+05 | [illegible] | [illegible] |
| 3180 | 1 | P00367 | P00367 | Glutamate de | GLUD1 | Homo sapien | 61.398 | 12 | 5 | 7.6E+05 | 1.0E+06 | 1.3E+06 | 1.1E+06 | 9.5E+05 | 6.7E+05 | [illegible] | [illegible] |
| 3181 | 1 | P00736 | P00736 | Complement | C1R | Homo sapien | 80.119 | 5 | 5 | 7.6E+07 | 7.8E+07 | 3.9E+07 | 8.2E+07 | 6.8E+07 | 8.5E+07 | [illegible] | [illegible] |
| 3182 | 1 | P00738 | P00738 | Haptoglobin | HP | Homo sapien | 45.205 | 9 | 5 | 6.7E+07 | 2.9E+07 | 3.0E+07 | 1.6E+07 | 3.0E+07 | 2.8E+07 | [illegible] | [illegible] |
| 3183 | 1 | P00739 | P00739 | Haptoglobin-r | HPR | Homo sapien | 39.03 | 5 | 5 | 4.4E+07 | 1.5E+07 | 1.8E+07 | 4.2E+07 | 2.9E+07 | 3.5E+07 | [illegible] | [illegible] |
| 3184 | 1 | P00748 | P00748 | Coagulation f | F12 | Homo sapien | 67.792 | 5 | 5 | 1.3E+07 | 1.8E+07 | 6.9E+06 | 1.5E+07 | 1.5E+07 | 1.1E+07 | [illegible] | [illegible] |
| [illegible] | | | | | | | | | | | | | | | | | |

| | | | | | | | | | | | | | | |
|---|---|---|---|---|---|---|---|---|---|---|---|---|---|---|
| 3666 | 1 | Q9Y6V7 | Q9Y6V7 | Probable ATIDDX49 | Homo sapien | 54.227 | 5 | 5 | 0.0E+00 | 0.0E+00 | 2.4E+04 | 5.4E+03 | 3.2E+05 | 0.0E+00 |
| 3667 | 1 | Q9Y6X9 | Q9Y6X9 | ATPase MOFMORC2 | Homo sapien | 117.825 | 5 | 5 | 1.9E+04 | 1.3E+05 | 0.0E+00 | 0.0E+00 | 1.7E+04 | 0.0E+00 |
| 3668 | 1 | A0A075B6K4 | A0A075B6K4 | Immunoglobu IGLV3-10 | Homo sapien | 12.438 | 4 | 4 | 3.7E+06 | 6.3E+06 | 7.1E+06 | 9.4E+06 | 4.8E+06 | 1.7E+06 |
| 3669 | 1 | A0JNW5 | A0JNW5 | Bridge-like lipBLTP3B | Homo sapien | 164.202 | 4 | 4 | 1.3E+04 | 1.5E+04 | 0.0E+00 | 2.9E+06 | 0.0E+00 | 0.0E+00 |
| 3670 | 1 | A2RUC4 | A2RUC4 | tRNA wybutoTYW5 | Homo sapien | 36.549 | 4 | 4 | 2.4E+04 | 7.0E+04 | 4.5E+04 | 5.4E+04 | 1.9E+04 | 2.7E+04 |
| 3671 | 1 | A4D1E9 | A4D1E9 | GTP-binding GTPBP10 | Homo sapien | 42.934 | 4 | 4 | 6.1E+04 | 9.9E+04 | 1.3E+05 | 6.2E+04 | 9.8E+04 | 8.8E+04 |
| 3672 | 1 | A4UGR9 | A4UGR9 | Xin actin-bindXIRP2 | Homo sapien | 382.308 | 4 | 4 | 0.0E+00 | 2.6E+04 | 6.1E+04 | 1.1E+05 | 3.4E+03 | 5.0E+03 |
| 3673 | 1 | A6NDG6 | A6NDG6 | Glycerol-3-pPGP | Homo sapien | 34.003 | 4 | 4 | 3.6E+06 | 3.4E+06 | 2.9E+06 | 3.2E+06 | 3.3E+06 | 3.1E+06 |
| 3674 | 1 | A6NFX1 | A6NFX1 | Sphingosine-MFSD2B | Homo sapien | 53.743 | 4 | 4 | 2.8E+05 | 3.2E+05 | 2.9E+05 | 2.1E+05 | 2.5E+05 | 3.4E+05 |
| 3675 | 1 | A8K2U0 | A8K2U0 | Alpha-2-macA2ML1 | Homo sapien | 161.109 | 4 | 4 | 0.0E+00 | 0.0E+00 | 0.0E+00 | 0.0E+00 | 0.0E+00 | 0.0E+00 |
| 3676 | 1 | C9JLW8 | C9JLW8 | Mapk-regulatMCRIP1 | Homo sapien | 10.918 | 4 | 4 | 1.3E+05 | 8.2E+04 | 1.3E+05 | 0.0E+00 | 1.3E+06 | 1.3E+05 |
| 3677 | 2 | G9CGD6;Q8 | G9CGD6 | CNK3/IPCEFCNK3/IPCEF | Homo sapien | 100.369 | 4 | 4 | 1.3E+06 | 1.3E+06 | 1.3E+06 | 1.3E+06 | 1.2E+06 | 8.7E+05 |
| 3678 | 1 | H0Y944 | H0Y944 | Calpastatin (ICAST | Homo sapien | 30.565 | 4 | 4 | 1.5E+05 | 1.7E+05 | 6.7E+05 | 1.2E+05 | 1.2E+05 | 5.8E+04 |
| 3679 | 1 | O00408 | O00408 | cGMP-deperPDE2A | Homo sapien | 105.718 | 4 | 4 | 0.0E+00 | 0.0E+00 | 0.0E+00 | 0.0E+00 | 0.0E+00 | 0.0E+00 |
| 3680 | 1 | O00505 | O00505 | Importin subuKPNA3 | Homo sapien | 57.809 | 6 | 4 | 1.5E+05 | 6.6E+04 | 1.2E+05 | 2.3E+05 | 7.0E+04 | 7.7E+04 |
| 3681 | 1 | O00566 | O00566 | U3 small nuclMPHOSPH1 | Homo sapien | 78.864 | 4 | 4 | 1.9E+05 | 1.8E+05 | 2.0E+05 | 2.5E+04 | 2.8E+05 | 1.5E+05 |
| 3682 | 1 | O00743 | O00743 | Serine/threonPPP6C | Homo sapien | 35.144 | 4 | 4 | 3.6E+05 | 2.1E+05 | 2.3E+05 | 2.7E+05 | 1.0E+05 | 1.7E+05 |
| 3683 | 1 | O00754 | O00754 | Lysosomal alMAN2B1 | Homo sapien | 113.743 | 4 | 4 | 6.0E+03 | 0.0E+00 | 7.1E+03 | 0.0E+00 | 1.8E+04 | 0.0E+00 |
| 3684 | 1 | O14545 | O14545 | TRAF-type zTRAFD1 | Homo sapien | 64.844 | 4 | 4 | 4.1E+04 | 5.6E+04 | 5.6E+07 | 4.8E+04 | 2.9E+04 | 3.7E+04 |
| 3685 | 1 | O14579 | O14579 | Coatomer suCOPE | Homo sapien | 34.482 | 4 | 4 | 1.1E+06 | 1.6E+05 | 1.0E+06 | 1.1E+06 | 1.0E+06 | 1.0E+06 |
| 3686 | 1 | O14641 | O14641 | Segment poloDVL2 | Homo sapien | 78.95 | 7 | 4 | 0.0E+00 | 0.0E+00 | 0.0E+00 | 2.0E+04 | 2.7E+06 | 0.0E+00 |
| 3687 | 1 | O14683 | O14683 | Tumor proteiTP53I11 | Homo sapien | 21.052 | 4 | 4 | 1.2E+05 | 1.8E+06 | 4.0E+04 | 9.4E+04 | 9.9E+04 | 1.4E+06 |
| 3688 | 1 | O14735 | O14735 | CDP-diacylglCDIPT | Homo sapien | 23.538 | 4 | 4 | 4.3E+05 | 7.4E+05 | 5.6E+05 | 4.5E+05 | 9.3E+05 | 7.0E+05 |
| 3689 | 1 | O14772 | O14772 | Fucose-1-phFPGT | Homo sapien | 68.01 | 4 | 4 | 2.9E+03 | 5.3E+03 | 0.0E+00 | 7.0E+03 | 0.0E+00 | 1.2E+04 |
| 3690 | 1 | O14773 | O14773 | Tripeptidyl-peTPP1 | Homo sapien | 61.247 | 4 | 4 | 4.5E+05 | 5.2E+05 | 4.7E+05 | 3.5E+05 | 4.9E+05 | 3.9E+05 |
| 3691 | 1 | O15021 | O15021 | Microtubule-aMAST4 | Homo sapien | 284.102 | 4 | 4 | 0.0E+00 | 1.2E+04 | 6.5E+03 | 0.0E+00 | 1.2E+04 | 1.3E+04 |
| 3692 | 1 | O15047 | O15047 | Histone-lysinSETD1A | Homo sapien | 186.038 | 5 | 4 | 0.0E+00 | 0.0E+00 | 0.0E+00 | 1.5E+05 | 1.1E+05 | 0.0E+00 |
| 3693 | 1 | O15212 | O15212 | Prefoldin subPFDN6 | Homo sapien | 14.58 | 4 | 4 | 4.6E+04 | 8.6E+04 | 5.9E+04 | 4.6E+04 | 5.2E+04 | 3.1E+04 |
| 3694 | 1 | O15213 | O15213 | WD repeat-coWDR46 | Homo sapien | 68.07 | 4 | 4 | 0.0E+00 | 0.0E+00 | 1.2E+04 | 2.7E+04 | 9.0E+04 | 0.0E+00 |
| 3695 | 1 | O15229 | O15229 | Kynurenine 3KMO | Homo sapien | 55.808 | 4 | 4 | 0.0E+00 | 0.0E+00 | 0.0E+00 | 0.0E+00 | 5.7E+03 | 0.0E+00 |
| 3696 | 1 | O15254 | O15254 | Peroxisomal ACOX3 | Homo sapien | 77.63 | 4 | 4 | 0.0E+00 | 1.6E+05 | 5.3E+03 | 5.9E+03 | 0.0E+00 | 4.9E+03 |
| 3697 | 1 | O15260 | O15260 | Surfeit locus SURF4 | Homo sapien | 30.396 | 4 | 4 | 3.5E+06 | 4.2E+06 | 3.4E+06 | 4.0E+06 | 3.8E+06 | 3.2E+06 |
| 3698 | 1 | O15264 | O15264 | Mitogen-activMAPK13 | Homo sapien | 42.089 | 4 | 4 | 0.0E+00 | 0.0E+00 | 3.9E+03 | 0.0E+00 | 0.0E+00 | 1.8E+04 |
| 3699 | 1 | O15305 | O15305 | PhosphomanPMM2 | Homo sapien | 28.082 | 4 | 4 | 6.4E+04 | 6.8E+04 | 6.2E+05 | 6.5E+04 | 5.9E+05 | 1.9E+05 |
| 3700 | 1 | O15397 | O15397 | Importin-8 IPO8 | Homo sapien | 119.941 | 4 | 4 | 0.0E+00 | 0.0E+00 | 0.0E+00 | 0.0E+00 | 0.0E+00 | 0.0E+00 |
| 3701 | 1 | O15400 | O15400 | Syntaxin-7 STX7 | Homo sapien | 29.815 | 4 | 4 | 3.6E+05 | 6.4E+05 | 8.1E+05 | 5.8E+05 | 8.6E+05 | 8.1E+05 |
| 3702 | 1 | O15511 | O15511 | Actin-related ARPC5 | Homo sapien | 16.321 | 4 | 4 | 1.7E+06 | 2.1E+06 | 2.2E+06 | 1.9E+06 | 1.8E+06 | 1.7E+06 |
| 3703 | 1 | O15541 | O15541 | E3 ubiquitin-pRNF113A | Homo sapien | 38.787 | 4 | 4 | 2.9E+04 | 2.4E+04 | 5.1E+04 | 3.5E+04 | 2.8E+04 | 1.5E+04 |
| 3704 | 1 | O43175 | O43175 | D-3-phosphoPHGDH | Homo sapien | 56.652 | 4 | 4 | 5.6E+05 | 6.2E+05 | 4.7E+05 | 5.4E+05 | 4.7E+05 | 3.7E+05 |
| 3705 | 1 | O43181 | O43181 | NADH dehydNDUFS4 | Homo sapien | 20.108 | 4 | 4 | 1.3E+06 | 1.2E+06 | 1.6E+06 | 1.1E+06 | 1.1E+06 | 1.2E+06 |
| 3706 | 1 | O43236 | O43236 | Septin-4 SEPTIN4 | Homo sapien | 55.099 | 4 | 4 | 2.0E+07 | 0.0E+00 | 0.0E+00 | 2.6E+07 | 8.6E+05 | 1.8E+06 |
| 3707 | 1 | O43299 | O43299 | AP-5 complexAP5Z1 | Homo sapien | 88.606 | 4 | 4 | 5.2E+03 | 0.0E+00 | 0.0E+00 | 0.0E+00 | 4.5E+04 | 4.4E+04 |
| 3708 | 1 | O43402 | O43402 | ER membranEMC8 | Homo sapien | 23.771 | 4 | 4 | 5.0E+04 | 5.7E+04 | 9.6E+04 | 1.1E+05 | 1.1E+05 | 6.1E+04 |
| 3709 | 1 | O43422 | O43422 | 52 kDa repreTHAP12 | Homo sapien | 87.707 | 4 | 4 | 0.0E+00 | 7.1E+04 | 6.6E+04 | 0.0E+00 | 1.3E+05 | 6.6E+04 |

[illegible]

| | | | | | | | | | | | | | | | | |
|---|---|---|---|---|---|---|---|---|---|---|---|---|---|---|---|---|
| 4190 | 1 | Q9BPX5 | Q9BPX5 | Actin-related | ARPC5L | Homo sapien | 16.941 | 4 | 4 | 6.5E+04 | 3.6E+05 | 2.8E+05 | 4.1E+05 | 2.4E+05 | 1.8E+05 | [illegible] |
| 4191 | 1 | Q9BQG2 | Q9BQG2 | NAD-capped | NUDT12 | Homo sapien | 52.075 | 4 | 4 | 4.2E+04 | 0.0E+00 | 5.4E+04 | 0.0E+00 | 5.4E+04 | 5.7E+04 | [illegible] |
| 4192 | 1 | Q9BR61 | Q9BR61 | Acyl-CoA-bin | ACBD6 | Homo sapien | 31.15 | 4 | 4 | 1.3E+05 | 1.0E+05 | 8.6E+04 | 1.0E+05 | 1.2E+05 | 9.1E+04 | [illegible] |
| 4193 | 1 | Q9BRA2 | Q9BRA2 | Thioredoxin d | TXNDC17 | Homo sapien | 13.941 | 4 | 4 | 5.2E+06 | 7.8E+06 | 8.0E+06 | 6.0E+06 | 5.8E+06 | 4.5E+06 | [illegible] |
| 4194 | 1 | Q9BRJ7 | Q9BRJ7 | Tudor-interac | NUDT16L1 | Homo sapien | 23.339 | 5 | 4 | 6.6E+03 | 0.0E+00 | 0.0E+00 | 1.0E+04 | 0.0E+00 | 5.3E+03 | [illegible] |
| 4195 | 1 | Q9BSD7 | Q9BSD7 | Cancer-relate | NTPCR | Homo sapien | 20.712 | 4 | 4 | 1.4E+04 | 1.7E+04 | 1.4E+04 | 2.0E+05 | 2.1E+05 | 9.1E+03 | [illegible] |
| 4196 | 1 | Q9BT25 | Q9BT25 | HAUS augmi | HAUS8 | Homo sapien | 44.857 | 4 | 4 | 4.0E+06 | 2.5E+06 | 2.5E+06 | 2.6E+06 | 2.4E+06 | 2.4E+06 | [illegible] |
| 4197 | 1 | Q9BTY2 | Q9BTY2 | Plasma alpha | FUCA2 | Homo sapien | 54.066 | 4 | 4 | 1.8E+05 | 7.7E+04 | 0.0E+00 | 0.0E+00 | 1.2E+05 | 0.0E+00 | [illegible] |
| 4198 | 1 | Q9BU23 | Q9BU23 | Lipase matur | LMF2 | Homo sapien | 79.698 | 4 | 4 | 1.5E+05 | 1.5E+05 | 1.1E+05 | 1.2E+05 | 1.4E+05 | 1.1E+05 | [illegible] |
| 4199 | 1 | Q9BUB7 | Q9BUB7 | Transmembr | TMEM70 | Homo sapien | 28.972 | 4 | 4 | 0.0E+00 | 1.2E+06 | 1.3E+05 | 1.1E+05 | 1.3E+05 | 2.5E+04 | [illegible] |
| 4200 | 1 | Q9BUL5 | Q9BUL5 | PHD finger pr | PHF23 | Homo sapien | 43.817 | 4 | 4 | 2.6E+06 | 2.0E+06 | 2.4E+06 | 2.3E+06 | 3.3E+06 | 2.2E+06 | [illegible] |
| 4201 | 1 | Q9BV20 | Q9BV20 | Methylthiorib | MRI1 | Homo sapien | 39.147 | 4 | 4 | 3.5E+05 | 4.1E+05 | 2.9E+05 | 4.2E+05 | 7.1E+05 | 3.6E+05 | [illegible] |
| 4202 | 1 | Q9BVI4 | Q9BVI4 | Nucleolar con | NOC4L | Homo sapien | 58.47 | 4 | 4 | 1.3E+04 | 1.3E+04 | 1.3E+04 | 0.0E+00 | 2.5E+04 | 0.0E+00 | [illegible] |
| 4203 | 1 | Q9BW71 | Q9BW71 | HIRA-interac | HIRIP3 | Homo sapien | 61.956 | 4 | 4 | 3.5E+05 | 5.1E+05 | 5.1E+05 | 9.3E+05 | 7.8E+05 | 5.9E+05 | [illegible] |
| 4204 | 1 | Q9BWD1 | Q9BWD1 | Acetyl-CoA a | ACAT2 | Homo sapien | 41.351 | 4 | 4 | 6.5E+05 | 7.2E+05 | 5.0E+05 | 1.3E+06 | 1.3E+06 | 6.6E+05 | [illegible] |
| 4205 | 1 | Q9BXJ4 | Q9BXJ4 | Complement | C1QTNF3 | Homo sapien | 26.993 | 4 | 4 | 1.7E+04 | 5.8E+04 | 3.9E+04 | 5.4E+04 | 3.1E+04 | 2.8E+04 | [illegible] |
| 4206 | 1 | Q9BXK5 | Q9BXK5 | Bcl-2-like pro | BCL2L13 | Homo sapien | 52.724 | 4 | 4 | 1.2E+05 | 1.4E+05 | 1.3E+05 | 8.4E+04 | 1.9E+05 | 1.5E+05 | [illegible] |
| 4207 | 1 | Q9BXL7 | Q9BXL7 | Caspase rec | CARD11 | Homo sapien | 133.286 | 5 | 4 | 0.0E+00 | 0.0E+00 | 1.4E+05 | 1.2E+05 | 1.1E+05 | 0.0E+00 | [illegible] |
| 4208 | 1 | Q9BXP2 | Q9BXP2 | Solute carrier | SLC12A9 | Homo sapien | 96.111 | 4 | 4 | 3.4E+04 | 6.6E+04 | 6.6E+04 | 2.9E+04 | 2.9E+04 | 4.5E+04 | [illegible] |
| 4209 | 1 | Q9BXR6 | Q9BXR6 | Complement | CFHR5 | Homo sapien | 64.42 | 4 | 4 | 1.8E+07 | 1.6E+07 | 2.0E+07 | 2.2E+07 | 2.4E+07 | 2.4E+07 | [illegible] |
| 4210 | 1 | Q9BXX0 | Q9BXX0 | EMILIN-2 | EMILIN2 | Homo sapien | 115.686 | 4 | 4 | 1.2E+04 | 5.0E+04 | 2.2E+04 | 0.0E+00 | 0.0E+00 | 0.0E+00 | [illegible] |
| 4211 | 1 | Q9BYD2 | Q9BYD2 | Large ribosor | MRPL9 | Homo sapien | 30.242 | 4 | 4 | 6.3E+04 | 7.1E+04 | 1.4E+05 | 5.8E+04 | 8.3E+04 | 6.4E+04 | [illegible] |
| 4212 | 1 | Q9BYD6 | Q9BYD6 | Large ribosor | MRPL1 | Homo sapien | 36.909 | 4 | 4 | 1.7E+05 | 1.7E+05 | 2.0E+05 | 1.1E+05 | 2.4E+05 | 1.6E+05 | [illegible] |
| 4213 | 1 | Q9BYE9 | Q9BYE9 | Cadherin-rela | CDHR2 | Homo sapien | 141.544 | 4 | 4 | 0.0E+00 | 0.0E+00 | 0.0E+00 | 0.0E+00 | 0.0E+00 | 0.0E+00 | [illegible] |
| 4214 | 1 | Q9BYK8 | Q9BYK8 | 3'-5' exoribon | HELZ2 | Homo sapien | 322.32 | 4 | 4 | 0.0E+00 | 0.0E+00 | 5.7E+03 | 2.1E+03 | 0.0E+00 | 0.0E+00 | [illegible] |
| 4215 | 1 | Q9BYN8 | Q9BYN8 | Small ribosor | MRPS26 | Homo sapien | 24.212 | 4 | 4 | 6.8E+04 | 1.3E+04 | 8.7E+04 | 1.3E+04 | 1.1E+05 | 6.1E+04 | [illegible] |
| 4216 | 1 | Q9BZK7 | Q9BZK7 | F-box-like/W | TBL1XR1 | Homo sapien | 55.597 | 4 | 4 | 2.4E+04 | 1.2E+05 | 8.5E+04 | 6.1E+04 | 9.2E+04 | 0.0E+00 | [illegible] |
| 4217 | 1 | Q9C0B0 | Q9C0B0 | RING finger p | UNK | Homo sapien | 88.087 | 4 | 4 | 4.8E+04 | 0.0E+00 | 1.1E+04 | 2.3E+04 | 2.2E+04 | 5.1E+04 | [illegible] |
| 4218 | 1 | Q9C0D3 | Q9C0D3 | Protein zyg-1 | ZYG11B | Homo sapien | 83.923 | 4 | 4 | 0.0E+00 | 0.0E+00 | 0.0E+00 | 2.7E+04 | 0.0E+00 | 0.0E+00 | [illegible] |
| 4219 | 1 | Q9C0F1 | Q9C0F1 | Centrosomal | CEP44 | Homo sapien | 44.14 | 4 | 4 | 1.0E+06 | 9.6E+05 | 6.1E+05 | 9.4E+05 | 7.3E+05 | 9.0E+05 | [illegible] |
| 4220 | 1 | Q9C0H2 | Q9C0H2 | Protein tweet | TTYH3 | Homo sapien | 57.544 | 4 | 4 | 1.9E+05 | 0.0E+00 | 1.5E+05 | 2.0E+05 | 1.8E+05 | 1.7E+05 | [illegible] |
| 4221 | 1 | Q9GZY8 | Q9GZY8 | Mitochondrial | MFF | Homo sapien | 38.466 | 4 | 4 | 3.0E+06 | 4.3E+06 | 4.3E+06 | 2.7E+06 | 1.8E+06 | 2.8E+06 | [illegible] |
| 4222 | 1 | Q9H0B6 | Q9H0B6 | Kinesin light c | KLC2 | Homo sapien | 68.935 | 4 | 4 | 2.9E+04 | 2.2E+04 | 2.1E+04 | 0.0E+00 | 0.0E+00 | 2.8E+04 | [illegible] |
| 4223 | 1 | Q9H0M0 | Q9H0M0 | NEDD4-like E | WWP1 | Homo sapien | 105.203 | 4 | 4 | 5.7E+04 | 0.0E+00 | 0.0E+00 | 0.0E+00 | 3.3E+04 | 0.0E+00 | [illegible] |
| 4224 | 1 | Q9H0R4 | Q9H0R4 | Haloacid deh | HDHD2 | Homo sapien | 28.536 | 4 | 4 | 3.5E+05 | 2.9E+05 | 3.2E+05 | 3.7E+05 | 3.4E+05 | 2.7E+05 | [illegible] |
| 4225 | 1 | Q9H0W9 | Q9H0W9 | Ester hydrola | C11orf54 | Homo sapien | 35.114 | 4 | 4 | 1.8E+04 | 3.9E+04 | 3.2E+04 | 7.1E+05 | 3.3E+04 | 5.0E+04 | [illegible] |
| 4226 | 1 | Q9H0X9 | Q9H0X9 | Oxysterol-bin | OSBPL5 | Homo sapien | 98.619 | 4 | 4 | 0.0E+00 | 1.4E+04 | 0.0E+00 | 7.0E+03 | 1.0E+05 | 0.0E+00 | [illegible] |
| 4227 | 1 | Q9H1B5 | Q9H1B5 | Xylosyltransf | XYLT2 | Homo sapien | 96.769 | 4 | 4 | 4.8E+04 | 7.5E+04 | 2.2E+04 | 7.0E+04 | 2.9E+04 | 2.5E+04 | [illegible] |
| 4228 | 1 | Q9H300 | Q9H300 | Presenilin-as | PARL | Homo sapien | 42.191 | 4 | 4 | 0.0E+00 | 0.0E+00 | 8.3E+04 | 0.0E+00 | 5.5E+05 | 5.2E+05 | [illegible] |
| 4229 | 1 | Q9H330 | Q9H330 | Transmembr | TMEM245 | Homo sapien | 97.356 | 4 | 4 | 1.3E+05 | 1.8E+05 | 2.1E+05 | 2.1E+05 | 1.9E+05 | 2.2E+05 | [illegible] |
| 4230 | 1 | Q9H336 | Q9H336 | Cysteine-rich | CRISPLD1 | Homo sapien | 56.89 | 4 | 4 | 2.3E+05 | 3.1E+05 | 2.8E+05 | 3.8E+05 | 2.1E+05 | 2.2E+05 | [illegible] |
| 4231 | 1 | Q9H3M7 | Q9H3M7 | Thioredoxin-i | TXNIP | Homo sapien | 43.662 | 4 | 4 | 1.5E+05 | 1.5E+05 | 1.1E+05 | 1.3E+05 | 1.3E+05 | 5.6E+04 | [illegible] |
| 4232 | 1 | Q9H490 | Q9H490 | Phosphatidyli | PIGU | Homo sapien | 50.051 | 4 | 4 | 0.0E+00 | 0.0E+00 | 4.0E+05 | 2.1E+05 | 2.7E+05 | 2.6E+05 | [illegible] |
| [illegible] | | | | | | | | | | | | | | | | |

| | | | | | | | | | | | | | | | | | |
|---|---|---|---|---|---|---|---|---|---|---|---|---|---|---|---|---|---|
| 4714 | 1 Q14202 | Q14202 | Zinc finger MZMYM3 | Homo sapien | 152.381 | 3 | 3 | 0.0E+00 | 2.3E+03 | 5.7E+03 | 0.0E+00 | 1.1E+05 | 9.0E+04 | 0.0E+00 | 8.1E+03 | 9.1E+03 | 0.0E+00 |
| 4715 | 1 Q14314 | Q14314 | Fibroleukin FGL2 | Homo sapien | 50.229 | 3 | 3 | 7.6E+05 | 1.0E+06 | 8.6E+05 | 5.0E+05 | 4.4E+05 | 6.9E+05 | 9.9E+05 | 8.3E+05 | 5.0E+05 | 8.6E+05 |
| 4716 | 1 Q14315 | Q14315 | Filamin-C FLNC | Homo sapien | 291.025 | 3 | 3 | 0.0E+00 | 9.9E+05 | 0.0E+00 | 9.2E+05 | 7.2E+05 | 9.0E+05 | 1.2E+04 | 0.0E+00 | 0.0E+00 | 0.0E+00 |
| 4717 | 1 Q14318 | Q14318 | Peptidyl-prolyFKBP8 | Homo sapien | 44.561 | 3 | 3 | 1.2E+04 | 1.4E+04 | 2.0E+04 | 1.2E+04 | 2.8E+04 | 1.7E+04 | 1.8E+04 | 3.3E+04 | 2.4E+04 | 3.9E+04 |
| 4718 | 1 Q14376 | Q14376 | UDP-glucoseGALE | Homo sapien | 38.28 | 3 | 3 | 2.3E+05 | 8.4E+05 | 7.1E+05 | 9.3E+05 | 2.2E+05 | 6.4E+05 | 8.5E+05 | 1.2E+06 | 9.8E+05 | 4.6E+05 |
| 4719 | 1 Q14517 | Q14517 | ProtocadheriFAT1 | Homo sapien | 506.28 | 3 | 3 | 0.0E+00 | 0.0E+00 | 0.0E+00 | 0.0E+00 | 0.0E+00 | 0.0E+00 | 0.0E+00 | 0.0E+00 | 0.0E+00 | 0.0E+00 |
| 4720 | 1 Q14761 | Q14761 | Protein tyrosiPTPRCAP | Homo sapien | 21.194 | 3 | 3 | 1.1E+04 | 5.5E+03 | 2.2E+04 | 0.0E+00 | 4.1E+04 | 2.1E+04 | 1.9E+04 | 3.8E+04 | 0.0E+00 | 1.7E+04 |
| 4721 | 1 Q14773 | Q14773 | Intercellular aICAM4 | Homo sapien | 29.265 | 3 | 3 | 1.8E+07 | 1.2E+07 | 1.3E+07 | 2.3E+07 | 2.6E+07 | 2.6E+07 | 2.2E+07 | 2.4E+07 | 2.3E+07 | 3.5E+07 |
| 4722 | 1 Q14814 | Q14814 | Myocyte-speMEF2D | Homo sapien | 55.938 | 3 | 3 | 9.7E+04 | 7.6E+04 | 4.4E+04 | 3.8E+04 | 1.1E+05 | 4.3E+04 | 3.7E+04 | 3.6E+04 | 3.0E+04 | 4.6E+04 |
| 4723 | 1 Q14849 | Q14849 | StAR-related STARD3 | Homo sapien | 50.501 | 3 | 3 | 5.9E+04 | 4.5E+04 | 0.0E+00 | 0.0E+00 | 7.1E+04 | 7.0E+04 | 8.6E+04 | 4.9E+04 | 6.1E+04 | 4.0E+04 |
| 4724 | 1 Q14938 | Q14938 | Nuclear factoNFIX | Homo sapien | 55.097 | 3 | 3 | 2.2E+06 | 1.4E+06 | 1.9E+06 | 1.7E+06 | 1.8E+06 | 1.8E+06 | 1.5E+06 | 1.4E+06 | 1.8E+06 | 1.7E+06 |
| 4725 | 1 Q15040 | Q15040 | Josephin-1 JOSD1 | Homo sapien | 23.199 | 3 | 3 | 2.1E+05 | 1.6E+05 | 1.6E+05 | 2.4E+05 | 1.8E+05 | 1.9E+05 | 3.2E+04 | 9.4E+04 | 1.2E+05 | 2.7E+04 |
| 4726 | 1 Q15102 | Q15102 | Platelet-activPAFAH1B3 | Homo sapien | 25.734 | 3 | 3 | 3.5E+06 | 2.6E+06 | 2.6E+06 | 3.2E+06 | 3.2E+06 | 2.4E+06 | 6.3E+06 | 4.6E+06 | 3.9E+06 | 6.2E+06 |
| 4727 | 1 Q15113 | Q15113 | Procollagen cPCOLCE | Homo sapien | 47.972 | 3 | 3 | 7.0E+05 | 4.7E+05 | 1.7E+05 | 1.9E+05 | 6.5E+05 | 6.8E+05 | 3.9E+05 | 6.0E+05 | 3.8E+05 | 4.9E+05 |
| 4728 | 1 Q15121 | Q15121 | Astrocytic phPEA15 | Homo sapien | 15.037 | 3 | 3 | 3.0E+05 | 4.3E+05 | 4.4E+05 | 2.8E+05 | 2.2E+05 | 2.1E+05 | 8.5E+05 | 6.2E+05 | 5.2E+05 | 6.9E+05 |
| 4729 | 1 Q15326 | Q15326 | Zinc finger MZMYND11 | Homo sapien | 70.963 | 3 | 3 | 0.0E+00 | 1.1E+05 | 1.4E+04 | 0.0E+00 | 9.6E+04 | 0.0E+00 | 0.0E+00 | 7.7E+03 | 0.0E+00 | 8.1E+03 |
| 4730 | 1 Q15366 | Q15366 | Poly(rC)-bindPCBP2 | Homo sapien | 38.579 | 4 | 3 | 5.1E+06 | 5.1E+06 | 6.4E+06 | 1.0E+07 | 7.9E+06 | 7.9E+06 | 7.5E+06 | 5.2E+06 | 3.7E+06 | 6.4E+06 |
| 4731 | 1 Q15369 | Q15369 | Elongin-C ELOC | Homo sapien | 12.473 | 3 | 3 | 2.0E+06 | 2.0E+06 | 2.4E+06 | 2.6E+06 | 2.6E+06 | 2.6E+06 | 3.8E+06 | 2.6E+06 | 2.3E+06 | 3.6E+06 |
| 4732 | 1 Q15417 | Q15417 | Calponin-3 CNN3 | Homo sapien | 36.414 | 4 | 3 | 9.6E+05 | 1.1E+06 | 1.3E+06 | 2.2E+06 | 1.7E+06 | 1.6E+06 | 1.6E+06 | 0.0E+00 | 1.0E+06 | 1.7E+06 |
| 4733 | 1 Q15526 | Q15526 | Surfeit locus SURF1 | Homo sapien | 33.33 | 3 | 3 | 9.5E+04 | 1.4E+05 | 5.8E+05 | 8.6E+04 | 1.4E+05 | 5.0E+04 | 7.1E+05 | 1.5E+05 | 6.3E+05 | 1.9E+05 |
| 4734 | 1 Q15528 | Q15528 | Mediator of RMED22 | Homo sapien | 22.221 | 3 | 3 | 0.0E+00 | 0.0E+00 | 0.0E+00 | 0.0E+00 | 1.3E+04 | 0.0E+00 | 0.0E+00 | 0.0E+00 | 0.0E+00 | 0.0E+00 |
| 4735 | 1 Q15545 | Q15545 | TranscriptionTAF7 | Homo sapien | 40.258 | 3 | 3 | 1.1E+04 | 5.9E+04 | 1.2E+04 | 1.4E+04 | 4.4E+04 | 3.5E+04 | 0.0E+00 | 0.0E+00 | 1.6E+04 | 0.0E+00 |
| 4736 | 1 Q15645 | Q15645 | Pachytene cl'TRIP13 | Homo sapien | 48.553 | 3 | 3 | 9.6E+03 | 1.2E+04 | 2.3E+04 | 8.9E+03 | 1.2E+04 | 0.0E+00 | 1.4E+04 | 0.0E+00 | 1.5E+04 | 1.6E+04 |
| 4737 | 1 Q15652 | Q15652 | Probable JmjJMJD1C | Homo sapien | 284.529 | 3 | 3 | 0.0E+00 | 1.6E+05 | 2.5E+04 | 0.0E+00 | 0.0E+00 | 0.0E+00 | 0.0E+00 | 0.0E+00 | 0.0E+00 | 0.0E+00 |
| 4738 | 1 Q15654 | Q15654 | Thyroid recepTRIP6 | Homo sapien | 50.286 | 3 | 3 | 0.0E+00 | 0.0E+00 | 4.2E+03 | 5.9E+04 | 0.0E+00 | 4.5E+04 | 7.7E+05 | 1.0E+06 | 8.6E+05 | 9.4E+05 |
| 4739 | 1 Q15738 | Q15738 | Sterol-4-alphNSDHL | Homo sapien | 41.9 | 3 | 3 | 4.7E+05 | 6.2E+05 | 0.0E+00 | 4.9E+04 | 6.4E+05 | 0.0E+00 | 0.0E+00 | 7.1E+05 | 0.0E+00 | 5.1E+05 |
| 4740 | 1 Q15771 | Q15771 | Ras-related pRAB30 | Homo sapien | 23.057 | 3 | 3 | 1.5E+05 | 1.3E+05 | 2.1E+05 | 4.8E+04 | 1.2E+05 | 7.6E+04 | 2.1E+05 | 0.0E+00 | 2.1E+05 | 2.6E+05 |
| 4741 | 1 Q15796 | Q15796 | Mothers agaiSMAD2 | Homo sapien | 52.307 | 3 | 3 | 7.7E+06 | 9.4E+06 | 6.7E+06 | 1.1E+07 | 7.4E+06 | 8.3E+06 | 1.0E+07 | 6.7E+06 | 7.2E+06 | 8.4E+06 |
| 4742 | 1 Q15814 | Q15814 | Tubulin-speciTBCC | Homo sapien | 39.248 | 3 | 3 | 4.6E+05 | 4.3E+05 | 2.0E+05 | 3.5E+05 | 4.4E+05 | 5.5E+05 | 4.6E+05 | 3.6E+05 | 3.7E+05 | 2.1E+05 |
| 4743 | 1 Q15819 | Q15819 | Ubiquitin-conUBE2V2 | Homo sapien | 16.363 | 6 | 3 | 5.0E+06 | 6.1E+06 | 4.8E+06 | 5.5E+06 | 5.9E+06 | 5.3E+06 | 5.8E+06 | 6.4E+06 | 4.5E+06 | 6.4E+06 |
| 4744 | 1 Q15853 | Q15853 | Upstream stirUSF2 | Homo sapien | 36.955 | 4 | 3 | 0.0E+00 | 0.0E+00 | 0.0E+00 | 0.0E+00 | 1.3E+04 | 0.0E+00 | 0.0E+00 | 0.0E+00 | 1.4E+04 | 0.0E+00 |
| 4745 | 1 Q16637 | Q16637 | Survival motoSMN1 | Homo sapien | 31.847 | 3 | 3 | 7.6E+04 | 7.2E+04 | 8.4E+04 | 5.6E+04 | 9.1E+04 | 7.5E+04 | 1.8E+05 | 9.7E+04 | 3.7E+05 | 1.2E+05 |
| 4746 | 1 Q16656 | Q16656 | Nuclear respiNRF1 | Homo sapien | 53.54 | 3 | 3 | 6.5E+04 | 2.3E+05 | 3.0E+04 | 2.0E+04 | 2.6E+05 | 1.6E+05 | 2.9E+04 | 2.3E+05 | 1.9E+05 | 2.3E+05 |
| 4747 | 1 Q16760 | Q16760 | DiacylglycerolDGKD | Homo sapien | 134.527 | 3 | 3 | 3.3E+04 | 4.0E+04 | 1.3E+05 | 1.6E+04 | 1.3E+04 | 1.3E+04 | 0.0E+00 | 2.1E+04 | 2.4E+04 | 2.4E+04 |
| 4748 | 1 Q16831 | Q16831 | Uridine phosUPP1 | Homo sapien | 33.933 | 3 | 3 | 0.0E+00 | 0.0E+00 | 0.0E+00 | 0.0E+00 | 0.0E+00 | 0.0E+00 | 3.2E+04 | 0.0E+00 | 3.1E+04 | 0.0E+00 |
| 4749 | 1 Q16875 | Q16875 | 6-phosphofruPFKFB3 | Homo sapien | 59.609 | 3 | 3 | 3.4E+05 | 0.0E+00 | 2.0E+04 | 3.2E+04 | 0.0E+00 | 2.8E+04 | 5.1E+04 | 4.3E+04 | 6.2E+04 | 6.2E+04 |
| 4750 | 1 Q16877 | Q16877 | 6-phosphofruPFKFB4 | Homo sapien | 54.039 | 4 | 3 | 0.0E+00 | 1.2E+04 | 1.8E+04 | 0.0E+00 | 0.0E+00 | 0.0E+00 | 1.4E+04 | 1.7E+04 | 1.4E+04 | 0.0E+00 |
| 4751 | 1 Q32NB8 | Q32NB8 | CDP-diacylglPGS1 | Homo sapien | 62.733 | 3 | 3 | 0.0E+00 | 0.0E+00 | 3.0E+04 | 0.0E+00 | 0.0E+00 | 0.0E+00 | 0.0E+00 | 3.7E+04 | 4.5E+04 | 5.2E+04 |
| 4752 | 1 Q3ZCW2 | Q3ZCW2 | Galectin-relatLGALSL | Homo sapien | 18.987 | 3 | 3 | 6.8E+05 | 7.4E+05 | 6.8E+05 | 8.1E+05 | 6.4E+05 | 6.0E+05 | 1.0E+06 | 1.2E+06 | 1.7E+06 | 1.7E+06 |
| 4753 | 1 Q4AE62 | Q4AE62 | tRNA-queuosQTDC1 | Homo sapien | 52.598 | 3 | 3 | 1.3E+05 | 1.5E+05 | 1.4E+05 | 1.8E+05 | 1.7E+05 | 2.0E+05 | 1.1E+05 | 2.5E+04 | 4.0E+04 | 1.1E+05 |
| 4754 | 1 Q4LE39 | Q4LE39 | AT-rich interaARID4B | Homo sapien | 147.812 | 3 | 3 | 0.0E+00 | 0.0E+00 | 8.8E+04 | 6.1E+04 | 7.8E+04 | 7.0E+04 | 0.0E+00 | 7.6E+04 | 0.0E+00 | 8.9E+04 |
| 4755 | 1 Q53EU6 | Q53EU6 | Glycerol-3-phGPAT3 | Homo sapien | 48.707 | 3 | 3 | 0.0E+00 | 0.0E+00 | 2.0E+04 | 0.0E+00 | 0.0E+00 | 0.0E+00 | 3.8E+04 | 2.2E+04 | 0.0E+00 | 1.6E+04 |
| 4756 | 1 Q53HC9 | Q53HC9 | EARP and GAEIPR1 | Homo sapien | 43.602 | 3 | 3 | 6.8E+04 | 0.0E+00 | 9.3E+04 | 3.9E+05 | 6.9E+04 | 4.7E+04 | 7.9E+04 | 8.9E+05 | 6.3E+04 | 1.2E+05 |
| 4757 | 1 Q56VL3 | Q56VL3 | OCIA domainOCIAD2 | Homo sapien | 16.953 | 3 | 3 | 0.0E+00 | 0.0E+00 | 9.5E+03 | 7.6E+03 | 1.0E+04 | 0.0E+00 | 0.0E+00 | 2.9E+04 | 0.0E+00 | 1.6E+04 |
| 4758 | 1 Q5HYI8 | Q5HYI8 | Rab-like protRABL3 | Homo sapien | 26.423 | 3 | 3 | 9.2E+05 | 1.2E+06 | 3.4E+05 | 3.4E+04 | 1.2E+06 | 1.2E+06 | 1.1E+06 | 9.5E+05 | 1.2E+06 | 1.1E+06 |
| 4759 | 1 Q5J8M3 | Q5J8M3 | ER membranEMC4 | Homo sapien | 20.086 | 3 | 3 | 1.0E+05 | 0.0E+00 | 0.0E+00 | 0.0E+00 | 0.0E+00 | 0.0E+00 | 3.7E+06 | 3.3E+06 | 3.8E+06 | 2.8E+06 |
| 4760 | 1 Q5JTD0 | Q5JTD0 | Tight junctionTJAP1 | Homo sapien | 61.821 | 3 | 3 | 1.3E+05 | 1.5E+05 | 8.7E+04 | 1.1E+05 | 1.7E+05 | 1.3E+05 | 8.0E+04 | 5.1E+04 | 2.6E+04 | 5.8E+04 |
| 4761 | 1 Q5JU69 | Q5JU69 | Torsin-2A TOR2A | Homo sapien | 35.711 | 3 | 3 | 4.3E+04 | 4.9E+04 | 2.7E+04 | 4.4E+04 | 4.2E+04 | 3.8E+04 | 4.2E+04 | 6.7E+04 | 4.2E+04 | 4.8E+04 |
| 4762 | 1 Q5M8T2 | Q5M8T2 | Solute carrierSLC35D3 | Homo sapien | 44.184 | 3 | 3 | 7.6E+04 | 9.7E+04 | 9.5E+04 | 8.5E+04 | 6.7E+04 | 8.9E+04 | 1.1E+05 | 6.7E+04 | 9.6E+04 | 1.4E+05 |
| 4763 | 1 Q5SQ64 | Q5SQ64 | Lymphocyte aLY6G6F | Homo sapien | 32.464 | 3 | 3 | 9.3E+05 | 9.3E+05 | 7.9E+05 | 8.7E+05 | 8.0E+05 | 8.2E+05 | 5.7E+05 | 8.2E+05 | 7.9E+05 | 1.1E+06 |
| 4764 | 1 Q5ST30 | Q5ST30 | Valine--tRNA VARS2 | Homo sapien | 118.49 | 3 | 3 | 0.0E+00 | 0.0E+00 | 0.0E+00 | 0.0E+00 | 0.0E+00 | 0.0E+00 | 0.0E+00 | 0.0E+00 | 0.0E+00 | 0.0E+00 |
| 4765 | 1 Q5SWX8 | Q5SWX8 | Protein odr-4ODR4 | Homo sapien | 51.103 | 3 | 3 | 0.0E+00 | 3.4E+04 | 3.5E+04 | 0.0E+00 | 0.0E+00 | 0.0E+00 | 4.3E+04 | 0.0E+00 | 0.0E+00 | 0.0E+00 |
| 4766 | 1 Q5SY16 | Q5SY16 | PolynucleotidNOL9 | Homo sapien | 79.324 | 3 | 3 | 7.0E+03 | 0.0E+00 | 0.0E+00 | 2.3E+04 | 1.3E+05 | 7.8E+03 | 0.0E+00 | 0.0E+00 | 0.0E+00 | 0.0E+00 |
| 4767 | 1 Q5T280 | Q5T280 | Putative methSPOUT1 | Homo sapien | 42.009 | 3 | 3 | 1.9E+04 | 2.5E+04 | 1.6E+04 | 2.8E+04 | 3.0E+04 | 0.0E+00 | 0.0E+00 | 0.0E+00 | 0.0E+00 | 0.0E+00 |
| 4768 | 1 Q5TFE4 | Q5TFE4 | 5'-nucleotidasNT5DC1 | Homo sapien | 51.845 | 3 | 3 | 5.8E+05 | 8.1E+05 | 1.5E+04 | 6.7E+05 | 5.8E+05 | 5.5E+05 | 1.3E+06 | 1.6E+06 | 1.0E+06 | 1.4E+06 |
| 4769 | 1 Q5VTQ0 | Q5VTQ0 | TetratricopepTTC39B | Homo sapien | 76.959 | 3 | 3 | 0.0E+00 | 7.5E+05 | 0.0E+00 | 0.0E+00 | 1.2E+04 | 0.0E+00 | 0.0E+00 | 0.0E+00 | 0.0E+00 | 0.0E+00 |
| 4770 | 1 Q5VU43 | Q5VU43 | Myomegalin PDE4DIP | Homo sapien | 265.106 | 3 | 3 | 0.0E+00 | 1.4E+07 | 1.8E+07 | 0.0E+00 | 2.4E+07 | 0.0E+00 | 0.0E+00 | 0.0E+00 | 1.8E+07 | 0.0E+00 |
| 4771 | 1 Q5VWC8 | Q5VWC8 | Very-long-chaHACD4 | Homo sapien | 27.518 | 3 | 3 | 7.0E+05 | 6.1E+05 | 7.7E+05 | 6.2E+05 | 6.5E+05 | 5.1E+05 | 4.3E+04 | 5.2E+05 | 2.8E+06 | 4.7E+04 |
| 4772 | 1 Q5VWG9 | Q5VWG9 | TranscriptionTAF3 | Homo sapien | 103.583 | 3 | 3 | 0.0E+00 | 0.0E+00 | 0.0E+00 | 0.0E+00 | 0.0E+00 | 0.0E+00 | 0.0E+00 | 0.0E+00 | 0.0E+00 | 0.0E+00 |

[illegible]

| | | | | | | | | | | | | | | | | | | | | | |
|---|---|---|---|---|---|---|---|---|---|---|---|---|---|---|---|---|---|---|---|---|---|
| 5238 | 1 | O14526 | O14526 | F-BAR domaiFCHO1 | Homo sapien | 96.863 | 2 | 2 | 0.0E+00 | 2.0E+04 | 0.0E+00 | 0.0E+00 | 2.1E+04 | 1.3E+04 | 0.0E+00 | 0.0E+00 | 0.0E+00 | 0.0E+00 | 0.0E+00 | 0.0E+00 | [illegible] |
| 5239 | 1 | O14530 | O14530 | Thioredoxin cTXNDC9 | Homo sapien | 26.533 | 2 | 2 | 2.2E+05 | 1.4E+05 | 1.2E+05 | 2.0E+05 | 2.0E+05 | 1.9E+05 | 1.5E+05 | 1.8E+05 | 1.3E+05 | 1.5E+05 | 1.6E+05 | 1.8E+05 | [illegible] |
| 5240 | 1 | O14618 | O14618 | Copper chapCCS | Homo sapien | 29.037 | 2 | 2 | 8.3E+06 | 7.5E+06 | 6.4E+06 | 6.9E+06 | 7.6E+06 | 7.5E+06 | 1.5E+07 | 1.8E+07 | 1.4E+07 | 1.9E+07 | 2.0E+07 | 1.9E+07 | [illegible] |
| 5241 | 1 | O14653 | O14653 | Golgi SNAP rGOSR2 | Homo sapien | 24.774 | 2 | 2 | 2.3E+03 | 0.0E+00 | 9.2E+03 | 0.0E+00 | 6.4E+03 | 6.8E+03 | 0.0E+00 | 0.0E+00 | 5.5E+03 | 4.4E+03 | 4.9E+03 | 4.3E+03 | [illegible] |
| 5242 | 1 | O14657 | O14657 | Torsin-1B TOR1B | Homo sapien | 37.979 | 2 | 2 | 0.0E+00 | 0.0E+00 | 0.0E+00 | 0.0E+00 | 7.0E+05 | 0.0E+00 | 0.0E+00 | 0.0E+00 | 0.0E+00 | 0.0E+00 | 0.0E+00 | 0.0E+00 | [illegible] |
| 5243 | 1 | O14686 | O14686 | Histone-lysinKMT2D | Homo sapien | 593.401 | 2 | 2 | 0.0E+00 | 0.0E+00 | 0.0E+00 | 0.0E+00 | 6.6E+03 | 6.9E+03 | 0.0E+00 | 4.8E+03 | 0.0E+00 | 0.0E+00 | 1.1E+04 | 0.0E+00 | [illegible] |
| 5244 | 1 | O14713 | O14713 | Integrin beta-ITGB1BP1 | Homo sapien | 21.783 | 2 | 2 | 7.1E+04 | 9.3E+04 | 0.0E+00 | 0.0E+00 | 6.1E+04 | 8.5E+04 | 0.0E+00 | 0.0E+00 | 0.0E+00 | 0.0E+00 | 0.0E+00 | 7.7E+04 | [illegible] |
| 5245 | 1 | O14734 | O14734 | Acyl-coenzymACOT8 | Homo sapien | 35.915 | 2 | 2 | 9.0E+04 | 7.3E+04 | 1.3E+05 | 9.3E+04 | 6.4E+04 | 7.8E+04 | 1.1E+05 | 8.8E+04 | 1.0E+05 | 9.5E+04 | 1.1E+05 | 8.3E+04 | [illegible] |
| 5246 | 1 | O14802 | O14802 | DNA-directedPOLR3A | Homo sapien | 155.645 | 2 | 2 | 0.0E+00 | 0.0E+00 | 0.0E+00 | 0.0E+00 | 0.0E+00 | 0.0E+00 | 0.0E+00 | 0.0E+00 | 0.0E+00 | 0.0E+00 | 0.0E+00 | 0.0E+00 | [illegible] |
| 5247 | 1 | O14828 | O14828 | Secretory caSCAMP3 | Homo sapien | 38.286 | 2 | 2 | 5.4E+04 | 6.1E+04 | 7.0E+04 | 5.1E+04 | 4.8E+04 | 3.4E+04 | 4.7E+04 | 0.0E+00 | 4.9E+04 | 4.5E+04 | 4.3E+04 | 5.2E+04 | [illegible] |
| 5248 | 1 | O14880 | O14880 | Glutathione SMGST3 | Homo sapien | 16.516 | 2 | 2 | 2.0E+06 | 2.4E+06 | 2.7E+06 | 2.7E+06 | 2.9E+06 | 2.4E+06 | 1.4E+06 | 1.8E+06 | 5.4E+05 | 2.6E+06 | 1.5E+06 | 1.9E+06 | [illegible] |
| 5249 | 1 | O14908 | O14908 | PDZ domain-GIPC1 | Homo sapien | 36.05 | 2 | 2 | 0.0E+00 | 0.0E+00 | 0.0E+00 | 3.6E+03 | 0.0E+00 | 0.0E+00 | 0.0E+00 | 0.0E+00 | 0.0E+00 | 0.0E+00 | 0.0E+00 | 0.0E+00 | [illegible] |
| 5250 | 1 | O14933 | O14933 | Ubiquitin/ISGUBE2L6 | Homo sapien | 17.768 | 2 | 2 | 0.0E+00 | 0.0E+00 | 0.0E+00 | 0.0E+00 | 1.1E+04 | 0.0E+00 | 0.0E+00 | 1.2E+04 | 0.0E+00 | 0.0E+00 | 0.0E+00 | 0.0E+00 | [illegible] |
| 5251 | 1 | O14936 | O14936 | Peripheral plaCASK | Homo sapien | 105.126 | 2 | 2 | 0.0E+00 | 1.1E+05 | 0.0E+00 | 9.0E+04 | 0.0E+00 | 3.8E+06 | 3.7E+06 | 8.3E+04 | 9.0E+04 | 0.0E+00 | 9.3E+04 | 5.0E+06 | [illegible] |
| 5252 | 2 | O14950;P19 | O14950 | Myosin regulMYL12B | Homo sapien | 19.779 | 2 | 2 | 2.2E+08 | 1.8E+08 | 1.4E+08 | 1.5E+08 | 1.6E+08 | 1.4E+08 | 2.3E+08 | 1.7E+08 | 2.1E+08 | 2.0E+08 | 2.3E+08 | 2.8E+08 | [illegible] |
| 5253 | 1 | O14966 | O14966 | Ras-related pRAB29 | Homo sapien | 23.152 | 2 | 2 | 0.0E+00 | 4.4E+04 | 5.1E+04 | 0.0E+00 | 6.6E+04 | 0.0E+00 | 6.1E+04 | 0.0E+00 | 8.1E+04 | 2.2E+05 | 7.8E+04 | 0.0E+00 | [illegible] |
| 5254 | 1 | O14972 | O14972 | Vacuolar proVPS26C | Homo sapien | 33.01 | 2 | 2 | 0.0E+00 | 0.0E+00 | 0.0E+00 | 0.0E+00 | 0.0E+00 | 0.0E+00 | 0.0E+00 | 0.0E+00 | 1.1E+04 | 1.9E+04 | 2.7E+04 | 1.6E+04 | [illegible] |
| 5255 | 1 | O15014 | O15014 | Zinc finger prZNF609 | Homo sapien | 151.193 | 2 | 2 | 0.0E+00 | 0.0E+00 | 1.4E+04 | 0.0E+00 | 1.2E+04 | 1.1E+04 | 0.0E+00 | 1.4E+04 | 0.0E+00 | 0.0E+00 | 0.0E+00 | 0.0E+00 | [illegible] |
| 5256 | 1 | O15091 | O15091 | MitochondrialPRORP | Homo sapien | 67.316 | 2 | 2 | 0.0E+00 | 0.0E+00 | 2.5E+03 | 0.0E+00 | 4.6E+03 | 1.7E+03 | 0.0E+00 | 0.0E+00 | 1.9E+04 | 0.0E+00 | 0.0E+00 | 0.0E+00 | [illegible] |
| 5257 | 1 | O15118 | O15118 | NPC intracellNPC1 | Homo sapien | 142.169 | 2 | 2 | 0.0E+00 | 0.0E+00 | 0.0E+00 | 0.0E+00 | 5.6E+03 | 5.3E+03 | 0.0E+00 | 0.0E+00 | 0.0E+00 | 0.0E+00 | 0.0E+00 | 0.0E+00 | [illegible] |
| 5258 | 1 | O15120 | O15120 | 1-acyl-sn-glyAGPAT2 | Homo sapien | 30.916 | 2 | 2 | 1.6E+05 | 2.2E+05 | 1.6E+05 | 1.5E+05 | 1.6E+05 | 1.8E+05 | 4.0E+05 | 4.5E+04 | 4.5E+04 | 3.6E+04 | 3.4E+04 | 4.7E+04 | [illegible] |
| 5259 | 1 | O15121 | O15121 | Sphingolipid cDEGS1 | Homo sapien | 37.867 | 2 | 2 | 2.7E+05 | 3.3E+05 | 2.5E+05 | 0.0E+00 | 0.0E+00 | 0.0E+00 | 5.7E+05 | 0.0E+00 | 0.0E+00 | 5.5E+05 | 0.0E+00 | 0.0E+00 | [illegible] |
| 5260 | 1 | O15126 | O15126 | Secretory caSCAMP1 | Homo sapien | 37.921 | 2 | 2 | 1.7E+04 | 0.0E+00 | 1.5E+04 | 1.1E+04 | 1.7E+03 | 0.0E+00 | 1.1E+04 | 0.0E+00 | 0.0E+00 | 1.3E+04 | 7.3E+03 | 9.0E+03 | [illegible] |
| 5261 | 1 | O15127 | O15127 | Secretory caSCAMP2 | Homo sapien | 36.647 | 2 | 2 | 8.2E+04 | 4.3E+04 | 9.6E+04 | 8.6E+04 | 5.2E+04 | 6.5E+04 | 6.8E+04 | 8.7E+04 | 5.5E+04 | 9.6E+04 | 6.5E+04 | 9.0E+04 | [illegible] |
| 5262 | 1 | O15160 | O15160 | DNA-directedPOLR1C | Homo sapien | 39.251 | 2 | 2 | 0.0E+00 | 0.0E+00 | 0.0E+00 | 0.0E+00 | 0.0E+00 | 0.0E+00 | 0.0E+00 | 0.0E+00 | 0.0E+00 | 0.0E+00 | 0.0E+00 | 0.0E+00 | [illegible] |
| 5263 | 1 | O15204 | O15204 | ADAM DEC1 ADAMDEC1 | Homo sapien | 52.776 | 2 | 2 | 1.1E+05 | 2.1E+05 | 8.6E+04 | 1.6E+05 | 1.2E+05 | 1.0E+05 | 2.1E+05 | 1.5E+05 | 9.8E+04 | 2.1E+05 | 2.1E+05 | 1.9E+05 | [illegible] |
| 5264 | 1 | O15232 | O15232 | Matrilin-3 MATN3 | Homo sapien | 52.817 | 2 | 2 | 0.0E+00 | 0.0E+00 | 0.0E+00 | 0.0E+00 | 0.0E+00 | 0.0E+00 | 0.0E+00 | 0.0E+00 | 0.0E+00 | 0.0E+00 | 0.0E+00 | 0.0E+00 | [illegible] |
| 5265 | 1 | O15258 | O15258 | Protein RER1RER1 | Homo sapien | 22.958 | 2 | 2 | 3.3E+05 | 5.4E+05 | 5.3E+05 | 4.0E+05 | 4.8E+05 | 5.0E+05 | 5.7E+05 | 5.9E+05 | 5.8E+05 | 7.8E+05 | 6.7E+05 | 6.6E+05 | [illegible] |
| 5266 | 1 | O15321 | O15321 | TransmembrTM9SF1 | Homo sapien | 68.861 | 2 | 2 | 1.3E+04 | 1.6E+04 | 1.5E+04 | 1.4E+04 | 2.7E+04 | 0.0E+00 | 3.0E+04 | 2.2E+04 | 1.6E+04 | 2.2E+04 | 0.0E+00 | 0.0E+00 | [illegible] |
| 5267 | 1 | O15427 | O15427 | MonocarboxySLC16A3 | Homo sapien | 49.47 | 2 | 2 | 1.9E+05 | 2.3E+05 | 1.9E+05 | 2.3E+05 | 2.7E+05 | 2.3E+05 | 3.6E+05 | 3.5E+05 | 3.1E+05 | 3.9E+05 | 3.5E+05 | 3.6E+05 | [illegible] |
| 5268 | 1 | O15514 | O15514 | DNA-directedPOLR2D | Homo sapien | 16.312 | 2 | 2 | 3.1E+03 | 0.0E+00 | 3.1E+03 | 0.0E+00 | 3.9E+03 | 0.0E+00 | 0.0E+00 | 0.0E+00 | 0.0E+00 | 0.0E+00 | 0.0E+00 | 0.0E+00 | [illegible] |
| 5269 | 1 | O15530 | O15530 | 3-phosphoincPDPK1 | Homo sapien | 63.155 | 7 | 2 | 0.0E+00 | 0.0E+00 | 0.0E+00 | 0.0E+00 | 0.0E+00 | 0.0E+00 | 1.7E+05 | 0.0E+00 | 0.0E+00 | 0.0E+00 | 0.0E+00 | 1.6E+05 | [illegible] |
| 5270 | 1 | O43156 | O43156 | TELO2-interaTTI1 | Homo sapien | 122.071 | 2 | 2 | 0.0E+00 | 0.0E+00 | 0.0E+00 | 0.0E+00 | 0.0E+00 | 0.0E+00 | 1.2E+05 | 0.0E+00 | 0.0E+00 | 1.4E+05 | 0.0E+00 | 1.4E+05 | [illegible] |
| 5271 | 1 | O43169 | O43169 | Cytochrome lCYB5B | Homo sapien | 16.697 | 2 | 2 | 1.8E+05 | 2.0E+05 | 2.2E+05 | 2.6E+05 | 2.9E+05 | 2.2E+05 | 2.2E+05 | 3.4E+05 | 3.1E+05 | 4.6E+05 | 3.6E+05 | 3.2E+05 | [illegible] |
| 5272 | 1 | O43293 | O43293 | Death-associDAPK3 | Homo sapien | 52.535 | 2 | 2 | 9.2E+04 | 3.3E+04 | 7.6E+04 | 0.0E+00 | 5.9E+04 | 3.1E+04 | 1.8E+04 | 2.2E+04 | 4.3E+04 | 4.7E+04 | 1.6E+04 | 1.6E+04 | [illegible] |
| 5273 | 1 | O43303 | O43303 | Centriolar coCCP110 | Homo sapien | 113.425 | 2 | 2 | 6.1E+03 | 0.0E+00 | 0.0E+00 | 0.0E+00 | 7.8E+03 | 7.6E+03 | 0.0E+00 | 0.0E+00 | 0.0E+00 | 0.0E+00 | 0.0E+00 | 0.0E+00 | [illegible] |
| 5274 | 1 | O43464 | O43464 | Serine proteaHTRA2 | Homo sapien | 48.841 | 3 | 2 | 0.0E+00 | 0.0E+00 | 0.0E+00 | 0.0E+00 | 0.0E+00 | 0.0E+00 | 0.0E+00 | 0.0E+00 | 0.0E+00 | 0.0E+00 | 0.0E+00 | 0.0E+00 | [illegible] |
| 5275 | 1 | O43548 | O43548 | Protein-glutarTGM5 | Homo sapien | 80.779 | 2 | 2 | 0.0E+00 | 0.0E+00 | 0.0E+00 | 0.0E+00 | 0.0E+00 | 0.0E+00 | 0.0E+00 | 0.0E+00 | 0.0E+00 | 0.0E+00 | 0.0E+00 | 0.0E+00 | [illegible] |
| 5276 | 1 | O43561 | O43561 | Linker for actLAT | Homo sapien | 27.929 | 2 | 2 | 2.4E+05 | 1.3E+05 | 3.0E+05 | 1.8E+05 | 2.0E+05 | 2.1E+05 | 4.6E+05 | 2.4E+05 | 4.5E+05 | 0.0E+00 | 1.0E+06 | 4.4E+05 | [illegible] |
| 5277 | 1 | O43676 | O43676 | NADH dehydNDUFB3 | Homo sapien | 11.4 | 2 | 2 | 1.0E+06 | 1.0E+06 | 1.8E+06 | 1.0E+06 | 9.3E+05 | 7.9E+05 | 1.0E+06 | 1.2E+06 | 1.3E+06 | 1.2E+06 | 1.0E+06 | 1.2E+06 | [illegible] |
| 5278 | 1 | O43677 | O43677 | NADH dehydNDUFC1 | Homo sapien | 8.731 | 2 | 2 | 0.0E+00 | 0.0E+00 | 0.0E+00 | 0.0E+00 | 0.0E+00 | 0.0E+00 | 5.0E+03 | 6.1E+03 | 6.1E+03 | 0.0E+00 | 0.0E+00 | 7.1E+03 | [illegible] |
| 5279 | 1 | O43678 | O43678 | NADH dehydNDUFA2 | Homo sapien | 10.921 | 2 | 2 | 0.0E+00 | 0.0E+00 | 1.5E+04 | 0.0E+00 | 0.0E+00 | 0.0E+00 | 0.0E+00 | 1.6E+05 | 0.0E+00 | 0.0E+00 | 0.0E+00 | 2.4E+05 | [illegible] |
| 5280 | 1 | O43708 | O43708 | MaleylacetoaGSTZ1 | Homo sapien | 24.21 | 2 | 2 | 7.6E+04 | 9.8E+04 | 1.3E+05 | 1.2E+05 | 1.3E+05 | 7.6E+04 | 3.2E+05 | 1.8E+05 | 1.6E+05 | 3.7E+05 | 3.9E+05 | 3.0E+05 | [illegible] |
| [illegible] | | | | | | | | | | | | | | | | | | | | | |

| | | | | | | | | | | | | | | | | | | | | | | |
|---|---|---|---|---|---|---|---|---|---|---|---|---|---|---|---|---|---|---|---|---|---|---|
| 5762 | 1 | Q6ICB4 | Q6ICB4 | Sesquipedal | PHETA2 | Homo sapien | 28.339 | 2 | 2 | 4.3E+04 | 4.0E+04 | 4.8E+04 | 2.7E+04 | 1.2E+05 | 5.6E+04 | 0.0E+00 | 0.0E+00 | 5.1E+04 | 0.0E+00 | 7.5E+04 | 0.0E+00 | [illegible] |
| 5763 | 1 | Q6IQ23 | Q6IQ23 | Pleckstrin ho | PLEKHA7 | Homo sapien | 127.137 | 2 | 2 | 0.0E+00 | 0.0E+00 | 1.7E+04 | 0.0E+00 | 2.2E+04 | 3.6E+04 | 0.0E+00 | 0.0E+00 | 0.0E+00 | 0.0E+00 | 0.0E+00 | 0.0E+00 | [illegible] |
| 5764 | 1 | Q6N063 | Q6N063 | 2-oxoglutarat | OGFOD2 | Homo sapien | 38.996 | 2 | 2 | 0.0E+00 | 0.0E+00 | 0.0E+00 | 0.0E+00 | 1.5E+03 | 0.0E+00 | 0.0E+00 | 0.0E+00 | 4.5E+04 | 0.0E+00 | 0.0E+00 | 0.0E+00 | [illegible] |
| 5765 | 1 | Q6NW34 | Q6NW34 | Nucleolus an | NEPRO | Homo sapien | 64.553 | 2 | 2 | 0.0E+00 | 0.0E+00 | 0.0E+00 | 0.0E+00 | 0.0E+00 | 0.0E+00 | 0.0E+00 | 0.0E+00 | 0.0E+00 | 0.0E+00 | 0.0E+00 | 0.0E+00 | [illegible] |
| 5766 | 1 | Q6P1L5 | Q6P1L5 | Protein FAM1 | FAM117B | Homo sapien | 61.968 | 2 | 2 | 2.1E+04 | 2.5E+04 | 0.0E+00 | 0.0E+00 | 2.5E+04 | 0.0E+00 | 1.7E+04 | 3.3E+04 | 0.0E+00 | 0.0E+00 | 0.0E+00 | 1.4E+04 | [illegible] |
| 5767 | 1 | Q6P1L8 | Q6P1L8 | Large ribosor | MRPL14 | Homo sapien | 15.949 | 2 | 2 | 3.5E+04 | 3.5E+04 | 3.7E+04 | 2.7E+04 | 3.5E+04 | 3.3E+04 | 7.7E+04 | 7.1E+04 | 8.6E+04 | 7.7E+04 | 7.0E+04 | 7.5E+04 | [illegible] |
| 5768 | 1 | Q6P4R8 | Q6P4R8 | Nuclear facto | NFRKB | Homo sapien | 139.004 | 2 | 2 | 0.0E+00 | 0.0E+00 | 0.0E+00 | 0.0E+00 | 0.0E+00 | 0.0E+00 | 0.0E+00 | 0.0E+00 | 0.0E+00 | 0.0E+00 | 0.0E+00 | 0.0E+00 | [illegible] |
| 5769 | 1 | Q6P587 | Q6P587 | Oxaloacetate | FAHD1 | Homo sapien | 24.541 | 2 | 2 | 0.0E+00 | 0.0E+00 | 0.0E+00 | 0.0E+00 | 0.0E+00 | 4.9E+04 | 0.0E+00 | 1.5E+04 | 1.3E+04 | 1.3E+04 | 0.0E+00 | 1.1E+04 | [illegible] |
| 5770 | 1 | Q6P9B6 | Q6P9B6 | MTOR-assoc | MEAK7 | Homo sapien | 50.996 | 2 | 2 | 0.0E+00 | 1.7E+05 | 0.0E+00 | 0.0E+00 | 0.0E+00 | 2.1E+05 | 0.0E+00 | 0.0E+00 | 0.0E+00 | 0.0E+00 | 0.0E+00 | 0.0E+00 | [illegible] |
| 5771 | 1 | Q6PCB5 | Q6PCB5 | Lysine-specif | RSBN1L | Homo sapien | 94.871 | 2 | 2 | 0.0E+00 | 0.0E+00 | 0.0E+00 | 0.0E+00 | 0.0E+00 | 0.0E+00 | 0.0E+00 | 0.0E+00 | 0.0E+00 | 0.0E+00 | 0.0E+00 | 0.0E+00 | [illegible] |
| 5772 | 1 | Q6PI47 | Q6PI47 | BTB/POZ dor | KCTD18 | Homo sapien | 46.74 | 2 | 2 | 0.0E+00 | 0.0E+00 | 6.1E+03 | 6.7E+03 | 6.2E+04 | 3.5E+04 | 0.0E+00 | 0.0E+00 | 0.0E+00 | 5.0E+03 | 1.7E+04 | 0.0E+00 | [illegible] |
| 5773 | 1 | Q6PIY7 | Q6PIY7 | Poly(A) RNA | TENT2 | Homo sapien | 56.027 | 2 | 2 | 0.0E+00 | 0.0E+00 | 1.4E+04 | 0.0E+00 | 5.4E+03 | 0.0E+00 | 0.0E+00 | 0.0E+00 | 0.0E+00 | 0.0E+00 | 0.0E+00 | 0.0E+00 | [illegible] |
| 5774 | 1 | Q6RW13 | Q6RW13 | Type-1 angio | AGTRAP | Homo sapien | 17.42 | 2 | 2 | 3.0E+05 | 2.8E+05 | 1.6E+05 | 2.7E+05 | 3.4E+05 | 2.7E+05 | 0.0E+00 | 3.2E+05 | 0.0E+00 | 0.0E+00 | 2.8E+05 | 0.0E+00 | [illegible] |
| 5775 | 1 | Q6SPF0 | Q6SPF0 | Sterile alpha | SAMD1 | Homo sapien | 56.05 | 2 | 2 | 2.7E+04 | 2.6E+04 | 2.8E+04 | 1.9E+04 | 2.5E+04 | 0.0E+00 | 4.0E+04 | 5.4E+04 | 0.0E+00 | 0.0E+00 | 0.0E+00 | 0.0E+00 | [illegible] |
| 5776 | 1 | Q6UW68 | Q6UW68 | Transmembr | TMEM205 | Homo sapien | 21.197 | 2 | 2 | 2.5E+05 | 2.7E+05 | 3.0E+05 | 2.6E+05 | 3.4E+05 | 2.4E+05 | 4.6E+05 | 3.2E+05 | 3.0E+05 | 4.1E+05 | 2.8E+05 | 1.0E+05 | [illegible] |
| 5777 | 1 | Q6UWJ1 | Q6UWJ1 | Transmembr | TMCO3 | Homo sapien | 75.598 | 2 | 2 | 0.0E+00 | 0.0E+00 | 0.0E+00 | 0.0E+00 | 0.0E+00 | 0.0E+00 | 4.4E+04 | 4.2E+04 | 4.0E+04 | 4.0E+04 | 0.0E+00 | 0.0E+00 | [illegible] |
| 5778 | 1 | Q6UXI7 | Q6UXI7 | Vitrin | VIT | Homo sapien | 73.93 | 2 | 2 | 0.0E+00 | 0.0E+00 | 0.0E+00 | 0.0E+00 | 1.3E+04 | 1.3E+04 | 0.0E+00 | 8.8E+03 | 0.0E+00 | 0.0E+00 | 0.0E+00 | 0.0E+00 | [illegible] |
| 5779 | 1 | Q6UXI9 | Q6UXI9 | Nephronectin | NPNT | Homo sapien | 61.906 | 2 | 2 | 1.3E+05 | 0.0E+00 | 1.0E+05 | 1.6E+05 | 1.0E+05 | 2.3E+05 | 0.0E+00 | 0.0E+00 | 0.0E+00 | 0.0E+00 | 0.0E+00 | 4.1E+04 | [illegible] |
| 5780 | 1 | Q6ZMD2 | Q6ZMD2 | Protein spins | SPNS3 | Homo sapien | 54.769 | 2 | 2 | 0.0E+00 | 0.0E+00 | 0.0E+00 | 0.0E+00 | 0.0E+00 | 0.0E+00 | 0.0E+00 | 0.0E+00 | 0.0E+00 | 0.0E+00 | 0.0E+00 | 0.0E+00 | [illegible] |
| 5781 | 1 | Q6ZNE5 | Q6ZNE5 | Beclin 1-assc | ATG14 | Homo sapien | 55.309 | 2 | 2 | 1.1E+05 | 0.0E+00 | 0.0E+00 | 6.4E+03 | 6.9E+03 | 3.4E+03 | 0.0E+00 | 1.4E+05 | 1.2E+05 | 0.0E+00 | 1.1E+05 | 0.0E+00 | [illegible] |
| 5782 | 1 | Q6ZPD9 | Q6ZPD9 | Protein C-ma | DPY19L3 | Homo sapien | 83.199 | 2 | 2 | 0.0E+00 | 0.0E+00 | 0.0E+00 | 3.0E+04 | 1.4E+04 | 0.0E+00 | 0.0E+00 | 0.0E+00 | 0.0E+00 | 0.0E+00 | 0.0E+00 | 0.0E+00 | [illegible] |
| 5783 | 1 | Q6ZS30 | Q6ZS30 | Neurobeachi | NBEAL1 | Homo sapien | 307.243 | 2 | 2 | 0.0E+00 | 0.0E+00 | 0.0E+00 | 0.0E+00 | 0.0E+00 | 0.0E+00 | 0.0E+00 | 0.0E+00 | 0.0E+00 | 0.0E+00 | 0.0E+00 | 0.0E+00 | [illegible] |
| 5784 | 1 | Q6ZU80 | Q6ZU80 | Centrosomal | CEP128 | Homo sapien | 128.016 | 2 | 2 | 0.0E+00 | 0.0E+00 | 0.0E+00 | 0.0E+00 | 2.7E+04 | 0.0E+00 | 0.0E+00 | 3.7E+04 | 2.4E+04 | 0.0E+00 | 0.0E+00 | 0.0E+00 | [illegible] |
| 5785 | 1 | Q6ZUX7 | Q6ZUX7 | LHFPL tetras | LHFPL2 | Homo sapien | 24.483 | 2 | 2 | 2.3E+05 | 6.4E+04 | 2.9E+05 | 4.5E+04 | 2.0E+05 | 0.0E+00 | 3.8E+05 | 4.1E+05 | 2.7E+05 | 3.5E+05 | 2.2E+05 | 3.7E+05 | [illegible] |
| 5786 | 1 | Q6ZV70 | Q6ZV70 | LanC-like pro | LANCL3 | Homo sapien | 46.318 | 2 | 2 | 1.4E+04 | 2.6E+04 | 2.4E+04 | 9.6E+03 | 1.5E+04 | 3.2E+04 | 1.8E+04 | 1.0E+04 | 0.0E+00 | 2.2E+04 | 0.0E+00 | 2.4E+04 | [illegible] |
| 5787 | 1 | Q6ZVD8 | Q6ZVD8 | PH domain le | PHLPP2 | Homo sapien | 146.753 | 2 | 2 | 0.0E+00 | 0.0E+00 | 0.0E+00 | 0.0E+00 | 0.0E+00 | 0.0E+00 | 2.2E+04 | 0.0E+00 | 0.0E+00 | 0.0E+00 | 0.0E+00 | 0.0E+00 | [illegible] |
| 5788 | 1 | Q6ZVF9 | Q6ZVF9 | G protein-reg | GPRIN3 | Homo sapien | 82.441 | 2 | 2 | 0.0E+00 | 0.0E+00 | 0.0E+00 | 0.0E+00 | 0.0E+00 | 0.0E+00 | 0.0E+00 | 0.0E+00 | 0.0E+00 | 0.0E+00 | 0.0E+00 | 0.0E+00 | [illegible] |
| 5789 | 1 | Q6ZXV5 | Q6ZXV5 | Protein O-ma | TMTC3 | Homo sapien | 104.01 | 2 | 2 | 5.6E+03 | 6.1E+03 | 0.0E+00 | 0.0E+00 | 1.6E+05 | 0.0E+00 | 0.0E+00 | 0.0E+00 | 7.3E+03 | 0.0E+00 | 0.0E+00 | 0.0E+00 | [illegible] |
| 5790 | 1 | Q70IA8 | Q70IA8 | MOB kinase a | MOB3C | Homo sapien | 25.623 | 2 | 2 | 7.9E+05 | 8.3E+05 | 6.4E+05 | 5.2E+05 | 5.6E+05 | 4.7E+05 | 6.2E+05 | 6.9E+05 | 4.3E+05 | 8.7E+05 | 9.3E+05 | 8.8E+05 | [illegible] |
| 5791 | 1 | Q712K3 | Q712K3 | Ubiquitin-con | UBE2R2 | Homo sapien | 27.164 | 3 | 2 | 6.8E+04 | 0.0E+00 | 0.0E+00 | 6.2E+04 | 6.9E+04 | 6.0E+04 | 9.7E+04 | 1.5E+05 | 1.1E+05 | 5.6E+04 | 0.0E+00 | 1.6E+05 | [illegible] |
| 5792 | 1 | Q76FK4 | Q76FK4 | Nucleolar pro | NOL8 | Homo sapien | 131.619 | 2 | 2 | 0.0E+00 | 0.0E+00 | 0.0E+00 | 0.0E+00 | 0.0E+00 | 0.0E+00 | 0.0E+00 | 0.0E+00 | 0.0E+00 | 0.0E+00 | 0.0E+00 | 0.0E+00 | [illegible] |
| 5793 | 1 | Q7KYR7 | Q7KYR7 | Butyrophilin s | BTN2A1 | Homo sapien | 59.633 | 2 | 2 | 0.0E+00 | 0.0E+00 | 0.0E+00 | 0.0E+00 | 0.0E+00 | 4.2E+03 | 0.0E+00 | 0.0E+00 | 0.0E+00 | 0.0E+00 | 0.0E+00 | 0.0E+00 | [illegible] |
| 5794 | 1 | Q7L5A3 | Q7L5A3 | Atos homolog | ATOSB | Homo sapien | 56.687 | 2 | 2 | 0.0E+00 | 2.4E+04 | 0.0E+00 | 1.1E+05 | 1.7E+04 | 1.3E+04 | 0.0E+00 | 0.0E+00 | 0.0E+00 | 9.2E+04 | 0.0E+00 | 0.0E+00 | [illegible] |
| 5795 | 1 | Q7Z2E3 | Q7Z2E3 | Aprataxin | APTX | Homo sapien | 40.738 | 2 | 2 | 0.0E+00 | 0.0E+00 | 0.0E+00 | 0.0E+00 | 7.0E+03 | 0.0E+00 | 0.0E+00 | 0.0E+00 | 0.0E+00 | 0.0E+00 | 0.0E+00 | 0.0E+00 | [illegible] |
| 5796 | 1 | Q7Z3F1 | Q7Z3F1 | Lysosomal c | GPR155 | Homo sapien | 96.92 | 2 | 2 | 2.0E+03 | 0.0E+00 | 0.0E+00 | 0.0E+00 | 0.0E+00 | 0.0E+00 | 0.0E+00 | 7.6E+03 | 0.0E+00 | 6.1E+03 | 0.0E+00 | 0.0E+00 | [illegible] |
| 5797 | 1 | Q7Z3Y7 | Q7Z3Y7 | Keratin, type | KRT28 | Homo sapien | 50.568 | 2 | 2 | 0.0E+00 | 0.0E+00 | 0.0E+00 | 0.0E+00 | 0.0E+00 | 0.0E+00 | 0.0E+00 | 0.0E+00 | 0.0E+00 | 0.0E+00 | 0.0E+00 | 0.0E+00 | [illegible] |
| 5798 | 1 | Q7Z3Z0 | Q7Z3Z0 | Keratin, type | KRT25 | Homo sapien | 49.317 | 7 | 2 | 0.0E+00 | 0.0E+00 | 0.0E+00 | 0.0E+00 | 0.0E+00 | 0.0E+00 | 0.0E+00 | 0.0E+00 | 0.0E+00 | 0.0E+00 | 0.0E+00 | 0.0E+00 | [illegible] |
| 5799 | 1 | Q7Z6K3 | Q7Z6K3 | Protein preny | PTAR1 | Homo sapien | 46.406 | 2 | 2 | 0.0E+00 | 0.0E+00 | 0.0E+00 | 0.0E+00 | 0.0E+00 | 0.0E+00 | 0.0E+00 | 0.0E+00 | 4.7E+04 | 0.0E+00 | 0.0E+00 | 0.0E+00 | [illegible] |
| 5800 | 1 | Q7Z7A3 | Q7Z7A3 | Cytoplasmic | CTU1 | Homo sapien | 36.45 | 2 | 2 | 0.0E+00 | 0.0E+00 | 0.0E+00 | 0.0E+00 | 0.0E+00 | 0.0E+00 | 0.0E+00 | 0.0E+00 | 0.0E+00 | 0.0E+00 | 0.0E+00 | 0.0E+00 | [illegible] |
| 5801 | 1 | Q7Z7E8 | Q7Z7E8 | Ubiquitin-con | UBE2Q1 | Homo sapien | 46.127 | 3 | 2 | 5.1E+04 | 0.0E+00 | 4.8E+04 | 5.4E+04 | 4.8E+04 | 5.0E+04 | 1.8E+04 | 0.0E+00 | 1.5E+04 | 1.8E+04 | 4.8E+04 | 5.5E+04 | [illegible] |
| 5802 | 1 | Q86SZ2 | Q86SZ2 | Trafficking pr | TRAPPC6B | Homo sapien | 17.984 | 2 | 2 | 3.3E+04 | 0.0E+00 | 2.7E+04 | 3.1E+04 | 2.1E+04 | 0.0E+00 | 0.0E+00 | 0.0E+00 | 0.0E+00 | 0.0E+00 | 0.0E+00 | 0.0E+00 | [illegible] |
| 5803 | 1 | Q86TJ2 | Q86TJ2 | Transcription | TADA2B | Homo sapien | 48.47 | 2 | 2 | 4.7E+03 | 0.0E+00 | 4.8E+04 | 3.4E+03 | 0.0E+00 | 3.0E+03 | 7.2E+04 | 0.0E+00 | 0.0E+00 | 8.9E+04 | 0.0E+00 | 0.0E+00 | [illegible] |
| 5804 | 1 | Q86TY3 | Q86TY3 | Armadillo-like | ARMH4 | Homo sapien | 84.173 | 2 | 2 | 0.0E+00 | 9.7E+03 | 0.0E+00 | 3.1E+04 | 4.8E+03 | 1.5E+04 | 0.0E+00 | 0.0E+00 | 0.0E+00 | 3.8E+04 | 0.0E+00 | 0.0E+00 | [illegible] |
| 5805 | 1 | Q86UB9 | Q86UB9 | Transmembr | TMEM135 | Homo sapien | 52.289 | 2 | 2 | 0.0E+00 | 0.0E+00 | 0.0E+00 | 0.0E+00 | 0.0E+00 | 0.0E+00 | 0.0E+00 | 0.0E+00 | 0.0E+00 | 9.0E+05 | 0.0E+00 | 0.0E+00 | [illegible] |
| 5806 | 1 | Q86UE8 | Q86UE8 | Serine/threon | TLK2 | Homo sapien | 87.662 | 2 | 2 | 0.0E+00 | 4.7E+03 | 0.0E+00 | 1.7E+03 | 3.6E+03 | 0.0E+00 | 0.0E+00 | 0.0E+00 | 0.0E+00 | 0.0E+00 | 0.0E+00 | 0.0E+00 | [illegible] |
| 5807 | 1 | Q86UF1 | Q86UF1 | Tetraspanin- | TSPAN33 | Homo sapien | 31.539 | 2 | 2 | 5.9E+07 | 1.1E+06 | 7.7E+07 | 1.9E+06 | 5.6E+07 | 1.8E+06 | 1.2E+06 | 9.2E+05 | 8.8E+05 | 0.0E+00 | 1.3E+06 | 1.0E+06 | [illegible] |
| 5808 | 1 | Q86UL3 | Q86UL3 | Glycerol-3-ph | GPAT4 | Homo sapien | 52.07 | 2 | 2 | 2.4E+04 | 9.1E+03 | 0.0E+00 | 1.0E+04 | 3.2E+04 | 2.9E+05 | 0.0E+00 | 1.2E+04 | 2.1E+05 | 1.7E+04 | 0.0E+00 | 0.0E+00 | [illegible] |
| 5809 | 1 | Q86UW6 | Q86UW6 | NEDD4-bindi | N4BP2 | Homo sapien | 198.806 | 2 | 2 | 6.8E+03 | 0.0E+00 | 3.3E+03 | 0.0E+00 | 0.0E+00 | 0.0E+00 | 0.0E+00 | 0.0E+00 | 0.0E+00 | 0.0E+00 | 0.0E+00 | 0.0E+00 | [illegible] |
| 5810 | 1 | Q86V21 | Q86V21 | Acetoacetyl- | AACS | Homo sapien | 75.144 | 2 | 2 | 0.0E+00 | 0.0E+00 | 0.0E+00 | 0.0E+00 | 0.0E+00 | 0.0E+00 | 0.0E+00 | 0.0E+00 | 0.0E+00 | 0.0E+00 | 0.0E+00 | 0.0E+00 | [illegible] |
| 5811 | 1 | Q86VP1 | Q86VP1 | Tax1-binding | TAX1BP1 | Homo sapien | 90.88 | 2 | 2 | 4.0E+03 | 0.0E+00 | 0.0E+00 | 2.6E+04 | 0.0E+00 | 0.0E+00 | 4.9E+03 | 0.0E+00 | 0.0E+00 | 0.0E+00 | 0.0E+00 | 0.0E+00 | [illegible] |
| 5812 | 1 | Q86VW0 | Q86VW0 | SEC14 doma | SESTD1 | Homo sapien | 79.348 | 2 | 2 | 0.0E+00 | 0.0E+00 | 2.4E+04 | 2.7E+04 | 0.0E+00 | 0.0E+00 | 3.7E+04 | 2.8E+04 | 0.0E+00 | 3.3E+04 | 6.0E+04 | 4.5E+04 | [illegible] |
| 5813 | 1 | Q86WA8 | Q86WA8 | Lon protease | LONP2 | Homo sapien | 94.62 | 2 | 2 | 0.0E+00 | 0.0E+00 | 3.2E+04 | 4.3E+04 | 0.0E+00 | 0.0E+00 | 0.0E+00 | 0.0E+00 | 0.0E+00 | 5.2E+04 | 0.0E+00 | 0.0E+00 | [illegible] |
| 5814 | 1 | Q86WB0 | Q86WB0 | Zinc finger C | ZC3HC1 | Homo sapien | 55.262 | 2 | 2 | 0.0E+00 | 0.0E+00 | 0.0E+00 | 0.0E+00 | 0.0E+00 | 0.0E+00 | 0.0E+00 | 0.0E+00 | 0.0E+00 | 0.0E+00 | 0.0E+00 | 0.0E+00 | [illegible] |
| 5815 | 1 | Q86XN7 | Q86XN7 | Proline and s | PROSER1 | Homo sapien | 95.699 | 2 | 2 | 0.0E+00 | 8.0E+05 | 0.0E+00 | 0.0E+00 | 4.6E+05 | 4.1E+05 | 3.3E+06 | 0.0E+00 | 0.0E+00 | 0.0E+00 | 0.0E+00 | 0.0E+00 | [illegible] |
| 5816 | 1 | Q86Y39 | Q86Y39 | NADH dehyd | NDUFA11 | Homo sapien | 14.849 | 2 | 2 | 0.0E+00 | 0.0E+00 | 1.4E+05 | 2.2E+05 | 1.3E+05 | 1.5E+05 | 0.0E+00 | 0.0E+00 | 0.0E+00 | 0.0E+00 | 0.0E+00 | 0.0E+00 | [illegible] |
| 5817 | 1 | Q86YB8 | Q86YB8 | ERO1-like pr | ERO1B | Homo sapien | 53.543 | 2 | 2 | 3.7E+04 | 2.9E+04 | 3.2E+04 | 3.7E+04 | 0.0E+00 | 3.5E+03 | 7.2E+04 | 3.7E+04 | 6.6E+04 | 5.5E+04 | 0.0E+00 | 0.0E+00 | [illegible] |
| 5818 | 1 | Q86YT9 | Q86YT9 | Junctional ad | JAML | Homo sapien | 44.339 | 2 | 2 | 1.2E+04 | 0.0E+00 | 7.0E+03 | 8.3E+03 | 2.2E+04 | 0.0E+00 | 1.1E+04 | 0.0E+00 | 0.0E+00 | 0.0E+00 | 0.0E+00 | 0.0E+00 | [illegible] |
| 5819 | 1 | Q8IU85 | Q8IU85 | Calcium/calm | CAMK1D | Homo sapien | 42.915 | 2 | 2 | 0.0E+00 | 0.0E+00 | 0.0E+00 | 0.0E+00 | 2.1E+04 | 0.0E+00 | 0.0E+00 | 8.9E+03 | 0.0E+00 | 0.0E+00 | 8.8E+03 | 0.0E+00 | [illegible] |
| 5820 | 1 | Q8IUR0 | Q8IUR0 | Trafficking pr | TRAPPC5 | Homo sapien | 20.781 | 2 | 2 | 3.7E+04 | 3.7E+04 | 5.1E+04 | 5.3E+04 | 4.2E+04 | 3.1E+04 | 7.7E+04 | 2.6E+04 | 2.5E+04 | 9.5E+04 | 9.0E+04 | 6.2E+04 | [illegible] |
| [illegible] | | | | | | | | | | | | | | | | | | | | | | |

| | | | | | | | | | | | | | | | | | | | | |
|---|---|---|---|---|---|---|---|---|---|---|---|---|---|---|---|---|---|---|---|---|
| 6286 | 1 | A0A0A0MRZ | A0A0A0MRZ | ImmunoglobuIGLV5-52 | Homo sapien | 13.447 | 1 | 1 | 0.0E+00 | 0.0E+00 | 0.0E+00 | 0.0E+00 | 8.2E+03 | 0.0E+00 | 0.0E+00 | 0.0E+00 | 0.0E+00 | 0.0E+00 | 0.0E+00 | 0.0E+00 |
| 6287 | 1 | A0A0A0MS1 | A0A0A0MS1 | ImmunoglobuIGHV1-45 | Homo sapien | 13.505 | 1 | 1 | 0.0E+00 | 5.6E+05 | 7.6E+05 | 4.5E+05 | 0.0E+00 | 8.2E+05 | 0.0E+00 | 2.6E+05 | 0.0E+00 | 7.3E+05 | 2.6E+06 | 0.0E+00 |
| 6288 | 1 | A0A0B4J1U | A0A0B4J1U | ImmunoglobuIGLV1-36 | Homo sapien | 12.479 | 1 | 1 | 0.0E+00 | 0.0E+00 | 0.0E+00 | 0.0E+00 | 0.0E+00 | 0.0E+00 | 1.1E+04 | 0.0E+00 | 2.6E+04 | 3.0E+04 | 0.0E+00 | 0.0E+00 |
| 6289 | 1 | A0A0B4J1U | A0A0B4J1U7 | ImmunoglobuIGHV6-1 | Homo sapien | 13.48 | 1 | 1 | 9.9E+05 | 1.0E+06 | 3.6E+06 | 1.5E+06 | 1.5E+06 | 7.0E+05 | 1.3E+06 | 5.2E+05 | 8.9E+05 | 1.8E+06 | 1.9E+06 | 5.5E+05 |
| 6290 | 1 | A0A0B4J1V2 | A0A0B4J1V2 | ImmunoglobuIGHV2-26 | Homo sapien | 13.18 | 1 | 1 | 2.0E+05 | 3.6E+05 | 8.6E+05 | 4.5E+05 | 1.4E+05 | 3.8E+05 | 8.7E+05 | 0.0E+00 | 2.4E+05 | 0.0E+00 | 0.0E+00 | 0.0E+00 |
| 6291 | 1 | A0A0B4J1X5 | A0A0B4J1X5 | ImmunoglobuIGHV3-74 | Homo sapien | 12.838 | 1 | 1 | 0.0E+00 | 0.0E+00 | 0.0E+00 | 0.0E+00 | 0.0E+00 | 0.0E+00 | 0.0E+00 | 0.0E+00 | 0.0E+00 | 0.0E+00 | 0.0E+00 | 0.0E+00 |
| 6292 | 1 | A0A0B4J1X8 | A0A0B4J1X8 | ImmunoglobuIGHV3-43 | Homo sapien | 13.077 | 1 | 1 | 0.0E+00 | 0.0E+00 | 0.0E+00 | 0.0E+00 | 0.0E+00 | 0.0E+00 | 3.8E+04 | 0.0E+00 | 0.0E+00 | 0.0E+00 | 0.0E+00 | 0.0E+00 |
| 6293 | 1 | A0A0B4J265 | A0A0B4J265 | T cell receptTRAV26-2 | Homo sapien | 12.172 | 1 | 1 | 0.0E+00 | 0.0E+00 | 0.0E+00 | 0.0E+00 | 0.0E+00 | 0.0E+00 | 0.0E+00 | 0.0E+00 | 0.0E+00 | 0.0E+00 | 0.0E+00 | 0.0E+00 |
| 6294 | 1 | A0A0B4J2F0 | A0A0B4J2F0 | Protein PIGBPIGBOS1 | Homo sapien | 6.311 | 1 | 1 | 8.3E+03 | 8.2E+03 | 7.4E+03 | 0.0E+00 | 1.1E+04 | 1.1E+04 | 1.9E+04 | 2.0E+04 | 0.0E+00 | 8.9E+03 | 1.4E+04 | 1.5E+04 |
| 6295 | 1 | A0A0C4DH2 | A0A0C4DH2 | ImmunoglobuIGKV6-21 | Homo sapien | 12.428 | 2 | 1 | 2.1E+06 | 1.4E+05 | 3.8E+06 | 7.7E+05 | 1.0E+06 | 0.0E+00 | 0.0E+00 | 2.6E+05 | 6.1E+05 | 1.1E+06 | 3.1E+06 | 0.0E+00 |
| 6296 | 1 | A0A0C4DH3 | A0A0C4DH3 | ImmunoglobuIGHV1-24 | Homo sapien | 12.824 | 1 | 1 | 3.4E+04 | 8.4E+04 | 1.1E+05 | 1.0E+05 | 7.8E+04 | 6.9E+04 | 5.4E+04 | 1.8E+04 | 6.6E+04 | 9.3E+04 | 1.0E+05 | 3.4E+04 |
| 6297 | 1 | A0A0C4DH3 | A0A0C4DH3 | Probable nonIGHV3-38 | Homo sapien | 12.761 | 1 | 1 | 1.4E+07 | 1.3E+07 | 1.9E+07 | 9.6E+06 | 8.6E+06 | 8.9E+06 | 1.8E+07 | 9.1E+06 | 1.9E+07 | 1.4E+07 | 2.7E+07 | 2.1E+07 |
| 6298 | 1 | A0A0C4DH3 | A0A0C4DH3 | ImmunoglobuIGHV5-51 | Homo sapien | 12.673 | 2 | 1 | 0.0E+00 | 0.0E+00 | 2.9E+04 | 3.1E+04 | 1.7E+04 | 1.8E+04 | 9.6E+04 | 0.0E+00 | 0.0E+00 | 1.2E+05 | 0.0E+00 | 0.0E+00 |
| 6299 | 2 | A0A0C4DH6 | A0A0C4DH6 | ImmunoglobuIGKV1-8 | Homo sapien | 12.535 | 1 | 1 | 2.3E+05 | 3.0E+06 | 9.4E+05 | 5.7E+05 | 6.0E+06 | 3.3E+05 | 5.0E+06 | 0.0E+00 | 0.0E+00 | 0.0E+00 | 0.0E+00 | 0.0E+00 |
| 6300 | 1 | A0A0C4DH7 | A0A0C4DH7 | ImmunoglobuIGKV1-6 | Homo sapien | 12.698 | 2 | 1 | 0.0E+00 | 0.0E+00 | 0.0E+00 | 0.0E+00 | 0.0E+00 | 0.0E+00 | 4.0E+05 | 2.3E+05 | 3.6E+05 | 4.1E+05 | 0.0E+00 | 0.0E+00 |
| 6301 | 2 | A0A0C4DH7 | A0A0C4DH7 | ImmunoglobuIGKV1-12 | Homo sapien | 12.646 | 1 | 1 | 2.4E+05 | 1.3E+05 | 3.8E+05 | 1.9E+05 | 1.6E+05 | 1.2E+05 | 1.7E+05 | 1.4E+05 | 2.4E+05 | 2.5E+05 | 5.9E+05 | 1.8E+05 |
| 6302 | 1 | A0A0G2JKW | A0A0G2JKW | Uncharacteri- | Homo sapien | 3.491 | 1 | 1 | 0.0E+00 | 0.0E+00 | 0.0E+00 | 0.0E+00 | 3.1E+04 | 0.0E+00 | 0.0E+00 | 0.0E+00 | 0.0E+00 | 0.0E+00 | 0.0E+00 | 0.0E+00 |
| 6303 | 1 | A0A0J9YW | A0A0J9YW | Immunoglobu- | Homo sapien | 13.249 | 1 | 1 | 0.0E+00 | 0.0E+00 | 3.3E+04 | 2.3E+04 | 1.2E+04 | 2.9E+04 | 1.1E+04 | 7.2E+04 | 0.0E+00 | 2.2E+04 | 2.4E+04 | 8.0E+04 |
| 6304 | 1 | A0A0J9YWN | A0A0J9YWN | ImmunoglobuIGHJ2 | Homo sapien | 2.047 | 1 | 1 | 3.5E+04 | 7.7E+04 | 4.3E+04 | 8.4E+04 | 2.8E+04 | 3.2E+04 | 2.4E+05 | 0.0E+00 | 0.0E+00 | 4.0E+05 | 0.0E+00 | 1.0E+05 |
| 6305 | 1 | A0A0J9YX35 | A0A0J9YX35 | ImmunoglobuIGHV3-64D | Homo sapien | 12.822 | 1 | 1 | 2.4E+08 | 2.8E+08 | 3.4E+08 | 2.4E+08 | 3.2E+08 | 2.1E+08 | 3.4E+08 | 2.2E+08 | 3.9E+08 | 4.8E+08 | 4.0E+08 | 2.4E+08 |
| 6306 | 1 | A0A0J9YY99 | A0A0J9YY99 | Ig-like domain- | Homo sapien | 12.962 | 2 | 1 | 7.2E+05 | 3.9E+05 | 3.6E+05 | 4.1E+05 | 2.3E+05 | 2.2E+05 | 6.6E+05 | 3.4E+05 | 3.4E+05 | 8.5E+05 | 4.7E+05 | 4.2E+05 |
| 6307 | 1 | A0A1B0GTC | A0A1B0GTC | UncharacteriC3orf85 | Homo sapien | 10.386 | 1 | 1 | 0.0E+00 | 8.2E+03 | 0.0E+00 | 8.4E+03 | 0.0E+00 | 0.0E+00 | 0.0E+00 | 1.5E+04 | 0.0E+00 | 1.3E+04 | 0.0E+00 | 9.5E+03 |
| 6308 | 1 | A0AAG2SXZ | A0AAG2SXZ | Ig-like domain- | Homo sapien | 10.777 | 3 | 1 | 0.0E+00 | 0.0E+00 | 0.0E+00 | 0.0E+00 | 0.0E+00 | 5.8E+04 | 0.0E+00 | 0.0E+00 | 3.9E+04 | 7.5E+04 | 3.6E+04 | 3.5E+04 |
| 6309 | 1 | A0AAG2TV0 | A0AAG2TV0 | HCG2003024hCG_200302 | Homo sapien | 13.051 | 1 | 1 | 6.5E+05 | 2.3E+05 | 2.1E+05 | 3.1E+05 | 4.2E+05 | 2.1E+05 | 4.7E+05 | 4.6E+05 | 3.8E+05 | 6.9E+05 | 4.8E+05 | 3.0E+05 |
| 6310 | 1 | A0AAG2TVE | A0AAG2TVE | Ig-like domain- | Homo sapien | 13.04 | 1 | 1 | 0.0E+00 | 0.0E+00 | 0.0E+00 | 0.0E+00 | 0.0E+00 | 0.0E+00 | 0.0E+00 | 0.0E+00 | 1.5E+04 | 0.0E+00 | 2.3E+04 | 0.0E+00 |
| 6311 | 1 | A0AV96 | A0AV96 | RNA-binding RBM47 | Homo sapien | 64.1 | 1 | 1 | 6.4E+03 | 7.6E+03 | 5.9E+03 | 0.0E+00 | 0.0E+00 | 0.0E+00 | 6.8E+03 | 0.0E+00 | 0.0E+00 | 0.0E+00 | 0.0E+00 | 0.0E+00 |
| 6312 | 1 | A1L3X0 | A1L3X0 | Very long chELOVL7 | Homo sapien | 33.356 | 1 | 1 | 6.3E+04 | 0.0E+00 | 5.8E+04 | 0.0E+00 | 0.0E+00 | 7.8E+04 | 8.3E+04 | 7.4E+04 | 6.9E+04 | 1.0E+05 | 9.4E+04 | 6.4E+04 |
| 6313 | 1 | A2A288 | A2A288 | Probable riboZC3H12D | Homo sapien | 58.076 | 1 | 1 | 0.0E+00 | 0.0E+00 | 0.0E+00 | 2.6E+04 | 0.0E+00 | 0.0E+00 | 0.0E+00 | 0.0E+00 | 0.0E+00 | 0.0E+00 | 0.0E+00 | 0.0E+00 |
| 6314 | 1 | A2RU67 | A2RU67 | Protein FAMFAM234B | Homo sapien | 67.041 | 1 | 1 | 0.0E+00 | 0.0E+00 | 0.0E+00 | 0.0E+00 | 0.0E+00 | 0.0E+00 | 0.0E+00 | 0.0E+00 | 0.0E+00 | 0.0E+00 | 0.0E+00 | 0.0E+00 |
| 6315 | 1 | A2VDJ0 | A2VDJ0 | TransmembrTMEM131L | Homo sapien | 179.344 | 1 | 1 | 0.0E+00 | 0.0E+00 | 0.0E+00 | 0.0E+00 | 0.0E+00 | 0.0E+00 | 0.0E+00 | 0.0E+00 | 0.0E+00 | 0.0E+00 | 0.0E+00 | 0.0E+00 |
| 6316 | 1 | A5D8V6 | A5D8V6 | Vacuolar proVPS37C | Homo sapien | 38.66 | 1 | 1 | 4.4E+05 | 5.9E+05 | 2.5E+05 | 3.0E+05 | 5.0E+05 | 4.9E+05 | 4.9E+05 | 6.3E+05 | 5.2E+05 | 7.0E+05 | 8.3E+05 | 9.9E+05 |
| 6317 | 1 | A5PLL7 | A5PLL7 | PlasmanylethPEDS1 | Homo sapien | 31.137 | 1 | 1 | 0.0E+00 | 0.0E+00 | 0.0E+00 | 0.0E+00 | 0.0E+00 | 0.0E+00 | 0.0E+00 | 0.0E+00 | 0.0E+00 | 0.0E+00 | 0.0E+00 | 0.0E+00 |
| 6318 | 1 | A6NED2 | A6NED2 | RCC1 domaiRCCD1 | Homo sapien | 40.08 | 1 | 1 | 0.0E+00 | 0.0E+00 | 0.0E+00 | 0.0E+00 | 0.0E+00 | 0.0E+00 | 0.0E+00 | 0.0E+00 | 0.0E+00 | 0.0E+00 | 0.0E+00 | 0.0E+00 |
| 6319 | 1 | A6NEE1 | A6NEE1 | Pleckstrin hoPLEKHD1 | Homo sapien | 59.204 | 1 | 1 | 0.0E+00 | 0.0E+00 | 0.0E+00 | 0.0E+00 | 5.8E+04 | 5.0E+04 | 0.0E+00 | 4.4E+04 | 0.0E+00 | 0.0E+00 | 4.8E+04 | 0.0E+00 |
| 6320 | 1 | A6NFQ2 | A6NFQ2 | TRPM8 chanTCAF2 | Homo sapien | 100.908 | 3 | 1 | 0.0E+00 | 0.0E+00 | 0.0E+00 | 0.0E+00 | 0.0E+00 | 0.0E+00 | 0.0E+00 | 0.0E+00 | 0.0E+00 | 0.0E+00 | 0.0E+00 | 0.0E+00 |
| 6321 | 3 | A6NHP3;P0 | A6NHP3 | Speedy proteSPDYE2B | Homo sapien | 48.292 | 1 | 1 | 0.0E+00 | 0.0E+00 | 0.0E+00 | 0.0E+00 | 0.0E+00 | 0.0E+00 | 0.0E+00 | 0.0E+00 | 0.0E+00 | 0.0E+00 | 0.0E+00 | 0.0E+00 |
| 6322 | 1 | A6NI56 | A6NI56 | Coiled-coil doCCDC154 | Homo sapien | 75.41 | 1 | 1 | 0.0E+00 | 0.0E+00 | 1.2E+04 | 8.6E+03 | 9.0E+03 | 7.8E+03 | 0.0E+00 | 0.0E+00 | 0.0E+00 | 0.0E+00 | 0.0E+00 | 0.0E+00 |
| 6323 | 1 | A6NIH7 | A6NIH7 | Protein unc-1UNC119B | Homo sapien | 28.137 | 1 | 1 | 7.8E+03 | 7.9E+03 | 6.6E+03 | 6.5E+03 | 8.4E+03 | 9.5E+03 | 6.7E+03 | 7.4E+03 | 6.6E+03 | 7.1E+03 | 8.5E+03 | 9.4E+03 |
| 6324 | 1 | A6NJ69 | A6NJ69 | IgA-inducing IGIP | Homo sapien | 5.936 | 1 | 1 | 0.0E+00 | 0.0E+00 | 0.0E+00 | 0.0E+00 | 0.0E+00 | 0.0E+00 | 2.0E+05 | 3.5E+05 | 0.0E+00 | 0.0E+00 | 3.5E+05 | 2.3E+05 |
| 6325 | 1 | A6NJ78 | A6NJ78 | 12S rRNA N(METTL15 | Homo sapien | 46.123 | 2 | 1 | 0.0E+00 | 0.0E+00 | 0.0E+00 | 0.0E+00 | 0.0E+00 | 0.0E+00 | 0.0E+00 | 0.0E+00 | 0.0E+00 | 0.0E+00 | 0.0E+00 | 0.0E+00 |
| 6326 | 1 | A6NK02 | A6NK02 | Tripartite motTRIM75 | Homo sapien | 53.677 | 1 | 1 | 0.0E+00 | 0.0E+00 | 2.3E+03 | 1.6E+03 | 1.2E+03 | 0.0E+00 | 0.0E+00 | 8.7E+03 | 0.0E+00 | 0.0E+00 | 8.0E+03 | 0.0E+00 |
| 6327 | 1 | A8MSI8 | A8MSI8 | LYR motif-coLYRM9 | Homo sapien | 9.37 | 1 | 1 | 1.7E+04 | 1.3E+04 | 0.0E+00 | 1.6E+04 | 0.0E+00 | 0.0E+00 | 0.0E+00 | 0.0E+00 | 0.0E+00 | 0.0E+00 | 0.0E+00 | 0.0E+00 |
| 6328 | 1 | A8MW92 | A8MW92 | PHD finger prPHF20L1 | Homo sapien | 115.011 | 1 | 1 | 2.3E+06 | 2.7E+06 | 0.0E+00 | 2.3E+06 | 3.3E+06 | 2.8E+06 | 3.3E+06 | 0.0E+00 | 2.8E+06 | 2.9E+06 | 3.2E+06 | 2.6E+06 |
| 6329 | 2 | A8MWD9;P6 | A8MWD9 | Putative smaSNRPGP15 | Homo sapien | 8.541 | 1 | 1 | 8.8E+05 | 5.8E+05 | 9.9E+05 | 6.6E+05 | 7.1E+05 | 5.4E+05 | 7.0E+05 | 6.9E+05 | 8.2E+05 | 9.1E+05 | 7.1E+05 | 5.4E+05 |
| 6330 | 1 | B0YJ81 | B0YJ81 | Very-long-chHACD1 | Homo sapien | 32.389 | 1 | 1 | 0.0E+00 | 0.0E+00 | 0.0E+00 | 0.0E+00 | 0.0E+00 | 0.0E+00 | 0.0E+00 | 0.0E+00 | 0.0E+00 | 0.0E+00 | 0.0E+00 | 0.0E+00 |
| 6331 | 3 | B7ZW38;O6 | B7ZW38 | HeterogeneoHNRNPCL3 | Homo sapien | 32.03 | 1 | 1 | 0.0E+00 | 0.0E+00 | 0.0E+00 | 0.0E+00 | 0.0E+00 | 0.0E+00 | 0.0E+00 | 6.7E+04 | 8.3E+04 | 0.0E+00 | 0.0E+00 | 0.0E+00 |
| 6332 | 4 | I6L899;A6N | I6L899 | Golgin subfaGOLGA8R | Homo sapien | 71.491 | 2 | 1 | 0.0E+00 | 0.0E+00 | 0.0E+00 | 0.0E+00 | 0.0E+00 | 0.0E+00 | 0.0E+00 | 0.0E+00 | 0.0E+00 | 0.0E+00 | 0.0E+00 | 0.0E+00 |
| 6333 | 1 | O00148 | O00148 | ATP-dependeDDX39A | Homo sapien | 49.129 | 1 | 1 | 5.3E+04 | 4.4E+04 | 5.0E+04 | 4.3E+04 | 4.8E+04 | 3.8E+04 | 4.1E+04 | 3.5E+04 | 3.7E+04 | 4.5E+04 | 7.3E+04 | 4.2E+04 |
| 6334 | 1 | O00193 | O00193 | Small acidic pSMAP | Homo sapien | 20.333 | 1 | 1 | 0.0E+00 | 0.0E+00 | 0.0E+00 | 0.0E+00 | 0.0E+00 | 0.0E+00 | 0.0E+00 | 3.0E+04 | 0.0E+00 | 0.0E+00 | 0.0E+00 | 0.0E+00 |
| 6335 | 1 | O00244 | O00244 | Copper transATOX1 | Homo sapien | 7.398 | 1 | 1 | 4.5E+04 | 6.3E+04 | 3.8E+04 | 2.6E+04 | 6.9E+04 | 5.0E+04 | 6.9E+04 | 3.3E+04 | 3.5E+04 | 4.8E+04 | 6.6E+04 | 4.6E+04 |
| 6336 | 1 | O00287 | O00287 | Regulatory faRFXAP | Homo sapien | 28.233 | 1 | 1 | 0.0E+00 | 0.0E+00 | 4.1E+04 | 0.0E+00 | 3.8E+04 | 0.0E+00 | 0.0E+00 | 0.0E+00 | 0.0E+00 | 0.0E+00 | 0.0E+00 | 0.0E+00 |
| 6337 | 1 | O00292 | O00292 | Left-right deteLEFTY2 | Homo sapien | 40.92 | 1 | 1 | 0.0E+00 | 9.0E+04 | 7.1E+04 | 0.0E+00 | 0.0E+00 | 0.0E+00 | 0.0E+00 | 0.0E+00 | 0.0E+00 | 1.5E+04 | 0.0E+00 | 2.5E+04 |
| 6338 | 1 | O00401 | O00401 | Actin nucleatiWASL | Homo sapien | 54.828 | 1 | 1 | 0.0E+00 | 0.0E+00 | 0.0E+00 | 0.0E+00 | 0.0E+00 | 0.0E+00 | 0.0E+00 | 0.0E+00 | 0.0E+00 | 0.0E+00 | 0.0E+00 | 0.0E+00 |
| 6339 | 1 | O00421 | O00421 | C-C chemokiCCRL2 | Homo sapien | 39.513 | 1 | 1 | 4.5E+04 | 4.6E+04 | 5.9E+04 | 7.4E+04 | 7.9E+04 | 1.1E+05 | 1.0E+05 | 8.9E+04 | 7.6E+04 | 9.4E+04 | 4.4E+04 | 1.1E+05 |

[illegible]

| | | | | | | | | | | | | | | | | |
|---|---|---|---|---|---|---|---|---|---|---|---|---|---|---|---|---|
| 6810 | 1 | Q15056 | Q15056 | Eukaryotic tr | EIF4H | Homo sapien | 27.385 | 1 | 1 | 1.9E+06 | 2.0E+06 | 2.1E+06 | 2.5E+06 | 2.0E+06 | 2.0E+06 | [illegible] |
| 6811 | 1 | Q15070 | Q15070 | Mitochondrial | OXA1L | Homo sapien | 48.521 | 1 | 1 | 0.0E+00 | 0.0E+00 | 0.0E+00 | 0.0E+00 | 4.9E+03 | 0.0E+00 | [illegible] |
| 6812 | 1 | Q15072 | Q15072 | Zinc finger pr | ZNF146 | Homo sapien | 33.306 | 1 | 1 | 0.0E+00 | 0.0E+00 | 0.0E+00 | 0.0E+00 | 0.0E+00 | 0.0E+00 | [illegible] |
| 6813 | 1 | Q15119 | Q15119 | [Pyruvate de | PDK2 | Homo sapien | 46.154 | 1 | 1 | 0.0E+00 | 0.0E+00 | 0.0E+00 | 0.0E+00 | 0.0E+00 | 0.0E+00 | [illegible] |
| 6814 | 1 | Q15155 | Q15155 | BOS complex | NOMO1 | Homo sapien | 134.323 | 17 | 1 | 0.0E+00 | 0.0E+00 | 0.0E+00 | 0.0E+00 | 0.0E+00 | 0.0E+00 | [illegible] |
| 6815 | 1 | Q15223 | Q15223 | Nectin-1 | NECTIN1 | Homo sapien | 57.159 | 1 | 1 | 0.0E+00 | 0.0E+00 | 0.0E+00 | 0.0E+00 | 0.0E+00 | 4.4E+04 | [illegible] |
| 6816 | 1 | Q15287 | Q15287 | RNA-binding | RNPS1 | Homo sapien | 34.209 | 1 | 1 | 0.0E+00 | 0.0E+00 | 4.9E+04 | 9.7E+04 | 5.2E+04 | 5.3E+04 | [illegible] |
| 6817 | 1 | Q15388 | Q15388 | Mitochondrial | TOMM20 | Homo sapien | 16.295 | 1 | 1 | 5.6E+05 | 2.8E+05 | 4.5E+05 | 0.0E+00 | 0.0E+00 | 4.4E+05 | [illegible] |
| 6818 | 1 | Q15398 | Q15398 | Disks large-a | DLGAP5 | Homo sapien | 95.117 | 1 | 1 | 0.0E+00 | 0.0E+00 | 0.0E+00 | 0.0E+00 | 0.0E+00 | 0.0E+00 | [illegible] |
| 6819 | 1 | Q15427 | Q15427 | Splicing facto | SF3B4 | Homo sapien | 44.386 | 1 | 1 | 3.7E+04 | 0.0E+00 | 2.6E+04 | 5.4E+04 | 0.0E+00 | 0.0E+00 | [illegible] |
| 6820 | 1 | Q15517 | Q15517 | Corneodesm | CDSN | Homo sapien | 51.607 | 1 | 1 | 0.0E+00 | 0.0E+00 | 0.0E+00 | 0.0E+00 | 1.0E+05 | 6.2E+04 | [illegible] |
| 6821 | 1 | Q15543 | Q15543 | Transcription | TAF13 | Homo sapien | 14.287 | 1 | 1 | 2.9E+03 | 0.0E+00 | 3.0E+03 | 0.0E+00 | 3.4E+03 | 3.3E+03 | [illegible] |
| 6822 | 2 | Q15583;Q9[illegible] | Q15583 | Homeobox pr | TGIF1 | Homo sapien | 43.013 | 1 | 1 | 0.0E+00 | 0.0E+00 | 0.0E+00 | 0.0E+00 | 0.0E+00 | 0.0E+00 | [illegible] |
| 6823 | 1 | Q15723 | Q15723 | ETS-related t | ELF2 | Homo sapien | 63.967 | 1 | 1 | 0.0E+00 | 0.0E+00 | 0.0E+00 | 0.0E+00 | 2.1E+04 | 0.0E+00 | [illegible] |
| 6824 | 1 | Q15788 | Q15788 | Nuclear rece | NCOA1 | Homo sapien | 156.759 | 1 | 1 | 0.0E+00 | 0.0E+00 | 0.0E+00 | 0.0E+00 | 0.0E+00 | 0.0E+00 | [illegible] |
| 6825 | 1 | Q15847 | Q15847 | Adipogenesis | ADIRF | Homo sapien | 7.851 | 1 | 1 | 0.0E+00 | 0.0E+00 | 0.0E+00 | 0.0E+00 | 0.0E+00 | 0.0E+00 | [illegible] |
| 6826 | 1 | Q15906 | Q15906 | Vacuolar pro | VPS72 | Homo sapien | 40.595 | 1 | 1 | 1.7E+04 | 2.6E+04 | 1.8E+04 | 1.7E+04 | 2.4E+04 | 0.0E+00 | [illegible] |
| 6827 | 1 | Q15910 | Q15910 | Histone-lysin | EZH2 | Homo sapien | 85.366 | 1 | 1 | 0.0E+00 | 0.0E+00 | 0.0E+00 | 0.0E+00 | 0.0E+00 | 0.0E+00 | [illegible] |
| 6828 | 1 | Q16254 | Q16254 | Transcription | E2F4 | Homo sapien | 43.959 | 1 | 1 | 0.0E+00 | 7.4E+03 | 7.6E+03 | 6.9E+03 | 5.9E+03 | 0.0E+00 | [illegible] |
| 6829 | 1 | Q16514 | Q16514 | Transcription | TAF12 | Homo sapien | 17.922 | 1 | 1 | 3.5E+04 | 0.0E+00 | 3.3E+04 | 4.1E+04 | 3.6E+04 | 0.0E+00 | [illegible] |
| 6830 | 1 | Q16563 | Q16563 | Synaptophys | SYPL1 | Homo sapien | 28.565 | 1 | 1 | 3.1E+05 | 4.1E+05 | 3.8E+05 | 5.9E+05 | 6.3E+05 | 5.7E+05 | [illegible] |
| 6831 | 1 | Q16584 | Q16584 | Mitogen-activ | MAP3K11 | Homo sapien | 92.691 | 1 | 1 | 0.0E+00 | 0.0E+00 | 0.0E+00 | 0.0E+00 | 0.0E+00 | 0.0E+00 | [illegible] |
| 6832 | 1 | Q16594 | Q16594 | Transcription | TAF9 | Homo sapien | 28.973 | 4 | 1 | 0.0E+00 | 0.0E+00 | 0.0E+00 | 0.0E+00 | 7.8E+04 | 1.1E+05 | [illegible] |
| 6833 | 1 | Q16611 | Q16611 | Bcl-2 homolo | BAK1 | Homo sapien | 23.407 | 1 | 1 | 0.0E+00 | 0.0E+00 | 0.0E+00 | 0.0E+00 | 0.0E+00 | 0.0E+00 | [illegible] |
| 6834 | 1 | Q16635 | Q16635 | Tafazzin | TAFAZZIN | Homo sapien | 30.203 | 1 | 1 | 0.0E+00 | 0.0E+00 | 0.0E+00 | 0.0E+00 | 0.0E+00 | 0.0E+00 | [illegible] |
| 6835 | 4 | Q16695;P68[illegible] | Q16695 | Histone H3.1 | H3-4 | Homo sapien | 15.506 | 2 | 1 | 2.8E+04 | 3.0E+04 | 0.0E+00 | 0.0E+00 | 2.5E+04 | 2.7E+04 | [illegible] |
| 6836 | 1 | Q16719 | Q16719 | Kynureninase | KYNU | Homo sapien | 52.351 | 1 | 1 | 0.0E+00 | 0.0E+00 | 0.0E+00 | 0.0E+00 | 9.7E+03 | 0.0E+00 | [illegible] |
| 6837 | 1 | Q16740 | Q16740 | ATP-depend | CLPP | Homo sapien | 30.179 | 1 | 1 | 0.0E+00 | 0.0E+00 | 0.0E+00 | 0.0E+00 | 0.0E+00 | 0.0E+00 | [illegible] |
| 6838 | 1 | Q16773 | Q16773 | Kynurenine-- | KYAT1 | Homo sapien | 47.873 | 1 | 1 | 0.0E+00 | 0.0E+00 | 0.0E+00 | 0.0E+00 | 0.0E+00 | 0.0E+00 | [illegible] |
| 6839 | 1 | Q16819 | Q16819 | Meprin A sub | MEP1A | Homo sapien | 84.422 | 1 | 1 | 0.0E+00 | 0.0E+00 | 0.0E+00 | 0.0E+00 | 0.0E+00 | 0.0E+00 | [illegible] |
| 6840 | 1 | Q16850 | Q16850 | Lanosterol 14 | CYP51A1 | Homo sapien | 57.277 | 1 | 1 | 0.0E+00 | 0.0E+00 | 0.0E+00 | 0.0E+00 | 0.0E+00 | 0.0E+00 | [illegible] |
| 6841 | 1 | Q17RY0 | Q17RY0 | Cytoplasmic | CPEB4 | Homo sapien | 80.152 | 2 | 1 | 0.0E+00 | 0.0E+00 | 0.0E+00 | 0.0E+00 | 0.0E+00 | 0.0E+00 | [illegible] |
| 6842 | 1 | Q2M1Z3 | Q2M1Z3 | Rho GTPase | ARHGAP31 | Homo sapien | 156.988 | 1 | 1 | 0.0E+00 | 0.0E+00 | 0.0E+00 | 0.0E+00 | 0.0E+00 | 0.0E+00 | [illegible] |
| 6843 | 1 | Q2TB10 | Q2TB10 | Zinc finger pr | ZNF800 | Homo sapien | 75.237 | 1 | 1 | 9.3E+03 | 0.0E+00 | 0.0E+00 | 0.0E+00 | 8.1E+03 | 0.0E+00 | [illegible] |
| 6844 | 1 | Q2VY69 | Q2VY69 | Zinc finger pr | ZNF284 | Homo sapien | 69.019 | 2 | 1 | 0.0E+00 | 0.0E+00 | 0.0E+00 | 0.0E+00 | 0.0E+00 | 0.0E+00 | [illegible] |
| 6845 | 1 | Q3B7J2 | Q3B7J2 | Glucose-fruc | GFOD2 | Homo sapien | 42.255 | 1 | 1 | 0.0E+00 | 0.0E+00 | 0.0E+00 | 0.0E+00 | 0.0E+00 | 0.0E+00 | [illegible] |
| 6846 | 1 | Q3KP31 | Q3KP31 | Zinc finger pr | ZNF791 | Homo sapien | 66.873 | 1 | 1 | 0.0E+00 | 0.0E+00 | 0.0E+00 | 0.0E+00 | 0.0E+00 | 0.0E+00 | [illegible] |
| 6847 | 1 | Q3KRA6 | Q3KRA6 | UPF0538 pro | C2orf76 | Homo sapien | 14.606 | 1 | 1 | 0.0E+00 | 0.0E+00 | 0.0E+00 | 0.0E+00 | 0.0E+00 | 0.0E+00 | [illegible] |
| 6848 | 1 | Q3SXY8 | Q3SXY8 | ADP-ribosyla | ARL13B | Homo sapien | 48.644 | 1 | 1 | 0.0E+00 | 0.0E+00 | 0.0E+00 | 0.0E+00 | 0.0E+00 | 0.0E+00 | [illegible] |
| 6849 | 1 | Q3SY69 | Q3SY69 | Mitochondrial | ALDH1L2 | Homo sapien | 101.747 | 1 | 1 | 0.0E+00 | 0.0E+00 | 0.0E+00 | 0.0E+00 | 0.0E+00 | 0.0E+00 | [illegible] |
| 6850 | 1 | Q3ZAQ7 | Q3ZAQ7 | Vacuolar ATF | VMA21 | Homo sapien | 11.353 | 1 | 1 | 2.6E+05 | 3.1E+05 | 3.5E+05 | 3.9E+05 | 3.9E+05 | 2.3E+05 | [illegible] |
| 6851 | 1 | Q4G0I0 | Q4G0I0 | Ubiquinol-cyt | UQCC4 | Homo sapien | 15.003 | 1 | 1 | 0.0E+00 | 0.0E+00 | 1.5E+06 | 0.0E+00 | 9.4E+05 | 0.0E+00 | [illegible] |
| 6852 | 3 | Q4VC05;Q9[illegible] | Q4VC05 | B-cell CLL/lyr | BCL7A | Homo sapien | 22.812 | 1 | 1 | 1.5E+05 | 1.6E+05 | 2.1E+05 | 9.1E+04 | 2.4E+05 | 0.0E+00 | [illegible] |
| 6853 | 1 | Q4ZHG4 | Q4ZHG4 | Fibronectin ty | FNDC1 | Homo sapien | 205.561 | 1 | 1 | 0.0E+00 | 0.0E+00 | 0.0E+00 | 0.0E+00 | 0.0E+00 | 0.0E+00 | [illegible] |
| 6854 | 1 | Q53EL9 | Q53EL9 | Seizure prote | SEZ6 | Homo sapien | 107.426 | 1 | 1 | 0.0E+00 | 0.0E+00 | 0.0E+00 | 0.0E+00 | 0.0E+00 | 0.0E+00 | [illegible] |
| 6855 | 3 | Q53FV1;Q8[illegible] | Q53FV1 | ORM1-like pr | ORMDL2 | Homo sapien | 17.364 | 1 | 1 | 0.0E+00 | 1.9E+05 | 0.0E+00 | 5.4E+04 | 0.0E+00 | 2.1E+05 | [illegible] |
| 6856 | 1 | Q53RT3 | Q53RT3 | Retroviral-lik | ASPRV1 | Homo sapien | 36.99 | 1 | 1 | 0.0E+00 | 4.0E+04 | 0.0E+00 | 0.0E+00 | 0.0E+00 | 9.7E+04 | [illegible] |
| 6857 | 1 | Q562R1 | Q562R1 | Beta-actin-lik | ACTBL2 | Homo sapien | 42.003 | 2 | 1 | 0.0E+00 | 0.0E+00 | 6.1E+03 | 0.0E+00 | 0.0E+00 | 0.0E+00 | [illegible] |
| 6858 | 1 | Q587I9 | Q587I9 | Vesicle trans | SFT2D3 | Homo sapien | 21.79 | 1 | 1 | 3.2E+04 | 2.8E+04 | 2.3E+04 | 2.0E+04 | 2.3E+04 | 2.5E+04 | [illegible] |

[illegible]

| | | | | | | | | | | | | | | | | | | | | |
|---|---|---|---|---|---|---|---|---|---|---|---|---|---|---|---|---|---|---|---|---|
| 7334 | 1 | Q9NRX5 | Q9NRX5 | Serine incorp | SERINC1 | Homo sapien | 50.493 | 1 | 1 | 0.0E+00 | 0.0E+00 | 0.0E+00 | 0.0E+00 | 1.8E+06 | 2.0E+06 | 0.0E+00 | 0.0E+00 | 0.0E+00 | 0.0E+00 | [illegible] |
| 7335 | 1 | Q9NRY2 | Q9NRY2 | SOSS compl | INIP | Homo sapien | 11.423 | 1 | 1 | 3.0E+04 | 0.0E+00 | 4.2E+04 | 0.0E+00 | 5.0E+04 | 0.0E+00 | 0.0E+00 | 0.0E+00 | 0.0E+00 | 7.6E+04 | [illegible] |
| 7336 | 1 | Q9NS71 | Q9NS71 | Gastrokine-1 | GKN1 | Homo sapien | 20.33 | 1 | 1 | 0.0E+00 | 0.0E+00 | 0.0E+00 | 0.0E+00 | 0.0E+00 | 0.0E+00 | 0.0E+00 | 0.0E+00 | 0.0E+00 | 0.0E+00 | [illegible] |
| 7337 | 1 | Q9NSJ1 | Q9NSJ1 | Putative zinc | ZNF355P | Homo sapien | 49.69 | 2 | 1 | 0.0E+00 | 0.0E+00 | 0.0E+00 | 0.0E+00 | 0.0E+00 | 0.0E+00 | 0.0E+00 | 0.0E+00 | 0.0E+00 | 0.0E+00 | [illegible] |
| 7338 | 1 | Q9NSK7 | Q9NSK7 | Protein C19o | C19orf12 | Homo sapien | 15.005 | 1 | 1 | 0.0E+00 | 0.0E+00 | 0.0E+00 | 0.0E+00 | 0.0E+00 | 0.0E+00 | 0.0E+00 | 0.0E+00 | 0.0E+00 | 0.0E+00 | [illegible] |
| 7339 | 1 | Q9NSY2 | Q9NSY2 | StAR-related | STARD5 | Homo sapien | 23.795 | 1 | 1 | 0.0E+00 | 0.0E+00 | 0.0E+00 | 0.0E+00 | 0.0E+00 | 0.0E+00 | 8.6E+05 | 0.0E+00 | 8.5E+05 | 0.0E+00 | [illegible] |
| 7340 | 1 | Q9NUG6 | Q9NUG6 | p53 and DNA | PDRG1 | Homo sapien | 15.51 | 1 | 1 | 0.0E+00 | 0.0E+00 | 0.0E+00 | 0.0E+00 | 0.0E+00 | 0.0E+00 | 0.0E+00 | 0.0E+00 | 0.0E+00 | 0.0E+00 | [illegible] |
| 7341 | 1 | Q9NUL5 | Q9NUL5 | Shiftless anti | SHFL | Homo sapien | 33.11 | 1 | 1 | 6.5E+05 | 4.3E+05 | 0.0E+00 | 4.6E+05 | 0.0E+00 | 5.6E+05 | 0.0E+00 | 0.0E+00 | 0.0E+00 | 0.0E+00 | [illegible] |
| 7342 | 1 | Q9NUM3 | Q9NUM3 | Zinc transpor | SLC39A9 | Homo sapien | 32.253 | 1 | 1 | 0.0E+00 | 0.0E+00 | 0.0E+00 | 0.0E+00 | 0.0E+00 | 0.0E+00 | 0.0E+00 | 0.0E+00 | 0.0E+00 | 0.0E+00 | [illegible] |
| 7343 | 1 | Q9NUP7 | Q9NUP7 | tRNA:m(4)X r | TRMT13 | Homo sapien | 54.247 | 1 | 1 | 0.0E+00 | 0.0E+00 | 0.0E+00 | 0.0E+00 | 7.4E+04 | 0.0E+00 | 0.0E+00 | 0.0E+00 | 0.0E+00 | 0.0E+00 | [illegible] |
| 7344 | 1 | Q9NUP9 | Q9NUP9 | Protein lin-7 h | LIN7C | Homo sapien | 21.833 | 5 | 1 | 2.2E+06 | 2.3E+06 | 1.1E+06 | 1.6E+06 | 1.6E+06 | 1.9E+06 | 2.1E+06 | 1.2E+06 | 1.6E+06 | 1.2E+06 | [illegible] |
| 7345 | 1 | Q9NV56 | Q9NV56 | MRG/MORF4 | MRGBP | Homo sapien | 22.416 | 1 | 1 | 0.0E+00 | 0.0E+00 | 0.0E+00 | 0.0E+00 | 0.0E+00 | 0.0E+00 | 0.0E+00 | 0.0E+00 | 0.0E+00 | 0.0E+00 | [illegible] |
| 7346 | 1 | Q9NVE5 | Q9NVE5 | Ubiquitin carb | USP40 | Homo sapien | 140.132 | 1 | 1 | 0.0E+00 | 0.0E+00 | 0.0E+00 | 0.0E+00 | 0.0E+00 | 0.0E+00 | 0.0E+00 | 0.0E+00 | 0.0E+00 | 0.0E+00 | [illegible] |
| 7347 | 1 | Q9NVS2 | Q9NVS2 | Large ribosor | MRPS18A | Homo sapien | 22.184 | 1 | 1 | 5.9E+03 | 0.0E+00 | 0.0E+00 | 0.0E+00 | 0.0E+00 | 0.0E+00 | 0.0E+00 | 0.0E+00 | 0.0E+00 | 0.0E+00 | [illegible] |
| 7348 | 1 | Q9NVV5 | Q9NVV5 | Androgen-ind | AIG1 | Homo sapien | 27.459 | 1 | 1 | 0.0E+00 | 0.0E+00 | 0.0E+00 | 0.0E+00 | 0.0E+00 | 0.0E+00 | 0.0E+00 | 0.0E+00 | 0.0E+00 | 0.0E+00 | [illegible] |
| 7349 | 1 | Q9NW81 | Q9NW81 | Distal membr | DMAC2 | Homo sapien | 29.267 | 1 | 1 | 0.0E+00 | 0.0E+00 | 5.0E+04 | 0.0E+00 | 0.0E+00 | 0.0E+00 | 0.0E+00 | 0.0E+00 | 0.0E+00 | 0.0E+00 | [illegible] |
| 7350 | 1 | Q9NWA0 | Q9NWA0 | Mediator of R | MED9 | Homo sapien | 16.401 | 1 | 1 | 0.0E+00 | 0.0E+00 | 0.0E+00 | 0.0E+00 | 0.0E+00 | 0.0E+00 | 0.0E+00 | 0.0E+00 | 0.0E+00 | 0.0E+00 | [illegible] |
| 7351 | 1 | Q9NWW6 | Q9NWW6 | Nicotinamide | NMRK1 | Homo sapien | 23.191 | 1 | 1 | 0.0E+00 | 0.0E+00 | 0.0E+00 | 0.0E+00 | 0.0E+00 | 0.0E+00 | 2.2E+04 | 2.8E+04 | 0.0E+00 | 3.1E+04 | [illegible] |
| 7352 | 1 | Q9NWX5 | Q9NWX5 | Ankyrin repea | ASB6 | Homo sapien | 47.138 | 1 | 1 | 0.0E+00 | 2.7E+03 | 2.6E+03 | 3.1E+03 | 3.4E+03 | 3.0E+03 | 0.0E+00 | 0.0E+00 | 0.0E+00 | 0.0E+00 | [illegible] |
| 7353 | 1 | Q9NWY4 | Q9NWY4 | Histone PAR | HPF1 | Homo sapien | 39.435 | 1 | 1 | 0.0E+00 | 0.0E+00 | 0.0E+00 | 0.0E+00 | 2.9E+04 | 0.0E+00 | 0.0E+00 | 0.0E+00 | 0.0E+00 | 0.0E+00 | [illegible] |
| 7354 | 1 | Q9NWZ8 | Q9NWZ8 | Gem-associa | GEMIN8 | Homo sapien | 28.637 | 1 | 1 | 0.0E+00 | 0.0E+00 | 0.0E+00 | 0.0E+00 | 0.0E+00 | 0.0E+00 | 0.0E+00 | 0.0E+00 | 0.0E+00 | 0.0E+00 | [illegible] |
| 7355 | 1 | Q9NX04 | Q9NX04 | AFG2-interac | AIRIM | Homo sapien | 23.371 | 1 | 1 | 0.0E+00 | 0.0E+00 | 0.0E+00 | 0.0E+00 | 4.7E+05 | 0.0E+00 | 0.0E+00 | 0.0E+00 | 0.0E+00 | 0.0E+00 | [illegible] |
| 7356 | 1 | Q9NX07 | Q9NX07 | tRNA seleno | TRNAU1AP | Homo sapien | 32.5 | 1 | 1 | 0.0E+00 | 0.0E+00 | 0.0E+00 | 0.0E+00 | 0.0E+00 | 0.0E+00 | 0.0E+00 | 0.0E+00 | 0.0E+00 | 0.0E+00 | [illegible] |
| 7357 | 1 | Q9NX24 | Q9NX24 | H/ACA ribon | NHP2 | Homo sapien | 17.2 | 1 | 1 | 0.0E+00 | 2.8E+05 | 0.0E+00 | 1.2E+05 | 0.0E+00 | 1.7E+05 | 1.3E+05 | 2.1E+05 | 0.0E+00 | 2.4E+05 | [illegible] |
| 7358 | 1 | Q9NY59 | Q9NY59 | Sphingomyeli | SMPD3 | Homo sapien | 71.08 | 1 | 1 | 0.0E+00 | 0.0E+00 | 0.0E+00 | 0.0E+00 | 1.7E+05 | 0.0E+00 | 0.0E+00 | 0.0E+00 | 0.0E+00 | 0.0E+00 | [illegible] |
| 7359 | 1 | Q9NYA1 | Q9NYA1 | Sphingosine k | SPHK1 | Homo sapien | 42.516 | 1 | 1 | 3.0E+04 | 3.2E+04 | 2.2E+04 | 3.0E+04 | 3.2E+04 | 2.9E+04 | 0.0E+00 | 0.0E+00 | 0.0E+00 | 0.0E+00 | [illegible] |
| 7360 | 1 | Q9NYF3 | Q9NYF3 | Protein FAM5 | FAM53C | Homo sapien | 43.09 | 1 | 1 | 0.0E+00 | 1.0E+05 | 0.0E+00 | 0.0E+00 | 0.0E+00 | 0.0E+00 | 0.0E+00 | 0.0E+00 | 0.0E+00 | 0.0E+00 | [illegible] |
| 7361 | 1 | Q9NYG2 | Q9NYG2 | Palmitoyltran | ZDHHC3 | Homo sapien | 34.171 | 1 | 1 | 1.9E+05 | 1.7E+05 | 1.4E+05 | 1.7E+05 | 2.4E+05 | 2.2E+05 | 3.7E+05 | 3.6E+05 | 3.8E+05 | 3.3E+05 | [illegible] |
| 7362 | 1 | Q9NYM9 | Q9NYM9 | BET1-like pro | BET1L | Homo sapien | 12.386 | 1 | 1 | 1.1E+06 | 1.1E+06 | 9.6E+05 | 8.6E+05 | 8.2E+05 | 0.0E+00 | 8.7E+05 | 7.9E+05 | 6.6E+05 | 8.6E+05 | [illegible] |
| 7363 | 1 | Q9NYZ1 | Q9NYZ1 | Golgi appara | TVP23B | Homo sapien | 23.577 | 3 | 1 | 0.0E+00 | 0.0E+00 | 7.1E+05 | 0.0E+00 | 6.6E+05 | 5.2E+05 | 0.0E+00 | 0.0E+00 | 0.0E+00 | 0.0E+00 | [illegible] |
| 7364 | 1 | Q9NYZ4 | Q9NYZ4 | Sialic acid-bir | SIGLEC8 | Homo sapien | 54.042 | 1 | 1 | 0.0E+00 | 0.0E+00 | 0.0E+00 | 0.0E+00 | 9.0E+04 | 0.0E+00 | 0.0E+00 | 0.0E+00 | 0.0E+00 | 0.0E+00 | [illegible] |
| 7365 | 1 | Q9NZ53 | Q9NZ53 | Podocalyxin-l | PODXL2 | Homo sapien | 65.077 | 1 | 1 | 0.0E+00 | 0.0E+00 | 0.0E+00 | 0.0E+00 | 0.0E+00 | 0.0E+00 | 0.0E+00 | 0.0E+00 | 0.0E+00 | 0.0E+00 | [illegible] |
| 7366 | 1 | Q9NZD8 | Q9NZD8 | Maspardin | SPG21 | Homo sapien | 34.959 | 1 | 1 | 0.0E+00 | 0.0E+00 | 0.0E+00 | 0.0E+00 | 0.0E+00 | 0.0E+00 | 0.0E+00 | 0.0E+00 | 4.6E+04 | 0.0E+00 | [illegible] |
| 7367 | 1 | Q9NZE8 | Q9NZE8 | Large ribosor | MRPL35 | Homo sapien | 21.514 | 1 | 1 | 0.0E+00 | 0.0E+00 | 0.0E+00 | 0.0E+00 | 1.3E+04 | 0.0E+00 | 1.4E+04 | 1.3E+04 | 0.0E+00 | 1.9E+04 | [illegible] |
| 7368 | 1 | Q9NZH8 | Q9NZH8 | Interleukin-36 | IL36G | Homo sapien | 18.722 | 1 | 1 | 0.0E+00 | 0.0E+00 | 0.0E+00 | 0.0E+00 | 0.0E+00 | 7.0E+03 | 0.0E+00 | 0.0E+00 | 0.0E+00 | 0.0E+00 | [illegible] |
| 7369 | 1 | Q9NZJ9 | Q9NZJ9 | Diphosphoino | NUDT4 | Homo sapien | 20.306 | 3 | 1 | 6.3E+05 | 5.2E+05 | 4.4E+05 | 6.0E+05 | 6.0E+05 | 4.8E+05 | 7.2E+05 | 6.8E+05 | 5.5E+05 | 7.3E+05 | [illegible] |
| 7370 | 1 | Q9NZM5 | Q9NZM5 | Ribosome bio | NOP53 | Homo sapien | 54.39 | 1 | 1 | 0.0E+00 | 1.8E+04 | 6.5E+03 | 0.0E+00 | 1.4E+04 | 0.0E+00 | 0.0E+00 | 0.0E+00 | 0.0E+00 | 0.0E+00 | [illegible] |
| 7371 | 1 | Q9NZU5 | Q9NZU5 | LIM and cyst | LMCD1 | Homo sapien | 40.834 | 1 | 1 | 0.0E+00 | 0.0E+00 | 0.0E+00 | 0.0E+00 | 0.0E+00 | 0.0E+00 | 0.0E+00 | 0.0E+00 | 0.0E+00 | 0.0E+00 | [illegible] |
| 7372 | 1 | Q9NZZ3 | Q9NZZ3 | Charged mult | CHMP5 | Homo sapien | 24.57 | 1 | 1 | 2.3E+05 | 0.0E+00 | 1.9E+05 | 3.7E+05 | 3.1E+05 | 4.0E+05 | 9.0E+05 | 5.7E+05 | 0.0E+00 | 4.6E+05 | [illegible] |
| 7373 | 1 | Q9P021 | Q9P021 | Cysteine-rich | CRIPT | Homo sapien | 11.213 | 1 | 1 | 5.5E+04 | 4.0E+04 | 8.3E+04 | 5.2E+04 | 4.4E+04 | 4.5E+04 | 2.5E+04 | 4.5E+04 | 2.5E+04 | 3.3E+04 | [illegible] |
| 7374 | 1 | Q9P0N8 | Q9P0N8 | E3 ubiquitin-p | MARCHF2 | Homo sapien | 26.995 | 1 | 1 | 0.0E+00 | 0.0E+00 | 6.0E+03 | 8.8E+03 | 5.7E+03 | 0.0E+00 | 0.0E+00 | 0.0E+00 | 0.0E+00 | 1.2E+04 | [illegible] |
| 7375 | 1 | Q9P0R6 | Q9P0R6 | GSK3B-intera | GSKIP | Homo sapien | 15.647 | 1 | 1 | 0.0E+00 | 0.0E+00 | 0.0E+00 | 0.0E+00 | 0.0E+00 | 0.0E+00 | 5.5E+06 | 0.0E+00 | 3.6E+06 | 5.3E+06 | [illegible] |
| 7376 | 1 | Q9P0S2 | Q9P0S2 | Cytochrome c | COX16 | Homo sapien | 12.293 | 1 | 1 | 1.9E+05 | 0.0E+00 | 0.0E+00 | 1.5E+05 | 0.0E+00 | 0.0E+00 | 0.0E+00 | 1.9E+05 | 1.7E+05 | 0.0E+00 | [illegible] |
| 7377 | 1 | Q9P0T4 | Q9P0T4 | Zinc finger pr | ZNF581 | Homo sapien | 22.009 | 1 | 1 | 0.0E+00 | 0.0E+00 | 0.0E+00 | 1.9E+05 | 0.0E+00 | 1.9E+05 | 0.0E+00 | 0.0E+00 | 0.0E+00 | 0.0E+00 | [illegible] |
| 7378 | 1 | Q9P109 | Q9P109 | Beta-1,3-gala | GCNT4 | Homo sapien | 53.052 | 1 | 1 | 0.0E+00 | 0.0E+00 | 0.0E+00 | 0.0E+00 | 0.0E+00 | 0.0E+00 | 0.0E+00 | 0.0E+00 | 0.0E+00 | 0.0E+00 | [illegible] |
| 7379 | 1 | Q9P203 | Q9P203 | BTB/POZ dor | BTBD7 | Homo sapien | 126.369 | 1 | 1 | 0.0E+00 | 0.0E+00 | 0.0E+00 | 0.0E+00 | 8.6E+03 | 6.5E+03 | 0.0E+00 | 0.0E+00 | 0.0E+00 | 0.0E+00 | [illegible] |
| 7380 | 1 | Q9P2G1 | Q9P2G1 | Ankyrin repea | ANKIB1 | Homo sapien | 122.004 | 1 | 1 | 0.0E+00 | 0.0E+00 | 0.0E+00 | 0.0E+00 | 0.0E+00 | 0.0E+00 | 0.0E+00 | 0.0E+00 | 0.0E+00 | 0.0E+00 | [illegible] |
| 7381 | 1 | Q9P2G3 | Q9P2G3 | Kelch-like pro | KLHL14 | Homo sapien | 70.715 | 1 | 1 | 1.8E+04 | 2.3E+04 | 2.1E+04 | 1.1E+04 | 9.3E+03 | 1.2E+04 | 1.4E+04 | 9.7E+03 | 8.6E+03 | 9.2E+03 | [illegible] |
| 7382 | 1 | Q9P2K6 | Q9P2K6 | Kelch-like pro | KLHL42 | Homo sapien | 56.869 | 1 | 1 | 0.0E+00 | 0.0E+00 | 0.0E+00 | 0.0E+00 | 0.0E+00 | 0.0E+00 | 0.0E+00 | 0.0E+00 | 0.0E+00 | 0.0E+00 | [illegible] |
| 7383 | 1 | Q9P2X0 | Q9P2X0 | Dolichol-phos | DPM3 | Homo sapien | 10.092 | 1 | 1 | 4.4E+04 | 6.2E+04 | 5.5E+04 | 7.0E+04 | 7.8E+04 | 5.8E+04 | 5.4E+04 | 9.2E+04 | 8.8E+04 | 1.2E+05 | [illegible] |
| 7384 | 1 | Q9UBD6 | Q9UBD6 | Ammonium tr | RHCG | Homo sapien | 53.176 | 1 | 1 | 0.0E+00 | 0.0E+00 | 0.0E+00 | 0.0E+00 | 0.0E+00 | 0.0E+00 | 0.0E+00 | 0.0E+00 | 0.0E+00 | 0.0E+00 | [illegible] |
| 7385 | 1 | Q9UBM7 | Q9UBM7 | 7-dehydroch | DHCR7 | Homo sapien | 54.49 | 1 | 1 | 1.6E+05 | 1.2E+05 | 1.3E+05 | 1.3E+05 | 1.3E+05 | 8.0E+04 | 0.0E+00 | 0.0E+00 | 1.6E+05 | 2.3E+05 | [illegible] |
| 7386 | 1 | Q9UBP4 | Q9UBP4 | Dickkopf-rela | DKK3 | Homo sapien | 38.39 | 1 | 1 | 2.9E+04 | 5.9E+04 | 3.9E+04 | 3.0E+04 | 3.9E+04 | 0.0E+00 | 6.7E+04 | 0.0E+00 | 4.3E+04 | 7.0E+04 | [illegible] |
| 7387 | 1 | Q9UBQ6 | Q9UBQ6 | Exostosin-lik | EXTL2 | Homo sapien | 37.464 | 1 | 1 | 0.0E+00 | 0.0E+00 | 0.0E+00 | 0.0E+00 | 0.0E+00 | 0.0E+00 | 0.0E+00 | 2.0E+05 | 0.0E+00 | 0.0E+00 | [illegible] |
| 7388 | 1 | Q9UBU8 | Q9UBU8 | Mortality fact | MORF4L1 | Homo sapien | 41.471 | 1 | 1 | 0.0E+00 | 0.0E+00 | 0.0E+00 | 0.0E+00 | 0.0E+00 | 0.0E+00 | 0.0E+00 | 0.0E+00 | 0.0E+00 | 0.0E+00 | [illegible] |
| 7389 | 1 | Q9UBV7 | Q9UBV7 | Beta-1,4-gala | B4GALT7 | Homo sapien | 37.405 | 1 | 1 | 0.0E+00 | 2.6E+04 | 0.0E+00 | 1.4E+04 | 0.0E+00 | 0.0E+00 | 0.0E+00 | 1.0E+04 | 0.0E+00 | 0.0E+00 | [illegible] |
| 7390 | 1 | Q9UBY8 | Q9UBY8 | Protein CLN8 | CLN8 | Homo sapien | 32.787 | 1 | 1 | 0.0E+00 | 4.8E+05 | 0.0E+00 | 5.3E+05 | 3.9E+05 | 2.8E+05 | 0.0E+00 | 0.0E+00 | 0.0E+00 | 0.0E+00 | [illegible] |
| 7391 | 1 | Q9UDX5 | Q9UDX5 | Mitochondria | MTFP1 | Homo sapien | 18.008 | 1 | 1 | 0.0E+00 | 0.0E+00 | 5.9E+05 | 0.0E+00 | 4.7E+05 | 2.0E+05 | 0.0E+00 | 0.0E+00 | 0.0E+00 | 0.0E+00 | [illegible] |
| 7392 | 1 | Q9UET6 | Q9UET6 | tRNA (cytidin | FTSJ1 | Homo sapien | 36.079 | 1 | 1 | 6.3E+04 | 1.5E+05 | 4.8E+04 | 0.0E+00 | 7.2E+04 | 0.0E+00 | 0.0E+00 | 0.0E+00 | 0.0E+00 | 6.2E+04 | [illegible] |
| 7393 | 1 | Q9UEW3 | Q9UEW3 | Macrophage | MARCO | Homo sapien | 52.656 | 1 | 1 | 0.0E+00 | 0.0E+00 | 0.0E+00 | 0.0E+00 | 0.0E+00 | 0.0E+00 | 0.0E+00 | 0.0E+00 | 0.0E+00 | 0.0E+00 | [illegible] |
| 7394 | 1 | Q9UFG5 | Q9UFG5 | UPF0449 pro | C19orf25 | Homo sapien | 12.874 | 1 | 1 | 0.0E+00 | 0.0E+00 | 0.0E+00 | 0.0E+00 | 0.0E+00 | 0.0E+00 | 0.0E+00 | 0.0E+00 | 0.0E+00 | 0.0E+00 | [illegible] |
| 7395 | 1 | Q9UGM3 | Q9UGM3 | Scavenger re | DMBT1 | Homo sapien | 260.74 | 1 | 1 | 0.0E+00 | 0.0E+00 | 0.0E+00 | 0.0E+00 | 0.0E+00 | 0.0E+00 | 0.0E+00 | 0.0E+00 | 0.0E+00 | 0.0E+00 | [illegible] |
| 7396 | 1 | Q9UH17 | Q9UH17 | DNA dC->dU | APOBEC3B | Homo sapien | 45.925 | 4 | 1 | 0.0E+00 | 0.0E+00 | 0.0E+00 | 0.0E+00 | 0.0E+00 | 0.0E+00 | 0.0E+00 | 0.0E+00 | 0.0E+00 | 0.0E+00 | [illegible] |
| 7397 | 1 | Q9UH73 | Q9UH73 | Transcription | EBF1 | Homo sapien | 64.466 | 2 | 1 | 0.0E+00 | 0.0E+00 | 0.0E+00 | 0.0E+00 | 0.0E+00 | 0.0E+00 | 0.0E+00 | 0.0E+00 | 0.0E+00 | 0.0E+00 | [illegible] |
| 7398 | 1 | Q9UHB4 | Q9UHB4 | NADPH-depe | NDOR1 | Homo sapien | 66.762 | 1 | 1 | 0.0E+00 | 0.0E+00 | 0.0E+00 | 0.0E+00 | 0.0E+00 | 0.0E+00 | 0.0E+00 | 0.0E+00 | 0.0E+00 | 0.0E+00 | [illegible] |
| 7399 | 1 | Q9UHD9 | Q9UHD9 | Ubiquilin-2 | UBQLN2 | Homo sapien | 65.697 | 1 | 1 | 0.0E+00 | 1.7E+04 | 0.0E+00 | 1.3E+04 | 0.0E+00 | 0.0E+00 | 0.0E+00 | 0.0E+00 | 0.0E+00 | 1.5E+04 | [illegible] |
| 7400 | 1 | Q9UHG2 | Q9UHG2 | ProSAAS | PCSK1N | Homo sapien | 27.369 | 1 | 1 | 0.0E+00 | 0.0E+00 | 0.0E+00 | 0.0E+00 | 0.0E+00 | 0.0E+00 | 0.0E+00 | 0.0E+00 | 0.0E+00 | 0.0E+00 | [illegible] |
| 7401 | 1 | Q9UHY8 | Q9UHY8 | Fasciculation | FEZ2 | Homo sapien | 39.669 | 1 | 1 | 1.7E+04 | 1.3E+04 | 1.6E+04 | 0.0E+00 | 1.3E+04 | 1.5E+04 | 9.4E+03 | 1.2E+04 | 0.0E+00 | 1.2E+04 | [illegible] |
| 7402 | 1 | Q9UI14 | Q9UI14 | Prenylated R | RABAC1 | Homo sapien | 20.646 | 1 | 1 | 0.0E+00 | 0.0E+00 | 0.0E+00 | 0.0E+00 | 0.0E+00 | 0.0E+00 | 0.0E+00 | 0.0E+00 | 0.0E+00 | 0.0E+00 | [illegible] |
| 7403 | 1 | Q9UI38 | Q9UI38 | Probable thre | PRSS50 | Homo sapien | 43.086 | 1 | 1 | 0.0E+00 | 0.0E+00 | 0.0E+00 | 0.0E+00 | 0.0E+00 | 0.0E+00 | 0.0E+00 | 0.0E+00 | 0.0E+00 | 0.0E+00 | [illegible] |
| 7404 | 1 | Q9UIF9 | Q9UIF9 | Bromodomain | BAZ2A | Homo sapien | 211.2 | 1 | 1 | 0.0E+00 | 0.0E+00 | 3.7E+03 | 0.0E+00 | 0.0E+00 | 0.0E+00 | 0.0E+00 | 0.0E+00 | 0.0E+00 | 0.0E+00 | [illegible] |
| 7405 | 1 | Q9UII2 | Q9UII2 | ATPase inhib | ATP5IF1 | Homo sapien | 12.248 | 1 | 1 | 0.0E+00 | 0.0E+00 | 0.0E+00 | 0.0E+00 | 0.0E+00 | 0.0E+00 | 0.0E+00 | 0.0E+00 | 0.0E+00 | 0.0E+00 | [illegible] |
| 7406 | 1 | Q9UIL1 | Q9UIL1 | Short coiled-c | SCOC | Homo sapien | 18.043 | 1 | 1 | 1.2E+04 | 2.1E+04 | 1.0E+04 | 8.7E+03 | 1.4E+04 | 1.1E+04 | 0.0E+00 | 0.0E+00 | 5.3E+04 | 0.0E+00 | [illegible] |
| 7407 | 1 | Q9UIV1 | Q9UIV1 | CCR4-NOT t | CNOT7 | Homo sapien | 32.744 | 1 | 1 | 0.0E+00 | 1.0E+05 | 1.3E+05 | 8.0E+04 | 1.5E+05 | 7.6E+04 | 0.0E+00 | 9.7E+04 | 1.1E+05 | 1.4E+05 | [illegible] |
| 7408 | 4 | Q9UJ98;P0C | Q9UJ98 | Putative STA | STAG3 | Homo sapien | 139.034 | 1 | 1 | 0.0E+00 | 0.0E+00 | 0.0E+00 | 0.0E+00 | 0.0E+00 | 0.0E+00 | 0.0E+00 | 0.0E+00 | 0.0E+00 | 0.0E+00 | [illegible] |
| 7409 | 1 | Q9UJF2 | Q9UJF2 | Ras GTPase | RASAL2 | Homo sapien | 128.559 | 2 | 1 | 0.0E+00 | 0.0E+00 | 0.0E+00 | 0.0E+00 | 0.0E+00 | 0.0E+00 | 0.0E+00 | 0.0E+00 | 0.0E+00 | 0.0E+00 | [illegible] |
| 7410 | 1 | Q9UJK0 | Q9UJK0 | 18S rRNA am | TSR3 | Homo sapien | 33.597 | 1 | 1 | 6.2E+03 | 1.1E+04 | 0.0E+00 | 0.0E+00 | 9.5E+03 | 9.5E+03 | 0.0E+00 | 5.4E+03 | 9.8E+03 | 1.0E+04 | [illegible] |
| 7411 | 1 | Q9UK05 | Q9UK05 | Growth/differ | GDF2 | Homo sapien | 47.322 | 1 | 1 | 0.0E+00 | 0.0E+00 | 0.0E+00 | 0.0E+00 | 0.0E+00 | 0.0E+00 | 1.8E+04 | 2.1E+04 | 0.0E+00 | 1.8E+04 | [illegible] |
| 7412 | 1 | Q9UK53 | Q9UK53 | Inhibitor of gr | ING1 | Homo sapien | 46.737 | 1 | 1 | 4.7E+03 | 0.0E+00 | 4.6E+03 | 3.9E+03 | 4.3E+03 | 4.7E+03 | 0.0E+00 | 0.0E+00 | 1.4E+04 | 0.0E+00 | [illegible] |
| 7413 | 1 | Q9UKI2 | Q9UKI2 | Cdc42 effect | CDC42EP3 | Homo sapien | 27.676 | 1 | 1 | 1.3E+06 | 1.8E+06 | 0.0E+00 | 1.2E+06 | 0.0E+00 | 0.0E+00 | 0.0E+00 | 0.0E+00 | 0.0E+00 | 0.0E+00 | [illegible] |
| 7414 | 1 | Q9UKY3 | Q9UKY3 | Putative inact | CES1P1 | Homo sapien | 30.679 | 1 | 1 | 2.2E+05 | 1.8E+05 | 1.7E+05 | 1.3E+05 | 1.4E+05 | 1.6E+05 | 1.7E+05 | 1.0E+05 | 1.6E+05 | 0.0E+00 | [illegible] |
| 7415 | 1 | Q9UL45 | Q9UL45 | Biogenesis of | BLOC1S6 | Homo sapien | 19.745 | 1 | 1 | 1.1E+05 | 1.5E+05 | 8.1E+04 | 1.2E+05 | 1.3E+05 | 1.8E+05 | 0.0E+00 | 0.0E+00 | 0.0E+00 | 0.0E+00 | [illegible] |
| 7416 | 1 | Q9ULR3 | Q9ULR3 | Protein phos | PPM1H | Homo sapien | 56.448 | 1 | 1 | 4.5E+03 | 0.0E+00 | 5.3E+03 | 8.4E+03 | 0.0E+00 | 0.0E+00 | 0.0E+00 | 0.0E+00 | 0.0E+00 | 0.0E+00 | [illegible] |
| 7417 | 1 | Q9ULW0 | Q9ULW0 | Targeting pro | TPX2 | Homo sapien | 85.655 | 1 | 1 | 0.0E+00 | 0.0E+00 | 0.0E+00 | 0.0E+00 | 0.0E+00 | 0.0E+00 | 0.0E+00 | 0.0E+00 | 0.0E+00 | 0.0E+00 | [illegible] |
| 7418 | 1 | Q9UM82 | Q9UM82 | Spermatogen | SPATA2 | Homo sapien | 58.428 | 1 | 1 | 0.0E+00 | 0.0E+00 | 1.1E+06 | 0.0E+00 | 0.0E+00 | 0.0E+00 | 0.0E+00 | 0.0E+00 | 0.0E+00 | 0.0E+00 | [illegible] |
| 7419 | 1 | Q9UMR2 | Q9UMR2 | ATP-depend | DDX19B | Homo sapien | 53.928 | 11 | 1 | 0.0E+00 | 0.0E+00 | 0.0E+00 | 1.9E+04 | 0.0E+00 | 0.0E+00 | 0.0E+00 | 0.0E+00 | 0.0E+00 | 0.0E+00 | [illegible] |
| 7420 | 1 | Q9UMS0 | Q9UMS0 | NFU1 iron-su | NFU1 | Homo sapien | 28.461 | 1 | 1 | 0.0E+00 | 0.0E+00 | 0.0E+00 | 0.0E+00 | 0.0E+00 | 0.0E+00 | 0.0E+00 | 0.0E+00 | 0.0E+00 | 0.0E+00 | [illegible] |
| 7421 | 1 | Q9UNL2 | Q9UNL2 | Translocon-a | SSR3 | Homo sapien | 21.079 | 1 | 1 | 0.0E+00 | 0.0E+00 | 0.0E+00 | 0.0E+00 | 0.0E+00 | 0.0E+00 | 1.1E+06 | 1.4E+06 | 1.2E+06 | 1.5E+06 | [illegible] |
| 7422 | 1 | Q9UNL4 | Q9UNL4 | Inhibitor of gr | ING4 | Homo sapien | 28.531 | 1 | 1 | 0.0E+00 | 0.0E+00 | 1.3E+04 | 0.0E+00 | 1.5E+04 | 0.0E+00 | 0.0E+00 | 0.0E+00 | 0.0E+00 | 0.0E+00 | [illegible] |
| 7423 | 1 | Q9UNX3 | Q9UNX3 | Ribosomal pr | RPL26L1 | Homo sapien | 17.255 | 1 | 1 | 0.0E+00 | 0.0E+00 | 0.0E+00 | 0.0E+00 | 0.0E+00 | 0.0E+00 | 0.0E+00 | 0.0E+00 | 0.0E+00 | 0.0E+00 | [illegible] |
| 7424 | 1 | Q9UP95 | Q9UP95 | Solute carrier | SLC12A4 | Homo sapien | 120.65 | 1 | 1 | 0.0E+00 | 0.0E+00 | 0.0E+00 | 0.0E+00 | 0.0E+00 | 0.0E+00 | 0.0E+00 | 0.0E+00 | 0.0E+00 | 0.0E+00 | [illegible] |
| 7425 | 1 | Q9UPR3 | Q9UPR3 | Nonsense-m | SMG5 | Homo sapien | 113.928 | 1 | 1 | 3.1E+04 | 3.7E+04 | 3.0E+04 | 2.7E+04 | 3.0E+04 | 2.1E+04 | 3.2E+04 | 0.0E+00 | 2.7E+04 | 0.0E+00 | [illegible] |
| 7426 | 1 | Q9UPT9 | Q9UPT9 | Ubiquitin carb | USP22 | Homo sapien | 59.961 | 2 | 1 | 0.0E+00 | 0.0E+00 | 0.0E+00 | 0.0E+00 | 0.0E+00 | 0.0E+00 | 0.0E+00 | 0.0E+00 | 0.0E+00 | 0.0E+00 | [illegible] |
| 7427 | 1 | Q9UPW0 | Q9UPW0 | Forkhead bo | FOXJ3 | Homo sapien | 68.96 | 1 | 1 | 0.0E+00 | 0.0E+00 | 0.0E+00 | 0.0E+00 | 0.0E+00 | 0.0E+00 | 0.0E+00 | 0.0E+00 | 0.0E+00 | 0.0E+00 | [illegible] |
| 7428 | 1 | Q9UPY8 | Q9UPY8 | Microtubule-a | MAPRE3 | Homo sapien | 31.983 | 1 | 1 | 0.0E+00 | 0.0E+00 | 6.7E+03 | 8.5E+03 | 0.0E+00 | 0.0E+00 | 0.0E+00 | 0.0E+00 | 0.0E+00 | 0.0E+00 | [illegible] |
| 7429 | 1 | Q9UQQ2 | Q9UQQ2 | SH2B adapte | SH2B3 | Homo sapien | 63.225 | 1 | 1 | 0.0E+00 | 0.0E+00 | 0.0E+00 | 0.0E+00 | 3.6E+04 | 0.0E+00 | 0.0E+00 | 0.0E+00 | 0.0E+00 | 0.0E+00 | [illegible] |
| 7430 | 1 | Q9UQR0 | Q9UQR0 | Sex comb on | SCML2 | Homo sapien | 77.257 | 1 | 1 | 8.2E+03 | 0.0E+00 | 1.1E+04 | 7.4E+03 | 0.0E+00 | 0.0E+00 | 0.0E+00 | 0.0E+00 | 0.0E+00 | 0.0E+00 | [illegible] |
| 7431 | 1 | Q9Y221 | Q9Y221 | 60S ribosom | NIP7 | Homo sapien | 20.463 | 1 | 1 | 0.0E+00 | 0.0E+00 | 0.0E+00 | 0.0E+00 | 9.9E+03 | 0.0E+00 | 0.0E+00 | 0.0E+00 | 0.0E+00 | 0.0E+00 | [illegible] |
| 7432 | 1 | Q9Y223 | Q9Y223 | Bifunctional U | GNE | Homo sapien | 79.275 | 1 | 1 | 0.0E+00 | 0.0E+00 | 0.0E+00 | 0.0E+00 | 0.0E+00 | 0.0E+00 | 0.0E+00 | 0.0E+00 | 0.0E+00 | 0.0E+00 | [illegible] |
| 7433 | 1 | Q9Y244 | Q9Y244 | Proteasome r | POMP | Homo sapien | 15.786 | 1 | 1 | 0.0E+00 | 0.0E+00 | 0.0E+00 | 0.0E+00 | 0.0E+00 | 0.0E+00 | 0.0E+00 | 0.0E+00 | 0.0E+00 | 0.0E+00 | [illegible] |
| 7434 | 1 | Q9Y247 | Q9Y247 | Protein FAM5 | FAM50B | Homo sapien | 38.708 | 1 | 1 | 0.0E+00 | 0.0E+00 | 0.0E+00 | 0.0E+00 | 0.0E+00 | 0.0E+00 | 0.0E+00 | 0.0E+00 | 0.0E+00 | 0.0E+00 | [illegible] |
| 7435 | 1 | Q9Y257 | Q9Y257 | Potassium ch | KCNK6 | Homo sapien | 33.745 | 1 | 1 | 2.8E+03 | 0.0E+00 | 0.0E+00 | 0.0E+00 | 0.0E+00 | 0.0E+00 | 0.0E+00 | 0.0E+00 | 0.0E+00 | 0.0E+00 | [illegible] |
| 7436 | 1 | Q9Y279 | Q9Y279 | V-set and imr | VSIG4 | Homo sapien | 43.987 | 1 | 1 | 0.0E+00 | 0.0E+00 | 0.0E+00 | 0.0E+00 | 0.0E+00 | 0.0E+00 | 0.0E+00 | 0.0E+00 | 0.0E+00 | 0.0E+00 | [illegible] |
| 7437 | 1 | Q9Y2B0 | Q9Y2B0 | Protein cano | CNPY2 | Homo sapien | 20.652 | 1 | 1 | 2.5E+04 | 3.4E+04 | 2.9E+04 | 6.2E+04 | 5.3E+04 | 5.0E+04 | 0.0E+00 | 0.0E+00 | 3.4E+04 | 5.3E+04 | [illegible] |
| 7438 | 1 | Q9Y2D2 | Q9Y2D2 | UDP-N-acety | SLC35A3 | Homo sapien | 35.983 | 1 | 1 | 0.0E+00 | 0.0E+00 | 5.4E+04 | 4.2E+04 | 3.0E+04 | 3.6E+04 | 2.0E+04 | 0.0E+00 | 2.6E+04 | 0.0E+00 | [illegible] |
| 7439 | 1 | Q9Y2D9 | Q9Y2D9 | Zinc finger pr | ZNF652 | Homo sapien | 69.745 | 2 | 1 | 1.3E+04 | 0.0E+00 | 1.1E+04 | 1.3E+04 | 8.0E+03 | 1.1E+04 | 0.0E+00 | 0.0E+00 | 0.0E+00 | 0.0E+00 | [illegible] |
| 7440 | 1 | Q9Y2G5 | Q9Y2G5 | GDP-fucose | POFUT2 | Homo sapien | 49.976 | 1 | 1 | 0.0E+00 | 0.0E+00 | 0.0E+00 | 0.0E+00 | 0.0E+00 | 0.0E+00 | 0.0E+00 | 0.0E+00 | 0.0E+00 | 0.0E+00 | [illegible] |
| 7441 | 1 | Q9Y2Q5 | Q9Y2Q5 | Ragulator co | LAMTOR2 | Homo sapien | 13.506 | 1 | 1 | 1.2E+04 | 8.7E+03 | 1.3E+04 | 6.5E+03 | 1.4E+04 | 6.1E+03 | 9.5E+03 | 7.6E+03 | 7.7E+03 | 7.1E+03 | [illegible] |
| 7442 | 1 | Q9Y2U5 | Q9Y2U5 | Mitogen-activ | MAP3K2 | Homo sapien | 69.741 | 1 | 1 | 0.0E+00 | 5.0E+04 | 4.8E+04 | 3.4E+04 | 5.0E+04 | 5.7E+04 | 4.7E+04 | 6.5E+04 | 7.5E+04 | 3.2E+04 | [illegible] |
| 7443 | 1 | Q9Y3C0 | Q9Y3C0 | WASH compl | WASHC3 | Homo sapien | 21.169 | 1 | 1 | 1.9E+04 | 0.0E+00 | 0.0E+00 | 1.5E+04 | 2.9E+04 | 1.9E+04 | 1.4E+04 | 0.0E+00 | 1.0E+04 | 1.7E+04 | [illegible] |
| 7444 | 1 | Q9Y3D3 | Q9Y3D3 | Small ribosor | MRPS16 | Homo sapien | 15.345 | 1 | 1 | 0.0E+00 | 0.0E+00 | 6.8E+05 | 0.0E+00 | 0.0E+00 | 0.0E+00 | 0.0E+00 | 0.0E+00 | 0.0E+00 | 4.4E+05 | [illegible] |
| 7445 | 1 | Q9Y3E2 | Q9Y3E2 | BolA-like prot | BOLA1 | Homo sapien | 14.287 | 1 | 1 | 0.0E+00 | 0.0E+00 | 0.0E+00 | 0.0E+00 | 0.0E+00 | 0.0E+00 | 4.6E+04 | 2.3E+04 | 0.0E+00 | 3.1E+04 | [illegible] |
| 7446 | 1 | Q9Y3E5 | Q9Y3E5 | Peptidyl-tRN | PTRH2 | Homo sapien | 19.193 | 1 | 1 | 3.1E+04 | 3.4E+04 | 3.3E+04 | 4.7E+04 | 2.6E+04 | 2.9E+04 | 5.1E+04 | 4.5E+04 | 4.1E+04 | 4.8E+04 | [illegible] |
| 7447 | 1 | Q9Y3P8 | Q9Y3P8 | Signaling thre | SIT1 | Homo sapien | 21.126 | 1 | 1 | 0.0E+00 | 0.0E+00 | 0.0E+00 | 0.0E+00 | 0.0E+00 | 0.0E+00 | 0.0E+00 | 0.0E+00 | 0.0E+00 | 0.0E+00 | [illegible] |
| 7448 | 1 | Q9Y3U8 | Q9Y3U8 | Large ribosor | RPL36 | Homo sapien | 12.251 | 1 | 1 | 1.7E+05 | 1.1E+05 | 2.1E+05 | 9.7E+04 | 2.2E+05 | 1.2E+05 | 1.6E+05 | 2.2E+05 | 1.6E+05 | 2.3E+05 | [illegible] |
| 7449 | 1 | Q9Y3V2 | Q9Y3V2 | RWD domain | RWDD3 | Homo sapien | 30.545 | 1 | 1 | 0.0E+00 | 0.0E+00 | 2.6E+03 | 0.0E+00 | 0.0E+00 | 0.0E+00 | 0.0E+00 | 0.0E+00 | 0.0E+00 | 0.0E+00 | [illegible] |
| 7450 | 1 | Q9Y421 | Q9Y421 | Protein FAM3 | FAM32A | Homo sapien | 13.176 | 1 | 1 | 2.0E+05 | 0.0E+00 | 0.0E+00 | 1.6E+05 | 2.4E+05 | 1.6E+05 | 0.0E+00 | 0.0E+00 | 0.0E+00 | 0.0E+00 | [illegible] |
| 7451 | 1 | Q9Y496 | Q9Y496 | Kinesin-like p | KIF3A | Homo sapien | 80.042 | 1 | 1 | 0.0E+00 | 0.0E+00 | 0.0E+00 | 0.0E+00 | 6.4E+03 | 0.0E+00 | 0.0E+00 | 0.0E+00 | 0.0E+00 | 0.0E+00 | [illegible] |
| 7452 | 1 | Q9Y4C2 | Q9Y4C2 | TRPM8 chan | TCAF1 | Homo sapien | 102.128 | 1 | 1 | 3.8E+03 | 0.0E+00 | 0.0E+00 | 0.0E+00 | 0.0E+00 | 0.0E+00 | 0.0E+00 | 0.0E+00 | 0.0E+00 | 0.0E+00 | [illegible] |
| 7453 | 1 | Q9Y4C5 | Q9Y4C5 | Carbohydrate | CHST2 | Homo sapien | 57.857 | 1 | 1 | 4.1E+03 | 1.1E+04 | 0.0E+00 | 0.0E+00 | 7.7E+03 | 0.0E+00 | 0.0E+00 | 0.0E+00 | 0.0E+00 | 6.4E+03 | [illegible] |
| 7454 | 1 | Q9Y4H4 | Q9Y4H4 | G-protein-sig | GPSM3 | Homo sapien | 17.862 | 1 | 1 | 3.2E+04 | 0.0E+00 | 1.2E+04 | 0.0E+00 | 0.0E+00 | 0.0E+00 | 0.0E+00 | 7.5E+04 | 0.0E+00 | 0.0E+00 | [illegible] |
| 7455 | 1 | Q9Y508 | Q9Y508 | E3 ubiquitin-p | RNF114 | Homo sapien | 25.694 | 1 | 1 | 6.5E+05 | 1.0E+06 | 7.8E+05 | 7.4E+05 | 5.9E+05 | 6.4E+05 | 1.5E+06 | 1.1E+06 | 1.1E+06 | 1.6E+06 | [illegible] |
| 7456 | 1 | Q9Y547 | Q9Y547 | Intraflagellar | IFT25 | Homo sapien | 16.295 | 1 | 1 | 0.0E+00 | 0.0E+00 | 4.7E+04 | 9.8E+04 | 2.7E+04 | 3.5E+04 | 0.0E+00 | 0.0E+00 | 0.0E+00 | 0.0E+00 | [illegible] |
| 7457 | 1 | Q9Y5G6 | Q9Y5G6 | Protocadheri | PCDHGA7 | Homo sapien | 101.721 | 1 | 1 | 0.0E+00 | 0.0E+00 | 0.0E+00 | 0.0E+00 | 0.0E+00 | 0.0E+00 | 0.0E+00 | 0.0E+00 | 0.0E+00 | 0.0E+00 | [illegible] |
| 7458 | 1 | Q9Y5J6 | Q9Y5J6 | Mitochondria | TIMM10B | Homo sapien | 11.586 | 1 | 1 | 0.0E+00 | 0.0E+00 | 1.3E+04 | 9.6E+03 | 1.2E+04 | 0.0E+00 | 0.0E+00 | 0.0E+00 | 1.4E+04 | 1.8E+04 | [illegible] |
| 7459 | 1 | Q9Y5J7 | Q9Y5J7 | Mitochondria | TIMM9 | Homo sapien | 10.376 | 1 | 1 | 2.8E+04 | 1.4E+04 | 3.2E+04 | 0.0E+00 | 4.1E+04 | 2.4E+04 | 2.8E+04 | 3.2E+04 | 3.9E+04 | 0.0E+00 | [illegible] |
| 7460 | 1 | Q9Y5J9 | Q9Y5J9 | Mitochondria | TIMM8B | Homo sapien | 9.342 | 1 | 1 | 0.0E+00 | 2.3E+04 | 0.0E+00 | 0.0E+00 | 0.0E+00 | 0.0E+00 | 0.0E+00 | 0.0E+00 | 0.0E+00 | 0.0E+00 | [illegible] |
| 7461 | 1 | Q9Y5Z9 | Q9Y5Z9 | UbiA prenyltr | UBIAD1 | Homo sapien | 36.831 | 1 | 1 | 0.0E+00 | 0.0E+00 | 0.0E+00 | 1.3E+04 | 1.7E+04 | 0.0E+00 | 0.0E+00 | 0.0E+00 | 0.0E+00 | 0.0E+00 | [illegible] |
| 7462 | 1 | Q9Y605 | Q9Y605 | MORF4 famil | MRFAP1 | Homo sapien | 14.648 | 1 | 1 | 0.0E+00 | 0.0E+00 | 1.2E+04 | 1.3E+04 | 0.0E+00 | 0.0E+00 | 0.0E+00 | 0.0E+00 | 0.0E+00 | 0.0E+00 | [illegible] |
| 7463 | 1 | Q9Y680 | Q9Y680 | Peptidyl-proly | FKBP7 | Homo sapien | 25.794 | 1 | 1 | 0.0E+00 | 0.0E+00 | 0.0E+00 | 0.0E+00 | 0.0E+00 | 0.0E+00 | 0.0E+00 | 0.0E+00 | 0.0E+00 | 0.0E+00 | [illegible] |
| 7464 | 1 | Q9Y6A9 | Q9Y6A9 | Signal peptid | SPCS1 | Homo sapien | 18.297 | 1 | 1 | 0.0E+00 | 0.0E+00 | 0.0E+00 | 0.0E+00 | 2.7E+05 | 1.8E+05 | 1.6E+05 | 3.0E+05 | 2.8E+05 | 0.0E+00 | [illegible] |
| 7465 | 1 | Q9Y6B6 | Q9Y6B6 | Small COPII c | SAR1B | Homo sapien | 22.41 | 2 | 1 | 1.3E+05 | 1.1E+05 | 1.5E+05 | 1.0E+05 | 1.1E+05 | 1.4E+05 | 1.8E+05 | 1.5E+05 | 2.0E+05 | 2.4E+05 | [illegible] |
| 7466 | 1 | Q9Y6D0 | Q9Y6D0 | Selenoprotein | SELENOK | Homo sapien | 10.617 | 1 | 1 | 0.0E+00 | 0.0E+00 | 0.0E+00 | 0.0E+00 | 0.0E+00 | 3.9E+04 | 0.0E+00 | 0.0E+00 | 0.0E+00 | 0.0E+00 | [illegible] |
| 7467 | 1 | Q9Y6I8 | Q9Y6I8 | Peroxisomal r | PXMP4 | Homo sapien | 24.263 | 1 | 1 | 4.8E+03 | 4.4E+03 | 5.8E+03 | 4.8E+03 | 6.0E+03 | 4.7E+03 | 0.0E+00 | 0.0E+00 | 0.0E+00 | 0.0E+00 | [illegible] |
| 7468 | 1 | Q9Y6K0 | Q9Y6K0 | Choline/ethar | CEPT1 | Homo sapien | 46.552 | 1 | 1 | 2.5E+05 | 3.4E+05 | 3.9E+05 | 3.2E+05 | 3.1E+05 | 3.0E+05 | 2.1E+05 | 3.2E+05 | 2.9E+05 | 3.7E+05 | [illegible] |
| 7469 | 1 | Q9Y6L7 | Q9Y6L7 | Tolloid-like pr | TLL2 | Homo sapien | 113.56 | 1 | 1 | 1.7E+06 | 1.6E+06 | 0.0E+00 | 2.1E+06 | 1.9E+06 | 2.5E+06 | 0.0E+00 | 0.0E+00 | 0.0E+00 | 0.0E+00 | [illegible] |
| 7470 | 1 | Q9Y6Q9 | Q9Y6Q9 | Nuclear rece | NCOA3 | Homo sapien | 155.295 | 1 | 1 | 0.0E+00 | 0.0E+00 | 0.0E+00 | 0.0E+00 | 0.0E+00 | 0.0E+00 | 0.0E+00 | 0.0E+00 | 0.0E+00 | 0.0E+00 | [illegible] |
| 7471 | 1 | Q9Y6R9 | Q9Y6R9 | Centrosomal | CCDC61 | Homo sapien | 57.369 | 1 | 1 | 0.0E+00 | 0.0E+00 | 0.0E+00 | 0.0E+00 | 0.0E+00 | 0.0E+00 | 0.0E+00 | 0.0E+00 | 0.0E+00 | 0.0E+00 | [illegible] |
| 7472 | 1 | Q9Y6T7 | Q9Y6T7 | Diacylglycero | DGKB | Homo sapien | 90.596 | 1 | 1 | 0.0E+00 | 0.0E+00 | 0.0E+00 | 0.0E+00 | 0.0E+00 | 0.0E+00 | 0.0E+00 | 0.0E+00 | 0.0E+00 | 0.0E+00 | [illegible] |
| 7473 | 1 | Q9Y6X5 | Q9Y6X5 | Bis(5'-adeno | ENPP4 | Homo sapien | 51.641 | 2 | 1 | 0.0E+00 | 0.0E+00 | 0.0E+00 | 1.2E+05 | 0.0E+00 | 0.0E+00 | 0.0E+00 | 0.0E+00 | 0.0E+00 | 0.0E+00 | [illegible] |
| 7474 | 1 | Q9Y6Y0 | Q9Y6Y0 | Influenza viru | IVNS1ABP | Homo sapien | 71.729 | 1 | 1 | 0.0E+00 | 7.6E+03 | 7.2E+03 | 0.0E+00 | 6.0E+03 | 5.3E+03 | 7.8E+03 | 0.0E+00 | 0.0E+00 | 1.3E+04 | [illegible] |

Table S4. Fragments' quantification detected with DIA LC-MS/MS proteomics.

| Master gene symb | Sequence pc | Sequence pc | Sequence | SS_1 | SS_2 | SS_3 | SS_4 | SS_5 | SS_6 | SS_7 | SS_8 | SS_9 | SS_10 | SS_11 | WS_1 | WS_2 | WS_3 | SS_12 |
|---|---|---|---|---|---|---|---|---|---|---|---|---|---|---|---|---|---|---|
| SLC25A13 | 57 | 69 | TVELLSGVVDQTK | 0.0E+00 | 0.0E+00 | 0.0E+00 | 0.0E+00 | 0.0E+00 | 0.0E+00 | 0.0E+00 | 0.0E+00 | 0.0E+00 | 0.0E+00 | 0.0E+00 | 0.0E+00 | 1.4E+04 | 1.6E+04 | 0.0E+00 |
| SLC25A13 | 252 | 260 | EEFVLAAQK | 0.0E+00 | 0.0E+00 | 0.0E+00 | 0.0E+00 | 0.0E+00 | 0.0E+00 | 0.0E+00 | 0.0E+00 | 0.0E+00 | 0.0E+00 | 0.0E+00 | 0.0E+00 | 0.0E+00 | 0.0E+00 | 0.0E+00 |
| SLC25A13 | 361 | 372 | STGSFVGELMYK | 0.0E+00 | 0.0E+00 | 0.0E+00 | 0.0E+00 | 0.0E+00 | 0.0E+00 | 0.0E+00 | 0.0E+00 | 0.0E+00 | 0.0E+00 | 0.0E+00 | 0.0E+00 | 0.0E+00 | 0.0E+00 | 0.0E+00 |
| SLC25A13 | 393 | 405 | GLLPQLLGVAPEK | 0.0E+00 | 0.0E+00 | 0.0E+00 | 0.0E+00 | 0.0E+00 | 0.0E+00 | 0.0E+00 | 0.0E+00 | 0.0E+00 | 0.0E+00 | 0.0E+00 | 3.9E+04 | 0.0E+00 | 3.6E+04 | 0.0E+00 |
| SLC25A13 | 589 | 605 | SSPQFGVTLLTYELL | 0.0E+00 | 0.0E+00 | 0.0E+00 | 0.0E+00 | 0.0E+00 | 0.0E+00 | 0.0E+00 | 0.0E+00 | 0.0E+00 | 0.0E+00 | 0.0E+00 | 0.0E+00 | 0.0E+00 | 0.0E+00 | 0.0E+00 |
| SLC25A13 | 627 | 641 | INLPAPNPDHVGGYI | 0.0E+00 | 0.0E+00 | 0.0E+00 | 0.0E+00 | 0.0E+00 | 0.0E+00 | 0.0E+00 | 0.0E+00 | 0.0E+00 | 0.0E+00 | 0.0E+00 | 2.1E+05 | 2.2E+05 | 1.9E+05 | 0.0E+00 |
| SLC25A13 | 642 | 653 | LAVATFAGIENK | 0.0E+00 | 0.0E+00 | 0.0E+00 | 0.0E+00 | 0.0E+00 | 0.0E+00 | 0.0E+00 | 0.0E+00 | 0.0E+00 | 0.0E+00 | 0.0E+00 | 2.4E+04 | 2.4E+04 | 2.1E+04 | 0.0E+00 |
| SLC25A13 | 654 | 669 | FGLYLPLFKPSVSTS | 0.0E+00 | 0.0E+00 | 0.0E+00 | 0.0E+00 | 0.0E+00 | 0.0E+00 | 0.0E+00 | 0.0E+00 | 0.0E+00 | 0.0E+00 | 0.0E+00 | 6.3E+03 | 0.0E+00 | 8.3E+03 | 0.0E+00 |

| Sequence | WS_4 | WS_5 | WS_6 | WS_7 | WS_8 | WS_9 | SM_1 | SM_2 | SM_3 | SM_4 | SM_5 | SM_6 | SM_7 | WW_1 | WW_2 | WW_3 | WW_4 | WW_5 | WW_6 |
|---|---|---|---|---|---|---|---|---|---|---|---|---|---|---|---|---|---|---|---|
| TVELLSGVVDQTK | 0.0E+00 | 0.0E+00 | 0.0E+00 | 2.5E+04 | 1.9E+04 | 2.5E+04 | 1.6E+04 | 2.0E+04 | 0.0E+00 | 2.2E+04 | 2.2E+04 | 0.0E+00 | 0.0E+00 | 2.9E+04 | 2.2E+04 | 3.7E+04 | 2.1E+04 | 2.5E+04 | 2.8E+04 |
| EEFVLAAQK | 0.0E+00 | 0.0E+00 | 0.0E+00 | 1.3E+05 | 1.4E+05 | 0.0E+00 | 1.4E+05 | 1.5E+05 | 1.2E+05 | 1.4E+05 | 1.1E+05 | 8.2E+04 | 1.1E+05 | 0.0E+00 | 0.0E+00 | 3.3E+05 | 2.0E+05 | 2.3E+05 | 2.1E+05 |
| STGSFVGELMYK | 0.0E+00 | 0.0E+00 | 2.0E+04 | 0.0E+00 | 0.0E+00 | 2.0E+04 | 0.0E+00 | 0.0E+00 | 0.0E+00 | 2.0E+04 | 1.2E+04 | 0.0E+00 | 0.0E+00 | 0.0E+00 | 0.0E+00 | 4.0E+04 | 0.0E+00 | 2.5E+04 | 1.8E+04 |
| GLLPQLLGVAPEK | 4.0E+04 | 0.0E+00 | 4.1E+04 | 3.7E+04 | 3.6E+04 | 5.0E+04 | 0.0E+00 | 0.0E+00 | 0.0E+00 | 0.0E+00 | 3.7E+04 | 0.0E+00 | 0.0E+00 | 1.0E+05 | 7.9E+04 | 1.1E+05 | 8.0E+04 | 5.5E+04 | 5.8E+04 |
| SSPQFGVTLLTYELL | 0.0E+00 | 0.0E+00 | 0.0E+00 | 0.0E+00 | 0.0E+00 | 0.0E+00 | 0.0E+00 | 0.0E+00 | 0.0E+00 | 0.0E+00 | 0.0E+00 | 0.0E+00 | 0.0E+00 | 0.0E+00 | 0.0E+00 | 0.0E+00 | 0.0E+00 | 0.0E+00 | 3.4E+04 |
| INLPAPNPDHVGGYI | 2.2E+05 | 1.8E+05 | 1.5E+05 | 0.0E+00 | 0.0E+00 | 0.0E+00 | 0.0E+00 | 0.0E+00 | 0.0E+00 | 0.0E+00 | 5.8E+04 | 0.0E+00 | 0.0E+00 | 5.5E+05 | 3.3E+05 | 4.0E+05 | 2.6E+05 | 2.2E+05 | 0.0E+00 |
| LAVATFAGIENK | 3.0E+04 | 3.0E+04 | 1.8E+04 | 1.9E+04 | 0.0E+00 | 2.7E+04 | 0.0E+00 | 2.8E+04 | 1.2E+04 | 2.0E+04 | 2.4E+04 | 1.9E+04 | 2.0E+04 | 4.2E+04 | 5.5E+04 | 5.9E+04 | 3.1E+04 | 3.8E+04 | 2.9E+04 |
| FGLYLPLFKPSVSTS | 7.8E+03 | 1.1E+04 | 6.5E+03 | 6.0E+03 | 4.6E+03 | 6.3E+03 | 6.3E+03 | 5.5E+03 | 4.1E+03 | 4.8E+03 | 4.7E+03 | 0.0E+00 | 0.0E+00 | 1.9E+04 | 1.5E+04 | 1.7E+04 | 1.3E+04 | 1.2E+04 | 7.7E+03 |

Table S5. Clinical manifestations between LoF/LoF and Lof/Mis genotype in citrin deficiency.

| Variable | Status | LoF/LoF (n = 16) | LoF/Mis (n = 7) |
|---|---|---|---|
| NBS | Positive | 12 (75.0%) | 3 (42.9%) |
| | Negative | 3 (18.8%) | 3 (42.9%) |
| | NA | 1 (6.3%) | 1 (14.3%) |
| NICCD | Present | 15 (93.8%) | 3 (42.9%) |
| | Absent | 1 (6.3%) | 3 (42.9%) |
| | NA | 0 (0.0%) | 1 (14.3%) |
| Food preferences | Present | 12 (75.0%) | 5 (71.4%) |
| | Absent | 2 (12.5%) | 2 (28.6%) |
| | NA | 2 (12.5%) | 0 (0.0%) |
| Growth restriction | Present | 1 (6.3%) | 0 (0.0%) |
| | Absent | 15 (93.8%) | 7 (100.0%) |
| | NA | 0 (0.0%) | 0 (0.0%) |
| Pancreatitis | Present | 0 (0.0%) | 0 (0.0%) |
| | Absent | 16 (100.0%) | 7 (100.0%) |
| | NA | 0 (0.0%) | 0 (0.0%) |
| Fatigue | Present | 0 (0.0%) | 2 (28.6%) |
| | Absent | 15 (93.8%) | 5 (71.4%) |
| | NA | 1 (6.3%) | 0 (0.0%) |
| Anorexia | Present | 0 (0.0%) | 0 (0.0%) |
| | Absent | 16 (100.0%) | 7 (100.0%) |
| | NA | 0 (0.0%) | 0 (0.0%) |

Data are shown as n (%). The lower proportion of NICCD observed in the LoF/Mis group may be due to the inclusion of sibling cases identified through family screening (individual SM_3 and SM_5).
LoF, Loss of function; Mis, missense; NBS, newborn screening; NICCD, neonatal intrahepatic cholestasis by citrin deficiency; NA, not available.

Table S6. Performance metrics of candidate panel sizes evaluated by leave-one-out cross-validation.

| m | AUC | Threshold | Sensitivity | Specificity | Precision | Accuracy | Balanced Accuracy |
|---|---|---|---|---|---|---|---|
| 5 | 0.8339768339768342 | 0.5835285160658178 | 0.5714285714285714 | 0.9459459459459459 | 0.6666666666666666 | 0.8863636363636364 | 0.7586872586872586 |
| 10 | 0.9343629343629345 | 0.8437300743356432 | 0.5714285714285714 | 1 | 1 | 0.9318181818181818 | 0.7857142857142857 |
| 15 | 0.9806949806949807 | 0.3697923014309361 | 0.8571428571428571 | 1 | 1 | 0.9772727272727273 | 0.9285714285714286 |
| 20 | 0.945945946 | 0.5801937261393447 | 0.7142857142857143 | 1 | 1 | 0.9545454545454546 | 0.8571428571428572 |
| 30 | 0.9884169884169884 | 0.21326918876872636 | 1 | 0.9459459459459459 | 0.7777777777777778 | 0.9545454545454546 | 0.972972973 |
| 40 | 0.972972973 | 0.23096071631298182 | 0.8571428571428571 | 0.972972973 | 0.8571428571428571 | 0.9545454545454546 | 0.915057915 |
| 50 | 0.9536679536679538 | 0.5487540118100296 | 0.7142857142857143 | 1 | 1 | 0.9545454545454546 | 0.8571428571428572 |

Table S7. Coefficients and intercept of the final elastic net model for the 30-protein panel.

| FeatureID | Gene | Protein | Coef | | Intercept |
|---|---|---|---|---|---|
| Q06203 | PPAT | Amidophosphoribosyltransferase | -0.175139727 | | -1.2851237 |
| Q8WUD1 | RAB2B | Ras-related protein Rab-2B | -0.330726019 | | |
| Q99829 | CPNE1 | Copine-1 | -0.101033232 | | |
| P01236 | PRL | Prolactin | 0.093157944 | | |
| O95757 | HSPA4L | Heat shock 70 kDa protein 4L | 0 | | |
| Q9UHG3 | PCYOX1 | Prenylcysteine oxidase 1 | -0.114235826 | | |
| O15013 | ARHGEF10 | Rho guanine nucleotide exchange factor 10 | 0.43705226257987007 | | |
| O94979 | SEC31A | Protein transport protein Sec31A | 0.1763246060842403 | | |
| P55157 | MTTP | Microsomal triglyceride transfer protein large subunit | 0.062974033 | | |
| Q9UBQ7 | GRHPR | Glyoxylate reductase/hydroxypyruvate reductase | -0.289822145 | | |
| Q9H115 | NAPB | Beta-soluble NSF attachment protein | -0.427724039 | | |
| O95983 | MBD3 | Methyl-CpG-binding domain protein 3 | 0 | | |
| Q99759 | MAP3K3 | Mitogen-activated protein kinase kinase kinase 3 | 0 | | |
| Q15369 | ELOC | Elongin-C | 0 | | |
| Q5JSL3 | DOCK11 | Dedicator of cytokinesis protein 11 | 0 | | |
| Q8N3P4 | VPS8 | Vacuolar protein sorting-associated protein 8 homolog | 0 | | |
| P78509 | RELN | Reelin | -0.091517091 | | |
| P06733 | ENO1 | Alpha-enolase | -0.014877945 | | |
| O00468 | AGRN | Agrin | 0 | | |
| Q9Y6U3 | SCIN | Scinderin | 0 | | |
| P00519 | ABL1 | Tyrosine-protein kinase ABL1 | 0 | | |
| Q8TEQ0 | SNX29 | Sorting nexin-29 | 0 | | |
| Q9Y5S1 | TRPV2 | Transient receptor potential cation channel subfamily V member 2 | -0.08408396 | | |
| O00522 | KRIT1 | Krev interaction trapped protein 1 | 0 | | |
| Q7Z3K3 | POGZ | Pogo transposable element with ZNF domain | 0 | | |
| Q96BS2 | TESC | Calcineurin B homologous protein 3 | -0.188204986 | | |
| Q9UBX1 | CTSF | Cathepsin F | 0 | | |
| Q6ZRP7 | QSOX2 | Sulfhydryl oxidase 2 | 0 | | |
| P56556 | NDUFA6 | NADH dehydrogenase [ubiquinone] 1 alpha subcomplex subunit 6 | -0.020275006 | | |
| Q86UY8 | NT5DC3 | 5'-nucleotidase domain-containing protein 3 | 0 | | |

Table S8. Leave-one-LoF/Mis-out stability metrics for the final proteomic panel.

| FeatureID | Gene | Protein | Retention across leave-one-LoF/MS-out runs | Mean EN selection frequency across leave-one-LoF/MS-out runs | Rank (median) | Rank (IQR) |
|---|---|---|---|---|---|---|
| Q99829 | CPNE1 | Copine-1 | 1 | 0.3476190476190476 | 2 | 2.5 |
| P01236 | PRL | Prolactin | 1 | 0.3 | 5 | 4.5 |
| Q06203 | PPAT | Amidophosphoribosyltransferase | 1 | 0.2904761904761905 | 3 | 3.5 |
| O15013 | ARHGEF10 | Rho guanine nucleotide exchange factor 10 | 1 | 0.2857142857142857 | 5 | 4 |
| O94979 | SEC31A | Protein transport protein Sec31A | 1 | 0.2809523809523809 | 5 | 5 |
| Q8WUD1 | RAB2B | Ras-related protein Rab-2B | 1 | 0.26666666666666666 | 6 | 4.5 |
| O00522 | KRIT1 | Krev interaction trapped protein 1 | 1 | 0.2571428571428571 | 5 | 4.5 |
| Q9UHG3 | PCYOX1 | Prenylcysteine oxidase 1 | 1 | 0.21428571428571427 | 7 | 6.5 |
| P55157 | MTTP | Microsomal triglyceride transfer protein large subunit | 1 | 0.21428571428571427 | 8 | 1.5 |
| Q9H115 | NAPB | Beta-soluble NSF attachment protein | 1 | 0.15238095238095237 | 12 | 6 |
| O95757 | HSPA4L | Heat shock 70 kDa protein 4L | 1 | 0.1476190476190476 | 12 | 9 |
| Q96BS2 | TESC | Calcineurin B homologous protein 3 | 1 | 0.13809523809523808 | 11 | 12 |
| Q7Z3K3 | POGZ | Pogo transposable element with ZNF domain | 1 | 0.1238095238095238 | 15 | 6 |
| Q5JSL3 | DOCK11 | Dedicator of cytokinesis protein 11 | 1 | 0.11904761904761904 | 14 | 6 |
| O95983 | MBD3 | Methyl-CpG-binding domain protein 3 | 1 | 0.11904761904761904 | 16 | 7.5 |
| Q9UBQ7 | GRHPR | Glyoxylate reductase/hydroxypyruvate reductase | 1 | 0.10952380952380951 | 13 | 5 |
| Q9Y6U3 | SCIN | Scinderin | 1 | 0.10476190476190476 | 16 | 5.5 |
| Q9Y5S1 | TRPV2 | Transient receptor potential cation channel subfamily V member 2 | 1 | 0.085714286 | 17 | 3.5 |
| P56556 | NDUFA6 | NADH dehydrogenase [ubiquinone] 1 alpha subcomplex subunit 6 | 1 | 0.080952381 | 18 | 7.5 |
| P06733 | ENO1 | Alpha-enolase | 1 | 0.076190476 | 20 | 8.5 |
| Q8TEQ0 | SNX29 | Sorting nexin-29 | 1 | 0.076190476 | 22 | 6 |
| O00468 | AGRN | Agrin | 1 | 0.061904762 | 21 | 11.5 |
| Q86UY8 | NT5DC3 | 5'-nucleotidase domain-containing protein 3 | 1 | 0.052380952 | 24 | 6.5 |
| Q8N3P4 | VPS8 | Vacuolar protein sorting-associated protein 8 homolog | 1 | 0.052380952380952375 | 21 | 4.5 |
| Q6ZRP7 | QSOX2 | Sulfhydryl oxidase 2 | 1 | 0.042857142857142864 | 25 | 7.5 |
| P78509 | RELN | Reelin | 1 | 0.038095238 | 23 | 1.5 |
| Q15369 | ELOC | Elongin-C | 1 | 0.033333333 | 22 | 3.5 |
| Q9UBX1 | CTSF | Cathepsin F | 1 | 0.023809523809523808 | 29 | 3 |
| Q99759 | MAP3K3 | Mitogen-activated protein kinase kinase kinase 3 | 1 | 0.014285714285714287 | 22 | 5 |
| P00519 | ABL1 | Tyrosine-protein kinase ABL1 | 1 | 0 | 28 | 1 |

Table S9. Performance of sPLS-DA across the prespecified keepX grid in leave-one-out cross-validation.

| input_topN | keepX | ncomp | Accuracy | Sensitivity | Specificity | Balanced Error Rate | True Positive | False Positive | False Negative | True Negative |
|---|---|---|---|---|---|---|---|---|---|---|
| 1000 | 5 | 2 | 0.886363636 | 0.285714285 | 1 | 0.35714285714285715 | 2 | 0 | 5 | 37 |
| 1000 | 10 | 2 | 0.886363636 | 0.285714285 | 1 | 0.35714285714285715 | 2 | 0 | 5 | 37 |
| 1000 | 15 | 2 | 0.909090909 | 0.428571428 | 1 | 0.2857142857142857 | 3 | 0 | 4 | 37 |
| 1000 | 20 | 2 | 0.909090909 | 0.428571428 | 1 | 0.2857142857142857 | 3 | 0 | 4 | 37 |
| 1000 | 30 | 2 | 0.931818181 | 0.571428571 | 1 | 0.2142857142857143 | 4 | 0 | 3 | 37 |
| 1000 | 40 | 2 | 0.931818181 | 0.571428571 | 1 | 0.2142857142857143 | 4 | 0 | 3 | 37 |
| 1000 | 50 | 2 | 0.931818181 | 0.571428571 | 1 | 0.2142857142857143 | 4 | 0 | 3 | 37 |

Table S10. Gene Ontology enrichment analysis of the 15-protein core proteomic signature.

| ID | Description | GeneRatio | BgRatio | RichFactor | FoldEnrichm | zScore | pvalue | p.adjust | qvalue | geneID | Count |
|---|---|---|---|---|---|---|---|---|---|---|---|
| GO:0051592 | response to calcium ion | 3/15 | 139/18805 | 0.02158273 | 27.05755395 | 8.712098477 | 1.684908426 | 0.047502804 | 0.032070204 | CPNE1/SEC31A/MTTP | 3 |
| GO:0009152 | purine ribonucleotide biosynthetic process | 3/15 | 157/18805 | 0.01910828 | 23.95541401 | 8.160676958 | 2.413067579 | 0.047502804 | 0.032070204 | PPAT/ENO1/NDUFA6 | 3 |
| GO:0009260 | ribonucleotide biosynthetic process | 3/15 | 173/18805 | 0.017341040 | 21.73988439 | 7.742959822 | 3.209606707 | 0.047502804 | 0.032070204 | PPAT/ENO1/NDUFA6 | 3 |
| GO:0046390 | ribose phosphate biosynthetic process | 3/15 | 180/18805 | 0.016666666 | 20.89444444 | 7.577523440 | 3.605525922 | 0.047502804 | 0.032070204 | PPAT/ENO1/NDUFA6 | 3 |

Table S11. Gene set enrichment analysis of RNA-seq data comparing LoF/LoF and Ref/Ref samples.

| ID | Description | setSize | enrichmentScore | NES | pvalue | p.adjust | qvalue | rank | leading_edge | core_enrichment |
|---|---|---|---|---|---|---|---|---|---|---|
| GO:0003735 | structural constituent of ribosome | 158 | 0.6665395 | 2.63363183 | 1e-10 | 1.221e-07 | 1.013684210 | 2137 | tags=51%, list=13%, signal=45% | [illegible] |
| GO:0022832 | voltage-gated channel activity | 121 | -0.5753198 | -1.9250661 | 5.879701425 | 0.00017948 | 0.000149 | 3298 | tags=41%, list=19%, signal=34% | [illegible] |
| GO:0005244 | voltage-gated monoatomic ion channel activity | 119 | -0.573977 | -1.9149109 | 4.558764332 | 0.00017948 | 0.000149 | 3298 | tags=41%, list=19%, signal=33% | [illegible] |
| GO:0022836 | gated channel activity | 208 | -0.5082778 | -1.7864673 | 3.043053043 | 0.00017948 | 0.000149 | 4099 | tags=42%, list=24%, signal=32% | [illegible] |
| GO:0003729 | mRNA binding | 315 | 0.38642055 | 1.63962349 | 1.346891703 | 0.00032891 | 0.00027306 | 2147 | tags=29%, list=13%, signal=26% | [illegible] |
| GO:0022843 | voltage-gated monoatomic cation channel activity | 92 | -0.5939542 | -1.9015756 | 3.406300651 | 0.00069318 | 0.00057549 | 3298 | tags=41%, list=19%, signal=34% | [illegible] |
| GO:0005249 | voltage-gated potassium channel activity | 58 | -0.6486533 | -1.9455824 | 1.014533570 | 0.00137638 | 0.00114269 | 3049 | tags=48%, list=18%, signal=40% | [illegible] |
| GO:0015079 | potassium ion transmembrane transporter activity | 98 | -0.5664347 | -1.8283113 | 9.375140058 | 0.00137638 | 0.00114269 | 3461 | tags=42%, list=20%, signal=34% | [illegible] |
| GO:0046873 | metal ion transmembrane transporter activity | 320 | -0.4363772 | -1.5842762 | 8.985461395 | 0.00137638 | 0.00114269 | 4519 | tags=39%, list=26%, signal=30% | [illegible] |
| GO:0005216 | monoatomic ion channel activity | 303 | -0.4362183 | -1.5872268 | 1.175860330 | 0.00143573 | 0.00119195 | 4001 | tags=37%, list=23%, signal=29% | [illegible] |
| GO:0015267 | channel activity | 334 | -0.4271803 | -1.552952 | 1.858341128 | 0.00189086 | 0.00156981 | 4001 | tags=36%, list=23%, signal=28% | [illegible] |
| GO:0022803 | passive transmembrane transporter activity | 334 | -0.4271803 | -1.552952 | 1.858341128 | 0.00189086 | 0.00156981 | 4001 | tags=36%, list=23%, signal=28% | [illegible] |
| GO:0019843 | rRNA binding | 63 | 0.57491166 | 2.00638429 | 2.564421643 | 0.00240858 | 0.00199963 | 2321 | tags=43%, list=14%, signal=37% | [illegible] |
| GO:0030594 | neurotransmitter receptor activity | 47 | -0.6490491 | -1.8806932 | 3.848022792 | 0.00276379 | 0.00229452 | 4184 | tags=47%, list=24%, signal=35% | [illegible] |
| GO:0005267 | potassium channel activity | 71 | -0.590277 | -1.8312933 | 3.815256820 | 0.00276379 | 0.00229452 | 3461 | tags=45%, list=20%, signal=36% | [illegible] |
| GO:0009055 | electron transfer activity | 108 | 0.48176191 | 1.78818478 | 3.522137005 | 0.00276379 | 0.00229452 | 2705 | tags=39%, list=16%, signal=33% | [illegible] |
| GO:0015103 | inorganic anion transmembrane transporter activity | 122 | -0.5286696 | -1.770985 | 3.511770482 | 0.00276379 | 0.00229452 | 4691 | tags=50%, list=27%, signal=37% | [illegible] |
| GO:0019199 | transmembrane receptor protein kinase activity | 60 | -0.6162184 | -1.8549955 | 4.971538625 | 0.00337236 | 0.00279976 | 4264 | tags=53%, list=25%, signal=40% | [illegible] |
| GO:0097493 | structural molecule activity conferring elasticity | 10 | -0.8926939 | -1.9120189 | 8.204812853 | 0.00527267 | 0.00437742 | 303 | tags=30%, list=2%, signal=29% | 2006/7273/79026 |
| GO:0015291 | secondary active transmembrane transporter activity | 224 | -0.4326572 | -1.5276179 | 0.00013273 | 0.00810312 | 0.00672728 | 3665 | tags=37%, list=21%, signal=29% | [illegible] |
| GO:0004714 | transmembrane receptor protein tyrosine kinase activity | 46 | -0.6259928 | -1.8074867 | 0.00018371 | 0.01068143 | 0.00886781 | 4264 | tags=50%, list=25%, signal=38% | [illegible] |
| GO:0004407 | histone deacetylase activity | 20 | -0.7511548 | -1.9033255 | 0.0002556 | 0.01073845 | 0.00891515 | 1384 | tags=45%, list=8%, signal=41% | [illegible] |
| GO:0033558 | protein lysine deacetylase activity | 20 | -0.7511548 | -1.9033255 | 0.0002556 | 0.01073845 | 0.00891515 | 1384 | tags=45%, list=8%, signal=41% | [illegible] |
| GO:0051959 | dynein light intermediate chain binding | 27 | -0.7072465 | -1.8770781 | 0.00024521 | 0.01073845 | 0.00891515 | 3453 | tags=59%, list=20%, signal=47% | [illegible] |
| GO:0038064 | collagen receptor activity | 41 | -0.6423559 | -1.8369421 | 0.00021215 | 0.01073845 | 0.00891515 | 4060 | tags=49%, list=24%, signal=37% | [illegible] |
| GO:0005017 | platelet-derived growth factor receptor activity | 38 | -0.6425663 | -1.8144387 | 0.00026631 | 0.01073845 | 0.00891515 | 4060 | tags=50%, list=24%, signal=38% | [illegible] |
| GO:0005019 | platelet-derived growth factor beta-receptor activity | 38 | -0.6425663 | -1.8144387 | 0.00026631 | 0.01073845 | 0.00891515 | 4060 | tags=50%, list=24%, signal=38% | [illegible] |
| GO:0016500 | protein-hormone receptor activity | 47 | -0.6145371 | -1.7806909 | 0.00025091 | 0.01073845 | 0.00891515 | 4060 | tags=47%, list=24%, signal=36% | [illegible] |
| GO:0015108 | chloride transmembrane transporter activity | 86 | -0.542811 | -1.7275011 | 0.00022635 | 0.01073845 | 0.00891515 | 4691 | tags=49%, list=27%, signal=36% | [illegible] |
| GO:0180051 | translation factor activity | 81 | 0.47919937 | 1.70669694 | 0.00027264 | 0.01073845 | 0.00891515 | 2147 | tags=36%, list=13%, signal=31% | [illegible] |
| GO:0030695 | GTPase regulator activity | 423 | -0.3869079 | -1.4323127 | 0.00024985 | 0.01073845 | 0.00891515 | 3675 | tags=34%, list=22%, signal=27% | [illegible] |
| GO:0008135 | translation factor activity, RNA binding | 80 | 0.47998282 | 1.70621478 | 0.00033789 | 0.01269469 | 0.01053923 | 2147 | tags=36%, list=13%, signal=32% | [illegible] |
| GO:0060589 | nucleoside-triphosphatase regulator activity | 424 | -0.3868905 | -1.4328904 | 0.0003431 | 0.01269469 | 0.01053923 | 3675 | tags=34%, list=22%, signal=27% | [illegible] |
| GO:0005003 | ephrin receptor activity | 40 | -0.6378438 | -1.8129916 | 0.00037395 | 0.01304552 | 0.0108305 | 4241 | tags=50%, list=25%, signal=38% | [illegible] |
| GO:0005005 | transmembrane-ephrin receptor activity | 40 | -0.6378438 | -1.8129916 | 0.00037395 | 0.01304552 | 0.0108305 | 4241 | tags=50%, list=25%, signal=38% | [illegible] |
| GO:0005004 | GPI-linked ephrin receptor activity | 37 | -0.6386547 | -1.7969327 | 0.0005297 | 0.01376092 | 0.01142443 | 4060 | tags=49%, list=24%, signal=37% | [illegible] |
| GO:0005006 | epidermal growth factor receptor activity | 37 | -0.6386547 | -1.7969327 | 0.0005297 | 0.01376092 | 0.01142443 | 4060 | tags=49%, list=24%, signal=37% | [illegible] |
| GO:0005008 | hepatocyte growth factor receptor activity | 37 | -0.6386547 | -1.7969327 | 0.0005297 | 0.01376092 | 0.01142443 | 4060 | tags=49%, list=24%, signal=37% | [illegible] |
| GO:0005009 | insulin receptor activity | 37 | -0.6386547 | -1.7969327 | 0.0005297 | 0.01376092 | 0.01142443 | 4060 | tags=49%, list=24%, signal=37% | [illegible] |
| GO:0005011 | macrophage colony-stimulating factor receptor activity | 37 | -0.6386547 | -1.7969327 | 0.0005297 | 0.01376092 | 0.01142443 | 4060 | tags=49%, list=24%, signal=37% | [illegible] |
| GO:0005018 | platelet-derived growth factor alpha-receptor activity | 37 | -0.6386547 | -1.7969327 | 0.0005297 | 0.01376092 | 0.01142443 | 4060 | tags=49%, list=24%, signal=37% | [illegible] |
| GO:0005020 | stem cell factor receptor activity | 37 | -0.6386547 | -1.7969327 | 0.0005297 | 0.01376092 | 0.01142443 | 4060 | tags=49%, list=24%, signal=37% | [illegible] |
| GO:0008288 | boss receptor activity | 37 | -0.6386547 | -1.7969327 | 0.0005297 | 0.01376092 | 0.01142443 | 4060 | tags=49%, list=24%, signal=37% | [illegible] |
| GO:0036332 | placental growth factor receptor activity | 37 | -0.6386547 | -1.7969327 | 0.0005297 | 0.01376092 | 0.01142443 | 4060 | tags=49%, list=24%, signal=37% | [illegible] |
| GO:0038062 | protein tyrosine kinase collagen receptor activity | 37 | -0.6386547 | -1.7969327 | 0.0005297 | 0.01376092 | 0.01142443 | 4060 | tags=49%, list=24%, signal=37% | [illegible] |
| GO:0060175 | brain-derived neurotrophic factor receptor activity | 37 | -0.6386547 | -1.7969327 | 0.0005297 | 0.01376092 | 0.01142443 | 4060 | tags=49%, list=24%, signal=37% | [illegible] |
| GO:0160215 | deacylase activity | 53 | -0.5948394 | -1.7579409 | 0.00051938 | 0.01376092 | 0.01142443 | 1803 | tags=26%, list=11%, signal=24% | [illegible] |
| GO:0033691 | sialic acid binding | 17 | -0.7630669 | -1.8489484 | 0.00058414 | 0.01480009 | 0.01228716 | 1549 | tags=47%, list=9%, signal=43% | [illegible] |
| GO:0005021 | vascular endothelial growth factor receptor activity | 39 | -0.6361982 | -1.7991286 | 0.00059394 | 0.01480009 | 0.01228716 | 4060 | tags=46%, list=24%, signal=35% | [illegible] |
| GO:0055105 | ubiquitin-protein transferase inhibitor activity | 14 | 0.7890666 | 1.93510298 | 0.00064301 | 0.01481346 | 0.01229826 | 1000 | tags=43%, list=6%, signal=40% | [illegible] |
| GO:0005007 | fibroblast growth factor receptor activity | 38 | -0.6297128 | -1.7781438 | 0.00062388 | 0.01481346 | 0.01229826 | 4264 | tags=50%, list=25%, signal=38% | [illegible] |
| GO:0005201 | extracellular matrix structural constituent | 107 | -0.4929037 | -1.6161879 | 0.00063866 | 0.01481346 | 0.01229826 | 4467 | tags=42%, list=26%, signal=31% | [illegible] |
| GO:0015078 | proton transmembrane transporter activity | 158 | 0.38589662 | 1.52475528 | 0.00062555 | 0.01481346 | 0.01229826 | 2839 | tags=35%, list=17%, signal=29% | [illegible] |
| GO:0004672 | protein kinase activity | 495 | -0.3657468 | -1.3646677 | 0.00072302 | 0.01634819 | 0.0135724 | 3840 | tags=32%, list=22%, signal=26% | [illegible] |
| GO:0048027 | mRNA 5'-UTR binding | 22 | 0.68482583 | 1.88235974 | 0.00078249 | 0.01706112 | 0.01416428 | 4021 | tags=77%, list=24%, signal=59% | [illegible] |
| GO:0005251 | delayed rectifier potassium channel activity | 16 | -0.7796657 | -1.8697979 | 0.0007715 | 0.01706112 | 0.01416428 | 1009 | tags=50%, list=6%, signal=47% | [illegible] |
| GO:0005030 | neurotrophin receptor activity | 39 | -0.6311099 | -1.7847391 | 0.00081176 | 0.01738868 | 0.01443623 | 4060 | tags=49%, list=24%, signal=37% | [illegible] |
| GO:0045499 | chemorepellent activity | 18 | -0.7563831 | -1.8458566 | 0.0008865 | 0.01848297 | 0.01534471 | 2580 | tags=44%, list=15%, signal=38% | [illegible] |
| GO:0005509 | calcium ion binding | 456 | -0.3651112 | -1.3580265 | 0.00089312 | 0.01848297 | 0.01534471 | 3985 | tags=31%, list=23%, signal=25% | [illegible] |
| GO:0098632 | cell-cell adhesion mediator activity | 36 | -0.6426614 | -1.8002936 | 0.00091953 | 0.01871243 | 0.01553521 | 4274 | tags=56%, list=25%, signal=42% | [illegible] |
| GO:0016922 | nuclear receptor binding | 124 | 0.40502882 | 1.54669586 | 0.00095089 | 0.01903344 | 0.01580172 | 2162 | tags=31%, list=13%, signal=27% | [illegible] |
| GO:0015453 | oxidoreduction-driven active transmembrane transporter activity | 65 | 0.49394679 | 1.71767886 | 0.00104104 | 0.02035974 | 0.01690282 | 2920 | tags=46%, list=17%, signal=38% | [illegible] |
| GO:0005254 | chloride channel activity | 57 | -0.559397 | -1.6804333 | 0.0010505 | 0.02035974 | 0.01690282 | 4299 | tags=49%, list=25%, signal=37% | [illegible] |
| GO:0005010 | insulin-like growth factor receptor activity | 39 | -0.6234044 | -1.7629483 | 0.00113918 | 0.02139908 | 0.01776569 | 4060 | tags=49%, list=24%, signal=37% | [illegible] |
| GO:0005516 | calmodulin binding | 173 | -0.4388438 | -1.5222802 | 0.0011225 | 0.02139908 | 0.01776569 | 4913 | tags=50%, list=29%, signal=36% | [illegible] |
| GO:0050661 | NADP binding | 42 | -0.6175446 | -1.7635247 | 0.00123983 | 0.02293678 | 0.0190423 | 3454 | tags=52%, list=20%, signal=42% | [illegible] |
| GO:0031625 | ubiquitin protein ligase binding | 277 | 0.32395589 | 1.36334233 | 0.00128002 | 0.02332695 | 0.01936623 | 2183 | tags=25%, list=13%, signal=22% | [illegible] |
| GO:1990948 | ubiquitin ligase inhibitor activity | 10 | 0.8355699 | 1.80059884 | 0.00156274 | 0.02775669 | 0.02304383 | 807 | tags=50%, list=5%, signal=48% | [illegible] |
| GO:0019200 | carbohydrate kinase activity | 19 | -0.7179122 | -1.7817983 | 0.00156856 | 0.02775669 | 0.02304383 | 1455 | tags=47%, list=9%, signal=43% | [illegible] |
| GO:0019213 | deacetylase activity | 32 | -0.6353857 | -1.7390607 | 0.00180901 | 0.03110988 | 0.02582768 | 1384 | tags=28%, list=8%, signal=26% | [illegible] |
| GO:0016758 | hexosyltransferase activity | 151 | -0.4368562 | -1.4981735 | 0.00179085 | 0.03110988 | 0.02582768 | 4862 | tags=43%, list=28%, signal=31% | [illegible] |
| GO:0022834 | ligand-gated channel activity | 119 | -0.4573556 | -1.5258369 | 0.00209819 | 0.03558183 | 0.02954033 | 4099 | tags=39%, list=24%, signal=30% | [illegible] |
| GO:0071813 | lipoprotein particle binding | 24 | -0.6635615 | -1.7347264 | 0.00235078 | 0.03776711 | 0.03135456 | 3723 | tags=50%, list=22%, signal=39% | [illegible] |
| GO:0071814 | protein-lipid complex binding | 24 | -0.6635615 | -1.7347264 | 0.00235078 | 0.03776711 | 0.03135456 | 3723 | tags=50%, list=22%, signal=39% | [illegible] |
| GO:0022824 | transmitter-gated monoatomic ion channel activity | 31 | -0.6320842 | -1.7201919 | 0.00230332 | 0.03776711 | 0.03135456 | 4099 | tags=45%, list=24%, signal=34% | [illegible] |
| GO:0022835 | transmitter-gated channel activity | 31 | -0.6320842 | -1.7201919 | 0.00230332 | 0.03776711 | 0.03135456 | 4099 | tags=45%, list=24%, signal=34% | [illegible] |
| GO:0005261 | monoatomic cation channel activity | 233 | -0.4099006 | -1.4537516 | 0.00256233 | 0.04063126 | 0.0337324 | 3717 | tags=33%, list=22%, signal=26% | [illegible] |
| GO:0008137 | NADH dehydrogenase (ubiquinone) activity | 41 | 0.53302071 | 1.68204815 | 0.00277685 | 0.04346837 | 0.03608779 | 1389 | tags=32%, list=8%, signal=29% | [illegible] |
| GO:0004984 | olfactory receptor activity | 30 | -0.6217142 | -1.6788784 | 0.00295252 | 0.04563329 | 0.03788513 | 4215 | tags=50%, list=25%, signal=38% | [illegible] |
| GO:0031420 | alkali metal ion binding | 17 | -0.7202807 | -1.7452755 | 0.0032323 | 0.04698375 | 0.03900629 | 1214 | tags=47%, list=7%, signal=44% | [illegible] |
| GO:0005253 | monoatomic anion channel activity | 71 | -0.5100654 | -1.5824422 | 0.00317518 | 0.04698375 | 0.03900629 | 4299 | tags=48%, list=25%, signal=36% | [illegible] |
| GO:0019955 | cytokine binding | 113 | -0.470266 | -1.5537062 | 0.00319022 | 0.04698375 | 0.03900629 | 3554 | tags=37%, list=21%, signal=30% | [illegible] |
| GO:0008509 | monoatomic anion transmembrane transporter activity | 109 | -0.4722176 | -1.553548 | 0.00315041 | 0.04698375 | 0.03900629 | 4691 | tags=47%, list=27%, signal=34% | [illegible] |
| GO:0044389 | ubiquitin-like protein ligase binding | 293 | 0.31947105 | 1.34085484 | 0.00321378 | 0.04698375 | 0.03900629 | 2549 | tags=28%, list=15%, signal=24% | [illegible] |
| GO:0001221 | transcription coregulator binding | 108 | 0.40043348 | 1.48631314 | 0.00339336 | 0.04874458 | 0.04046815 | 1902 | tags=27%, list=11%, signal=24% | [illegible] |
| GO:0061629 | RNA polymerase II-specific DNA-binding transcription factor binding | 306 | 0.31142687 | 1.31143679 | 0.00351703 | 0.0499337 | 0.04145537 | 1843 | tags=24%, list=11%, signal=22% | [illegible] |
